# Genome-wide association and Mendelian randomization analyses of host response to *Helicobacter pylori* infection implicate HLA polymorphisms and immune-mediated diseases

**DOI:** 10.64898/2026.03.27.26349357

**Authors:** Tomoki Kyosaka, Akira Narita, Jerzy K Kulski, Aye Ko Ko Minn, Akimitsu Miyake, Yurii Kotsar, Kyoga Hiraide, Takafumi Ojima, Masahiro Nakatochi, Shinichi Namba, Taiki Yamaji, Yoichi Sutoh, Yu Sasaki, Linda Broer, Fabian Frost, Yuriko N. Koyanagi, Yumiko Kasugai, Hidemi Ito, Norie Sawada, Shiori Nakano, Sadao Suzuki, Asahi Hishida, Teruhide Koyama, Yoko Kubo, Takamitsu Funayama, Satoshi Makino, Matsuyuki Shirota, Jun Takayama, Chinatsu Gocho, Sachiyo Sugimoto, Yayoi Otsuka-Yamasaki, Kozo Tanno, Yasuhiko Abe, Osamu Nakajima, Manon C W Spaander, Stefan Weiss, Markus M. Lerch, Daniel Levy, Shih-Jen Hwang, Alexis C Wood, Stephen S. Rich, Jerome I Rotter, Kent D. Taylor, Russell P. Tracy, Hannah Stocker, Hermann Brenner, Mārcis Leja, Raitis Peculis, Atsushi Hozawa, Kengo Kinoshita, Atsushi Shimizu, Masayuki Yamamoto, Keitaro Matsuo, Motoki Iwasaki, Kenji Wakai, Yoshiyuki Ueno, Gwenny M Fuhler, Georg Homuth, Maikel P Peppelenbosch, the Tohoku Medical Megabank Project Study Group, the Biobank Japan Project, Yukinori Okada, Shinichi Kuriyama, Motomichi Matsuzaki, Gen Tamiya

**Author notes:** Corresponding authors: Akira Narita, Motomichi Matsuzaki, and Gen Tamiya. A full list of members and their affiliations appears in the Supplementary Information.

## Abstract

*Helicobacter pylori* (*H. pylori*) infects the gastric epithelium of approximately half of the global population, and is a well-known risk factor for developing gastric cancer. Despite the clinical significance of *H. pylori* infection, many genetic factors underlying host response to *H. pylori* remain unidentified. Moreover, although *H. pylori* infection has been implicated in a range of extra-gastric diseases, whether these associations share a common genetic architecture remains unclear. We performed a genome-wide association study (GWAS) of anti-*H. pylori* IgG antibody titers, a composite host phenotype reflecting susceptibility to persistent infection and subsequent immune activation, followed by gene-based and pathway analyses, involving up to 140,863 individuals. This included 56,967 individuals in the discovery phase and 68,211 in the replication phase from Japanese cohorts, together with an additional 15,685 from European populations in a cross-ancestry meta-analysis. We reveal significant associations between IgG titers and polymorphisms in the Human Leukocyte Antigen (HLA) class II region within the Major Histocompatibility Complex (MHC), as well as genes related to innate immunity, including *CCDC80*, *NFKBIZ*, *TIFA*, *PSCA*, and *TRAF3*. Mendelian randomization (MR) analyses revealed directionally consistent associations between genetic liability to higher anti–*H. pylori* IgG levels and a range of diseases, including immune-mediated diseases such as type 1 diabetes, Hashimoto’s disease, atopic dermatitis, as well as quantitative traits like body height and weight. These genetic findings support the interpretation that host genetic determinants of response to *H. pylori*, particularly those mediated through HLA class II, form part of a shared immunogenetic architecture associated with susceptibility to multiple immune-related diseases.

## Main

*Helicobacter pylori* (*H. pylori*) is a spiral-shaped, gram-negative bacterium first identified in human gastric mucosa by Warren and Marshall in 1983^[1]^. Its infection is well-known for causing digestive diseases such as gastritis and peptic ulceration^[2]^. The link to gastric and esophageal cancer has been of particular interest^[3][4]^, leading to its classification as a Group 1 carcinogen (a confirmed cause of cancer) for gastric cancer by the International Agency for Research on Cancer (IARC) in 1994^[5]^. By 2018, it was recognized as one of the most significant infectious causes of cancer worldwide^[6]^. Beyond digestive diseases, host responses to *H. pylori*, often assessed using serological markers, have been linked epidemiologically to various other conditions, including autoimmune diseases such as Type 1 diabetes (T1D)^[7]^, Hashimoto’s disease^[8]^, Type 2 diabetes (T2D)^[9]^ and coronary artery disease^[10]^. Additionally, inverse associations have been reported between *H. pylori* infection and allergic diseases^[11]^, including asthma and atopic dermatitis.

*H. pylori* is one of the most prevalent infectious bacteria in humans, although its prevalence varies greatly by region and the ancestral background of the host population. For example, the infection rates are as low as 18.9% in Switzerland, and as high as 87.7% in Nigeria^[12]^. Globally, it is estimated that more than half the world’s population is infected^[12]^. In developed countries, the prevalence of *H. pylori* has declined due to improved hygiene and the widespread use of eradication treatments, whereas it remains high in developing regions, such as Africa, Latin America, and Asia^[13]^. Infection typically occurs during childhood, often before the age of five^[14]^. There also is considerable variation in how individuals acquire and retain the bacterial infection. Twin studies have shown higher concordance rates for *H. pylori* infection in monozygotic twins than in dizygotic twins^[15]^, suggesting that host genetic factors contribute to inter-individual variation in response to *H. pylori*, in addition to environmental influences on pathogen encounter.

Genome-wide association studies (GWAS) of anti-*H. pylori* IgG antibody titers in European populations have identified significant associations with genes on chromosome 4p14, which encode the Toll-like receptors (*TLR1*, *TLR6*, and *TLR10*), key regulators of pathogen recognition and initiation of innate immunity^[16][17]^. These findings mainly highlight innate immune recognition pathways, whereas a broader UK Biobank GWAS and HLA association study of infectious-disease serology implicated the MHC/HLA region as a recurrent determinant of pathogen-specific antibody responses^[18]^. However, large-scale GWAS have not yet been performed in Japanese or other East Asian populations, where more virulent strains of *H. pylori* are prevalent^[19]^, leaving the involvement of HLA variation in *H. pylori*–related serological host responses in these populations unresolved. While numerous epidemiological studies have examined the relationship between *H. pylori*–related phenotypes and a wide range of diseases^[7][8][9][10][11]^, large-scale genetic analyses that systematically characterize the host immunogenetic background underlying these associations remain limited.

To investigate the genetic architecture underlying host response to *H. pylori*, we performed the largest GWAS to date of anti-*H. pylori* IgG antibody titers, a composite host phenotype reflecting susceptibility to persistent infection and subsequent immune activation, and widely used in population-based screening settings^[20]^. This analysis leveraged data from large Japanese cohorts and biobank resources, and was complemented by a cross-ancestry meta-analysis incorporating summary statistics from previous GWAS of European populations^[17]^. In addition, we evaluated whether genetic liability to higher anti–*H. pylori* IgG levels shows consistent associations with a wide range of diseases and quantitative traits, using Mendelian randomization (MR) and genetic correlation analyses.

## Results

We performed a GWAS of anti-*H. pylori* IgG antibody titers followed by subsequent analyses, involving up to 140,863 individuals: 56,967 in the discovery phase, 68,211 in the replication phase from Japanese cohorts, and an additional 15,685 from European populations^[17]^ in a cross-ancestry meta-analysis. MR analyses identified directionally consistent positive and inverse associations between genetic liability to higher anti-*H. pylori* IgG levels and a range of diseases. An overview of the workflow is provided in **Fig. 1**.

**Fig. 1.**
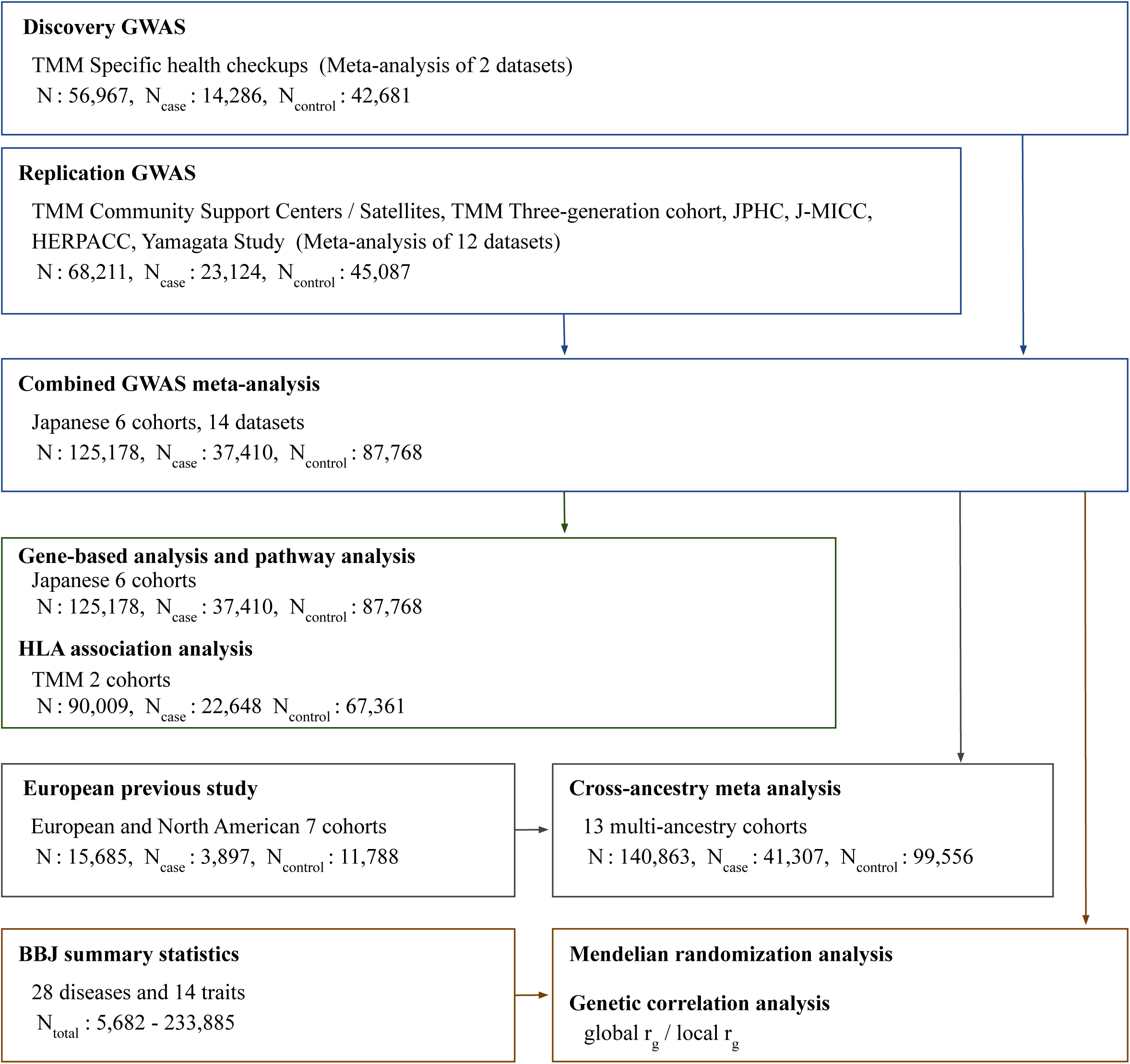
Overview of cohorts and study design. Genome-Wide Association Studies (GWAS) were performed, followed by gene-based analysis, pathway analysis, and HLA-association analysis. Cross-ancestry GWAS meta-analyses were performed between Japanese and European populations to investigate genetic differences. Additionally, Mendelian randomization (MR) and genetic correlation analyses were used to assess the causal relationships between genetic liability to high anti-*H. pylori* IgG levels and other diseases. The Japanese study cohorts included Tohoku Medical Megabank Organization (TMM), Japan Public Health Center-based Prospective Study (JPHC), Japan Multi-Institutional Collaborative Cohort Study (J-MICC), Hospital-based Epidemiologic Research Program at Aichi Cancer Center (HERPACC), and the Yamagata Study. See **Supplementary Information** for additional cohort details.

### Genome-wide association studies

In the discovery GWAS of 56,967 participants from specific health checkups in the Tohoku Medical Megabank Community-Based Cohort (TMM CommCohort)^[21]^, genome-wide significant associations (P-value < 5×10^-8^) of anti-*H. pylori* IgG antibody titers were identified in four genomic regions (**Fig. 2a**, **Table 1, and Supplementary Fig. 2**). No inflation of test statistics due to population stratification was observed (λGC score: 1.048). The strongest association was found in the Major Histocompatibility Complex (MHC) class II region at 6p21.3, with the lead SNP rs28383359 (Odds ratio (OR) = 1.155, 95% CI: 1.122 – 1.189, P-value = 4.44×10^-24^) located in UTR3 of the *HLA-DQA1* gene. The second strongest association was at 3q13.2, with the lead SNP rs146203219 (OR = 1.139, 95% CI: 1.105 – 1.174, P-value = 1.56×10^-18^) located in the intergenic region between *CCDC80* (Coiled-coil domain containing 80) and *CD200R1L* (CD200 receptor 1-Like). The third strongest association was at 17p12, with the lead SNP rs1078643 (OR = 1.140, 95% CI: 1.101 – 1.181, P-value = 2.74×10^-14^) located in the exonic region of *LINC00675* (long intergenic non-coding RNA 00675). The fourth strongest association was at 14q32.33, with the lead SNP rs62637720 (OR = 1.169, 95% CI: 1.114 – 1.227, P-value = 7.24×10^-11^) located in the intergenic region between *IGHE* (Immunoglobulin Heavy Constant Epsilon) and *IGHG4* (Immunoglobulin Heavy Constant Gamma 4).

**Fig. 2.**
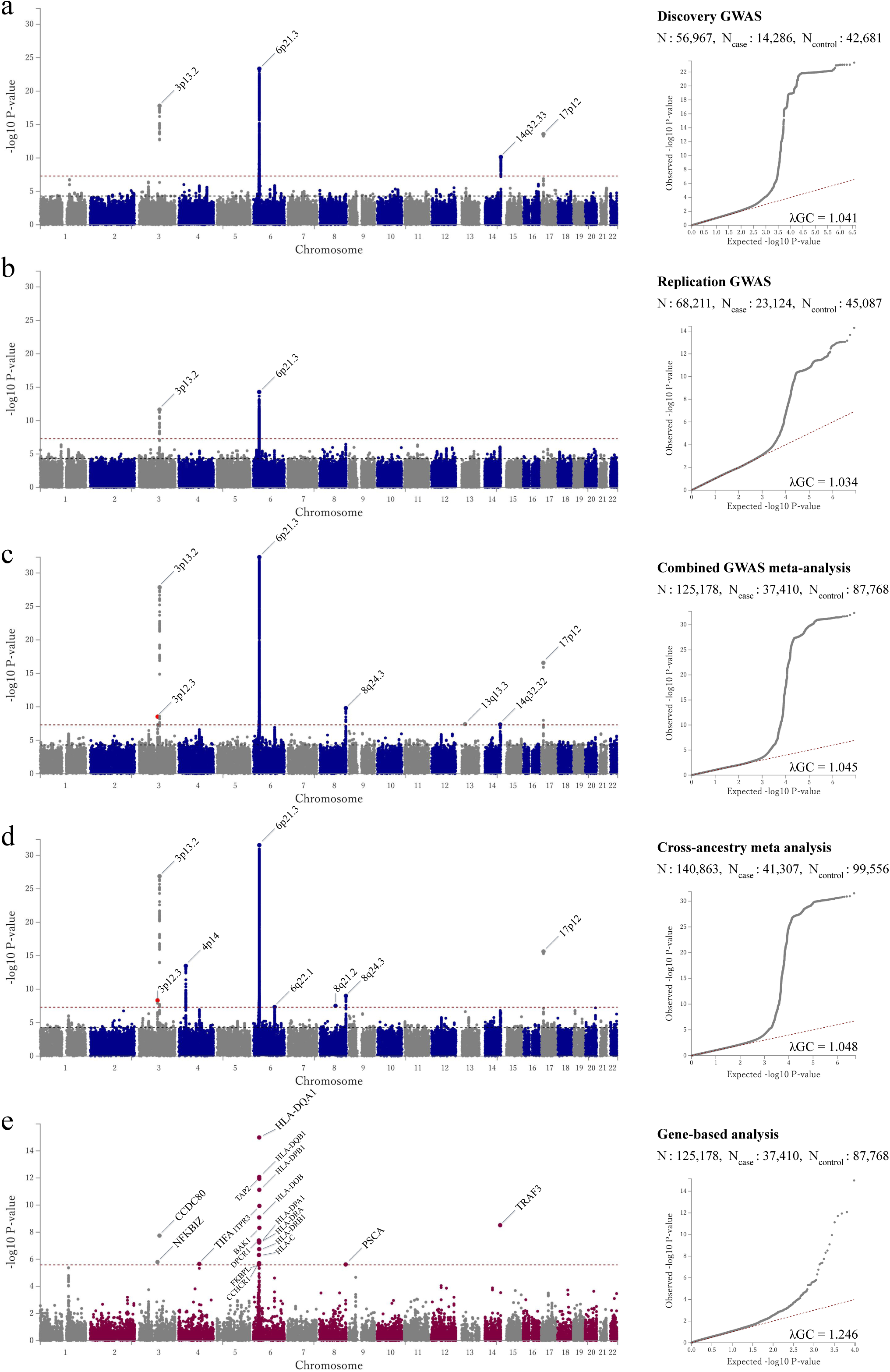
Manhattan and Q-Q plots of the study. Manhattan plots and Quantile-Quantile (Q-Q) plots were generated using the FUMA platform (v1.6.1). **a,** Plots of the discovery GWAS. P-values were derived from the meta-analysis of 14,286 cases and 42,681 controls of Japanese ancestry. The x-axis represents the chromosomal positions and the y-axis represents the -log10 P-values. The red dashed line indicates the genome-wide significance threshold (P-value < 5×10^-8^). The black dashed line indicates the suggestive threshold (P-value < 5×10^-5^). Variants are plotted against GRCh37 (hg19). In the Q-Q plot, the negative logarithm of the observed (y-axis) and the expected (x-axis) P-value were plotted for each SNP (dot), and the red dashed line (y = x) indicates the null hypothesis of no true association. The regression genomic inflation factor (λGC score) is 1.041. **b,** Plot of the replication GWAS. P-values were derived from the meta-analysis of 23,124 cases and 45,087 controls of Japanese ancestry. The λGC score is 1.034. **c,** Plots of the combined GWAS meta-analysis. P-values were derived from the meta-analysis of 37,410 cases and 87,768 controls of Japanese ancestry. The λGC score is 1.045. **d,** Manhattan plot of the cross-ancestry meta-GWAS. P-values were derived from the meta-analysis of 41,307 cases and 99,556 controls of Japanese and European ancestry. The λGC score is 1.048. **e,** Manhattan plot of the gene-based analysis. The x-axis represents the chromosomal positions and the y-axis represents the -log10 P-values. The red dashed line indicates the Bonferroni-corrected significance level (P-value < 0.05/19276 = 2.59×10^-6^). The λGC score is 1.246.

**Table 1.**
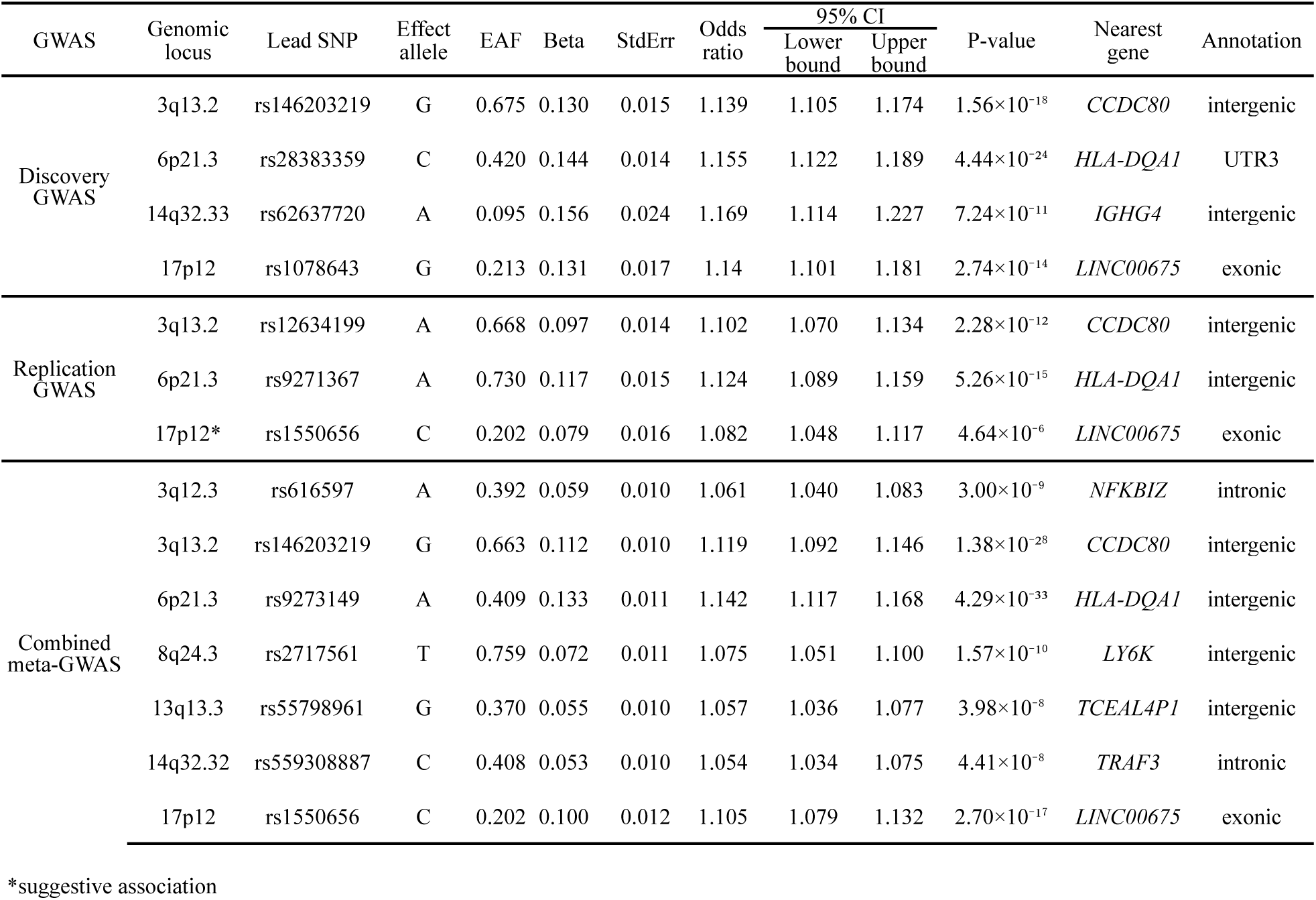
GWAS results: A list of regions reaching genome-wide significance.

For the replication phase, 12 datasets from six different cohorts within Japan were compiled for a GWAS meta-analysis involving a total of 68,211 individuals (details of the cohorts and datasets are provided in **Supplementary Table 1 and 2**). In this meta-analysis for the replication phase, two genomic regions, harboring *HLA-DQA1* and *CCDC80* reproducibly attained the genome-wide significance (**Fig. 2b**, **Table 1, and Supplementary Fig. 3**). The strongest association was found in the intergenic region between *HLA-DRB1* and *HLA-DQA1* at 6p21.3 (lead SNP: rs9271367, OR = 1.124, 95% CI: 1.089 – 1.159, P-value = 5.26×10^-15^), and the second strongest was in the intergenic region between *CCDC80* and *CD200R1L* at 3q13.2 (lead SNP: rs12634199, OR = 1.102, 95% CI: 1.070 – 1.134, P-value = 2.28×10^-12^). A suggestive association was found in the exonic region of *LINC00675* at 17p12 (lead SNP: rs1550656, OR = 1.082, 95% CI: 1.048 – 1.117, P-value = 4.64×10^-6^).

We then meta-analyzed the discovery and replication GWAS of 125,178 individuals. This analysis revealed that, in addition to *HLA-DQA1, CCDC80* and *LINC00675*, four other regions also reached genome-wide significance (**Fig. 2c and Table 1**). The strongest association was in the intergenic region between *HLA-DQA1* and *HLA-DQB1* at 6p21.3 (lead SNP: rs9273149, OR = 1.142, 95% CI: 1.117 – 1.168, P-value = 4.29×10^-33^). The second strongest was in the intergenic region between *CCDC80* and *CD200R1L* at 3q13.2 (lead SNP: rs146203219, OR = 1.119, 95% CI: 1.092 – 1.146, P-value = 1.38×10^-28^), and the third strongest was in the exonic region of LINC00675 at 17p12 (lead SNP: rs1550656, OR = 1.105, 95% CI: 1.079 – 1.132, P-value = 2.70×10^-17^). The fourth strongest association was in the intergenic region between *LY6K* (lymphocyte antigen 6 family member K) and *PSCA* (prostate stem cell antigen) at 8q24.3 (lead SNP: rs2717561, OR = 1.075, 95% CI: 1.051 – 1.100, P-value = 1.57×10^-10^). The fifth strongest association was in the intronic region of *NFKBIZ* (NF-kappa-B inhibitor zeta) at 3q12.3 (lead SNP: rs616597, OR = 1.061, 95% CI: 1.040 – 1.083, P-value = 3.00×10^-9^). The sixth strongest association was in the intronic region of *TRAF3* (Tumor necrosis factor receptor-associated factor 3) at 14q32.32 (lead SNP: rs559308887, OR = 1.054, 95% CI: 1.034 – 1.075, P-value = 4.41×10^-8^). The seventh strongest association was in the intergenic region between *TCEAL4P1* (transcription elongation factor A like 4 pseudogene 1) and *ARL2BPP3* (ADP ribosylation factor like GTPase 2 binding protein pseudogene 3) at 13q13.3 (lead SNP: rs55798961, OR = 1.057, 95% CI: 1.036 – 1.077, P-value = 3.98×10^-8^).

### Gene-based analysis and pathway analysis

In the gene-based analysis, which is based on the combined GWAS meta-analysis using 125,178 individuals, 19 genes were identified (**Fig. 2e and Supplementary Table 4**) with a Bonferroni-corrected significance threshold (P-value < 0.05/19276 = 2.59×10^-6^). *HLA-DQA1* was the most significantly associated gene (P-value = 1.03×10^-15^) closest to the lead nucleotide variants of the GWAS (rs28383359 in the discovery P-value = 4.44×10^-24^; rs9271367 in the replication P-value = 5.26×10^-15^; rs9273149 in the combined P-value = 4.29×10^-33^). Among the genes outside the MHC region, significant gene-based associations were observed with *TRAF3* (P-value = 3.05×10^-8^), *CCDC80* (P-value = 1.80×10^-8^), *NFKBIZ* (P-value = 1.59×10^-6^), *TIFA* (TRAF interacting protein with forkhead associated domain, P-value = 2.21×10^-6^), and *PSCA* genes (P-value = 2.42×10^-6^); all these genes overlapped with the GWAS signals, except for *TIFA*, and all of them are known to be involved in innate _immune pathways_[22][23][24][25].

For the pathway analysis, gene mapping was performed using annotations obtained from ANNOVAR^[26]^. A broader set of 112 genes, selected from the pool of 20,260 protein-coding genes, was identified, enabling a comprehensive exploration of biological pathways on the FUMA platform^[27]^. Pathways from the Kyoto Encyclopedia of Genes and Genomes (KEGG) database^[28]^ with significant associations (adjusted P-value calculated using a FDR approach < 0.05) included the gene sets for “Antigen processing and presentation” (adjusted P-value (adj P) = 1.47×10^-24^), “Type I diabetes mellitus” (adj P = 8.34×10^-20^), “Allograft rejection” (adj P = 4.34×10^-19^), “Graft-versus-host disease” (adj P = 4.34×10^-19^), and “Autoimmune thyroid disease” (adj P = 7.60×10^-17^) (**Fig. 3a and Supplementary Table 5**). Pathways from the Reactome database^[29]^ showed significant associations with gene sets involved in both the adaptive immune system (“Adaptive immune system” (adj P = 3.17×10^-9^), “MHC class II antigen presentation” (adj P = 5.89×10^-8^), and “TCR signaling” (adj P = 8.06×10^-6^)) and the innate immune system (“TNF receptor superfamily members mediating non canonical NF-κB pathway” (adj P = 4.79×10^-3^), “Innate immune system” (adj P = 6.32×10^-3^), and “Non-canonical NF-κB pathway” (adj P = 9.09×10^-3^)) (**Fig. 3b and Supplementary Table 6**).

**Fig. 3.**
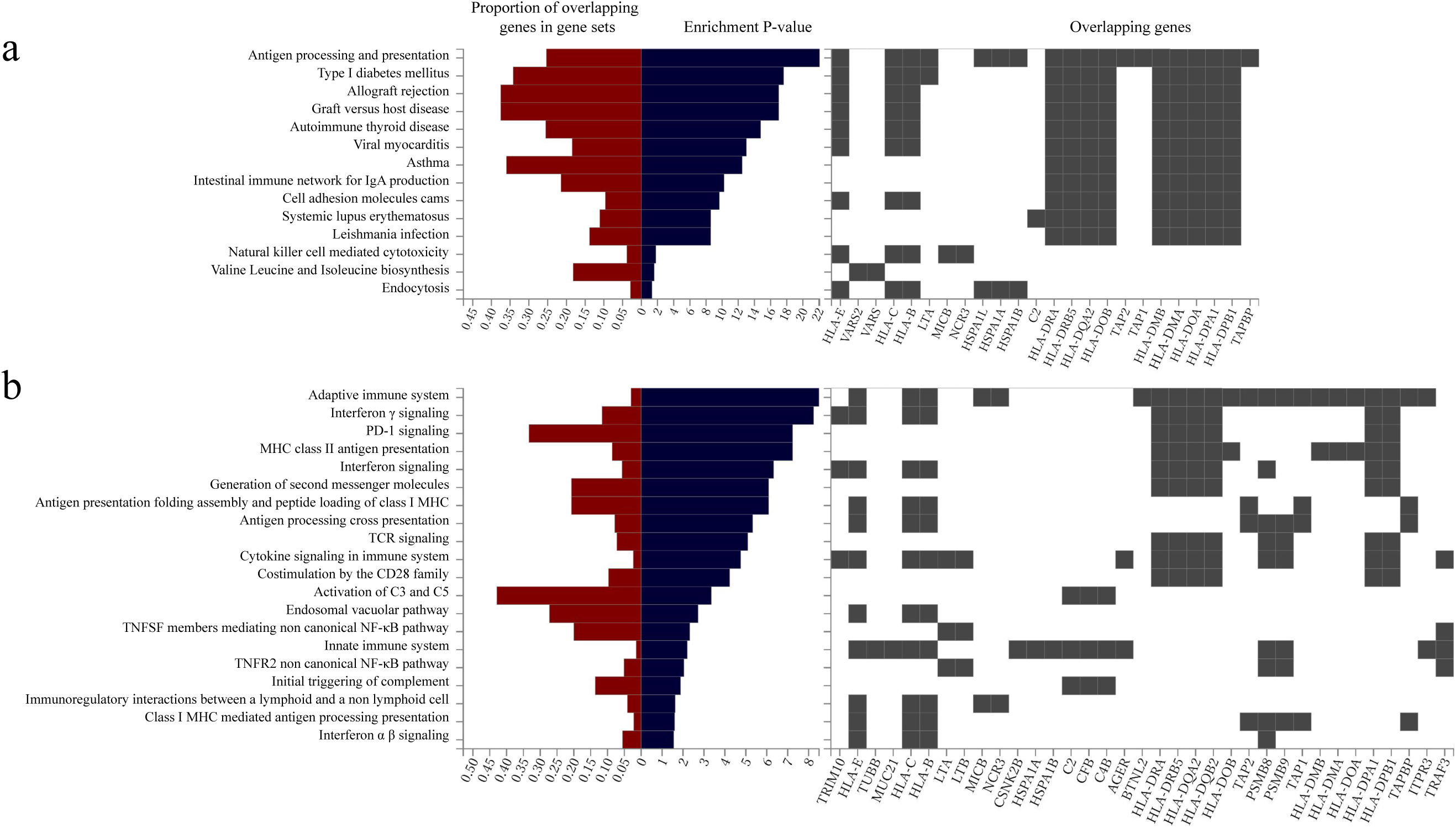
Pathway analysis using KEGG and Reactome pathways. Pathway analysis was performed using the GENE2FUNC function on the FUMA platform. Pathways with significant associations (adjusted P-value calculated using a FDR approach < 0.05) are displayed. The X-axis shows the proportion of overlapping genes in the gene set, the enrichment P-values (-log10 P-values), and the overlapping genes. The Y-axis displays a list of suspected related pathways and diseases. **a,** The results of the KEGG pathway analysis. **b,** The results of the Reactome pathway analysis.

### HLA imputation and HLA association analysis

When examining the linkage disequilibrium (LD) relationships using genome data from 90,009 individuals in the TMM cohorts (excluding those with close kinship), the lead variants identified in the GWAS of anti–*H. pylori* IgG antibody titers (rs28383359 in the discovery phase, rs9271367 in the replication phase, and rs9273149 in the combined analysis) were found to be in LD with each other. They were also in LD with the top variants in the promoter/enhancer regions of the MHC class II genes, as well as with the top missense variants of these genes. (**Supplementary Fig. 5a and 5b**).

Additionally, HLA imputation was performed using the DEEP*HLA software^[30]^ and HLA allele frequencies were calculated. The lead variants of the GWAS were found to be in particularly strong LD with HLA-DQA1*03:01 (rs28383359, D’ = 0.99, r² = 0.76; rs9271367, D’ = 0.99, r² = 0.19; and rs9273149, D’ = 0.99, r² = 0.78) (**Supplementary Fig. 5c and 5d**). To further clarify these associations, we closely examined the MHC region.

We identified several HLA alleles significantly associated with higher anti–*H. pylori* IgG antibody titers, including HLA-DQA1*03:01 (OR = 1.110, 95% CI: 1.086–1.134, P-value = 2.06×10⁻²⁰), DRB1*09:01 (OR = 1.124, 95% CI: 1.089–1.160, P-value = 1.46×10⁻¹³), DQB1*03:03 (OR = 1.116, 95% CI: 1.084–1.150, P-value = 9.12×10⁻¹³), and DPB1*05:01 (OR = 1.065, 95% CI: 1.042–1.088, P-value = 2.40×10⁻⁸). Conversely, several alleles were associated with lower anti–H. pylori IgG antibody titers, including HLA-DQB1*06:02 (OR = 0.869, 95% CI: 0.836–0.904, P-value = 1.41×10⁻¹²), DRB1*15:01 (OR = 0.899, 95% CI: 0.872–0.928, P-value = 9.34×10⁻¹¹), and DQA1*01:02 (OR = 0.918, 95% CI: 0.891–0.945, P-value = 2.23×10⁻⁹). A complete list of four-digit and two-digit HLA alleles meeting the Bonferroni-corrected significance threshold (P-value < 0.05/155 = 3.23×10⁻⁴) is provided in Table 2.

**Table 2.**
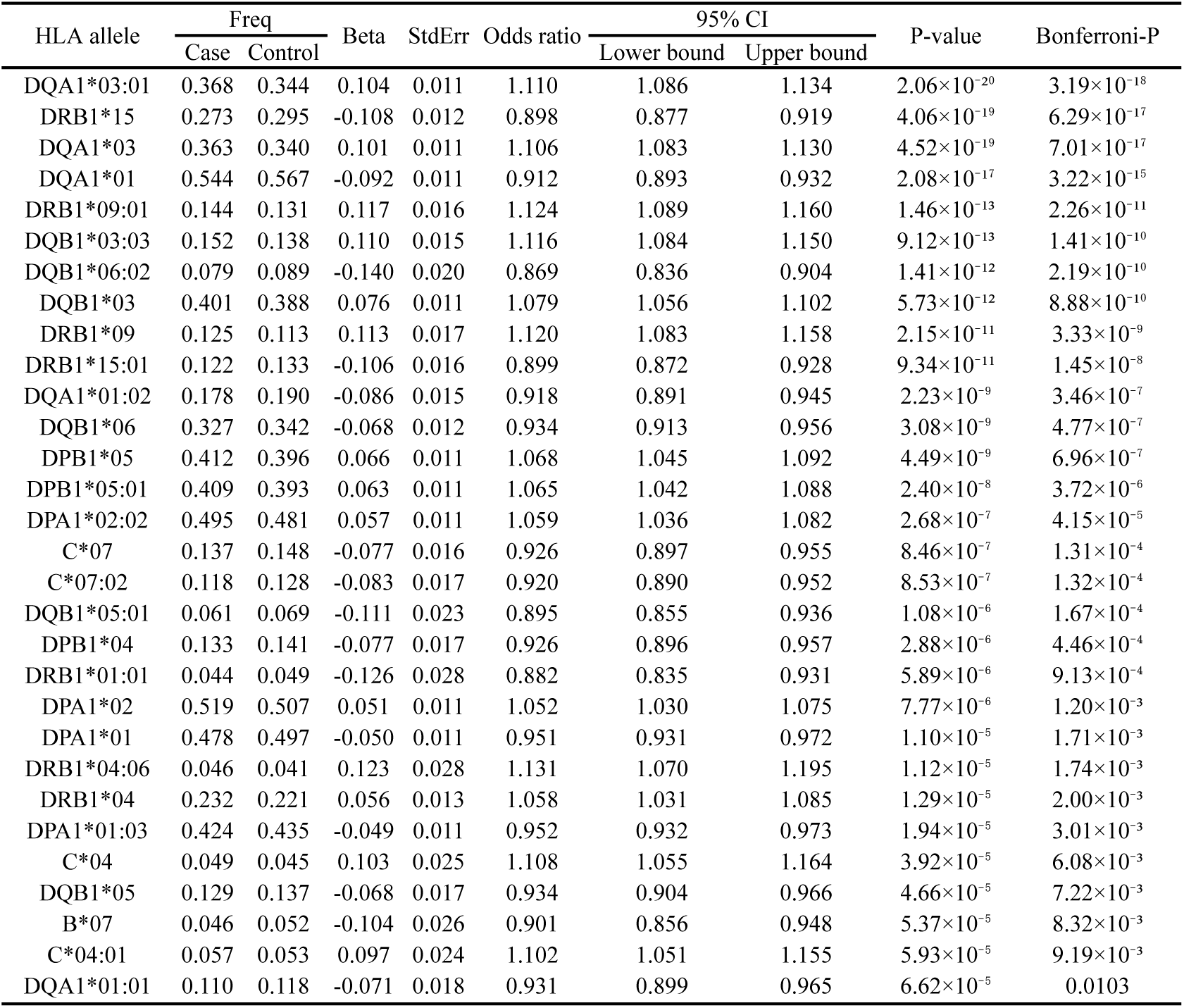
HLA association analysis (HLA alleles). The four-digit and two-digit HLA alleles that meet the significance level of the Bonferroni correction (P-value < 0.05/155 = 3.23×10 ⁻⁴).

| HLA allele | Freq |  | Beta | StdErr | Odds ratio | 95% CI |  | P-value | Bonferroni-P |
| --- | --- | --- | --- | --- | --- | --- | --- | --- | --- |
|  | Case | Control |  |  |  | Lower bound | Upper bound |  |  |
| DQA1*03:01 | 0.368 | 0.344 | 0.104 | 0.011 | 1.110 | 1.086 | 1.134 | $2.06 \times 10^{-20}$ | $3.19 \times 10^{-18}$ |
| DRB1*15 | 0.273 | 0.295 | -0.108 | 0.012 | 0.898 | 0.877 | 0.919 | $4.06 \times 10^{-19}$ | $6.29 \times 10^{-17}$ |
| DQA1*03 | 0.363 | 0.340 | 0.101 | 0.011 | 1.106 | 1.083 | 1.130 | $4.52 \times 10^{-19}$ | $7.01 \times 10^{-17}$ |
| DQA1*01 | 0.544 | 0.567 | -0.092 | 0.011 | 0.912 | 0.893 | 0.932 | $2.08 \times 10^{-17}$ | $3.22 \times 10^{-15}$ |
| DRB1*09:01 | 0.144 | 0.131 | 0.117 | 0.016 | 1.124 | 1.089 | 1.160 | $1.46 \times 10^{-13}$ | $2.26 \times 10^{-11}$ |
| DQB1*03:03 | 0.152 | 0.138 | 0.110 | 0.015 | 1.116 | 1.084 | 1.150 | $9.12 \times 10^{-13}$ | $1.41 \times 10^{-10}$ |
| DQB1*06:02 | 0.079 | 0.089 | -0.140 | 0.020 | 0.869 | 0.836 | 0.904 | $1.41 \times 10^{-12}$ | $2.19 \times 10^{-10}$ |
| DQB1*03 | 0.401 | 0.388 | 0.076 | 0.011 | 1.079 | 1.056 | 1.102 | $5.73 \times 10^{-12}$ | $8.88 \times 10^{-10}$ |
| DRB1*09 | 0.125 | 0.113 | 0.113 | 0.017 | 1.120 | 1.083 | 1.158 | $2.15 \times 10^{-11}$ | $3.33 \times 10^{-9}$ |
| DRB1*15:01 | 0.122 | 0.133 | -0.106 | 0.016 | 0.899 | 0.872 | 0.928 | $9.34 \times 10^{-11}$ | $1.45 \times 10^{-8}$ |
| DQA1*01:02 | 0.178 | 0.190 | -0.086 | 0.015 | 0.918 | 0.891 | 0.945 | $2.23 \times 10^{-9}$ | $3.46 \times 10^{-7}$ |
| DQB1*06 | 0.327 | 0.342 | -0.068 | 0.012 | 0.934 | 0.913 | 0.956 | $3.08 \times 10^{-9}$ | $4.77 \times 10^{-7}$ |
| DPB1*05 | 0.412 | 0.396 | 0.066 | 0.011 | 1.068 | 1.045 | 1.092 | $4.49 \times 10^{-9}$ | $6.96 \times 10^{-7}$ |
| DPB1*05:01 | 0.409 | 0.393 | 0.063 | 0.011 | 1.065 | 1.042 | 1.088 | $2.40 \times 10^{-8}$ | $3.72 \times 10^{-6}$ |
| DPA1*02:02 | 0.495 | 0.481 | 0.057 | 0.011 | 1.059 | 1.036 | 1.082 | $2.68 \times 10^{-7}$ | $4.15 \times 10^{-5}$ |
| C*07 | 0.137 | 0.148 | -0.077 | 0.016 | 0.926 | 0.897 | 0.955 | $8.46 \times 10^{-7}$ | $1.31 \times 10^{-4}$ |
| C*07:02 | 0.118 | 0.128 | -0.083 | 0.017 | 0.920 | 0.890 | 0.952 | $8.53 \times 10^{-7}$ | $1.32 \times 10^{-4}$ |
| DQB1*05:01 | 0.061 | 0.069 | -0.111 | 0.023 | 0.895 | 0.855 | 0.936 | $1.08 \times 10^{-6}$ | $1.67 \times 10^{-4}$ |
| DPB1*04 | 0.133 | 0.141 | -0.077 | 0.017 | 0.926 | 0.896 | 0.957 | $2.88 \times 10^{-6}$ | $4.46 \times 10^{-4}$ |
| DRB1*01:01 | 0.044 | 0.049 | -0.126 | 0.028 | 0.882 | 0.835 | 0.931 | $5.89 \times 10^{-6}$ | $9.13 \times 10^{-4}$ |
| DPA1*02 | 0.519 | 0.507 | 0.051 | 0.011 | 1.052 | 1.030 | 1.075 | $7.77 \times 10^{-6}$ | $1.20 \times 10^{-3}$ |
| DPA1*01 | 0.478 | 0.497 | -0.050 | 0.011 | 0.951 | 0.931 | 0.972 | $1.10 \times 10^{-5}$ | $1.71 \times 10^{-3}$ |
| DRB1*04:06 | 0.046 | 0.041 | 0.123 | 0.028 | 1.131 | 1.070 | 1.195 | $1.12 \times 10^{-5}$ | $1.74 \times 10^{-3}$ |
| DRB1*04 | 0.232 | 0.221 | 0.056 | 0.013 | 1.058 | 1.031 | 1.085 | $1.29 \times 10^{-5}$ | $2.00 \times 10^{-3}$ |
| DPA1*01:03 | 0.424 | 0.435 | -0.049 | 0.011 | 0.952 | 0.932 | 0.973 | $1.94 \times 10^{-5}$ | $3.01 \times 10^{-3}$ |
| C*04 | 0.049 | 0.045 | 0.103 | 0.025 | 1.108 | 1.055 | 1.164 | $3.92 \times 10^{-5}$ | $6.08 \times 10^{-3}$ |
| DQB1*05 | 0.129 | 0.137 | -0.068 | 0.017 | 0.934 | 0.904 | 0.966 | $4.66 \times 10^{-5}$ | $7.22 \times 10^{-3}$ |
| B*07 | 0.046 | 0.052 | -0.104 | 0.026 | 0.901 | 0.856 | 0.948 | $5.37 \times 10^{-5}$ | $8.32 \times 10^{-3}$ |
| C*04:01 | 0.057 | 0.053 | 0.097 | 0.024 | 1.102 | 1.051 | 1.155 | $5.93 \times 10^{-5}$ | $9.19 \times 10^{-3}$ |
| DQA1*01:01 | 0.110 | 0.118 | -0.071 | 0.018 | 0.931 | 0.899 | 0.965 | $6.62 \times 10^{-5}$ | 0.0103 |

Among the HLA amino acid polymorphism, residues at positions 98 and 104 of HLA-DRB1 protein (P-value = 3.71×10^-21^), positions 50, 53, and 215 of HLA-DQA1 protein (P-value = 8.66×10^-21^), and position 185 of HLA-DQB1 protein (P-value = 1.64×10^-19^) showed the most significant associations within their respective genes. A list of amino acid polymorphisms meeting the Bonferroni corrected significance level (P-value < 0.05/766 = 6.53×10^-5^) is presented in **Supplementary Table 7**. Structural mapping of these associated amino acid residues onto the HLA-DRB1, HLA-DQA1, and HLA-DQB1 protein structures is shown in **Supplementary Fig. 6a, 6b, and 6c**.

### Fine-mapping and conditional analyses in the MHC region

The MHC region is characterized by extreme polymorphism and extended linkage disequilibrium (LD), which can lead to apparent associations that reflect LD rather than true causal effects. To address this, we performed fine-mapping and conditional analyses.

Fine-mapping identified five variants within the HLA-DQA1 locus that showed equally high posterior inclusion probabilities (PIP = 1.00, logBF = 14.59) (**Supplementary Table 8**). These included both intronic and intergenic variants (e.g., rs533028614, rs114598250, rs7751376, rs9273382), as well as a missense variant, rs1047989 (C>A, p.Leu–16Met). This SNP is in moderate-to-strong LD with the HLA-DQA1*03:01 allele (D’ = 0.99, r² = 0.48), and a recent report suggested that this variant may affect the structural stability of HLA-DQA1^[31]^, supporting its biological plausibility.

When rs1047989 was included as a covariate in the conditional GWAS, the lead association signal in the HLA-DQA1 region was substantially attenuated, suggesting that this variant (or others in strong LD with it) explains the observed association to a meaningful extent. However, a residual peak remained within the MHC class II region, with a new lead SNP, rs9271501, emerging after conditioning (OR = 1.076, 95% CI: 1.053 – 1.099, P-value = 2.89×10^-11^; **Supplementary Fig. 7**). This variant lies between HLA-DRB1 and HLA-DQA1 and is in moderate-to-strong LD with the HLA-DRB1*15 allele (D’ = 0.91, r² = 0.48). In the HLA allele–level conditional analyses based on DEEP*HLA^[30]^ imputation, inclusion of rs1047989 as a covariate reduced the association of HLA-DQA1*03:01 allele to a non-significant level, while the signal for HLA-DRB1*15 allele remained statistically significant (OR = 0.934, 95% CI: 0.908 – 0.961, P-value = 1.52×10^-6^; **Supplementary Table 9**). These results suggest that the MHC class II association comprises at least two partly independent components driven by HLA-DQA1 and HLA-DRB1/DQB1 allelic polymorphisms.

### Cross-ancestry meta-analysis

A cross-ancestry meta-analysis was performed using summary statistics from a previous GWAS for anti-*H. pylori* IgG antibody titers involving 15,685 individuals of European ancestry^[17]^ along with our GWAS summary statistics from this study in six Japanese cohorts totaling 125,178 individuals. The MR-MEGA software^[32]^ analysis, based on principal components (PCs) for each dataset derived from a matrix of allele frequency similarities between GWASs, showed clear separation between the two main ancestral groups included in the study. Furthermore, no inflation of test statistics due to population stratification was observed (λGC score: 1.048), indicating that including the first principal component as a covariate sufficiently accounted for the differences in allele frequencies between populations.

The strongest association was found in the MHC class II region in intergenic between *HLA-DQA1* and *HLA-DQB1* at 6p21.3 (lead SNP: rs9273149, P-value = 2.99×10^-32^). The second strongest was in the intergenic region between *CCDC80* and *CD200R1L* at 3q13.2 (lead SNP: rs146203219, P-value = 1.32×10^-27^). The third strongest was in the exonic region of *LINC00675* at 17p12 (lead SNP: rs1550656, P-value = 2.34×10^-16^). The fourth strongest association was in the intergenic region between *TLR10* (Toll-Like Receptor 10) and *TLR1* (Toll-Like Receptor 1) at 4q14 (lead SNP: rs12233670, P-value = 3.31×10^-14^). The fifth strongest association was in the intergenic region between *LY6K* and *PSCA* at 8q24.3 (lead SNP: rs2717561, P-value = 1.02×10^-9^). The sixth strongest association was in the intronic region of *NFKBIZ* at 3q12.3 (lead SNP: rs616597, P-value = 4.81×10^-9^). Notably, for these signals, adding the European cohort to the cross-ancestry GWAS meta-analysis did not significantly weaken the strength of associations identified independently in each ancestry. On the other hand, the following associations were newly identified through the cross-ancestry meta-analysis. The seventh strongest association was in the intergenic region between *REXO1L2P* (REXO1 Like 2, Pseudogene) and *ATP6V0D2* (ATPase H+ Transporting V0 Subunit D2) at 8q21.2 (lead SNP: rs145917343, P-value = 3.16×10^-8^). The eighth strongest association was in the intronic region of *NT5DC1* (5’-Nucleotidase Domain Containing 1) at 6q22.1 (lead SNP: rs11153598, P-value = 4.69×10^-8^) (**Fig. 2d and Supplementary Table 10**).

In a previous GWAS of European population^[17]^, a genome-wide significant association was found at 4p14 with the top SNP rs12233670 located in the intergenic region between *TLR10* and *TLR1* (P-value = 4.42×10^-15^). This variant showed the highest heterogeneity compared to the Japanese population (Cochran’s Q-test P-value = 1.40×10^-13^, I² = 0.982), as we did not attain a genome-wide significance in the analysis of the Japanese population for this region (lead SNP: rs144493879, P-value = 1.11×10^-4^). Even after the cross-ancestry meta-analysis, no significant enhancement of the association was observed (P-value = 5.59×10^-4^). We present a comparison of the lead variants identified in both the Japanese and European ancestry GWAS across the loci found through the cross-ancestry meta-analysis (**Supplementary Table 11**). On the other hand, a close inspection of the previous GWAS result in the European population revealed a suggestive association in the intronic region of *HLA-DQA1* (lead SNP: rs9273255, P-value = 1.12×10^-5^; **Supplementary Fig. 8**).

### Mendelian randomization analysis and genetic correlation analysis

Mendelian randomization (MR) analyses were performed to evaluate whether genetic liability to higher anti–*H. pylori* IgG levels is associated with a range of diseases and quantitative traits in a directionally consistent manner. The combined GWAS meta-analysis results from 125,178 individuals in our Japanese cohorts were used as instrumental variables (IVs), while BioBank Japan (BBJ) summary statistics for a wide range of diseases/traits^[33]^ were used as the outcomes. Details of the outcome diseases/traits are provided in **Supplementary Table 12**. When genetic liability to higher anti–*H. pylori* IgG levels was used as the exposure, the inverse variance–weighted (IVW) MR analysis^[34]^ identified positive associations with autoimmune-related diseases, including type 1 diabetes, Hashimoto’s disease, pollinosis, and tuberculosis, as well as inverse associations with atopic dermatitis and anthropometric traits such as height and body weight (**Fig. 4, Supplementary Fig. 9, and Supplementary Table 13**). In addition to the IVW method, sensitivity analyses were performed using other statistical models (weighted median^[35]^, weighted mode^[36]^, MR-Egger^[37]^, and MR-cML^[38]^) to support the robustness of the results.

**Fig. 4.**
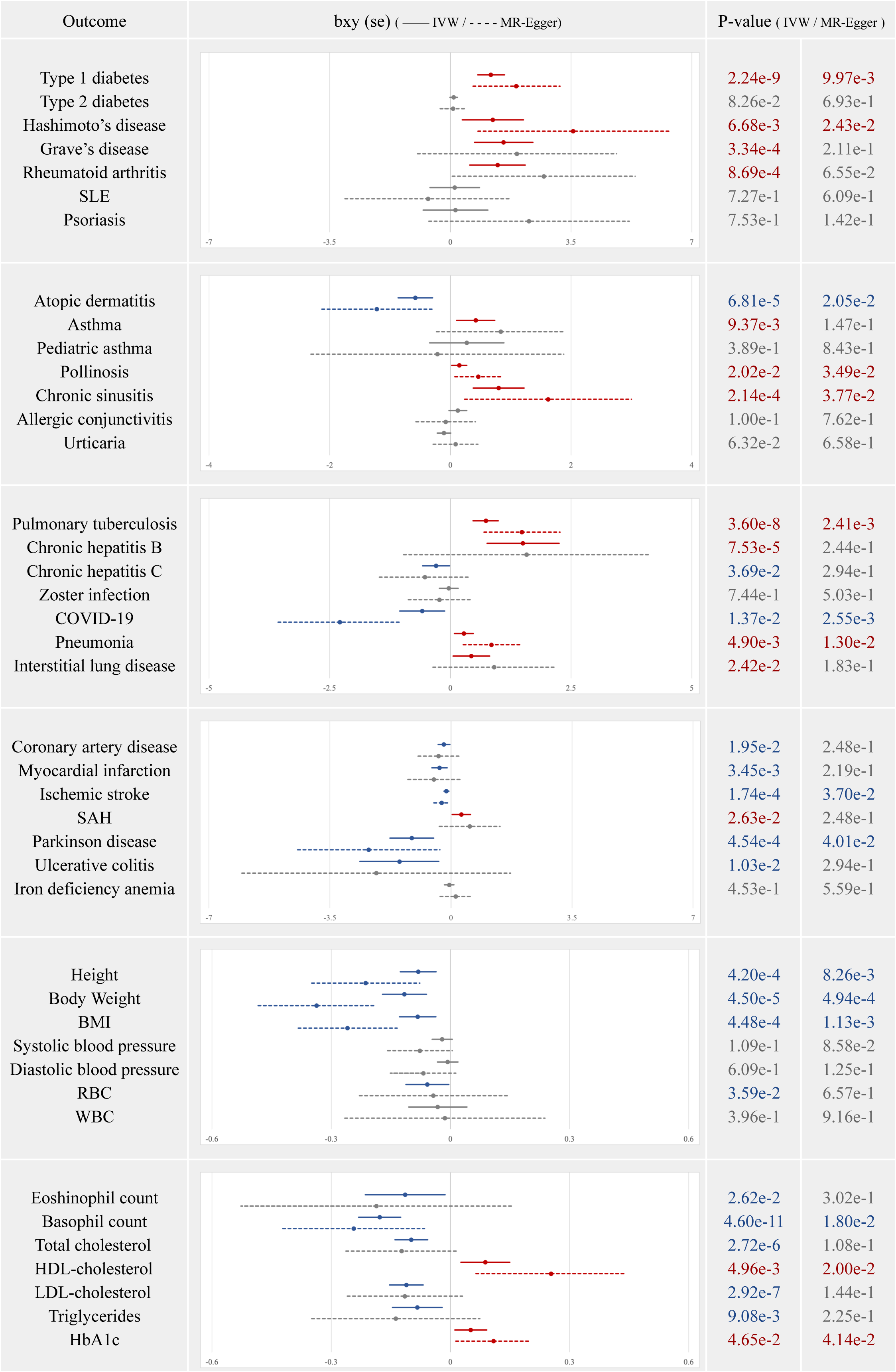
Forest plots of Mendelian randomization analysis (IVW, MR-Egger). Forest plots of Mendelian randomization (IVW and MR-Egger) for the association between genetic liability to high anti-*H. pylori* IgG levels and other diseases. The X-axis shows the beta coefficients for the presence of each disease (95% CI). The solid lines represent multiplicative random effects IVW, while the dashed lines represent MR-Egger. The red line indicates a positive association, the blue line indicates an inverse association, and the gray line indicates no significant association.

In the MR-Egger pleiotropy test, the presence of horizontal pleiotropy was suggested in the analysis involving three diseases/traits: COVID-19 infection, body weight, and BMI. For the other traits/diseases, no significant pleiotropy was observed; however, it is important to note that this merely supports the appropriateness of the selection of instrumental variables and does not rule out alternative biological pathways influencing the target diseases/traits^[37]^. Additionally, possibly due to the use of data from different cohorts for the instrumental variables and outcomes, heterogeneity was observed in the heterogeneity statistics (the Cochran’s Q P-value < FDR-adjusted threshold of 6.78×10^-3^). Nevertheless, a multiplicative random effects IVW analysis showed directionally consistent associations for many diseases/traits as the fixed effects IVW method. The leave-one-out sensitivity analysis shows that our results from the MR analysis are not dependent on any single variant.

In the global genetic correlation (global r_g_) analysis, significant correlations (FDR Q-value < 0.05) with anti–*H. pylori* IgG levels were not observed for any of the diseases or traits. However, in the local genetic correlation (local r_g_) analysis, a localized positive correlation within the MHC class II region was identified, particularly with autoimmune diseases: Type 1 diabetes (r_g_ = 0.360, P-value = 2.69×10^-11^), Graves’ disease (r_g_ = 0.172, P-value = 1.91×10^-6^), rheumatoid arthritis (r_g_ = 0.244, P-value = 2.48×10^-21^), and psoriasis (r_g_ = 0.238, P-value = 2.62×10^-11^; **Supplementary Table 15**). Furthermore, by focusing on the effect sizes of the variants used in the MR analysis, it was observed that particularly within the MHC region, certain variants confer shared risk for both anti–*H. pylori* IgG levels and these diseases (**Supplementary Fig. 10 and Supplementary Table 16**). Thus, the absence of significant global r_g_ signals is likely due in part to the opposing effects of the innate and adaptive immune functions across different genomic regions, which may cancel each other when averaging across the entire genome.

## Discussion

*H. pylori* infection is a significant factor influencing the onset and progression of various diseases[7][8][9][10][11], including but not limited to gastric cancer[5][6]. In light of its widespread prevalence and associated health implications, we performed the largest GWAS to date, subsequent analyses, cross-ancestry GWAS meta-analyses among Japanese and European populations, and MR and genetic correlation analyses to elucidate the genetic determinants of host response to *H. pylori* and to assess their broader disease relevance. Essentially, this comprehensive genetic study links these determinants with key regulators of innate and adaptive immunity—particularly genes in the nuclear factor-kappa B (NF-κB) pathway and the MHC class II region—and with multiple immune-related diseases.

The NF-κB transcription factor is pivotal in controlling cellular processes^[39]^ such as proliferation, survival, inflammatory responses, and immune reactions that can be activated by various stimuli, including microbial infections^[40]^. Our gene-based analysis identified *TRAF3*, *CCDC80*, *NFKBIZ*, *TIFA*, and *PSCA* that are involved with NF-κB or alternative immune regulatory pathways. The *CCDC80* gene plays a critical role in various physiological processes within the human body, particularly with cancer suppression^[41]^, metabolic regulation^[42]^, and has been implicated in atopic dermatitis^[43]^. Prior research demonstrated that *CCDC80* is involved in the suppression of the STAT3 pathway^[23]^ and the modulation of PPARγ expression^[42]^. Both STAT3 and PPARγ are known to regulate innate immune pathways, such as the JAK-STAT signaling pathway and the NF-κB pathway, in modulating inflammatory responses during *H. pylori* infection^[44][45]^. These findings raise the possibility that genetic variation in *CCDC80* influences immune pathways involved in the host response to *H. pylori*, which may contribute to inter-individual differences in anti–*H. pylori* IgG levels.

Several other genes that we identified also have essential roles in different feedback steps of the NF-κB signaling pathway. For instance, IκBζ, encoded by *NFKBIZ*, interacts with the p50 subunit of the NF-κB pathway in the nucleus, thereby regulating the expression of inflammatory cytokines such as IL-6^[24]^. *TIFA* has been shown to activate both the classical and alternative NF-κB pathways during *H. pylori* infection, particularly influencing the alternative pathway through *TRAF3*^[22]^. Additionally, *PSCA* has been reported to regulate IL-6 expression through the p38/NF-κB signaling axis in prostate cancer cells^[25]^. TLR10, highlighted in previous GWAS^[16][17]^, was shown experimentally to activate NF-κB in *H. pylori*-infected cells^[46]^, suggesting that this pathway may influence host response not only in Japanese populations but also across European cohorts. Although the *TLR1/6/10* gene region was significantly associated in a previous European study, our analysis revealed considerable effect-size heterogeneity between the European and Japanese populations, preventing the replication of these associations at a genome-wide significance level. This heterogeneity may arise from variations in allele frequencies of SNPs in the *TLR1/6/10* region between the two populations, necessitating further exploration of the genetic underpinnings of host response across diverse ethnic groups. Nevertheless, the role of the NF-κB pathway in *H. pylori* infection has been extensively documented in cellular and molecular studies^[47]^, and our findings further emphasize its critical importance in understanding the pathophysiology of host response to *H. pylori*.

The adaptive immune response and HLA class II gene expression is connected to the NF-κB pathway via T cell development and differentiation, the MHC class II transactivator (*CIITA*), and various other factors such as interferon-γ (IFN-γ), IκB kinase (IKK) complexes, and the regulatory genes in the MHC genomic region like *TNF* (Tumor Necrosis Factor) and *LT* (Lymphotoxin)^[48][49]^. Our analyses consistently revealed that anti–*H. pylori* IgG levels were most strongly associated with the MHC class II region. Although the LD structure in this region is complex, we observed that the top variants identified in the GWAS were in LD with lead variants located in the promoter/enhancer regions of MHC class II genes, as well as with significant missense variants of these genes. This prompted us to conduct HLA imputation and association studies, revealing that specific HLA alleles, including HLA-DQA1*03:01, -DRB1*09:01, -DQB1*03:03, and -DPB1*05:01, were strongly associated with higher anti–*H. pylori* IgG levels. Notably, these alleles were implicated in the risk of developing or exacerbating several autoimmune and collagen-related diseases, such as type 1 diabetes^[50]^, Hashimoto’s thyroiditis^[51]^, ulcerative colitis^[52]^, myasthenia gravis^[53]^, and ANCA-associated diseases^[54]^, particularly in East Asian populations. This observation suggests a shared genetic architecture within the MHC class II region linking host response to *H. pylori* and multiple immune-related diseases. Conversely, certain HLA alleles, including HLA-DQB1*06:02, -DRB1*15:01, and -DQA1*01:02, exhibited associations with lower anti–*H. pylori* IgG levels, consistent with a protective role. These polymorphisms previously were reported to confer protection against conditions such as type 1 diabetes^[50]^ and HTLV-1-associated myelopathy^[55]^. Our amino acid polymorphism analysis revealed widespread associations with amino acids encoded by *HLA-DRB1*, -*DQA1*, and -*DQB1*. Notably, a significant association was identified at amino acid position 185 of HLA-DQB1 protein. Although this position is not within the peptide-binding groove, the substitution of threonine with isoleucine at this site may alter the DQ-DM anchoring mechanism through interactions with adjacent residues, potentially influencing susceptibility to type 1 diabetes^[30]^, ANCA-associated vasculitis^[56]^, and narcolepsy^[57]^.

Given the complex LD architecture of the MHC region, we tested whether these associations were artifacts of long-range LD by performing fine-mapping and conditional analyses. Fine-mapping identified rs1047989, a missense variant in *HLA-DQA1*, as one of the high-confidence candidates with a high posterior probability. Notably, the amino-acid substitution corresponding to rs1047989 (position −16 of the HLA-DQA1 protein) was also significantly associated in an independent DEEP*HLA^[30]^–based amino-acid–level analysis, providing convergent evidence at the same site. Furthermore, conditional GWAS and HLA allele–level models showed that adjusting for rs1047989 reduced the association for HLA-DQA1*03:01 allele, whereas the signal for HLA-DRB1*15 allele remained statistically significant. This pattern indicates multiple, partly distinct associations driven by allelic variation within the HLA class II loci (e.g., *HLA-DRB1*, *DQA1*, and *DQB1*), although fine-mapping resolution is limited by the region’s LD complexity. Collectively, while pinpointing the exact causal variant remains challenging within the MHC region, these findings support genuine HLA class II–driven effects on host response to *H. pylori* rather than artifacts of extended LD.

Before our study, the association between host response to *H. pylori* and HLA alleles had not been clearly established, although several small HLA-association studies suggested that certain HLA class II alleles might be associated with variation in *H. pylori* infection status or immune response. For example, a study in Turkey^[58]^ reported that HLA-DRB1*11:04 was associated with higher prevalence of *H. pylori* infection compared with HLA-DRB1*03:01. However, the frequencies of these alleles in the Japanese population are extremely low (<0.1%), which raises questions about their relevance in the context of our study. Nonetheless, considering that HLA class II molecules play a crucial role in presenting extracellular antigens to CD4+ T cells, it is reasonable to surmise that these molecules are integral to immune responses against pathogens like *H. pylor*i. In fact, the Turkish study revealed through in silico analysis that HLA-DRB1*03:01 exhibits a stronger binding capacity for *H. pylori*-derived CagA peptides compared to HLA-DRB1*11:04, suggesting that HLA-DRB1*03:01 may confer protective effects against *H. pylori* infection by eliciting a more effective immune response.

Based on our findings from the HLA association and pathway analyses, we infer the existence of a common pathogenic mechanism linking host response to *H. pylori* with certain diseases, particularly those whose risks are influenced by HLA class II alleles. It should be noted that pathway analysis may not fully account for LD between genes, which may result in an overestimation of significance levels when including gene-dense regions such as the MHC^[59]^. Nonetheless, our study, including GWAS and other analyses, consistently highlights the importance of the MHC region, supporting the validity of these findings. The influence of HLA polymorphisms extends beyond this context, encompassing specific diseases such as autoimmune and allergic conditions. While the precise mechanisms by which specific HLA class II genes affect susceptibility and protection against these diseases remain ambiguous, their involvement in various immunological processes has been widely suggested. Mechanisms such as allelic differences in self-epitope presentation^[60]^, alterations in peptide presentation due to HLA protein stability^[61]^, and the activation of autoimmune responses resulting from misfolded protein/HLA class II molecule complex formations^[62]^ are all potential pathways worth exploring.

Our large-scale GWAS and MR analysis enabled a systematic evaluation of directionally consistent associations between genetic liability to high anti–*H. pylori* IgG levels and a broad range of diseases and quantitative traits, particularly immune-related conditions. MR is generally applied to infer causal effects by using genetic variants as instrumental variables^[63]^. In this context, several methodological challenges have been raised regarding MR analyses that use infectious diseases as exposures^[64]^. For example, genetic instruments for infection-related traits may capture not only biological susceptibility but also variation in exposure opportunity, which is often strongly influenced by environmental and social factors. However, the interpretation of our results requires careful consideration of the biological meaning of the exposure used in this study. Anti–*H. pylori* IgG levels do not simply reflect pathogen encounter; rather, they represent a composite host phenotype shaped by persistent *H. pylori* infection and the host immune response after infection has been established^[65]^. In this respect, our study may resemble prior examples, such as the reported relationship between malaria and iron deficiency^[66]^, in which the genetically predicted exposure is less reflective of exposure opportunity itself and more reflective of biologically meaningful post-infection processes, potentially making causal inference more tractable even in the setting of infectious disease MR^[64]^. Nevertheless, because anti–*H. pylori* IgG levels remain a composite phenotype and because substantial pleiotropy within the MHC region complicates causal interpretation, MR in the present study was used primarily to evaluate directional consistency rather than to infer direct causal effects. Details are provided in the **Supplementary Information**. Within this framework, we identified positive associations with autoimmune-related diseases, including type 1 diabetes, Hashimoto’s disease, pollinosis, and tuberculosis, as well as inverse associations with atopic dermatitis and anthropometric traits such as height and body weight.

Our MR results indicated significant positive associations between genetic liability to high anti–*H. pylori* IgG levels and T1D, potentially explained by molecular mimicry involving HLA molecules, a mechanism where immune responses to bacterial infections can mistakenly target self-antigens, triggering autoimmune diseases^[67]^. For Type 2 diabetes (T2D), the mechanism involving *H. pylori* infection remains unclear, although chronic inflammation and cytokine release may contribute to insulin resistance^[68]^. Our MR analysis showed weak positive associations between genetic liability to high IgG levels and elevated HbA1c levels, supporting previous findings^[69]^. Given the clinical overlap between adult-onset T1D and T2D^[70]^, the relationship between the host response to *H. pylori* and diabetes phenotypes is likely to be biologically complex. Further mechanistic studies are needed to better understand these links, as *H. pylori* eradication has already been shown to improve outcomes in a subtype of diabetes known as Type B insulin resistance syndrome^[71]^. In regard to autoimmune and allergic diseases, our MR analysis highlighted positive associations between genetic liability to high IgG levels and autoimmune thyroid diseases, such as Hashimoto’s and Graves’ disease, though the link with Graves’ disease was less robust in the MR-Egger sensitivity analyses. These results align with epidemiological studies^[8]^ and suggest potential involvement of both HLA alleles^[51]^ and the innate immune system^[72]^. In contrast, we found inverse associations between genetic liability to high IgG levels and atopic dermatitis, consistent with the hygiene hypothesis^[73]^, which posits that early microbial exposure can protect against allergic diseases. Genetic correlation analysis revealed significant local genetic correlations between anti–*H. pylori* IgG levels and some autoimmune diseases in the MHC class II region, suggesting that host genetic determinants of response to *H. pylori*—particularly those mediated through HLA class II—form part of a shared genetic architecture that is also associated with susceptibility to multiple immune-related diseases.

Although our study provides valuable insights into the complex relationships between host response to *H. pylori* and various diseases, it is essential to acknowledge the limitations of our research. Because our analysis used total anti-*H. pylori* IgG titers, it may have obscured antigen-specific associations by averaging heterogeneous antibody responses directed against distinct bacterial components, each with its own immunogenicity, kinetics, and relevance to disease pathogenesis^[74][75]^. A recent study analyzed six antigen-specific IgG traits—CagA, OMP, IgG, UREA, VacA, and Catalase—and reported distinct patterns of association; for example, OMP-specific antibodies were positively associated with Graves’ disease (OR = 1.70, P = 7.2×10^-12^)^[76]^. These findings highlight the potential for antigen-specific heterogeneity in causal inference, suggesting that total IgG levels may miss fine-grained antigen-specific patterns that could refine mechanistic interpretation of MR results. Even so, total IgG titers are interpreted not as markers of antigen-specific immunity, but as reflecting a broadly genetically regulated host response to *H. pylori*, including both variation in infection susceptibility and immune responsiveness^[17]^. Thus, our findings and those of Wang et al. are complementary rather than contradictory—their antigen-specific approach delineates mechanistic pathways that operate within the broader infection susceptibility captured by our study.

It is also important to discuss the interpretation of MR with binary exposures, such as disease status, which remains an area of ongoing methodological debate, including studies using infectious diseases as exposures^[64]^. Recent studies have emphasized the need to carefully consider what the MR estimate represents in such settings. For example, Burgess et al. have argued that MR with a binary exposure should be interpreted as estimating the causal effect of the underlying continuous risk factor or latent liability to that exposure, rather than the direct effect of the binary trait itself^[77]^. In our analysis, the exposure was not a clinically defined binary infection status per se, but genetic liability to high anti–*H. pylori* IgG levels, used as a proxy of host response to *H. pylori*. Therefore, the MR estimates should be interpreted as reflecting associations mediated through latent genetic liability—capturing variation in host susceptibility and immune responsiveness—rather than the direct effect of pathogen exposure itself.

Moreover, consistent with the methodological challenges outlined by Hamilton et al.^[64]^, further systematic validation would be warranted for excluding the possibility of residual confounding by socioeconomic factors or survival bias to determine the clinical and public health significance of these findings. Future studies should aim to clarify the underlying biological mechanisms and consider alternative methodologies, such as longitudinal studies incorporating direct measures of infection status, to better understand the relationship between *H. pylori* and health outcomes. Additionally, the application of large-scale genome sequencing techniques could facilitate a more comprehensive examination of the genetic variations associated with host response to *H. pylori*, ultimately leading to improved public health strategies regarding infection management and its potential links to disease.

In conclusion, our findings characterize the genetic architecture underlying host response to *H. pylori* and demonstrate its relationship with immunity and various diseases. The implications of these results extend beyond the realm of infectious diseases, highlighting the potential role of *H. pylori* in shaping immune responses through genetic predispositions. Clinically, this perspective suggests that individuals genetically predisposed to higher anti–*H. pylori* IgG levels may warrant consideration of immune-related multimorbidity, rather than single-disease risk alone, in clinical evaluation and risk stratification.

## Materials & Methods

### Study participants

The discovery GWAS utilized data from 56,967 individuals recruited at municipal specific health checkup sites (Type 1 cohort) in the Tohoku Medical Megabank Project Community-Based Cohort (TMM CommCohort)^[21]^. For the replication GWAS, data from 68,211 participants across the following six independent Japanese cohorts: the Type 2 cohort of the TMM CommCohort (n=18,107), the TMM Birth and Three-Generation Cohort (BirThree Cohort) (n=30,837)^[78]^, the Japan Public Health Center-based Prospective Study (JPHC) (n=5,344)^[79]^, the Japan Multi-Institutional Collaborative Cohort Study (J-MICC Study) (n=3,468)^[80]^, the Yamagata Study (n=5,266)^[81]^, and the Hospital-based Epidemiologic Research Program at Aichi Cancer Center (HERPACC) (n=5,189)^[82]^. The Type 2 cohort of the TMM CommCohort consists of 11,442 recruited at the Community Support Centers of the Tohoku Medical Megabank Organization (ToMMo) in Tohoku University, and 6,665 at the Satellites of Iwate Tohoku Medical Megabank Organization (IMM) in Iwate Medical University. The TMM cohorts, JPHC, J-MICC Study, and Yamagata Study are general population cohorts, while HERPACC is a hospital-based cohort. Details on each cohort are presented in the Supplementary Information and Supplementary Table 1. In addition, GWAS summary statistics from a previous study^[17]^ involving 15,685 individuals from an European ancestry population across seven cohorts in Europe and the USA were collected for our cross-ancestry GWAS meta-analysis. Informed consent was obtained from all study participants, and approval was granted by the institutional review board.

### Antibody titers

In all the Japanese cohorts studied, anti-*H. pylori* IgG antibody titers were measured using either ELISA (E-Plate Eiken *H pylori* antibody, Eiken Chemical Co. Ltd., Tokyo, Japan) or LIA (*H. pylori*-latex Seiken, Denka Seiken Co., Ltd., Tokyo, Japan). Due to the skewed distribution of antibody levels, the cutoff value was set at either the top 25% of the population or 3U/ml, based on previous studies^[17][83]^. Details of the cohort characteristics and method for determining the cutoff value are provided in the **Supplementary Information** and **Supplementary Table 1**.

### Genome-wide association studies (GWAS)

To ensure the quality and comparability of genotype data across cohorts, stringent quality control (QC) procedures were implemented prior to and after genotype imputation. QC for samples and SNPs was performed based on study-specific criteria. In the TMM study, samples with a call rate < 0.97, sex discrepancy, or excessive heterozygosity were excluded. Variants were filtered using thresholds of minor allele frequency (MAF) > 0.01 and Hardy–Weinberg equilibrium (HWE) P-value > 1×10^-5^. Imputation was performed with SHAPEIT (v2)^[84]^ for phasing and IMPUTE4^[85]^ using a cross-imputed reference panel combining the 3.5KJPNv2^[86]^ and the 1000 Genomes Project^[87]^ phase 3 panels. After imputation, additional QC was applied, excluding variants with an imputation INFO score < 0.4 or MAF < 0.01. In each cohort genotypic imputation for different genotyping array platforms, quality controls, and GWAS were performed separately for each dataset according to our provided protocol. Detailed information on the genotyping platforms, imputation methods, and study-specific QC thresholds for each cohort is summarized in Supplementary Table 2.

During GWAS, adjustments were made for age, sex, and the first ten principal components (PC1-10) as covariates. In the TMM cohorts, a linear mixed model was employed for the GWAS of anti-*H. pylori* IgG antibody titers using the Genome-wide Complex Trait Analysis (GCTA) software (version 1.94.0 beta)^[88]^. This approach was chosen due to the presence of a substantial number of closely related individuals (PI-HAT > 0.1875) in these cohorts. By accounting for both fixed and random effects, this method improves the accuracy of the association analysis. The GWAS results (summary statistics) from Japanese cohorts were integrated using inverse variance-weighted (IVW) meta-analysis with METAL software (version released on 2011-03-25)^[89]^. The genome-wide significance level was set at a P-value of < 5×10⁻⁸.

In the discovery GWAS, we analyzed the Type 1 cohort, consisting of 56,967 participants (14,286 cases with high anti-*H. pylori* IgG levels and 42,681 controls), of the TMM CommCohort. For the replication phase, 12 datasets from six different cohorts were compiled, involving a total of 68,211 individuals (23,124 cases and 45,087 controls). In addition to these discovery and replication GWAS phases, we performed a combined GWAS meta-analysis, comprising 125,178 individuals (37,410 cases and 87,768 controls).

### Gene-based analysis and pathway analysis

We performed a gene-based analysis on 125,178 individuals using summary statistics from our combined GWAS meta-analysis described above. The MAGMA software (v1.08)^[90]^ implemented in the FUMA platform (v1.6.1)^[27]^ was used to evaluate the aggregate effects of multiple variants within the same gene. The linkage disequilibrium (LD) reference panel was constructed from the East Asian population of the 1000 Genomes Project^[87]^. Pathway analysis was performed using the GENE2FUNC function on the FUMA platform. Briefly, genes within 10 kb of SNPs in LD with lead SNPs at each region (r² > 0.6) and with a p-value threshold of p < 0.05 were mapped using default positional settings from the set of 20,260 protein-coding genes (ENSEMBL v110^[91]^). Based on the resulting list of mapped genes, pathway analysis was performed using the Kyoto Encyclopedia of Genes and Genomes (KEGG)^[28]^ and Reactome pathway databases^[29]^. This analysis was used to visualize the characteristics of gene groups with expression variation and to identify pathways and diseases that are potentially related.

### HLA imputation and HLA association analysis

We performed association analyses on classical Human Leukocyte Antigen (HLA) genes (*HLA-A*, *-B*, *-C*, *-DRB1*, *-DQA1*, *-DQB1*, *-DPA1*, *-DPB1*) in the Major Histocompatibility Complex (MHC) region, including four-digit and two-digit HLA alleles and amino acid polymorphisms. Data was obtained from 90,009 unrelated individuals (PI-HAT < 0.1875) from the TMM cohorts. To estimate HLA alleles and amino acid polymorphisms, HLA imputation was performed using the DEEP*HLA software^[30]^, with the HapMap reference panel from the 1000 Genomes Project’s Pan-Asian dataset^[92]^.

Quality control (QC) procedures were applied both before and after imputation. For the pre-imputation variant QC, we restricted the analysis to the MHC region (chr6: 29,677,984–33,485,635, GRCh37/hg19) and retained variants meeting the following criteria: call rate > 0.97, minor allele frequency (MAF) > 0.01, and Hardy–Weinberg equilibrium (HWE) P-value > 1×10^-5^. After imputation, variants with an estimated allele frequency < 0.01 were excluded from downstream analyses. As a result, 155 classical HLA alleles and 766 amino acid polymorphisms passed these QC thresholds and were included in the final association analysis. The statistical analysis methods for HLA alleles and amino acid polymorphisms are described in detail elsewhere^[93][94]^. To identify associations with anti-*H. pylori* IgG antibody titers, logistic regression analysis, adjusted for age, sex, and the first ten principal components (PC1-10), was performed using PLINK 2.0 software (v2.00a3LM AVX2 Intel (2 Mar 2022))^[95]^.

### Fine-mapping and conditional analyses in the MHC region

Fine-mapping of association signals within the MHC region was performed using FINEMAP (v1.4.1)^[96]^ to account for the complex LD structure of this region. Summary statistics (β, standard error, and effective sample size) from the combined meta-analysis were used as input, and linkage disequilibrium (LD) was estimated using a reference panel from the East Asian population of the 1000 Genomes Project^[87]^. FINEMAP was run assuming up to five causal variants per region (--n-causal-snps 5) and a normal prior for effect sizes with a standard deviation of 0.05 (--prior-std 0.05). The number of causal variants per locus and their posterior inclusion probabilities (PIPs) were inferred under a Bayesian model. Variants with high PIPs were regarded as credible candidate variants underlying the association signals.

Conditional analyses were then performed using GCTA^[88]^. A linear mixed model was fitted while adjusting for age, sex, and the first ten principal components (PC1–10), with the variant identified by fine-mapping as a putative causal signal (rs1047989) included as an additional covariate. In parallel, HLA allele–level conditional analyses were conducted using genotype data imputed with DEEP*HLA^[30]^. These logistic regression models were implemented in PLINK 2.0^[95]^, following the same framework as the primary HLA association analyses. The association tests were repeated with rs1047989 added as a covariate to assess whether any residual associations were attributable to independent HLA allelic polymorphisms.

### Cross-ancestry meta-analysis

To explore the genetic architecture underlying host responses to *H. pylori*, across different ancestral groups, we performed a cross-ancestry GWAS meta-analysis utilizing GWAS summary statistics from 15,685 individuals of European ancestry, analyzed in a previous GWAS study[17] along with our GWAS summary statistics from 125,178 individuals of Japanese ancestry. MR-MEGA software (v.0.2)^[32]^ was used for the cross-ancestry GWAS meta-analysis, accounting for differences in allelic effects on disease risk between populations. Heterogeneity between GWAS summary statistics from different ancestral populations was assessed using Cochran’s Q and I^2^ statistic for each variant, with significant heterogeneity defined as a Q test P-value < 5×10⁻⁸.

### Mendelian randomization analysis and genetic correlation analysis

To assess the directionally consistent associations between genetic liability to high anti–*H. pylori* IgG levels and other diseases, Mendelian Randomization (MR) analyses were performed using the TwoSample MR (version 0.5.7) package^[97]^. For outcome traits, we used GWAS summary statistics for 28 diseases and 14 quantitative traits with potential immunological or epidemiological relevance to anti–*H. pylori* IgG levels, primarily obtained from the BioBank Japan (BBJ) PheWeb resource^[33]^; data sources for individual outcomes are detailed in **Supplementary Table 12**. The primary approach employed was Inverse variance-weighted (IVW)^[34]^, with weighted median^[35]^, weighted mode^[36]^, and MR-Egger^[37]^ used as sensitivity analysis methods. In addition, we applied MR-cML^[38]^, a likelihood-based method that does not rely on the InSIDE assumption and is robust to both outlier and correlated pleiotropy, to further evaluate the robustness and directional stability of the observed associations. Evidence for a robust association was defined when the direction of effect was concordant across multiple MR methods, with nominal statistical support (P-value < 0.05) in the primary IVW analysis.

Independent instrumental variables (IVs) were selected from the combined GWAS meta-analysis (N = 125,178) based on the following criteria: (i) genome-wide significance (P-value < 5×10⁻⁸); (ii) independence defined by linkage disequilibrium (LD) clumping (r² < 0.05 within a 10 Mb window); and (iii) sufficient instrument strength indicated by F-statistics greater than 20 (F = β² / SE²). SNPs meeting these criteria were harmonized with outcome GWAS summary statistics, with allele orientation aligned using allele frequency information. Palindromic variants with ambiguous strand orientation were excluded. The number of IVs varied across outcomes. A complete list of IVs, including effect sizes, standard errors, and F-statistics, is provided in **Supplementary Table 16**.

To assess the possibility of instrumental variables violating the underlying assumptions, we performed three tests. First, if the heterogeneity statistics were significantly high, we reviewed the results using an IVW model with multiplicative random effects, which is considered effective for estimating causal relationships while accounting for data heterogeneity^[98]^. To test for horizontal pleiotropy, we performed the MR-Egger pleiotropy test and assessed bias by evaluating the intercept. Furthermore, to determine whether the association between exposure and outcome was driven by a single SNP, we performed a leave-one-out sensitivity analysis to identify and exclude such instances. In the genetic correlation analysis, both global and local genetic correlations (global r_g_ and local r_g_) were computed to further characterize the shared genetic architectures. To calculate local rg, autosomal regions were divided into approximately 1,500 smaller regions (median size of 2.2 Mb, ranging from 85.4 kb to 31.3 Mb) using a method designed to efficiently identify nearly independent linkage disequilibrium (LD) blocks^[99]^. In the MHC class II region, the boundaries of these smaller regions occasionally overlapped with gene coding sequences, leading to the consolidation of multiple smaller regions in some cases (**Supplementary Table 15**). The LD Score Regression (LDSC) tool (Version 1.0.1)^[100]^ and the Local Analysis of [co]Variant Association (LAVA) tool (v0.1.0)^[101]^ were used for the analysis, employing a reference panel from the East Asian population of the 1000 Genomes Project^[87]^ to estimate LD.

## Supporting information

Supplementary Information (including Supplementary Figures 1-12)

## Author contributions

G.T. supervised this project. T.Kyosaka., A.N., M.M., and G.T. designed the study. A.K.K.M., A.M., T.F., Y.Kotsar., K.H., S.M., M.S., C.G., S.Sugimoto., and J.T. contributed to bioinformatics analyses. A.Hozawa., K.K., M.Y., and S.K. contributed to the management of data from ToMMo. Y.Sutoh., Y.Y., K.T., and A.S. contributed to the management of data from IMM. O.N., Y.A., Y.Sasaki., and Y.U. contributed to the management of data from Yamagata Study. T.Y. contributed to bioinformatics analyses of data from JPHC. N.S., S.Nakano., and M.I. contributed to the management of data from JPHC. M.N. contributed to bioinformatics analyses of data from J-MICC Study and HERPACC. S.Suzuki., A.Hishida., T.Koyama., Y.Kubo., and K.W. contributed to the management of data from J-MICC Study. Y.N.K., Y.Kasugai., H.I., and K.M. contributed to the management of data from HERPACC. S.Namba., T.O., and Y.O. contributed to the management of data from BBJ. G.M.F. and G.H. coordinated the collaboration with co-authors from the European-ancestry study group. L.B., M.C.W.S., M.P.P., M.M.L., F.F., S.W., S.-J.H., D.L., A.C.W., S.S.R., J.I.R., K.D.T., R.P.T., H.S., H.B., M.L., and R.P. contributed to data curation and analysis in the European study. T.Kyosaka. wrote the manuscript with critical input from A.N., J.K.K., Y.O., M.M., and G.T. All authors provided critical revision of the manuscript.

## Acknowledgements

We would like to thank all participants and investigators involved in TMM, Yamagata Study, JPHC, J-MICC Study, HERPACC, and BBJ. We also appreciate the contributions of participants and researchers from previous GWAS studies on populations of European descent^[17]^, including the Rotterdam Study (RS), the Study of Health in Pomerania (SHIP), the Framingham Heart Study (FHS), and the Multi-Ethnic Study of Atherosclerosis (MESA).

Special thanks to Barry J. Marshall, Alfred Chin Yen Tay, and Roger L. Dawkins for their insightful guidance and valuable suggestions. We are also grateful to Michio Shimabukuro and Hayato Tanabe for their constructive feedback and support. We also thank Hideki Katagiri, Toru Furukawa, Tetsuya Niihori, and Kaname Kojima for their insightful advice. Additionally, we thank Tatsuhiko Naito for his advice on DEEP*HLA^[30]^ and appreciate the valuable advice provided by Sun Ha Jee. We also acknowledge the contributions of Ben Schöttker and Bernd Holleczek to the European study group. We thank all the contributors to the J-MICC study. Contributors to the J-MICC study are listed at the following site. (as of Mar 2023): https://jmicc.com/en/contributors.

## Funding

This work was supported by the Japan Agency for Medical Research and Development (AMED) (JP23ek0410113) to Y.O. and G.T., Programs for Bridging the gap between R&D and the IDeal society (society 5.0) and Generating Economic and social value (BRIDGE) (JPJ012525) to G.T., the Japan Society for the Promotion of Science (JSPS) KAKENHI (25K00278 and 24K13523) to T.Kyosaka and A.M., Japan Science and Technology Agency (JST) Moonshot R&D (JPMJMS2023) to J.T. and G.T., the RIKEN Center for Advanced Intelligence Project “Genome Statistical Analysis Using Machine Learning and Artificial Intelligence Technologies” to G.T., and the Tohoku Kaihatsu Memorial Foundation to T.Kyosaka and A.M. This work was also supported by the National Cancer Center Research and Development Fund (28-A-19, 31-A-18, and 2022-A-20). The TMM project’s supercomputer and integrated database dbTMM were supported by AMED grant numbers JP21tm0424601 and JP21tm0124005, respectively. The HERPACC Study was supported by Grants-in-Aid for Scientific Research from the Ministry of Education, Culture, Sports, Science and Technology (MEXT) (Priority Areas of Cancer: 17015018; Innovative Areas: 221S0001) and JSPS KAKENHI (JP16H06277[CoBiA], JP26253041, JP20K10463, JP23K16316, and JP24K02697), as well as a Grant-in-Aid for the Third Term Comprehensive 10-year Strategy for Cancer Control from the Ministry of Health, Labour and Welfare (MHLW) of Japan. The JPHC Study was supported by the National Cancer Center Research and Development Fund (23-A-31[toku], 26-A-2, 29-A-4, 2020-J-4, and 2023-J-4), and a Grants-in-Aid for Cancer Research from MHLW of Japan (from 1989 to 2010). The J-MICC Study was supported by Grants-in-Aid for Scientific Research from MEXT (Priority Areas of Cancer: 17015018; Innovative Areas: 221S0001) and JSPS KAKENHI (JP16H06277 and JP22H04923[CoBiA]). It was also partially funded by the BBJ Project from AMED (since April 2015) and by MEXT (from April 2003 to March 2015). The Yamagata Study was supported by AMED (20ck0106561h0001 and 24ck0106887h0002) and JSPS KAKENHI (JP23K07454).

## Competing interests

The authors declare no competing interests.

## Data availability

Summary statistics for the GWAS of anti-*H. pylori* IgG antibody titers generated in this study will be made publicly available through the jMorp database (https://jmorp.megabank.tohoku.ac.jp/) upon publication. Most of the GWAS summary statistics used as outcome datasets in the Mendelian randomization analyses are available through BioBank Japan PheWeb (https://pheweb.jp/), whereas the remaining summary statistics are derived from controlled-access datasets governed by cohort-specific ethical and institutional restrictions and can be accessed upon reasonable request to the respective data access committees. European ancestry GWAS summary statistics for anti-*H. pylori* IgG antibody titers from Lam et al. (2022) are not publicly available and were obtained under a data-sharing agreement; access must be requested directly from the original authors. Individual-level genotype and phenotype data from the participating Japanese cohorts cannot be publicly shared due to privacy, ethical, and regulatory restrictions. All remaining data supporting the findings of this study are available within the article and its Supplementary Information. Source Data are provided with this paper.

## Abbreviations

BBJ: BioBank Japan
CCDC80: Coiled-coil domain containing 80
GWAS: genome-wide association study
FHS: Framingham Heart Study
HERPACC: Hospital-based Epidemiologic Research Program at Aichi Cancer Center
HLA: human leukocyte antigen
IMM: Iwate Tohoku Medical Megabank organization
IVW: inverse-variance weighted
J-MICC: Japan Multi-Institutional Collaborative Cohort Study
JPHC: Japan Public Health Centre-based Prospective Study
LD: linkage disequilibrium
MAF: minor allele frequency
MESA: Multi-Ethnic Study of Atherosclerosis
MHC: major histocompatibility complex
MR: Mendelian randomization
NFKBIZ: NF-kappa-B inhibitor zeta
NF-κB: nuclear factor-kappa B
OR: odds ratio
PC: principal component
PSCA: prostate stem cell antigen
r_g_: genetic correlation
RS: Rotterdam Study
SHIP: Study of Health in Pomerania
SNP: single nucleotide polymorphism
T1D: Type 1 diabetes
T2D: Type 2 diabetes
TIFA: TRAF interacting protein with forkhead associated domain
TLR: Toll-Like receptor
TRAF3: Tumor necrosis factor receptor-associated factor 3
TMM: Tohoku Medical Megabank
ToMMo: Tohoku Medical Megabank Organization (Miyagi prefecture)

## Tables and Supplementary tables

**Supplementary Table 1.**
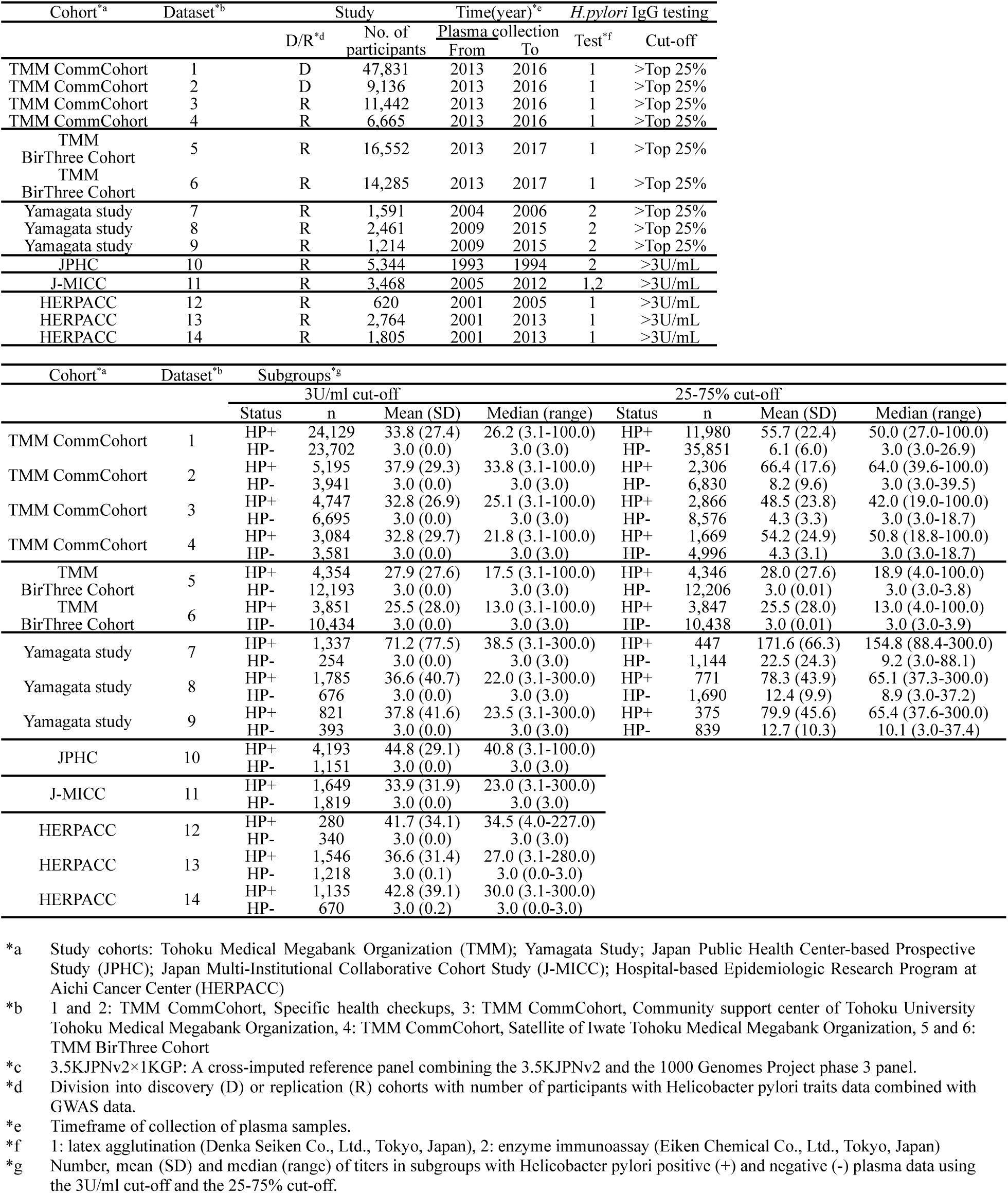
Characteristics of the study population.

**Supplementary Table 2.** Cohort-specific information on genotyping, imputation and GWAS.

| Cohort<sup>a</sup> | Dataset<sup>b</sup> | Genotyping |  |
| --- | --- | --- | --- |
|  |  | Genotyping array | Calling algorithm |
| TMM CommCohort | 1 | Japonica Array version 2 | Affymetrix Power Tools |
| TMM CommCohort | 2 | Japonica Array NEO | Affymetrix Power Tools |
| TMM CommCohort | 3 | Japonica Array version 2 | Affymetrix Power Tools |
| TMM CommCohort | 4 | Japonica Array version 2 | Affymetrix Power Tools |
| TMM<br>BirThree Cohort | 5 | Japonica Array version 2 | Affymetrix Power Tools |
| TMM<br>BirThree Cohort | 6 | Japonica Array NEO | Affymetrix Power Tools |
| Yamagata study | 7 | Human660W Quad | GenomeStudio |
| Yamagata study | 8 | Japonica Array NEO | Affymetrix Power Tools |
| Yamagata study | 9 | HumanOmniExpressExome | GenomeStudio |
| JPHC | 10 | HumanOmniExpressExome<br>BeadChip | GenomeStudio |
| J-MICC | 11 | HumanOmniExpressExome | GenomeStudio |
| HERPACC | 12 | Human610-Quad | GenomeStudio |
| HERPACC | 13 | Infinium Asian Screening | GenomeStudio |
| HERPACC | 14 | HumanCoreExome | GenomeStudio |

| Cohort<sup>a</sup> | Dataset<sup>b</sup> | Preimputation QC |  |  |  |  |
| --- | --- | --- | --- | --- | --- | --- |
|  |  | Sample<br>call rate cutoff | other criteria | SNP<br>call rate cutoff | SNP<br>MAF cutoff | SNP<br>HWE P cutoff |
| TMM CommCohort | 1 | 0.97 | history of gastric cancer,<br>sex mismatches | 0.99 | 0.01 | $1 \times 10^{-5}$ |
| TMM CommCohort | 2 | 0.97 | history of gastric cancer,<br>sex mismatches | 0.99 | 0.01 | $1 \times 10^{-5}$ |
| TMM CommCohort | 3 | 0.97 | history of gastric cancer,<br>sex mismatches | 0.99 | 0.01 | $1 \times 10^{-5}$ |
| TMM CommCohort | 4 | 0.97 | history of gastric cancer,<br>sex mismatches | 0.99 | 0.01 | $1 \times 10^{-5}$ |
| TMM<br>BirThree Cohort | 5 | 0.97 | history of gastric cancer,<br>sex mismatches | 0.99 | 0.01 | $1 \times 10^{-5}$ |
| TMM<br>BirThree Cohort | 6 | 0.97 | history of gastric cancer,<br>sex mismatches | 0.99 | 0.01 | $1 \times 10^{-5}$ |
| Yamagata study | 7 | 0.97 | history of gastric cancer,<br>sex mismatches | 0.99 | 0.01 | $1 \times 10^{-5}$ |
| Yamagata study | 8 | 0.97 | history of gastric cancer,<br>sex mismatches | 0.99 | 0.01 | $1 \times 10^{-5}$ |
| Yamagata study | 9 | 0.97 | history of gastric cancer,<br>sex mismatches | 0.99 | 0.01 | $1 \times 10^{-5}$ |
| JPHC | 10 | 0.98 | history of gastric cancer,<br>sex mismatches,<br>related samples (IBD 0.1875) | 0.99 | 0.01 | $1 \times 10^{-6}$ |
| J-MICC | 11 | 0.98 | history of gastric cancer,<br>sex mismatches,<br>related samples (IBD 0.1875) | 0.98 | 0.01 | $1 \times 10^{-6}$ |
| HERPACC | 12 | 0.98 | history of gastric cancer,<br>sex mismatches,<br>related samples (IBD 0.1875) | 0.98 | 0.01 | $1 \times 10^{-6}$ |
| HERPACC | 13 | 0.95 | history of gastric cancer,<br>sex mismatches,<br>related samples (IBD 0.1875) | 0.98 | 0.01 | $1 \times 10^{-6}$ |
| HERPACC | 14 | 0.98 | history of gastric cancer,<br>sex mismatches,<br>related samples (IBD 0.1875) | 0.98 | 0.01 | $1 \times 10^{-6}$ |

| Cohort<sup>*a</sup> | Dataset<sup>*b</sup> | Imputation |  | Post-imputation QC |  | GWAS |
| --- | --- | --- | --- | --- | --- | --- |
|  |  | Reference<sup>*c</sup> | Software | SNP info cutoff | SNP MAF cutoff | Software |
| TMM CommCohort | 1 | 3.5KJPNv2×1KGP | SHAPEIT2 & IMPUTE4 | 0.4 | 0.01 | GCTA |
| TMM CommCohort | 2 | 3.5KJPNv2×1KGP | SHAPEIT2 & IMPUTE4 | 0.4 | 0.01 | GCTA |
| TMM CommCohort | 3 | 3.5KJPNv2×1KGP | SHAPEIT2 & IMPUTE4 | 0.4 | 0.01 | GCTA |
| TMM CommCohort | 4 | 3.5KJPNv2×1KGP | SHAPEIT2 & IMPUTE4 | 0.4 | 0.01 | GCTA |
| TMM BirThree Cohort | 5 | 3.5KJPNv2×1KGP | SHAPEIT2 & IMPUTE4 | 0.4 | 0.01 | GCTA |
| TMM BirThree Cohort | 6 | 3.5KJPNv2×1KGP | SHAPEIT2 & IMPUTE4 | 0.4 | 0.01 | GCTA |
| Yamagata study | 7 | 1000 Genomes Project phase 3 | SHAPEIT2 & IMPUTE4 | 0.4 | 0.01 | GCTA |
| Yamagata study | 8 | 3.5KJPNv2×1KGP | SHAPEIT2 & IMPUTE4 | 0.4 | 0.01 | GCTA |
| Yamagata study | 9 | 1000 Genomes Project phase 3 | SHAPEIT2 & IMPUTE4 | 0.4 | 0.01 | GCTA |
| JPHC | 10 | 1000 Genomes Project phase 3 | SHAPEIT2 & IMPUTE2 | NA | 0.01 | SNPTEST |
| J-MICC | 11 | 1000 Genomes Project phase 3 | SHAPEIT2 & IMPUTE2 | 0.4 | 0.01 | REGENIE |
| HERPACC | 12 | 1000 Genomes Project phase 3 | SHAPEIT2 & minimac3 | 0.4 | 0.01 | REGENIE |
| HERPACC | 13 | 1000 Genomes Project phase 3 | SHAPEIT2 & minimac3 | 0.4 | 0.01 | REGENIE |
| HERPACC | 14 | 1000 Genomes Project phase 3 | SHAPEIT2 & minimac3 | 0.4 | 0.01 | REGENIE |

<sup>\*a</sup> Study cohorts: Tohoku Medical Megabank Organization (TMM); Yamagata Study; Japan Public Health Center-based Prospective Study (JPHC); Japan Multi-Institutional Collaborative Cohort Study (J-MICC); Hospital-based Epidemiologic Research Program at Aichi Cancer Center (HERPACC)
<sup>\*b</sup> 1 and 2: TMM CommCohort, Specific health checkups, 3: TMM CommCohort, Community support center of Tohoku University Tohoku Medical Megabank Organization, 4: TMM CommCohort, Satellite of Iwate Tohoku Medical Megabank Organization, 5 and 6: TMM BirThree Cohort
<sup>\*c</sup> 3.5KJPNv2×1KGP: A cross-imputed reference panel combining the 3.5KJPNv2 and the 1000 Genomes Project phase 3 panel.

**Supplementary Table 3.**
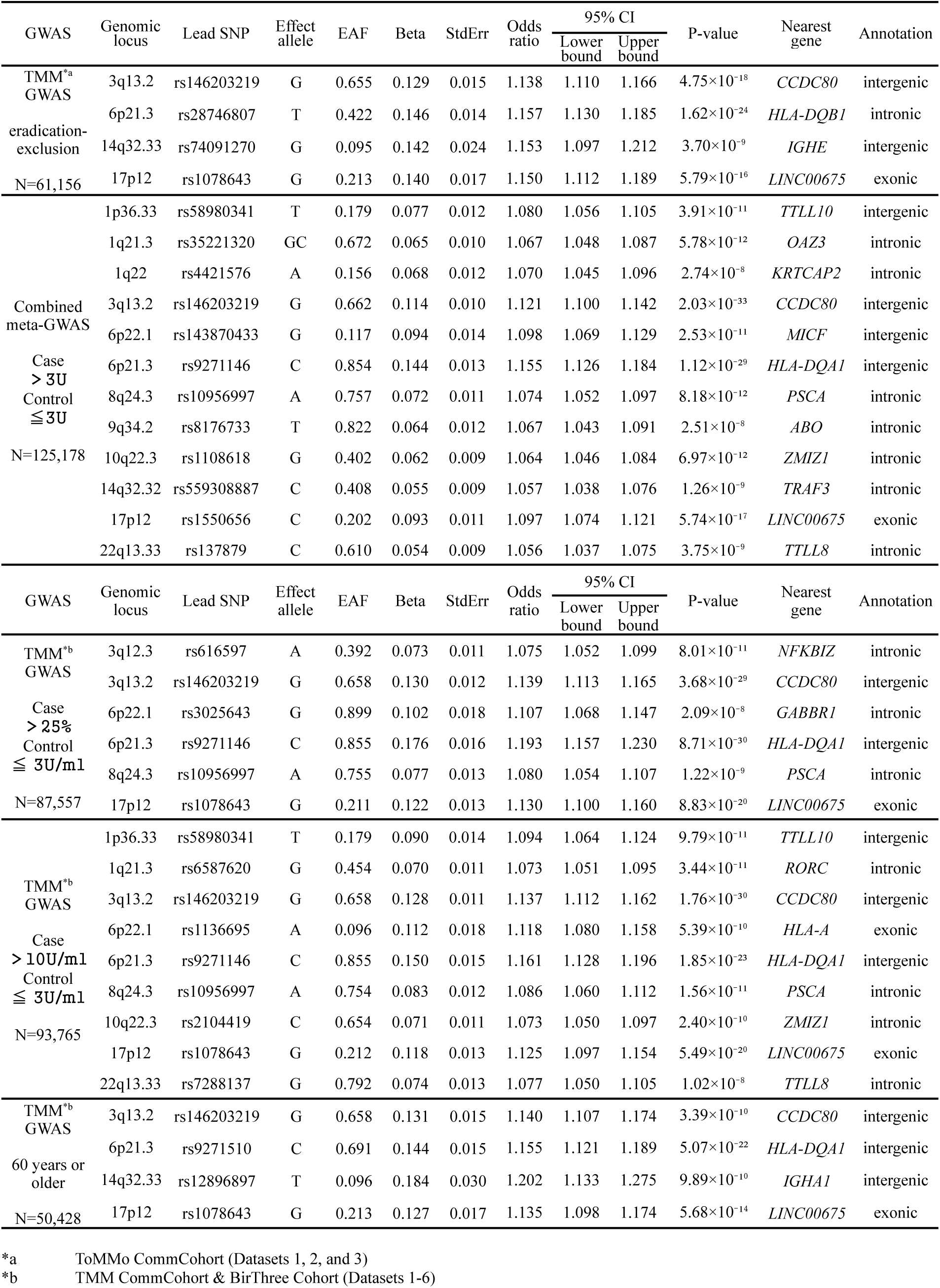
GWAS sensitivity analyses: A list of regions reaching genome-wide significance.

**Supplementary Table 4.** Gene-based Analysis. A list of 19 genes that meet the significance level set by the Bonferroni correction (P-value < 0.05/19,276 = 2.59×10 ⁻⁶). Base pair (BP) positions are referenced to the GRCh37 (hg19) genome assembly.

| Gene | CHR | BP: Start | BP: Stop | N of SNPs | Z statistics | P-value |
| --- | --- | --- | --- | --- | --- | --- |
| *HLA-DQA1* | 6 | 32595956 | 32614839 | 1917 | 7.9382 | $1.03 \times 10^{-15}$ |
| *HLA-DQB1* | 6 | 32627244 | 32636160 | 1021 | 7.0585 | $8.42 \times 10^{-13}$ |
| *TAP2* | 6 | 32781544 | 32806599 | 261 | 7.0149 | $1.15 \times 10^{-12}$ |
| *HLA-DPB1* | 6 | 33043703 | 33054978 | 376 | 6.7472 | $7.54 \times 10^{-12}$ |
| *ITPR3* | 6 | 33588142 | 33664351 | 540 | 6.3395 | $1.15 \times 10^{-10}$ |
| *HLA-DOB* | 6 | 32780540 | 32784825 | 54 | 6.0296 | $8.22 \times 10^{-10}$ |
| *TRAF3* | 14 | 103243813 | 103377837 | 551 | 5.8139 | $3.05 \times 10^{-9}$ |
| *BAK1* | 6 | 33540329 | 33548019 | 53 | 5.738 | $4.79 \times 10^{-9}$ |
| *CCDC80* | 3 | 112323407 | 112368377 | 222 | 5.5094 | $1.80 \times 10^{-8}$ |
| *DPCR1* | 6 | 30908749 | 30921998 | 72 | 5.3624 | $4.11 \times 10^{-8}$ |
| *HLA-DPA1* | 6 | 33032346 | 33048552 | 509 | 5.3405 | $4.64 \times 10^{-8}$ |
| *HLA-DRA* | 6 | 32407619 | 32412823 | 81 | 5.2986 | $5.83 \times 10^{-8}$ |
| *HLA-DRB1* | 6 | 32546546 | 32557625 | 801 | 5.0894 | $1.80 \times 10^{-7}$ |
| *HLA-C* | 6 | 31236526 | 31239907 | 221 | 4.8971 | $4.86 \times 10^{-7}$ |
| *NFKBIZ* | 3 | 101546835 | 101579866 | 91 | 4.6582 | $1.59 \times 10^{-6}$ |
| *FKBPL* | 6 | 32096484 | 32098068 | 7 | 4.6187 | $1.93 \times 10^{-6}$ |
| *TIFA* | 4 | 113195698 | 113207059 | 73 | 4.5907 | $2.21 \times 10^{-6}$ |
| *PSCA* | 8 | 143751726 | 143764142 | 96 | 4.5718 | $2.42 \times 10^{-6}$ |
| *CCHCR1* | 6 | 31110216 | 31126015 | 203 | 4.5651 | $2.50 \times 10^{-6}$ |

**Supplementary Table 5.**
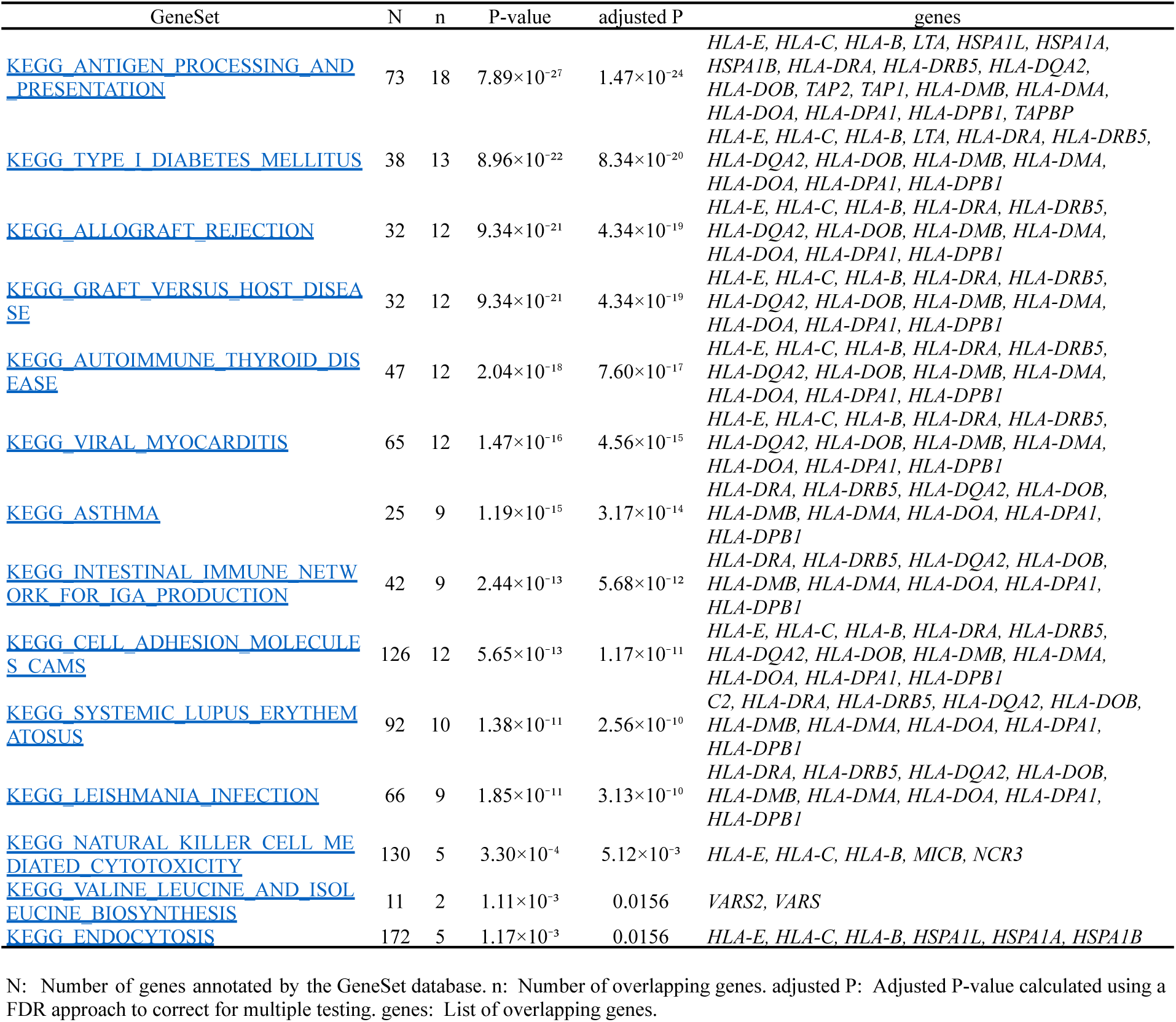
Pathway analysis using the KEGG database.

**Supplementary Table 6.**
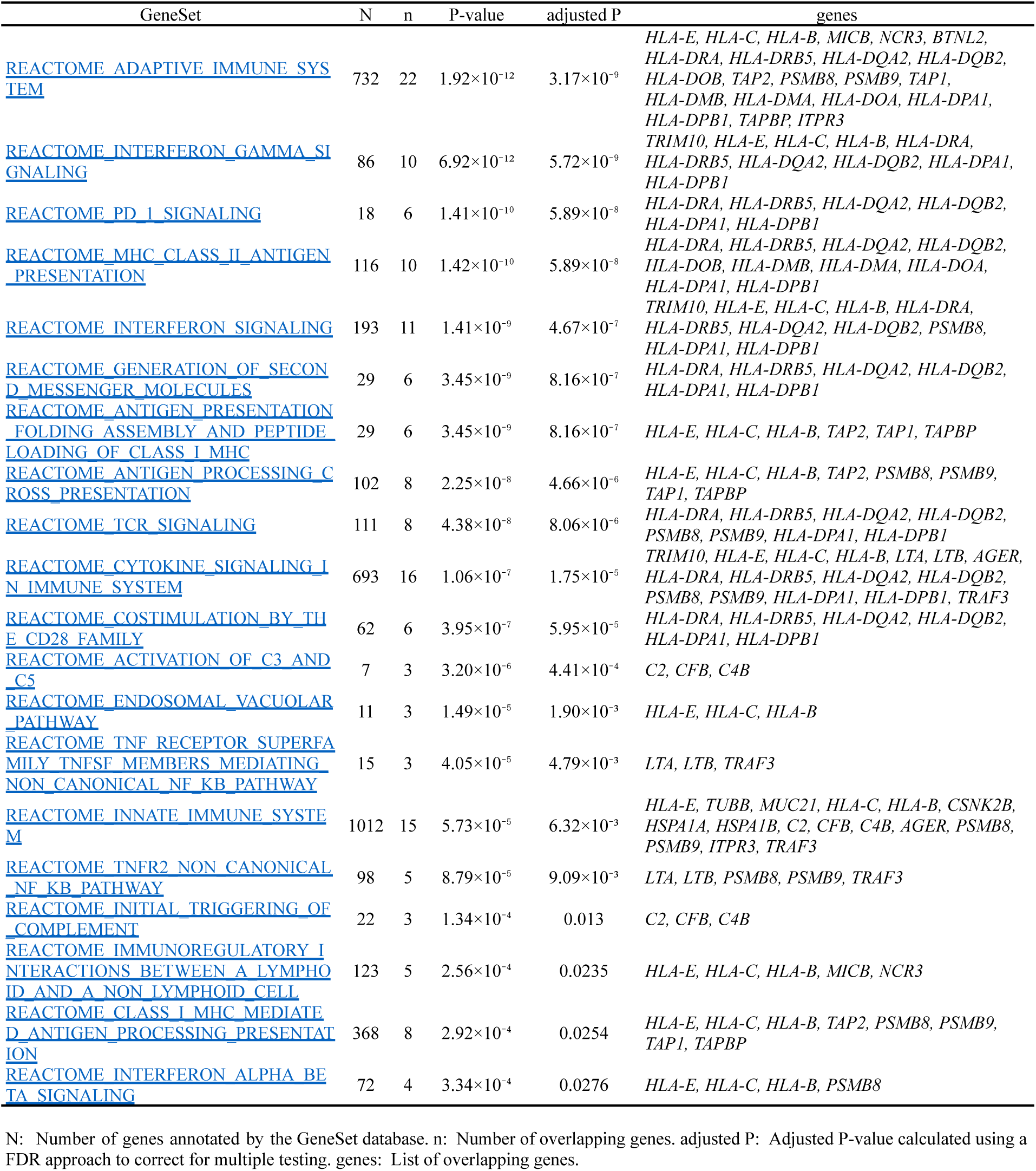
Pathway analysis using the Reactome database.

**Supplementary Table 7.** HLA association analysis (amino acid polymorphisms). Amino acid polymorphisms that meet the significance level of the Bonferroni correction (P < 0.05/766 = 6.53×10 ⁻⁵).

| Protein | Position | Amino-acid | Freq (Case) | Freq (Control) | Beta | StdErr | Odds ratio | 95% CI |  | P-value |
| --- | --- | --- | --- | --- | --- | --- | --- | --- | --- | --- |
|  |  |  |  |  |  |  |  | Lower bound | Upper bound |  |
| DRB1 | 104 | A | 0.380 | 0.356 | 0.106 | 0.011 | 1.112 | 1.088 | 1.136 | $3.71 \times 10^{-21}$ |
| DRB1 | 98 | E | 0.380 | 0.356 | 0.106 | 0.011 | 1.112 | 1.088 | 1.136 | $3.71 \times 10^{-21}$ |
| DRB1 | 98 | K | 0.620 | 0.644 | -0.106 | 0.011 | 0.899 | 0.880 | 0.919 | $3.73 \times 10^{-21}$ |
| DRB1 | 104 | S | 0.620 | 0.644 | -0.106 | 0.011 | 0.899 | 0.880 | 0.919 | $3.73 \times 10^{-21}$ |
| DQA1 | 215 | L | 0.374 | 0.350 | 0.105 | 0.011 | 1.111 | 1.087 | 1.135 | $8.66 \times 10^{-21}$ |
| DQA1 | 50 | L | 0.374 | 0.350 | 0.105 | 0.011 | 1.111 | 1.087 | 1.135 | $8.66 \times 10^{-21}$ |
| DQA1 | 53 | R | 0.374 | 0.350 | 0.105 | 0.011 | 1.111 | 1.087 | 1.135 | $8.66 \times 10^{-21}$ |
| DQA1 | 215 | F | 0.626 | 0.650 | -0.105 | 0.011 | 0.900 | 0.881 | 0.920 | $8.67 \times 10^{-21}$ |
| DQA1 | 26 | T | 0.632 | 0.656 | -0.104 | 0.011 | 0.901 | 0.881 | 0.921 | $2.06 \times 10^{-20}$ |
| DQA1 | 26 | S | 0.368 | 0.344 | 0.104 | 0.011 | 1.110 | 1.086 | 1.135 | $2.06 \times 10^{-20}$ |
| DQA1 | 47 | Q | 0.368 | 0.344 | 0.104 | 0.011 | 1.110 | 1.086 | 1.134 | $2.06 \times 10^{-20}$ |
| DQA1 | 56 | R | 0.368 | 0.344 | 0.104 | 0.011 | 1.110 | 1.086 | 1.134 | $2.06 \times 10^{-20}$ |
| DQA1 | 187 | T | 0.368 | 0.344 | 0.104 | 0.011 | 1.110 | 1.086 | 1.134 | $2.06 \times 10^{-20}$ |
| DQA1 | 76 | V | 0.368 | 0.344 | 0.104 | 0.011 | 1.110 | 1.086 | 1.134 | $2.06 \times 10^{-20}$ |
| DQA1 | 187 | A | 0.632 | 0.656 | -0.104 | 0.011 | 0.901 | 0.882 | 0.921 | $2.06 \times 10^{-20}$ |
| DQA1 | 69 | L | 0.419 | 0.394 | 0.101 | 0.011 | 1.106 | 1.083 | 1.130 | $5.42 \times 10^{-20}$ |
| DQA1 | -16 | M | 0.419 | 0.394 | 0.101 | 0.011 | 1.106 | 1.083 | 1.130 | $5.42 \times 10^{-20}$ |
| DQA1 | -16 | L | 0.581 | 0.606 | -0.101 | 0.011 | 0.904 | 0.885 | 0.924 | $5.42 \times 10^{-20}$ |
| DRB1 | -17 | T | 0.298 | 0.321 | -0.107 | 0.012 | 0.899 | 0.878 | 0.920 | $6.52 \times 10^{-20}$ |
| DQB1 | 185 | T | 0.554 | 0.578 | -0.099 | 0.011 | 0.906 | 0.886 | 0.925 | $1.64 \times 10^{-19}$ |
| DQB1 | 185 | I | 0.446 | 0.421 | 0.099 | 0.011 | 1.104 | 1.081 | 1.128 | $1.78 \times 10^{-19}$ |
| DRB1 | -17 | A | 0.691 | 0.668 | 0.104 | 0.012 | 1.110 | 1.084 | 1.136 | $5.72 \times 10^{-19}$ |
| DRB1 | 9 | W | 0.309 | 0.331 | -0.103 | 0.012 | 0.902 | 0.881 | 0.924 | $1.08 \times 10^{-18}$ |
| DQB1 | 221 | Q | 0.459 | 0.483 | -0.095 | 0.011 | 0.909 | 0.890 | 0.929 | $2.83 \times 10^{-18}$ |
| DQB1 | 220 | R | 0.459 | 0.483 | -0.095 | 0.011 | 0.909 | 0.890 | 0.929 | $2.83 \times 10^{-18}$ |
| DQB1 | 53 | L | 0.541 | 0.517 | 0.095 | 0.011 | 1.100 | 1.076 | 1.123 | $3.12 \times 10^{-18}$ |
| DQB1 | 53 | Q | 0.459 | 0.483 | -0.095 | 0.011 | 0.909 | 0.890 | 0.929 | $3.12 \times 10^{-18}$ |
| DQB1 | 90 | I | 0.459 | 0.483 | -0.095 | 0.011 | 0.909 | 0.890 | 0.929 | $3.12 \times 10^{-18}$ |
| DQB1 | 89 | G | 0.459 | 0.483 | -0.095 | 0.011 | 0.909 | 0.890 | 0.929 | $3.12 \times 10^{-18}$ |
| DQB1 | 85 | V | 0.459 | 0.483 | -0.095 | 0.011 | 0.909 | 0.890 | 0.929 | $3.12 \times 10^{-18}$ |
| DQB1 | 84 | E | 0.459 | 0.483 | -0.095 | 0.011 | 0.909 | 0.890 | 0.929 | $3.12 \times 10^{-18}$ |
| DQB1 | 87 | L | 0.541 | 0.517 | 0.095 | 0.011 | 1.100 | 1.076 | 1.124 | $3.13 \times 10^{-18}$ |
| DQB1 | 84 | Q | 0.541 | 0.517 | 0.095 | 0.011 | 1.100 | 1.076 | 1.124 | $3.13 \times 10^{-18}$ |
| DQB1 | 90 | T | 0.541 | 0.517 | 0.095 | 0.011 | 1.100 | 1.076 | 1.124 | $3.13 \times 10^{-18}$ |
| DQB1 | 85 | L | 0.541 | 0.517 | 0.095 | 0.011 | 1.100 | 1.076 | 1.124 | $3.13 \times 10^{-18}$ |
| DQB1 | 221 | H | 0.541 | 0.517 | 0.095 | 0.011 | 1.100 | 1.076 | 1.124 | $3.13 \times 10^{-18}$ |
| DQB1 | 89 | T | 0.541 | 0.517 | 0.095 | 0.011 | 1.100 | 1.076 | 1.124 | $3.13 \times 10^{-18}$ |
| DQB1 | 86 | E | 0.541 | 0.517 | 0.095 | 0.011 | 1.100 | 1.076 | 1.124 | $3.13 \times 10^{-18}$ |
| DQB1 | 125 | A | 0.541 | 0.517 | 0.095 | 0.011 | 1.100 | 1.076 | 1.124 | $3.13 \times 10^{-18}$ |
| DQB1 | 220 | H | 0.541 | 0.517 | 0.095 | 0.011 | 1.100 | 1.076 | 1.124 | $3.13 \times 10^{-18}$ |
| DQA1 | 175 | E | 0.422 | 0.399 | 0.095 | 0.011 | 1.100 | 1.076 | 1.124 | $4.05 \times 10^{-18}$ |
| DQA1 | 218 | R | 0.466 | 0.442 | 0.093 | 0.011 | 1.097 | 1.074 | 1.121 | $9.34 \times 10^{-18}$ |
| DQA1 | 56 | G | 0.534 | 0.558 | -0.093 | 0.011 | 0.911 | 0.892 | 0.931 | $9.34 \times 10^{-18}$ |
| DQA1 | 175 | Q | 0.534 | 0.558 | -0.093 | 0.011 | 0.911 | 0.892 | 0.931 | $9.34 \times 10^{-18}$ |
| DQA1 | 76 | M | 0.534 | 0.558 | -0.093 | 0.011 | 0.911 | 0.892 | 0.931 | $9.34 \times 10^{-18}$ |
| DQA1 | 50 | E | 0.534 | 0.558 | -0.093 | 0.011 | 0.911 | 0.892 | 0.931 | $9.34 \times 10^{-18}$ |
| DQA1 | 53 | K | 0.534 | 0.558 | -0.093 | 0.011 | 0.911 | 0.892 | 0.931 | $9.34 \times 10^{-18}$ |
| DQA1 | 52 | S | 0.534 | 0.558 | -0.093 | 0.011 | 0.911 | 0.892 | 0.931 | $9.34 \times 10^{-18}$ |
| DQA1 | 47 | R | 0.534 | 0.558 | -0.093 | 0.011 | 0.911 | 0.892 | 0.931 | $9.34 \times 10^{-18}$ |
| DQA1 | 69 | A | 0.534 | 0.558 | -0.093 | 0.011 | 0.911 | 0.892 | 0.931 | $9.34 \times 10^{-18}$ |
| DQA1 | 218 | Q | 0.534 | 0.558 | -0.093 | 0.011 | 0.911 | 0.892 | 0.931 | $9.34 \times 10^{-18}$ |
| DQA1 | 45 | A | 0.534 | 0.558 | -0.093 | 0.011 | 0.911 | 0.892 | 0.931 | $9.34 \times 10^{-18}$ |
| DQA1 | 55 | G | 0.534 | 0.558 | -0.093 | 0.011 | 0.911 | 0.892 | 0.931 | $9.34 \times 10^{-18}$ |
| DQA1 | 66 | I | 0.466 | 0.442 | 0.093 | 0.011 | 1.097 | 1.074 | 1.121 | $9.34 \times 10^{-18}$ |
| DQA1 | 64 | T | 0.466 | 0.442 | 0.093 | 0.011 | 1.097 | 1.074 | 1.121 | $9.34 \times 10^{-18}$ |
| DQA1 | 61 | G | 0.534 | 0.558 | -0.093 | 0.011 | 0.911 | 0.892 | 0.931 | $9.34 \times 10^{-18}$ |
| DQA1 | 48 | L | 0.466 | 0.442 | 0.093 | 0.011 | 1.097 | 1.074 | 1.121 | $9.34 \times 10^{-18}$ |
| DQA1 | 11 | C | 0.534 | 0.558 | -0.093 | 0.011 | 0.911 | 0.892 | 0.931 | $9.34 \times 10^{-18}$ |
| DQA1 | 80 | S | 0.466 | 0.442 | 0.093 | 0.011 | 1.097 | 1.074 | 1.121 | $9.34 \times 10^{-18}$ |
| DQA1 | 18 | F | 0.534 | 0.558 | -0.093 | 0.011 | 0.911 | 0.892 | 0.931 | $9.34 \times 10^{-18}$ |
| DQA1 | 11 | Y | 0.466 | 0.442 | 0.093 | 0.011 | 1.097 | 1.074 | 1.121 | $9.34 \times 10^{-18}$ |
| DQA1 | 80 | Y | 0.534 | 0.558 | -0.093 | 0.011 | 0.911 | 0.892 | 0.931 | $9.34 \times 10^{-18}$ |
| DQA1 | 48 | W | 0.534 | 0.558 | -0.093 | 0.011 | 0.911 | 0.892 | 0.931 | $9.34 \times 10^{-18}$ |
| DQA1 | 45 | V | 0.466 | 0.442 | 0.093 | 0.011 | 1.097 | 1.074 | 1.121 | $9.34 \times 10^{-18}$ |
| DQA1 | 18 | S | 0.466 | 0.442 | 0.093 | 0.011 | 1.097 | 1.074 | 1.121 | $9.34 \times 10^{-18}$ |
| DQA1 | 55 | R | 0.466 | 0.442 | 0.093 | 0.011 | 1.097 | 1.074 | 1.121 | $9.34 \times 10^{-18}$ |
| DQA1 | 64 | R | 0.534 | 0.558 | -0.093 | 0.011 | 0.911 | 0.892 | 0.931 | $9.34 \times 10^{-18}$ |
| DQA1 | 66 | M | 0.534 | 0.558 | -0.093 | 0.011 | 0.911 | 0.892 | 0.931 | $9.34 \times 10^{-18}$ |
| DQA1 | 61 | F | 0.466 | 0.442 | 0.093 | 0.011 | 1.097 | 1.074 | 1.121 | $9.34 \times 10^{-18}$ |
| DQB1 | 182 | S | 0.469 | 0.493 | -0.094 | 0.011 | 0.910 | 0.891 | 0.930 | $9.90 \times 10^{-18}$ |
| DQB1 | 140 | A | 0.469 | 0.493 | -0.094 | 0.011 | 0.910 | 0.891 | 0.930 | $9.90 \times 10^{-18}$ |
| DQB1 | 140 | T | 0.530 | 0.506 | 0.093 | 0.011 | 1.097 | 1.074 | 1.121 | $1.09 \times 10^{-17}$ |
| DQB1 | 182 | N | 0.530 | 0.506 | 0.093 | 0.011 | 1.097 | 1.074 | 1.121 | $1.09 \times 10^{-17}$ |
| DQA1 | 52 | R | 0.460 | 0.436 | 0.092 | 0.011 | 1.096 | 1.073 | 1.120 | $2.35 \times 10^{-17}$ |
| DRB1 | 181 | T | 0.831 | 0.847 | -0.124 | 0.015 | 0.883 | 0.858 | 0.910 | $5.60 \times 10^{-17}$ |
| DQB1 | 86 | A | 0.403 | 0.426 | -0.090 | 0.011 | 0.914 | 0.894 | 0.934 | $5.43 \times 10^{-16}$ |
| DRB1 | 181 | M | 0.160 | 0.145 | 0.122 | 0.015 | 1.130 | 1.097 | 1.163 | $6.20 \times 10^{-16}$ |
| DQB1 | 203 | V | 0.301 | 0.322 | -0.095 | 0.012 | 0.909 | 0.888 | 0.931 | $9.16 \times 10^{-16}$ |
| DQB1 | -21 | G | 0.699 | 0.678 | 0.095 | 0.012 | 1.100 | 1.074 | 1.126 | $9.18 \times 10^{-16}$ |
| DQB1 | -4 | V | 0.699 | 0.678 | 0.095 | 0.012 | 1.100 | 1.074 | 1.126 | $9.18 \times 10^{-16}$ |
| DQB1 | -5 | P | 0.699 | 0.678 | 0.095 | 0.012 | 1.100 | 1.074 | 1.126 | $9.18 \times 10^{-16}$ |
| DQB1 | 203 | I | 0.698 | 0.678 | 0.094 | 0.012 | 1.099 | 1.073 | 1.125 | $1.04 \times 10^{-15}$ |
| DRB1 | 47 | F | 0.382 | 0.402 | -0.089 | 0.011 | 0.915 | 0.895 | 0.935 | $2.17 \times 10^{-15}$ |
| DRB1 | 47 | Y | 0.618 | 0.598 | 0.089 | 0.011 | 1.093 | 1.069 | 1.117 | $2.17 \times 10^{-15}$ |
| DQA1 | 129 | Q | 0.288 | 0.307 | -0.096 | 0.012 | 0.908 | 0.887 | 0.930 | $4.24 \times 10^{-15}$ |
| DQA1 | 129 | H | 0.713 | 0.693 | 0.096 | 0.012 | 1.101 | 1.075 | 1.127 | $4.24 \times 10^{-15}$ |
| DQB1 | 70 | G | 0.246 | 0.264 | -0.098 | 0.013 | 0.907 | 0.884 | 0.930 | $8.61 \times 10^{-15}$ |
| DRB1 | -1 | S | 0.254 | 0.272 | -0.094 | 0.012 | 0.910 | 0.889 | 0.932 | $2.20 \times 10^{-14}$ |
| DRB1 | 71 | A | 0.254 | 0.272 | -0.094 | 0.012 | 0.910 | 0.889 | 0.932 | $2.20 \times 10^{-14}$ |
| DQB1 | -21 | D | 0.250 | 0.268 | -0.096 | 0.013 | 0.908 | 0.886 | 0.932 | $2.53 \times 10^{-14}$ |
| DQB1 | -6 | S | 0.250 | 0.268 | -0.096 | 0.013 | 0.908 | 0.886 | 0.932 | $2.53 \times 10^{-14}$ |
| DQB1 | -4 | L | 0.250 | 0.268 | -0.096 | 0.013 | 0.908 | 0.886 | 0.932 | $2.53 \times 10^{-14}$ |
| DRB1 | 71 | R | 0.674 | 0.655 | 0.087 | 0.012 | 1.091 | 1.066 | 1.117 | $3.71 \times 10^{-14}$ |
| DRB1 | 133 | R | 0.723 | 0.705 | 0.091 | 0.012 | 1.095 | 1.070 | 1.121 | $3.72 \times 10^{-14}$ |
| DRB1 | 142 | V | 0.723 | 0.705 | 0.091 | 0.012 | 1.095 | 1.070 | 1.121 | $3.72 \times 10^{-14}$ |
| DRB1 | 133 | L | 0.277 | 0.295 | -0.091 | 0.012 | 0.913 | 0.892 | 0.935 | 3.74×10<sup>-14</sup> |
| DRB1 | 142 | M | 0.277 | 0.295 | -0.091 | 0.012 | 0.913 | 0.892 | 0.935 | 3.74×10<sup>-14</sup> |
| DRB1 | 78 | V | 0.154 | 0.140 | 0.115 | 0.015 | 1.122 | 1.089 | 1.156 | 5.27×10<sup>-14</sup> |
| DRB1 | 78 | Y | 0.846 | 0.860 | -0.115 | 0.015 | 0.891 | 0.865 | 0.918 | 5.27×10<sup>-14</sup> |
| DRB1 | 4 | Q | 0.154 | 0.140 | 0.115 | 0.015 | 1.122 | 1.089 | 1.155 | 5.27×10<sup>-14</sup> |
| DRB1 | 11 | P | 0.256 | 0.273 | -0.092 | 0.012 | 0.912 | 0.891 | 0.934 | 6.15×10<sup>-14</sup> |
| DRB1 | 13 | R | 0.256 | 0.273 | -0.092 | 0.012 | 0.912 | 0.891 | 0.934 | 6.15×10<sup>-14</sup> |
| DRB1 | 9 | K | 0.144 | 0.131 | 0.117 | 0.016 | 1.124 | 1.089 | 1.160 | 1.46×10<sup>-13</sup> |
| DRB1 | 30 | G | 0.144 | 0.131 | 0.117 | 0.016 | 1.124 | 1.089 | 1.160 | 1.46×10<sup>-13</sup> |
| DRB1 | 28 | H | 0.144 | 0.131 | 0.117 | 0.016 | 1.124 | 1.089 | 1.160 | 1.46×10<sup>-13</sup> |
| DRB1 | 11 | D | 0.144 | 0.131 | 0.117 | 0.016 | 1.124 | 1.089 | 1.160 | 1.46×10<sup>-13</sup> |
| DRB1 | 74 | E | 0.286 | 0.269 | 0.091 | 0.012 | 1.095 | 1.070 | 1.121 | 1.56×10<sup>-13</sup> |
| DRB1 | -1 | A | 0.736 | 0.718 | 0.090 | 0.012 | 1.094 | 1.069 | 1.120 | 1.62×10<sup>-13</sup> |
| DQB1 | 55 | R | 0.601 | 0.621 | -0.082 | 0.011 | 0.921 | 0.902 | 0.941 | 2.35×10<sup>-13</sup> |
| DRB1 | 26 | Y | 0.145 | 0.131 | 0.115 | 0.016 | 1.122 | 1.087 | 1.158 | 2.52×10<sup>-13</sup> |
| DRB1 | 4 | R | 0.844 | 0.857 | -0.112 | 0.015 | 0.894 | 0.868 | 0.921 | 2.76×10<sup>-13</sup> |
| DQB1 | 55 | P | 0.388 | 0.369 | 0.082 | 0.011 | 1.085 | 1.062 | 1.109 | 4.61×10<sup>-13</sup> |
| DRB1 | 37 | S | 0.341 | 0.360 | -0.083 | 0.011 | 0.920 | 0.901 | 0.940 | 5.00×10<sup>-13</sup> |
| DRB1 | 57 | D | 0.561 | 0.581 | -0.080 | 0.011 | 0.923 | 0.903 | 0.943 | 5.72×10<sup>-13</sup> |
| DRB1 | 74 | A | 0.570 | 0.589 | -0.079 | 0.011 | 0.924 | 0.904 | 0.944 | 1.16×10<sup>-12</sup> |
| DRB1 | 96 | Q | 0.283 | 0.300 | -0.084 | 0.012 | 0.919 | 0.898 | 0.941 | 2.16×10<sup>-12</sup> |
| DRB1 | 60 | S | 0.194 | 0.179 | 0.098 | 0.014 | 1.103 | 1.073 | 1.134 | 3.00×10<sup>-12</sup> |
| DRB1 | 57 | V | 0.194 | 0.179 | 0.098 | 0.014 | 1.103 | 1.073 | 1.134 | 3.00×10<sup>-12</sup> |
| DRB1 | 67 | I | 0.437 | 0.455 | -0.074 | 0.011 | 0.929 | 0.909 | 0.949 | 1.70×10<sup>-11</sup> |
| DRB1 | 70 | R | 0.205 | 0.191 | 0.091 | 0.014 | 1.095 | 1.066 | 1.126 | 3.71×10<sup>-11</sup> |
| DRB1 | 60 | Y | 0.773 | 0.788 | -0.086 | 0.013 | 0.918 | 0.895 | 0.941 | 8.35×10<sup>-11</sup> |
| DQA1 | 34 | E | 0.483 | 0.467 | 0.071 | 0.011 | 1.073 | 1.050 | 1.096 | 8.56×10<sup>-11</sup> |
| DQA1 | 34 | Q | 0.517 | 0.534 | -0.071 | 0.011 | 0.932 | 0.912 | 0.952 | 8.56×10<sup>-11</sup> |
| DQB1 | 125 | G | 0.306 | 0.322 | -0.072 | 0.012 | 0.931 | 0.909 | 0.953 | 8.91×10<sup>-10</sup> |
| DQB1 | -6 | T | 0.595 | 0.579 | 0.068 | 0.011 | 1.070 | 1.048 | 1.094 | 9.57×10<sup>-10</sup> |
| DQB1 | -5 | L | 0.098 | 0.107 | -0.109 | 0.018 | 0.897 | 0.866 | 0.929 | 1.49×10<sup>-9</sup> |
| DQA1 | 207 | V | 0.822 | 0.810 | 0.086 | 0.015 | 1.090 | 1.058 | 1.122 | 2.23×10<sup>-9</sup> |
| DQA1 | 207 | M | 0.178 | 0.190 | -0.086 | 0.015 | 0.918 | 0.891 | 0.945 | 2.23×10<sup>-9</sup> |
| DRB1 | 37 | N | 0.222 | 0.210 | 0.075 | 0.013 | 1.078 | 1.051 | 1.106 | 1.06×10<sup>-8</sup> |
| DQB1 | 87 | F | 0.251 | 0.265 | -0.071 | 0.013 | 0.931 | 0.908 | 0.956 | 1.23×10<sup>-8</sup> |
| DPB1 | 205 | V | 0.582 | 0.597 | -0.063 | 0.011 | 0.939 | 0.919 | 0.959 | 1.74×10<sup>-8</sup> |
| DPB1 | 205 | M | 0.412 | 0.397 | 0.063 | 0.011 | 1.065 | 1.042 | 1.088 | 2.32×10<sup>-8</sup> |
| DPB1 | 55 | E | 0.432 | 0.416 | 0.061 | 0.011 | 1.063 | 1.040 | 1.086 | 3.85×10<sup>-8</sup> |
| DPB1 | 35 | L | 0.428 | 0.413 | 0.061 | 0.011 | 1.063 | 1.040 | 1.086 | 4.03×10<sup>-8</sup> |
| DQB1 | 70 | R | 0.612 | 0.599 | 0.060 | 0.011 | 1.062 | 1.039 | 1.085 | 9.69×10<sup>-8</sup> |
| DRB1 | 67 | F | 0.225 | 0.212 | 0.070 | 0.013 | 1.073 | 1.046 | 1.100 | 1.28×10<sup>-7</sup> |
| DPB1 | 35 | F | 0.560 | 0.574 | -0.059 | 0.011 | 0.943 | 0.923 | 0.963 | 1.29×10<sup>-7</sup> |
| DPA1 | 1 | I | 0.504 | 0.518 | -0.057 | 0.011 | 0.944 | 0.924 | 0.965 | 2.09×10<sup>-7</sup> |
| DPA1 | 4 | D | 0.504 | 0.518 | -0.057 | 0.011 | 0.944 | 0.924 | 0.965 | 2.09×10<sup>-7</sup> |
| DPA1 | 5 | H | 0.504 | 0.518 | -0.057 | 0.011 | 0.944 | 0.924 | 0.965 | 2.09×10<sup>-7</sup> |
| DPA1 | 6 | V | 0.504 | 0.518 | -0.057 | 0.011 | 0.944 | 0.924 | 0.965 | 2.09×10<sup>-7</sup> |
| DPA1 | 3 | A | 0.504 | 0.518 | -0.057 | 0.011 | 0.944 | 0.924 | 0.965 | 2.09×10<sup>-7</sup> |
| DPA1 | 7 | S | 0.504 | 0.518 | -0.057 | 0.011 | 0.944 | 0.924 | 0.965 | 2.09×10<sup>-7</sup> |
| DPA1 | 2 | K | 0.504 | 0.518 | -0.057 | 0.011 | 0.944 | 0.924 | 0.965 | 2.09×10<sup>-7</sup> |
| DPA1 | 11 | A | 0.505 | 0.519 | -0.057 | 0.011 | 0.945 | 0.925 | 0.965 | 2.68×10<sup>-7</sup> |
| DPA1 | 11 | M | 0.495 | 0.481 | 0.057 | 0.011 | 1.058 | 1.036 | 1.081 | 2.68×10<sup>-7</sup> |
| DRB1 | 11 | V | 0.232 | 0.221 | 0.066 | 0.013 | 1.068 | 1.041 | 1.096 | 2.88×10<sup>-7</sup> |
| DRB1 | 120 | N | 0.232 | 0.221 | 0.066 | 0.013 | 1.068 | 1.041 | 1.096 | 2.88×10<sup>-7</sup> |
| DRB1 | 120 | S | 0.768 | 0.779 | -0.066 | 0.013 | 0.936 | 0.913 | 0.960 | 2.89×10<sup>-7</sup> |
| C | -9 | A | 0.866 | 0.856 | 0.080 | 0.016 | 1.083 | 1.050 | 1.118 | 4.56×10<sup>-7</sup> |
| C | 184 | H | 0.866 | 0.856 | 0.080 | 0.016 | 1.083 | 1.050 | 1.118 | 4.56×10<sup>-7</sup> |
| C | -17 | T | 0.866 | 0.856 | 0.080 | 0.016 | 1.083 | 1.050 | 1.118 | 4.56×10<sup>-7</sup> |
| C | 194 | V | 0.866 | 0.856 | 0.080 | 0.016 | 1.083 | 1.050 | 1.118 | 4.56×10<sup>-7</sup> |
| C | 295 | A | 0.865 | 0.855 | 0.080 | 0.016 | 1.083 | 1.050 | 1.118 | 4.89×10<sup>-7</sup> |
| C | 261 | V | 0.865 | 0.855 | 0.080 | 0.016 | 1.083 | 1.050 | 1.118 | 4.89×10<sup>-7</sup> |
| C | 285 | V | 0.865 | 0.855 | 0.080 | 0.016 | 1.083 | 1.050 | 1.118 | 4.89×10<sup>-7</sup> |
| C | 307 | V | 0.865 | 0.855 | 0.080 | 0.016 | 1.083 | 1.050 | 1.118 | 4.89×10<sup>-7</sup> |
| C | 253 | E | 0.865 | 0.855 | 0.080 | 0.016 | 1.083 | 1.050 | 1.118 | 4.89×10<sup>-7</sup> |
| C | 306 | V | 0.865 | 0.855 | 0.080 | 0.016 | 1.083 | 1.050 | 1.118 | 4.89×10<sup>-7</sup> |
| C | 267 | P | 0.865 | 0.855 | 0.080 | 0.016 | 1.083 | 1.050 | 1.118 | 4.89×10<sup>-7</sup> |
| C | 305 | A | 0.865 | 0.855 | 0.080 | 0.016 | 1.083 | 1.050 | 1.118 | 4.89×10<sup>-7</sup> |
| C | 273 | R | 0.865 | 0.855 | 0.080 | 0.016 | 1.083 | 1.050 | 1.118 | 4.89×10<sup>-7</sup> |
| C | 339 | A | 0.865 | 0.855 | 0.080 | 0.016 | 1.083 | 1.050 | 1.118 | 4.89×10<sup>-7</sup> |
| C | 326 | S | 0.865 | 0.855 | 0.080 | 0.016 | 1.083 | 1.050 | 1.118 | 4.89×10<sup>-7</sup> |
| C | 285 | M | 0.133 | 0.143 | -0.079 | 0.016 | 0.924 | 0.896 | 0.953 | 6.42×10<sup>-7</sup> |
| C | 184 | P | 0.133 | 0.143 | -0.079 | 0.016 | 0.924 | 0.896 | 0.953 | 6.42×10<sup>-7</sup> |
| C | -9 | G | 0.133 | 0.143 | -0.079 | 0.016 | 0.924 | 0.896 | 0.953 | 6.42×10<sup>-7</sup> |
| C | 295 | V | 0.133 | 0.143 | -0.079 | 0.016 | 0.924 | 0.896 | 0.953 | 6.42×10<sup>-7</sup> |
| C | 307 | M | 0.133 | 0.143 | -0.079 | 0.016 | 0.924 | 0.896 | 0.953 | 6.42×10<sup>-7</sup> |
| C | 339 | T | 0.133 | 0.143 | -0.079 | 0.016 | 0.924 | 0.896 | 0.953 | 6.42×10<sup>-7</sup> |
| C | 273 | S | 0.133 | 0.143 | -0.079 | 0.016 | 0.924 | 0.896 | 0.953 | 6.42×10<sup>-7</sup> |
| C | 326 | C | 0.133 | 0.143 | -0.079 | 0.016 | 0.924 | 0.896 | 0.953 | 6.42×10<sup>-7</sup> |
| C | 267 | Q | 0.133 | 0.143 | -0.079 | 0.016 | 0.924 | 0.896 | 0.953 | 6.42×10<sup>-7</sup> |
| C | 253 | Q | 0.133 | 0.143 | -0.079 | 0.016 | 0.924 | 0.896 | 0.953 | 6.42×10<sup>-7</sup> |
| C | -17 | A | 0.133 | 0.143 | -0.079 | 0.016 | 0.924 | 0.896 | 0.953 | 6.42×10<sup>-7</sup> |
| C | 261 | M | 0.133 | 0.143 | -0.079 | 0.016 | 0.924 | 0.896 | 0.953 | 6.42×10<sup>-7</sup> |
| C | 305 | T | 0.133 | 0.143 | -0.079 | 0.016 | 0.924 | 0.896 | 0.953 | 6.42×10<sup>-7</sup> |
| C | 194 | L | 0.133 | 0.143 | -0.079 | 0.016 | 0.924 | 0.896 | 0.953 | 6.42×10<sup>-7</sup> |
| C | 306 | A | 0.133 | 0.143 | -0.079 | 0.016 | 0.924 | 0.896 | 0.953 | 6.42×10<sup>-7</sup> |
| C | 147 | L | 0.133 | 0.143 | -0.079 | 0.016 | 0.924 | 0.896 | 0.953 | 6.42×10<sup>-7</sup> |
| C | 147 | W | 0.867 | 0.857 | 0.079 | 0.016 | 1.082 | 1.049 | 1.116 | 6.42×10<sup>-7</sup> |
| C | 152 | A | 0.133 | 0.143 | -0.079 | 0.016 | 0.924 | 0.896 | 0.953 | 6.47×10<sup>-7</sup> |
| DRB1 | 180 | V | 0.764 | 0.775 | -0.063 | 0.013 | 0.939 | 0.915 | 0.963 | 8.22×10<sup>-7</sup> |
| C | 99 | S | 0.118 | 0.128 | -0.083 | 0.017 | 0.920 | 0.890 | 0.952 | 8.53×10<sup>-7</sup> |
| DRB1 | 70 | Q | 0.520 | 0.533 | -0.054 | 0.011 | 0.947 | 0.927 | 0.968 | 8.76×10<sup>-7</sup> |
| DRB1 | -24 | L | 0.762 | 0.773 | -0.061 | 0.013 | 0.941 | 0.917 | 0.965 | 1.91×10<sup>-6</sup> |
| DQB1 | 57 | V | 0.117 | 0.127 | -0.080 | 0.017 | 0.923 | 0.893 | 0.954 | 2.08×10<sup>-6</sup> |
| DRB1 | 28 | D | 0.746 | 0.757 | -0.060 | 0.013 | 0.942 | 0.918 | 0.966 | 2.20×10<sup>-6</sup> |
| DRB1 | 13 | F | 0.194 | 0.184 | 0.066 | 0.014 | 1.068 | 1.039 | 1.098 | 2.34×10<sup>-6</sup> |
| DRB1 | 31 | F | 0.806 | 0.816 | -0.066 | 0.014 | 0.936 | 0.911 | 0.962 | 2.34×10<sup>-6</sup> |
| DPB1 | 56 | A | 0.476 | 0.463 | 0.053 | 0.011 | 1.054 | 1.031 | 1.078 | 2.45×10<sup>-6</sup> |
| DPB1 | 56 | E | 0.524 | 0.537 | -0.053 | 0.011 | 0.949 | 0.928 | 0.970 | 2.45×10<sup>-6</sup> |
| DPB1 | 55 | D | 0.524 | 0.537 | -0.053 | 0.011 | 0.948 | 0.928 | 0.969 | 2.45×10<sup>-6</sup> |
| DPB1 | 84 | D | 0.581 | 0.568 | 0.053 | 0.011 | 1.054 | 1.032 | 1.077 | 2.48×10<sup>-6</sup> |
| DPB1 | 84 | G | 0.419 | 0.432 | -0.053 | 0.011 | 0.948 | 0.928 | 0.969 | 2.49×10<sup>-6</sup> |
| DPB1 | 86 | A | 0.581 | 0.568 | 0.053 | 0.011 | 1.054 | 1.031 | 1.078 | 2.49×10<sup>-6</sup> |
| DPB1 | 85 | G | 0.419 | 0.432 | -0.053 | 0.011 | 0.948 | 0.928 | 0.970 | 2.49×10<sup>-6</sup> |
| DPB1 | 87 | M | 0.419 | 0.432 | -0.053 | 0.011 | 0.948 | 0.928 | 0.970 | 2.49×10<sup>-6</sup> |
| DPB1 | 87 | V | 0.581 | 0.568 | 0.053 | 0.011 | 1.054 | 1.031 | 1.078 | 2.49×10<sup>-6</sup> |
| DPB1 | 86 | P | 0.419 | 0.432 | -0.053 | 0.011 | 0.948 | 0.928 | 0.970 | 2.49×10<sup>-6</sup> |
| DPB1 | 85 | E | 0.581 | 0.568 | 0.053 | 0.011 | 1.054 | 1.031 | 1.078 | 2.49×10<sup>-6</sup> |
| DPB1 | 170 | T | 0.421 | 0.434 | -0.053 | 0.011 | 0.948 | 0.928 | 0.969 | 2.52×10<sup>-6</sup> |
| DPB1 | 96 | R | 0.421 | 0.434 | -0.053 | 0.011 | 0.948 | 0.928 | 0.969 | 2.52×10<sup>-6</sup> |
| DPB1 | 170 | I | 0.577 | 0.564 | 0.053 | 0.011 | 1.054 | 1.032 | 1.077 | 2.74×10<sup>-6</sup> |
| DPB1 | 96 | K | 0.577 | 0.564 | 0.053 | 0.011 | 1.054 | 1.032 | 1.077 | 2.74×10<sup>-6</sup> |
| DQB1 | -27 | A | 0.796 | 0.785 | 0.064 | 0.014 | 1.066 | 1.037 | 1.096 | 2.91×10<sup>-6</sup> |
| DQB1 | -9 | M | 0.796 | 0.785 | 0.064 | 0.014 | 1.066 | 1.037 | 1.096 | 2.91×10<sup>-6</sup> |
| DRB1 | 13 | H | 0.227 | 0.216 | 0.060 | 0.013 | 1.062 | 1.035 | 1.089 | 3.98×10<sup>-6</sup> |
| DRB1 | 96 | Y | 0.227 | 0.216 | 0.060 | 0.013 | 1.062 | 1.035 | 1.089 | 3.98×10<sup>-6</sup> |
| DRB1 | 180 | L | 0.227 | 0.216 | 0.060 | 0.013 | 1.062 | 1.035 | 1.089 | 3.98×10<sup>-6</sup> |
| DRB1 | -24 | F | 0.227 | 0.216 | 0.060 | 0.013 | 1.062 | 1.035 | 1.089 | 3.98×10<sup>-6</sup> |
| DRB1 | 33 | H | 0.227 | 0.216 | 0.060 | 0.013 | 1.061 | 1.035 | 1.089 | 3.98×10<sup>-6</sup> |
| DRB1 | 33 | N | 0.774 | 0.784 | -0.060 | 0.013 | 0.942 | 0.919 | 0.966 | 3.98×10<sup>-6</sup> |
| DRB1 | 30 | Y | 0.755 | 0.765 | -0.059 | 0.013 | 0.943 | 0.919 | 0.967 | 4.73×10<sup>-6</sup> |
| DRB1 | 96 | H | 0.451 | 0.439 | 0.050 | 0.011 | 1.051 | 1.029 | 1.074 | 5.68×10<sup>-6</sup> |
| DQB1 | 87 | Y | 0.208 | 0.219 | -0.062 | 0.014 | 0.940 | 0.914 | 0.966 | 5.75×10<sup>-6</sup> |
| DRB1 | 11 | L | 0.044 | 0.049 | -0.126 | 0.028 | 0.882 | 0.835 | 0.931 | 5.89×10<sup>-6</sup> |
| DRB1 | 30 | C | 0.044 | 0.049 | -0.126 | 0.028 | 0.882 | 0.835 | 0.931 | 5.89×10<sup>-6</sup> |
| DRB1 | 96 | E | 0.044 | 0.049 | -0.126 | 0.028 | 0.882 | 0.835 | 0.931 | 5.89×10<sup>-6</sup> |
| C | -15 | I | 0.827 | 0.817 | 0.064 | 0.014 | 1.066 | 1.037 | 1.096 | 7.19×10<sup>-6</sup> |
| B | 99 | Y | 0.994 | 0.996 | -0.337 | 0.076 | 0.714 | 0.615 | 0.829 | 7.79×10<sup>-6</sup> |
| B | 77 | D | 0.006 | 0.004 | 0.337 | 0.076 | 1.401 | 1.207 | 1.626 | 7.92×10<sup>-6</sup> |
| B | 99 | S | 0.006 | 0.004 | 0.337 | 0.076 | 1.401 | 1.207 | 1.626 | 7.92×10<sup>-6</sup> |
| C | -15 | L | 0.173 | 0.182 | -0.063 | 0.014 | 0.939 | 0.914 | 0.965 | 9.45×10<sup>-6</sup> |
| DQB1 | 38 | A | 0.679 | 0.667 | 0.051 | 0.012 | 1.052 | 1.028 | 1.077 | 1.49×10<sup>-5</sup> |
| DQB1 | 38 | V | 0.321 | 0.333 | -0.051 | 0.012 | 0.950 | 0.929 | 0.973 | 1.49×10<sup>-5</sup> |
| DRB1 | 26 | F | 0.765 | 0.774 | -0.056 | 0.013 | 0.946 | 0.922 | 0.970 | 1.68×10<sup>-5</sup> |
| C | 275 | K | 0.062 | 0.058 | 0.101 | 0.024 | 1.106 | 1.055 | 1.160 | 1.71×10<sup>-5</sup> |
| DQB1 | 30 | H | 0.218 | 0.228 | -0.057 | 0.013 | 0.945 | 0.921 | 0.969 | 1.91×10<sup>-5</sup> |
| DPA1 | 50 | Q | 0.424 | 0.435 | -0.049 | 0.011 | 0.952 | 0.931 | 0.974 | 1.93×10<sup>-5</sup> |
| DPA1 | 83 | A | 0.577 | 0.565 | 0.049 | 0.011 | 1.050 | 1.027 | 1.074 | 1.93×10<sup>-5</sup> |
| DPA1 | 160 | F | 0.424 | 0.435 | -0.049 | 0.011 | 0.952 | 0.931 | 0.974 | 1.93×10<sup>-5</sup> |
| DPA1 | 228 | T | 0.424 | 0.435 | -0.049 | 0.011 | 0.952 | 0.931 | 0.974 | 1.93×10<sup>-5</sup> |
| DPA1 | 127 | L | 0.424 | 0.435 | -0.049 | 0.011 | 0.952 | 0.931 | 0.974 | 1.93×10<sup>-5</sup> |
| DPA1 | 160 | V | 0.577 | 0.565 | 0.049 | 0.011 | 1.050 | 1.027 | 1.074 | 1.93×10<sup>-5</sup> |
| DPA1 | 83 | T | 0.424 | 0.435 | -0.049 | 0.011 | 0.952 | 0.931 | 0.974 | 1.93×10<sup>-5</sup> |
| DPA1 | 50 | R | 0.577 | 0.565 | 0.049 | 0.011 | 1.050 | 1.027 | 1.074 | 1.93×10<sup>-5</sup> |
| DPA1 | 228 | P | 0.577 | 0.565 | 0.049 | 0.011 | 1.050 | 1.027 | 1.074 | 1.93×10<sup>-5</sup> |
| DPA1 | 127 | P | 0.577 | 0.565 | 0.049 | 0.011 | 1.050 | 1.027 | 1.074 | 1.93×10<sup>-5</sup> |
| DPA1 | -17 | L | 0.424 | 0.435 | -0.049 | 0.011 | 0.952 | 0.931 | 0.974 | 1.94×10<sup>-5</sup> |
| DPA1 | -30 | R | 0.424 | 0.435 | -0.049 | 0.011 | 0.952 | 0.931 | 0.974 | 1.94×10<sup>-5</sup> |
| DPA1 | -25 | M | 0.424 | 0.435 | -0.049 | 0.011 | 0.952 | 0.931 | 0.974 | 1.94×10<sup>-5</sup> |
| DPA1 | -20 | A | 0.424 | 0.435 | -0.049 | 0.011 | 0.952 | 0.931 | 0.974 | 1.94×10<sup>-5</sup> |
| DPA1 | -21 | R | 0.424 | 0.435 | -0.049 | 0.011 | 0.952 | 0.931 | 0.974 | 1.94×10<sup>-5</sup> |
| DPA1 | -28 | E | 0.424 | 0.435 | -0.049 | 0.011 | 0.952 | 0.931 | 0.974 | 1.94×10<sup>-5</sup> |
| DPA1 | -18 | I | 0.424 | 0.435 | -0.049 | 0.011 | 0.952 | 0.931 | 0.974 | 1.94×10<sup>-5</sup> |
| DPA1 | -14 | L | 0.424 | 0.435 | -0.049 | 0.011 | 0.952 | 0.931 | 0.974 | 1.94×10<sup>-5</sup> |
| DPA1 | -27 | D | 0.424 | 0.435 | -0.049 | 0.011 | 0.952 | 0.931 | 0.974 | 1.94×10<sup>-5</sup> |
| DPA1 | -16 | R | 0.424 | 0.435 | -0.049 | 0.011 | 0.952 | 0.931 | 0.974 | 1.94×10<sup>-5</sup> |
| DPA1 | -24 | F | 0.424 | 0.435 | -0.049 | 0.011 | 0.952 | 0.931 | 0.974 | 1.94×10<sup>-5</sup> |
| DPA1 | -11 | A | 0.424 | 0.435 | -0.049 | 0.011 | 0.952 | 0.931 | 0.974 | 1.94×10<sup>-5</sup> |
| DPA1 | -29 | P | 0.424 | 0.435 | -0.049 | 0.011 | 0.952 | 0.931 | 0.974 | 1.94×10<sup>-5</sup> |
| DPA1 | -23 | H | 0.424 | 0.435 | -0.049 | 0.011 | 0.952 | 0.931 | 0.974 | 1.94×10<sup>-5</sup> |
| DPA1 | -9 | L | 0.424 | 0.435 | -0.049 | 0.011 | 0.952 | 0.931 | 0.974 | 1.94×10<sup>-5</sup> |
| DPA1 | -22 | I | 0.424 | 0.435 | -0.049 | 0.011 | 0.952 | 0.931 | 0.974 | 1.94×10<sup>-5</sup> |
| DPA1 | -5 | R | 0.424 | 0.435 | -0.049 | 0.011 | 0.952 | 0.931 | 0.974 | 1.94×10<sup>-5</sup> |
| DPA1 | -19 | V | 0.424 | 0.435 | -0.049 | 0.011 | 0.952 | 0.931 | 0.974 | 1.94×10<sup>-5</sup> |
| DPA1 | -3 | A | 0.424 | 0.435 | -0.049 | 0.011 | 0.952 | 0.931 | 0.974 | 1.94×10<sup>-5</sup> |
| DPA1 | -8 | L | 0.424 | 0.435 | -0.049 | 0.011 | 0.952 | 0.931 | 0.974 | 1.94×10<sup>-5</sup> |
| DPA1 | -4 | G | 0.424 | 0.435 | -0.049 | 0.011 | 0.952 | 0.931 | 0.974 | 1.94×10<sup>-5</sup> |
| DPA1 | -31 | M | 0.424 | 0.435 | -0.049 | 0.011 | 0.952 | 0.931 | 0.974 | 1.94×10<sup>-5</sup> |
| DPA1 | -12 | L | 0.424 | 0.435 | -0.049 | 0.011 | 0.952 | 0.931 | 0.974 | 1.94×10<sup>-5</sup> |
| DPA1 | -2 | G | 0.424 | 0.435 | -0.049 | 0.011 | 0.952 | 0.931 | 0.974 | 1.94×10<sup>-5</sup> |
| DPA1 | -13 | S | 0.424 | 0.435 | -0.049 | 0.011 | 0.952 | 0.931 | 0.974 | 1.94×10<sup>-5</sup> |
| DPA1 | -26 | R | 0.424 | 0.435 | -0.049 | 0.011 | 0.952 | 0.931 | 0.974 | 1.94×10<sup>-5</sup> |
| DPA1 | -10 | F | 0.424 | 0.435 | -0.049 | 0.011 | 0.952 | 0.931 | 0.974 | 1.94×10<sup>-5</sup> |
| DPA1 | -7 | S | 0.424 | 0.435 | -0.049 | 0.011 | 0.952 | 0.931 | 0.974 | 1.94×10<sup>-5</sup> |
| DPA1 | -15 | A | 0.424 | 0.435 | -0.049 | 0.011 | 0.952 | 0.931 | 0.974 | 1.94×10<sup>-5</sup> |
| DPA1 | -6 | L | 0.424 | 0.435 | -0.049 | 0.011 | 0.952 | 0.931 | 0.974 | 1.94×10<sup>-5</sup> |
| DPA1 | -1 | A | 0.424 | 0.435 | -0.049 | 0.011 | 0.952 | 0.931 | 0.974 | 1.94×10<sup>-5</sup> |
| C | 219 | W | 0.633 | 0.620 | 0.048 | 0.011 | 1.049 | 1.027 | 1.072 | 2.02×10<sup>-5</sup> |
| DPA1 | 31 | M | 0.425 | 0.436 | -0.048 | 0.011 | 0.953 | 0.932 | 0.974 | 2.39×10<sup>-5</sup> |
| DPA1 | 111 | K | 0.425 | 0.436 | -0.048 | 0.011 | 0.953 | 0.932 | 0.974 | 2.39×10<sup>-5</sup> |
| DPA1 | 111 | R | 0.575 | 0.564 | 0.048 | 0.011 | 1.049 | 1.026 | 1.073 | 2.39×10<sup>-5</sup> |
| DPA1 | 31 | Q | 0.575 | 0.564 | 0.048 | 0.011 | 1.049 | 1.026 | 1.073 | 2.39×10<sup>-5</sup> |
| C | 219 | R | 0.365 | 0.378 | -0.047 | 0.011 | 0.954 | 0.934 | 0.975 | 2.49×10<sup>-5</sup> |
| C | 152 | E | 0.786 | 0.776 | 0.056 | 0.013 | 1.058 | 1.031 | 1.085 | 2.49×10<sup>-5</sup> |
| C | 77 | N | 0.112 | 0.105 | 0.074 | 0.018 | 1.077 | 1.041 | 1.115 | 2.56×10<sup>-5</sup> |
| C | 80 | K | 0.112 | 0.105 | 0.074 | 0.018 | 1.077 | 1.041 | 1.115 | 2.56×10<sup>-5</sup> |
| C | 77 | S | 0.888 | 0.895 | -0.074 | 0.018 | 0.928 | 0.897 | 0.961 | 2.56×10<sup>-5</sup> |
| C | 80 | N | 0.888 | 0.895 | -0.074 | 0.018 | 0.928 | 0.897 | 0.961 | 2.56×10<sup>-5</sup> |
| DRB1 | 31 | I | 0.188 | 0.180 | 0.059 | 0.014 | 1.061 | 1.032 | 1.090 | 3.20×10<sup>-5</sup> |
| DQB1 | 14 | L | 0.153 | 0.161 | -0.064 | 0.015 | 0.938 | 0.910 | 0.967 | 3.62×10<sup>-5</sup> |
| DQB1 | 14 | M | 0.847 | 0.839 | 0.064 | 0.015 | 1.066 | 1.034 | 1.098 | 3.62×10<sup>-5</sup> |
| DQB1 | 116 | I | 0.153 | 0.161 | -0.064 | 0.015 | 0.938 | 0.911 | 0.966 | 3.62×10<sup>-5</sup> |
| DQB1 | -27 | S | 0.153 | 0.161 | -0.064 | 0.015 | 0.938 | 0.911 | 0.966 | 3.62×10<sup>-5</sup> |
| DQB1 | 71 | A | 0.153 | 0.161 | -0.064 | 0.015 | 0.938 | 0.911 | 0.966 | 3.62×10<sup>-5</sup> |
| DQB1 | 224 | R | 0.153 | 0.161 | -0.064 | 0.015 | 0.938 | 0.911 | 0.966 | 3.62×10<sup>-5</sup> |
| DQB1 | 125 | S | 0.153 | 0.161 | -0.064 | 0.015 | 0.938 | 0.911 | 0.966 | 3.62×10<sup>-5</sup> |
| DQB1 | -9 | I | 0.153 | 0.161 | -0.064 | 0.015 | 0.938 | 0.911 | 0.966 | 3.62×10<sup>-5</sup> |
| DQB1 | -5 | S | 0.153 | 0.161 | -0.064 | 0.015 | 0.938 | 0.911 | 0.966 | 3.62×10<sup>-5</sup> |
| DQB1 | 224 | Q | 0.847 | 0.838 | 0.063 | 0.015 | 1.065 | 1.034 | 1.097 | 3.89×10<sup>-5</sup> |
| DQB1 | 116 | V | 0.847 | 0.838 | 0.063 | 0.015 | 1.065 | 1.034 | 1.097 | 3.89×10<sup>-5</sup> |
| C | 9 | D | 0.143 | 0.152 | -0.063 | 0.015 | 0.939 | 0.912 | 0.967 | 4.54×10<sup>-5</sup> |
| DQB1 | 30 | Y | 0.770 | 0.761 | 0.053 | 0.013 | 1.054 | 1.028 | 1.082 | 5.32×10<sup>-5</sup> |
| C | 99 | F | 0.177 | 0.169 | 0.058 | 0.014 | 1.060 | 1.031 | 1.089 | 5.89×10<sup>-5</sup> |
| C | 49 | A | 0.944 | 0.947 | -0.097 | 0.024 | 0.907 | 0.865 | 0.951 | 5.93×10<sup>-5</sup> |
| C | 14 | R | 0.944 | 0.947 | -0.097 | 0.024 | 0.907 | 0.865 | 0.951 | 5.93×10<sup>-5</sup> |
| C | 14 | W | 0.057 | 0.053 | 0.097 | 0.024 | 1.102 | 1.051 | 1.156 | 5.93×10<sup>-5</sup> |
| C | 49 | E | 0.057 | 0.053 | 0.097 | 0.024 | 1.102 | 1.051 | 1.156 | 5.93×10<sup>-5</sup> |
| C | 303 | M | 0.057 | 0.053 | 0.097 | 0.024 | 1.102 | 1.051 | 1.155 | 5.93×10<sup>-5</sup> |

**Supplementary Table 8.** Fine-mapping results across the MHC region. List of variants that satisfied a posterior inclusion probability (PIP) greater than 0.1 in the fine-mapping.

| SNP | Effect allele | EAF | Beta | StdErr | z | PIP | logBF | Nearest gene | Annotation |
| --- | --- | --- | --- | --- | --- | --- | --- | --- | --- |
| rs533028614 | T | 0.961 | 0.179 | 0.073 | 2.47 | 1.00 | 14.59 | *HLA-DQA1* | intronic |
| rs114598250 | C | 0.959 | 0.061 | 0.024 | 2.51 | 1.00 | 14.59 | *HLA-DQA1* | intronic |
| rs1047989 | A | 0.520 | 0.103 | 0.010 | 10.81 | 1.00 | 14.59 | *HLA-DQA1* | exonic |
| rs7751376 | G | 0.561 | 0.096 | 0.010 | 9.96 | 1.00 | 14.59 | *HLA-DQA1* | intronic |
| rs9273382 | G | 0.557 | 0.093 | 0.010 | 9.70 | 1.00 | 14.59 | *HLA-DQA1* | intergenic |

**Supplementary Table 9.** HLA allele-level conditional analyses (rs1047989 as covariate). The four-digit and two-digit HLA alleles that meet the significance level of the Bonferroni correction (P-value < 0.05 / 155 = 3.23×10⁻⁴).

| HLA allele | Freq |  | Beta | StdErr | Odds ratio | 95% CI |  | P-value | Bonferroni-P |
| --- | --- | --- | --- | --- | --- | --- | --- | --- | --- |
|  | Case | Control |  |  |  | Lower bound | Upper bound |  |  |
| DRB1*15 | 0.275 | 0.296 | -0.068 | 0.014 | 0.934 | 0.908 | 0.961 | $1.52 \times 10^{-6}$ | $2.36 \times 10^{-4}$ |
| DPB1*05 | 0.413 | 0.395 | 0.052 | 0.013 | 1.053 | 1.028 | 1.079 | $3.63 \times 10^{-5}$ | $5.63 \times 10^{-3}$ |
| DPA1*02 | 0.520 | 0.506 | 0.051 | 0.012 | 1.052 | 1.027 | 1.078 | $3.85 \times 10^{-5}$ | $5.63 \times 10^{-3}$ |
| DPA1*01:03 | 0.422 | 0.435 | -0.050 | 0.012 | 0.951 | 0.927 | 0.976 | $5.68 \times 10^{-5}$ | $8.81 \times 10^{-3}$ |
| DPA1*01 | 0.478 | 0.492 | -0.050 | 0.012 | 0.952 | 0.928 | 0.976 | $5.96 \times 10^{-5}$ | $9.24 \times 10^{-3}$ |
| DPB1*05:01 | 0.410 | 0.393 | 0.049 | 0.013 | 1.050 | 1.025 | 1.076 | $8.57 \times 10^{-5}$ | 0.0133 |
| A*33 | 0.058 | 0.057 | 0.095 | 0.026 | 1.100 | 1.045 | 1.158 | $2.93 \times 10^{-4}$ | 0.0454 |
| A*33:03 | 0.059 | 0.057 | 0.095 | 0.026 | 1.100 | 1.044 | 1.157 | $2.97 \times 10^{-4}$ | 0.046 |
| DPA1*02:02 | 0.500 | 0.481 | 0.044 | 0.012 | 1.045 | 1.020 | 1.071 | $3.14 \times 10^{-4}$ | 0.0486 |
| DRB1*15:01 | 0.125 | 0.135 | -0.065 | 0.018 | 0.938 | 0.905 | 0.972 | $3.25 \times 10^{-4}$ | 0.0504 |
| DQB1*06:02 | 0.080 | 0.089 | -0.077 | 0.022 | 0.926 | 0.885 | 0.968 | $6.45 \times 10^{-4}$ | 0.0538 |

**Supplementary Table 10.** Cross-ancestry Meta-GWAS. A list of regions reaching genome-wide significance.

| Genomic locus | Lead SNP | Effect allele | EAF | Beta_0 | StdErr_0 | P-value | Nearest gene | Annotation |
| --- | --- | --- | --- | --- | --- | --- | --- | --- |
| 3q12.3 | rs616597 | C | 0.608 | 0.118 | 0.077 | $4.81 \times 10^{-9}$ | *NFKBIZ* | intronic |
| 3q13.2 | rs146203219 | G | 0.664 | 0.203 | 0.093 | $1.32 \times 10^{-27}$ | *CCDC80* | intergenic |
| 4p14 | rs12233670 | C | 0.507 | 0.023 | 0.011 | $3.31 \times 10^{-14}$ | *TLR10* | intergenic |
| 6p21.3 | rs9273149 | A | 0.395 | 0.129 | 0.012 | $2.99 \times 10^{-32}$ | *HLA-DQA1* | intergenic |
| 6q22.1 | rs11153598 | A | 0.313 | 0.043 | 0.012 | $4.69 \times 10^{-8}$ | *NT5DC1* | intronic |
| 8q21.2 | rs145917343 | T | 0.033 | 0.487 | 0.135 | $3.16 \times 10^{-8}$ | *REXO1L2P* | intergenic |
| 8q24.3 | rs2717561 | T | 0.759 | 0.158 | 0.101 | $1.02 \times 10^{-9}$ | *LY6K* | intergenic |
| 17p12 | rs1550656 | C | 0.197 | 0.095 | 0.016 | $2.34 \times 10^{-16}$ | *LINC00675* | exonic |

Using MR-MEGA, we integrated the results with those of prior studies conducted in populations of European ancestry.
Beta\_0: Effect of the intercept meta-regression. SE\_0: Standard error of the intercept meta-regression.

**Supplementary Table 11.**
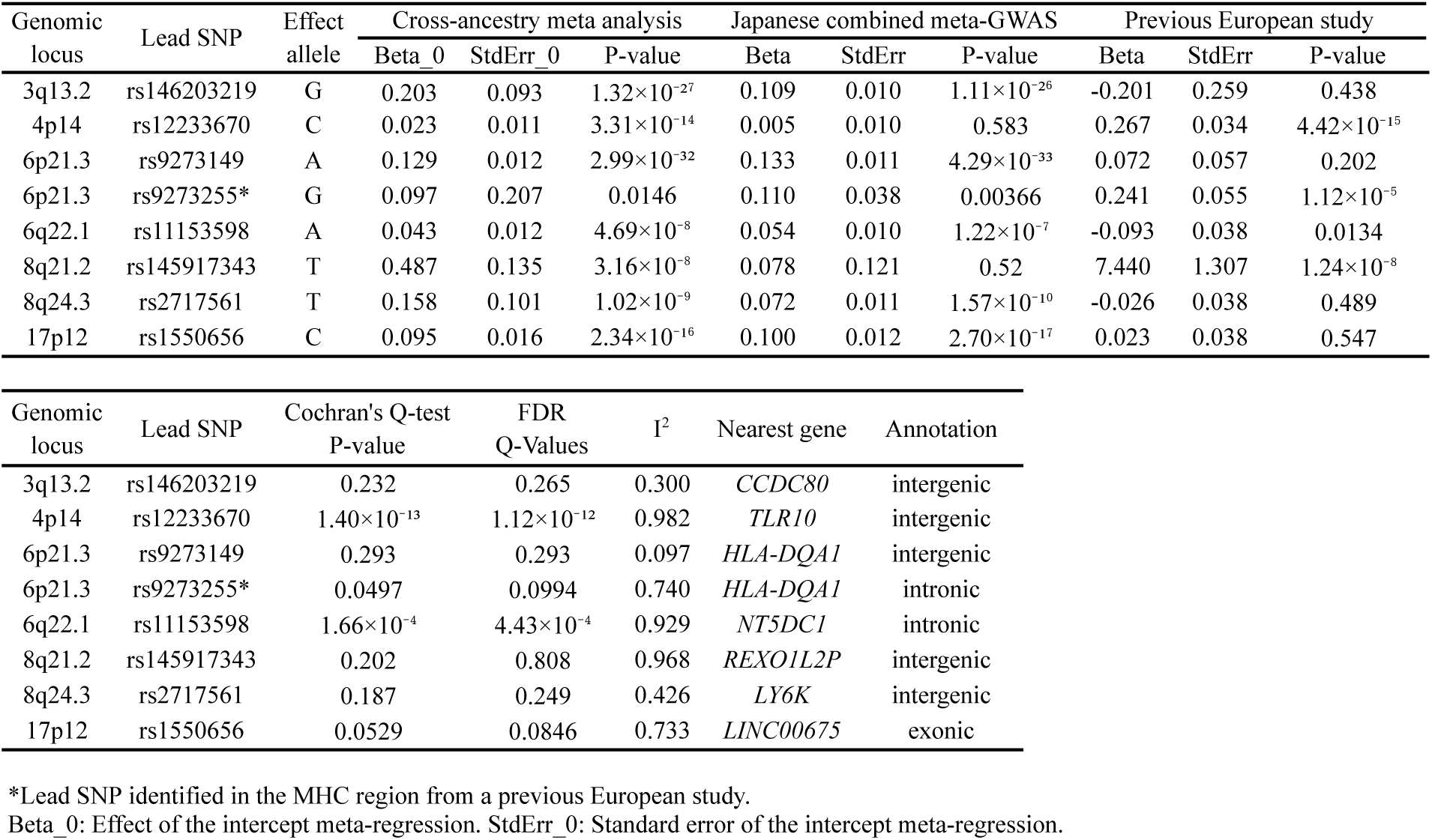
Comparison of the lead variants found in both the Japanese and the European ancestry GWAS. Comparison of the lead variants found in both the Japanese and the European ancestry GWAS among the loci identified through the cross-ancestry meta-analysis

**Supplementary Table 12.**
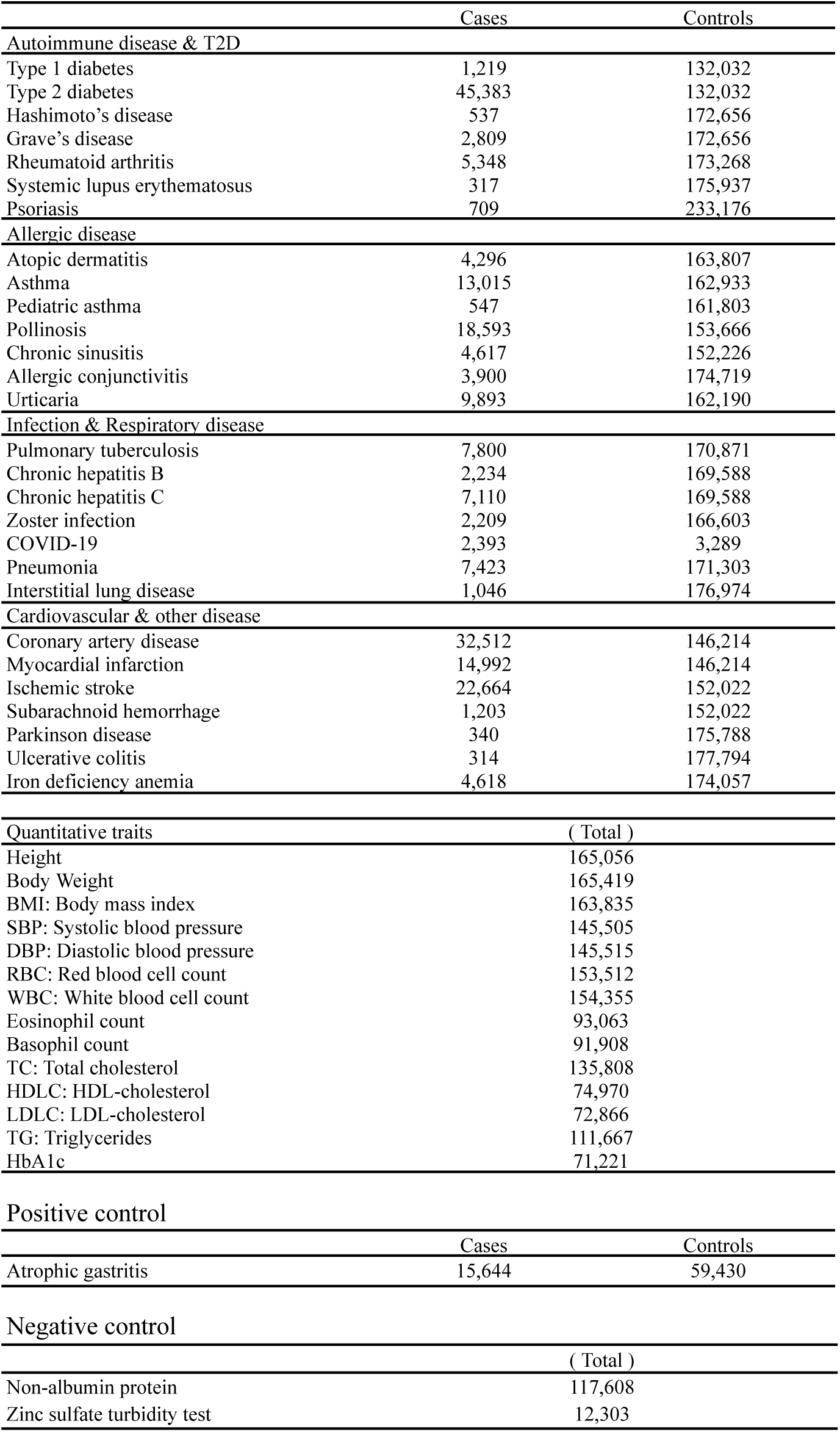
Details about summary statistics for MR outcome traits. The summary statistics used as outcomes were obtained from BBJ data published on PheWeb.jp (https://pheweb.jp/) and from separate GWAS analyses conducted on Psoriasis. A GWAS of atrophic gastritis was performed using the discovery GWAS dataset from the TMM CommCohort.

**Supplementary Table 13.**
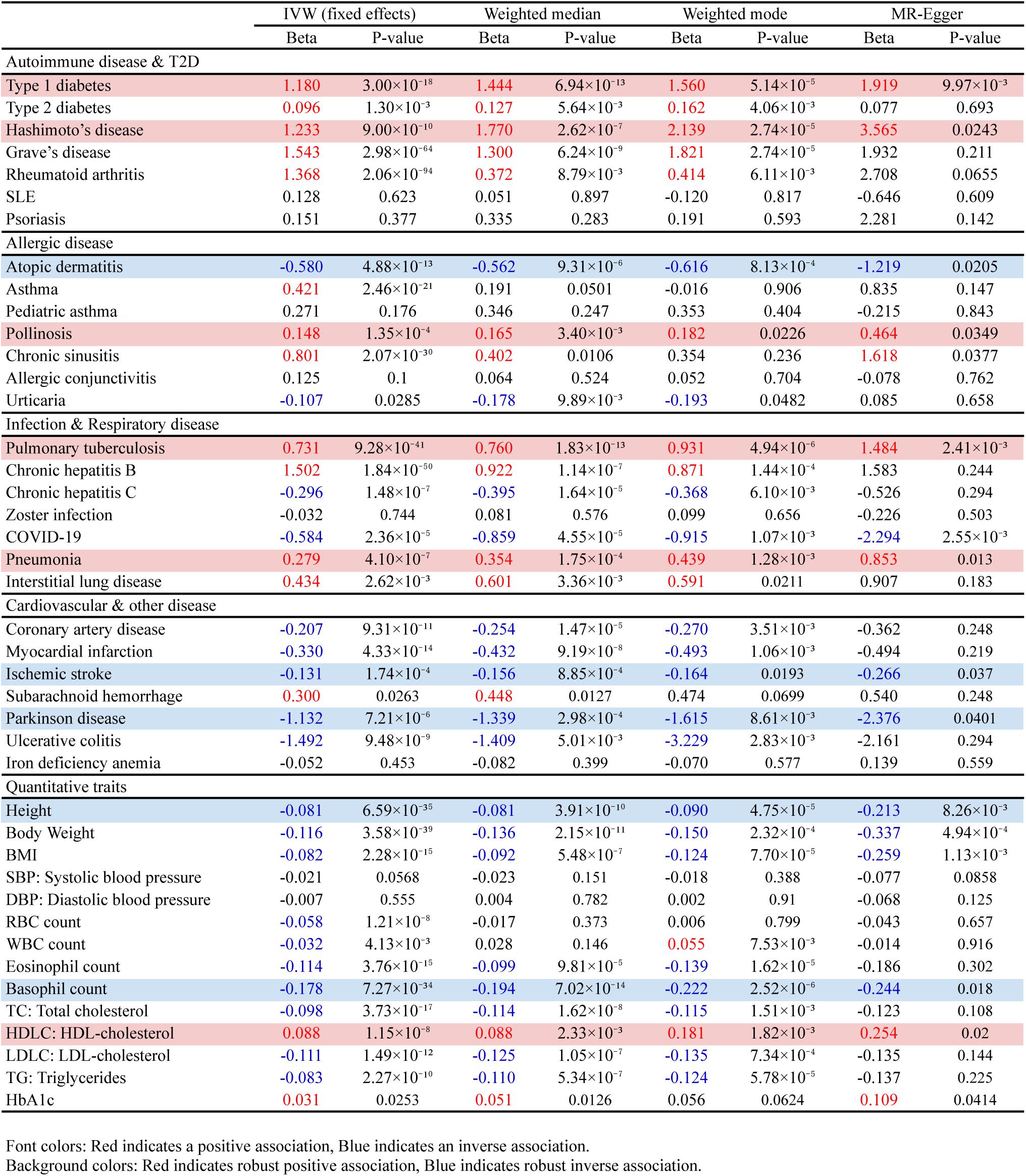
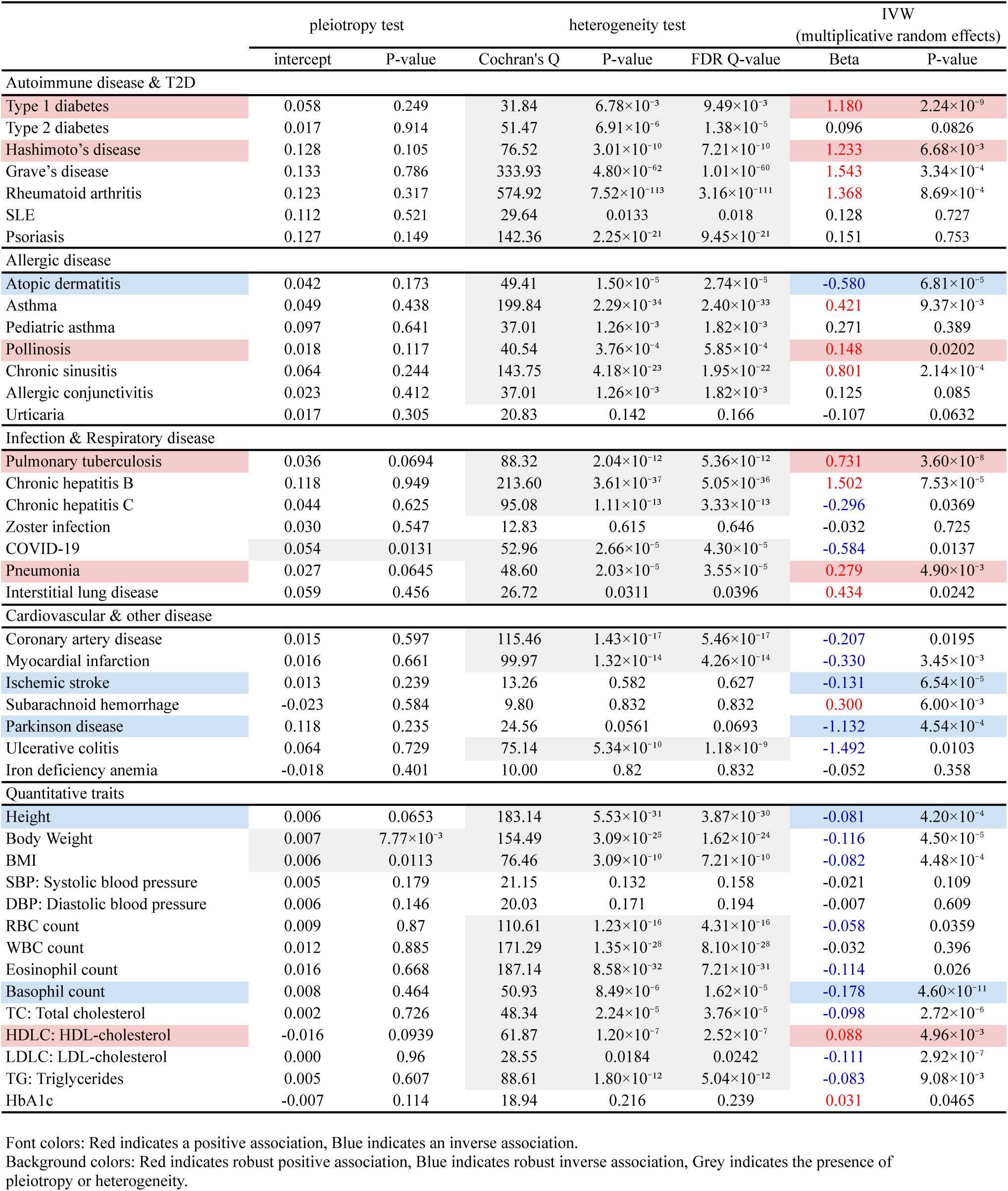
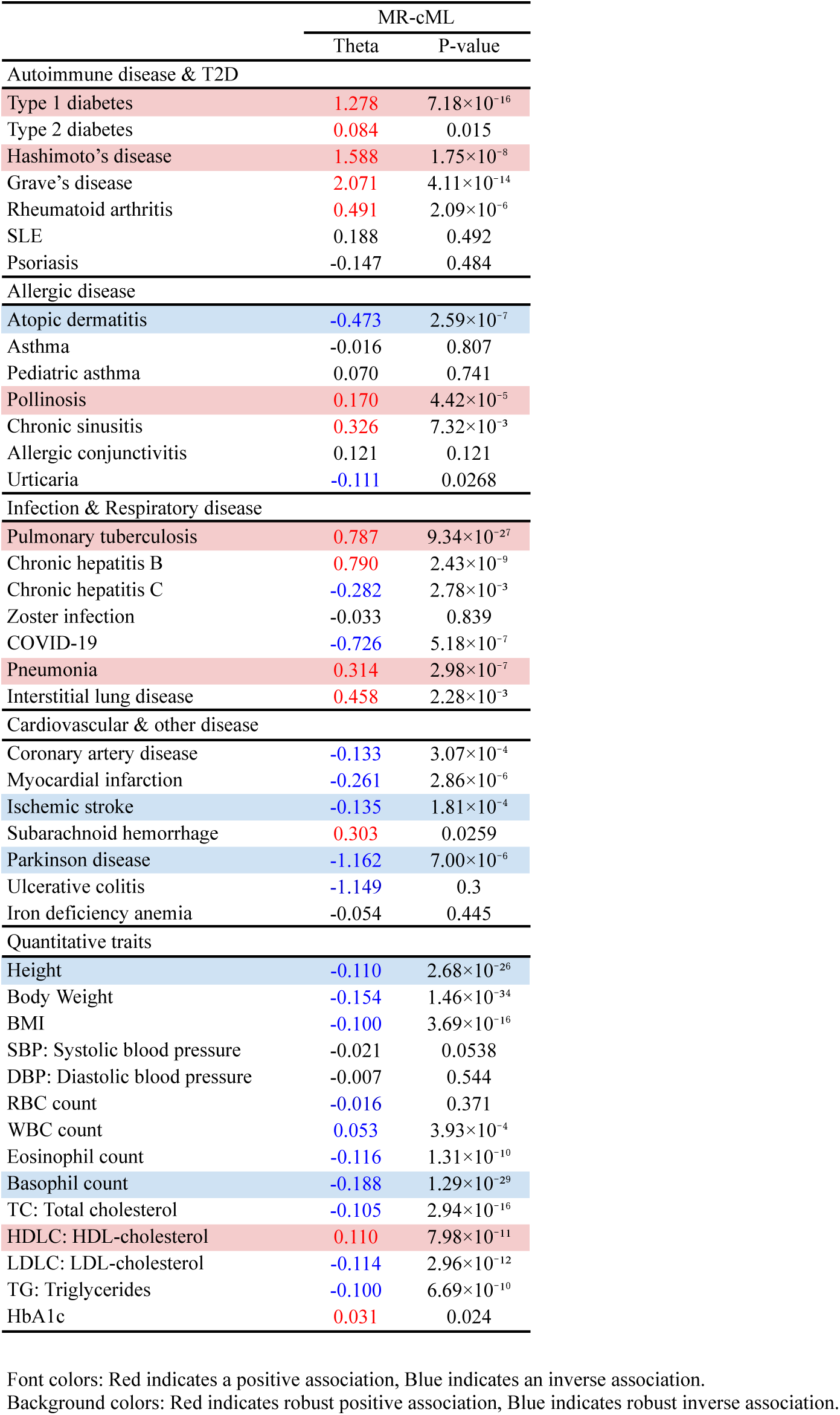
Mendelian randomization analysis.

|  | IVW (fixed effects) |  | Weighted median |  | Weighted mode |  | MR-Egger |  |
| --- | --- | --- | --- | --- | --- | --- | --- | --- |
|  | Beta | P-value | Beta | P-value | Beta | P-value | Beta | P-value |
| **Autoimmune disease & T2D** |  |  |  |  |  |  |  |  |
| Type 1 diabetes | 1.180 | 3.00×10<sup>-18</sup> | 1.444 | 6.94×10<sup>-13</sup> | 1.560 | 5.14×10<sup>-5</sup> | 1.919 | 9.97×10<sup>-3</sup> |
| Type 2 diabetes | 0.096 | 1.30×10<sup>-3</sup> | 0.127 | 5.64×10<sup>-3</sup> | 0.162 | 4.06×10<sup>-3</sup> | 0.077 | 0.693 |
| Hashimoto's disease | 1.233 | 9.00×10<sup>-10</sup> | 1.770 | 2.62×10<sup>-7</sup> | 2.139 | 2.74×10<sup>-5</sup> | 3.565 | 0.0243 |
| Grave's disease | 1.543 | 2.98×10<sup>-64</sup> | 1.300 | 6.24×10<sup>-9</sup> | 1.821 | 2.74×10<sup>-5</sup> | 1.932 | 0.211 |
| Rheumatoid arthritis | 1.368 | 2.06×10<sup>-94</sup> | 0.372 | 8.79×10<sup>-3</sup> | 0.414 | 6.11×10<sup>-3</sup> | 2.708 | 0.0655 |
| SLE | 0.128 | 0.623 | 0.051 | 0.897 | -0.120 | 0.817 | -0.646 | 0.609 |
| Psoriasis | 0.151 | 0.377 | 0.335 | 0.283 | 0.191 | 0.593 | 2.281 | 0.142 |
| **Allergic disease** |  |  |  |  |  |  |  |  |
| Atopic dermatitis | -0.580 | 4.88×10<sup>-13</sup> | -0.562 | 9.31×10<sup>-6</sup> | -0.616 | 8.13×10<sup>-4</sup> | -1.219 | 0.0205 |
| Asthma | 0.421 | 2.46×10<sup>-21</sup> | 0.191 | 0.0501 | -0.016 | 0.906 | 0.835 | 0.147 |
| Pediatric asthma | 0.271 | 0.176 | 0.346 | 0.247 | 0.353 | 0.404 | -0.215 | 0.843 |
| Pollinosis | 0.148 | 1.35×10<sup>-4</sup> | 0.165 | 3.40×10<sup>-3</sup> | 0.182 | 0.0226 | 0.464 | 0.0349 |
| Chronic sinusitis | 0.801 | 2.07×10<sup>-30</sup> | 0.402 | 0.0106 | 0.354 | 0.236 | 1.618 | 0.0377 |
| Allergic conjunctivitis | 0.125 | 0.1 | 0.064 | 0.524 | 0.052 | 0.704 | -0.078 | 0.762 |
| Urticaria | -0.107 | 0.0285 | -0.178 | 9.89×10<sup>-3</sup> | -0.193 | 0.0482 | 0.085 | 0.658 |
| **Infection & Respiratory disease** |  |  |  |  |  |  |  |  |
| Pulmonary tuberculosis | 0.731 | 9.28×10<sup>-41</sup> | 0.760 | 1.83×10<sup>-13</sup> | 0.931 | 4.94×10<sup>-6</sup> | 1.484 | 2.41×10<sup>-3</sup> |
| Chronic hepatitis B | 1.502 | 1.84×10<sup>-50</sup> | 0.922 | 1.14×10<sup>-7</sup> | 0.871 | 1.44×10<sup>-4</sup> | 1.583 | 0.244 |
| Chronic hepatitis C | -0.296 | 1.48×10<sup>-7</sup> | -0.395 | 1.64×10<sup>-5</sup> | -0.368 | 6.10×10<sup>-3</sup> | -0.526 | 0.294 |
| Zoster infection | -0.032 | 0.744 | 0.081 | 0.576 | 0.099 | 0.656 | -0.226 | 0.503 |
| COVID-19 | -0.584 | 2.36×10<sup>-5</sup> | -0.859 | 4.55×10<sup>-5</sup> | -0.915 | 1.07×10<sup>-3</sup> | -2.294 | 2.55×10<sup>-3</sup> |
| Pneumonia | 0.279 | 4.10×10<sup>-7</sup> | 0.354 | 1.75×10<sup>-4</sup> | 0.439 | 1.28×10<sup>-3</sup> | 0.853 | 0.013 |
| Interstitial lung disease | 0.434 | 2.62×10<sup>-3</sup> | 0.601 | 3.36×10<sup>-3</sup> | 0.591 | 0.0211 | 0.907 | 0.183 |
| **Cardiovascular & other disease** |  |  |  |  |  |  |  |  |
| Coronary artery disease | -0.207 | 9.31×10<sup>-11</sup> | -0.254 | 1.47×10<sup>-5</sup> | -0.270 | 3.51×10<sup>-3</sup> | -0.362 | 0.248 |
| Myocardial infarction | -0.330 | 4.33×10<sup>-14</sup> | -0.432 | 9.19×10<sup>-8</sup> | -0.493 | 1.06×10<sup>-3</sup> | -0.494 | 0.219 |
| Ischemic stroke | -0.131 | 1.74×10<sup>-4</sup> | -0.156 | 8.85×10<sup>-4</sup> | -0.164 | 0.0193 | -0.266 | 0.037 |
| Subarachnoid hemorrhage | 0.300 | 0.0263 | 0.448 | 0.0127 | 0.474 | 0.0699 | 0.540 | 0.248 |
| Parkinson disease | -1.132 | 7.21×10<sup>-6</sup> | -1.339 | 2.98×10<sup>-4</sup> | -1.615 | 8.61×10<sup>-3</sup> | -2.376 | 0.0401 |
| Ulcerative colitis | -1.492 | 9.48×10<sup>-9</sup> | -1.409 | 5.01×10<sup>-3</sup> | -3.229 | 2.83×10<sup>-3</sup> | -2.161 | 0.294 |
| Iron deficiency anemia | -0.052 | 0.453 | -0.082 | 0.399 | -0.070 | 0.577 | 0.139 | 0.559 |
| **Quantitative traits** |  |  |  |  |  |  |  |  |
| Height | -0.081 | 6.59×10<sup>-35</sup> | -0.081 | 3.91×10<sup>-10</sup> | -0.090 | 4.75×10<sup>-5</sup> | -0.213 | 8.26×10<sup>-3</sup> |
| Body Weight | -0.116 | 3.58×10<sup>-39</sup> | -0.136 | 2.15×10<sup>-11</sup> | -0.150 | 2.32×10<sup>-4</sup> | -0.337 | 4.94×10<sup>-4</sup> |
| BMI | -0.082 | 2.28×10<sup>-15</sup> | -0.092 | 5.48×10<sup>-7</sup> | -0.124 | 7.70×10<sup>-5</sup> | -0.259 | 1.13×10<sup>-3</sup> |
| SBP: Systolic blood pressure | -0.021 | 0.0568 | -0.023 | 0.151 | -0.018 | 0.388 | -0.077 | 0.0858 |
| DBP: Diastolic blood pressure | -0.007 | 0.555 | 0.004 | 0.782 | 0.002 | 0.91 | -0.068 | 0.125 |
| RBC count | -0.058 | 1.21×10<sup>-8</sup> | -0.017 | 0.373 | 0.006 | 0.799 | -0.043 | 0.657 |
| WBC count | -0.032 | 4.13×10<sup>-3</sup> | 0.028 | 0.146 | 0.055 | 7.53×10<sup>-3</sup> | -0.014 | 0.916 |
| Eosinophil count | -0.114 | 3.76×10<sup>-15</sup> | -0.099 | 9.81×10<sup>-5</sup> | -0.139 | 1.62×10<sup>-5</sup> | -0.186 | 0.302 |
| Basophil count | -0.178 | 7.27×10<sup>-34</sup> | -0.194 | 7.02×10<sup>-14</sup> | -0.222 | 2.52×10<sup>-6</sup> | -0.244 | 0.018 |
| TC: Total cholesterol | -0.098 | 3.73×10<sup>-17</sup> | -0.114 | 1.62×10<sup>-8</sup> | -0.115 | 1.51×10<sup>-3</sup> | -0.123 | 0.108 |
| HDLC: HDL-cholesterol | 0.088 | 1.15×10<sup>-8</sup> | 0.088 | 2.33×10<sup>-3</sup> | 0.181 | 1.82×10<sup>-3</sup> | 0.254 | 0.02 |
| LDLC: LDL-cholesterol | -0.111 | 1.49×10<sup>-12</sup> | -0.125 | 1.05×10<sup>-7</sup> | -0.135 | 7.34×10<sup>-4</sup> | -0.135 | 0.144 |
| TG: Triglycerides | -0.083 | 2.27×10<sup>-10</sup> | -0.110 | 5.34×10<sup>-7</sup> | -0.124 | 5.78×10<sup>-5</sup> | -0.137 | 0.225 |
| HbA1c | 0.031 | 0.0253 | 0.051 | 0.0126 | 0.056 | 0.0624 | 0.109 | 0.0414 |

Font colors: Red indicates a positive association, Blue indicates an inverse association. Background colors: Red indicates robust positive association, Blue indicates robust inverse association.
|  | pleiotropy test |  | heterogeneity test |  |  | IVW<br>(multiplicative random effects) |  |
| --- | --- | --- | --- | --- | --- | --- | --- |
|  | intercept | P-value | Cochran's Q | P-value | FDR Q-value | Beta | P-value |
| Autoimmune disease & T2D |  |  |  |  |  |  |  |
| Type 1 diabetes | 0.058 | 0.249 | 31.84 | $6.78 \times 10^{-3}$ | $9.49 \times 10^{-3}$ | 1.180 | $2.24 \times 10^{-9}$ |
| Type 2 diabetes | 0.017 | 0.914 | 51.47 | $6.91 \times 10^{-6}$ | $1.38 \times 10^{-5}$ | 0.096 | 0.0826 |
| Hashimoto's disease | 0.128 | 0.105 | 76.52 | $3.01 \times 10^{-10}$ | $7.21 \times 10^{-10}$ | 1.233 | $6.68 \times 10^{-3}$ |
| Grave's disease | 0.133 | 0.786 | 333.93 | $4.80 \times 10^{-62}$ | $1.01 \times 10^{-60}$ | 1.543 | $3.34 \times 10^{-4}$ |
| Rheumatoid arthritis | 0.123 | 0.317 | 574.92 | $7.52 \times 10^{-113}$ | $3.16 \times 10^{-111}$ | 1.368 | $8.69 \times 10^{-4}$ |
| SLE | 0.112 | 0.521 | 29.64 | 0.0133 | 0.018 | 0.128 | 0.727 |
| Psoriasis | 0.127 | 0.149 | 142.36 | $2.25 \times 10^{-21}$ | $9.45 \times 10^{-21}$ | 0.151 | 0.753 |
| Allergic disease |  |  |  |  |  |  |  |
| Atopic dermatitis | 0.042 | 0.173 | 49.41 | $1.50 \times 10^{-5}$ | $2.74 \times 10^{-5}$ | -0.580 | $6.81 \times 10^{-5}$ |
| Asthma | 0.049 | 0.438 | 199.84 | $2.29 \times 10^{-34}$ | $2.40 \times 10^{-33}$ | 0.421 | $9.37 \times 10^{-3}$ |
| Pediatric asthma | 0.097 | 0.641 | 37.01 | $1.26 \times 10^{-3}$ | $1.82 \times 10^{-3}$ | 0.271 | 0.389 |
| Pollinosis | 0.018 | 0.117 | 40.54 | $3.76 \times 10^{-4}$ | $5.85 \times 10^{-4}$ | 0.148 | 0.0202 |
| Chronic sinusitis | 0.064 | 0.244 | 143.75 | $4.18 \times 10^{-23}$ | $1.95 \times 10^{-22}$ | 0.801 | $2.14 \times 10^{-4}$ |
| Allergic conjunctivitis | 0.023 | 0.412 | 37.01 | $1.26 \times 10^{-3}$ | $1.82 \times 10^{-3}$ | 0.125 | 0.085 |
| Urticaria | 0.017 | 0.305 | 20.83 | 0.142 | 0.166 | -0.107 | 0.0632 |
| Infection & Respiratory disease |  |  |  |  |  |  |  |
| Pulmonary tuberculosis | 0.036 | 0.0694 | 88.32 | $2.04 \times 10^{-12}$ | $5.36 \times 10^{-12}$ | 0.731 | $3.60 \times 10^{-8}$ |
| Chronic hepatitis B | 0.118 | 0.949 | 213.60 | $3.61 \times 10^{-37}$ | $5.05 \times 10^{-36}$ | 1.502 | $7.53 \times 10^{-5}$ |
| Chronic hepatitis C | 0.044 | 0.625 | 95.08 | $1.11 \times 10^{-13}$ | $3.33 \times 10^{-13}$ | -0.296 | 0.0369 |
| Zoster infection | 0.030 | 0.547 | 12.83 | 0.615 | 0.646 | -0.032 | 0.725 |
| COVID-19 | 0.054 | 0.0131 | 52.96 | $2.66 \times 10^{-5}$ | $4.30 \times 10^{-5}$ | -0.584 | 0.0137 |
| Pneumonia | 0.027 | 0.0645 | 48.60 | $2.03 \times 10^{-5}$ | $3.55 \times 10^{-5}$ | 0.279 | $4.90 \times 10^{-3}$ |
| Interstitial lung disease | 0.059 | 0.456 | 26.72 | 0.0311 | 0.0396 | 0.434 | 0.0242 |
| Cardiovascular & other disease |  |  |  |  |  |  |  |
| Coronary artery disease | 0.015 | 0.597 | 115.46 | $1.43 \times 10^{-17}$ | $5.46 \times 10^{-17}$ | -0.207 | 0.0195 |
| Myocardial infarction | 0.016 | 0.661 | 99.97 | $1.32 \times 10^{-14}$ | $4.26 \times 10^{-14}$ | -0.330 | $3.45 \times 10^{-3}$ |
| Ischemic stroke | 0.013 | 0.239 | 13.26 | 0.582 | 0.627 | -0.131 | $6.54 \times 10^{-5}$ |
| Subarachnoid hemorrhage | -0.023 | 0.584 | 9.80 | 0.832 | 0.832 | 0.300 | $6.00 \times 10^{-3}$ |
| Parkinson disease | 0.118 | 0.235 | 24.56 | 0.0561 | 0.0693 | -1.132 | $4.54 \times 10^{-4}$ |
| Ulcerative colitis | 0.064 | 0.729 | 75.14 | $5.34 \times 10^{-10}$ | $1.18 \times 10^{-9}$ | -1.492 | 0.0103 |
| Iron deficiency anemia | -0.018 | 0.401 | 10.00 | 0.82 | 0.832 | -0.052 | 0.358 |
| Quantitative traits |  |  |  |  |  |  |  |
| Height | 0.006 | 0.0653 | 183.14 | $5.53 \times 10^{-31}$ | $3.87 \times 10^{-30}$ | -0.081 | $4.20 \times 10^{-4}$ |
| Body Weight | 0.007 | $7.77 \times 10^{-3}$ | 154.49 | $3.09 \times 10^{-25}$ | $1.62 \times 10^{-24}$ | -0.116 | $4.50 \times 10^{-5}$ |
| BMI | 0.006 | 0.0113 | 76.46 | $3.09 \times 10^{-10}$ | $7.21 \times 10^{-10}$ | -0.082 | $4.48 \times 10^{-4}$ |
| SBP: Systolic blood pressure | 0.005 | 0.179 | 21.15 | 0.132 | 0.158 | -0.021 | 0.109 |
| DBP: Diastolic blood pressure | 0.006 | 0.146 | 20.03 | 0.171 | 0.194 | -0.007 | 0.609 |
| RBC count | 0.009 | 0.87 | 110.61 | $1.23 \times 10^{-16}$ | $4.31 \times 10^{-16}$ | -0.058 | 0.0359 |
| WBC count | 0.012 | 0.885 | 171.29 | $1.35 \times 10^{-28}$ | $8.10 \times 10^{-28}$ | -0.032 | 0.396 |
| Eosinophil count | 0.016 | 0.668 | 187.14 | $8.58 \times 10^{-32}$ | $7.21 \times 10^{-31}$ | -0.114 | 0.026 |
| Basophil count | 0.008 | 0.464 | 50.93 | $8.49 \times 10^{-6}$ | $1.62 \times 10^{-5}$ | -0.178 | $4.60 \times 10^{-11}$ |
| TC: Total cholesterol | 0.002 | 0.726 | 48.34 | $2.24 \times 10^{-5}$ | $3.76 \times 10^{-5}$ | -0.098 | $2.72 \times 10^{-6}$ |
| HDLC: HDL-cholesterol | -0.016 | 0.0939 | 61.87 | $1.20 \times 10^{-7}$ | $2.52 \times 10^{-7}$ | 0.088 | $4.96 \times 10^{-3}$ |
| LDLC: LDL-cholesterol | 0.000 | 0.96 | 28.55 | 0.0184 | 0.0242 | -0.111 | $2.92 \times 10^{-7}$ |
| TG: Triglycerides | 0.005 | 0.607 | 88.61 | $1.80 \times 10^{-12}$ | $5.04 \times 10^{-12}$ | -0.083 | $9.08 \times 10^{-3}$ |
| HbA1c | -0.007 | 0.114 | 18.94 | 0.216 | 0.239 | 0.031 | 0.0465 |

|  | MR-cML |  |
| --- | --- | --- |
|  | Theta | P-value |
| Autoimmune disease & T2D |  |  |
| Type 1 diabetes | 1.278 | $7.18 \times 10^{-16}$ |
| Type 2 diabetes | 0.084 | 0.015 |
| Hashimoto's disease | 1.588 | $1.75 \times 10^{-8}$ |
| Grave's disease | 2.071 | $4.11 \times 10^{-14}$ |
| Rheumatoid arthritis | 0.491 | $2.09 \times 10^{-6}$ |
| SLE | 0.188 | 0.492 |
| Psoriasis | -0.147 | 0.484 |
| Allergic disease |  |  |
| Atopic dermatitis | -0.473 | $2.59 \times 10^{-7}$ |
| Asthma | -0.016 | 0.807 |
| Pediatric asthma | 0.070 | 0.741 |
| Pollinosis | 0.170 | $4.42 \times 10^{-5}$ |
| Chronic sinusitis | 0.326 | $7.32 \times 10^{-3}$ |
| Allergic conjunctivitis | 0.121 | 0.121 |
| Urticaria | -0.111 | 0.0268 |
| Infection & Respiratory disease |  |  |
| Pulmonary tuberculosis | 0.787 | $9.34 \times 10^{-27}$ |
| Chronic hepatitis B | 0.790 | $2.43 \times 10^{-9}$ |
| Chronic hepatitis C | -0.282 | $2.78 \times 10^{-3}$ |
| Zoster infection | -0.033 | 0.839 |
| COVID-19 | -0.726 | $5.18 \times 10^{-7}$ |
| Pneumonia | 0.314 | $2.98 \times 10^{-7}$ |
| Interstitial lung disease | 0.458 | $2.28 \times 10^{-3}$ |
| Cardiovascular & other disease |  |  |
| Coronary artery disease | -0.133 | $3.07 \times 10^{-4}$ |
| Myocardial infarction | -0.261 | $2.86 \times 10^{-6}$ |
| Ischemic stroke | -0.135 | $1.81 \times 10^{-4}$ |
| Subarachnoid hemorrhage | 0.303 | 0.0259 |
| Parkinson disease | -1.162 | $7.00 \times 10^{-6}$ |
| Ulcerative colitis | -1.149 | 0.3 |
| Iron deficiency anemia | -0.054 | 0.445 |
| Quantitative traits |  |  |
| Height | -0.110 | $2.68 \times 10^{-26}$ |
| Body Weight | -0.154 | $1.46 \times 10^{-34}$ |
| BMI | -0.100 | $3.69 \times 10^{-16}$ |
| SBP: Systolic blood pressure | -0.021 | 0.0538 |
| DBP: Diastolic blood pressure | -0.007 | 0.544 |
| RBC count | -0.016 | 0.371 |
| WBC count | 0.053 | $3.93 \times 10^{-4}$ |
| Eosinophil count | -0.116 | $1.31 \times 10^{-10}$ |
| Basophil count | -0.188 | $1.29 \times 10^{-29}$ |
| TC: Total cholesterol | -0.105 | $2.94 \times 10^{-16}$ |
| HDLC: HDL-cholesterol | 0.110 | $7.98 \times 10^{-11}$ |
| LDLC: LDL-cholesterol | -0.114 | $2.96 \times 10^{-12}$ |
| TG: Triglycerides | -0.100 | $6.69 \times 10^{-10}$ |
| HbA1c | 0.031 | 0.024 |

Font colors: Red indicates a positive association, Blue indicates an inverse association.
Background colors: Red indicates robust positive association, Blue indicates robust inverse association.

**Supplementary Table 14.**
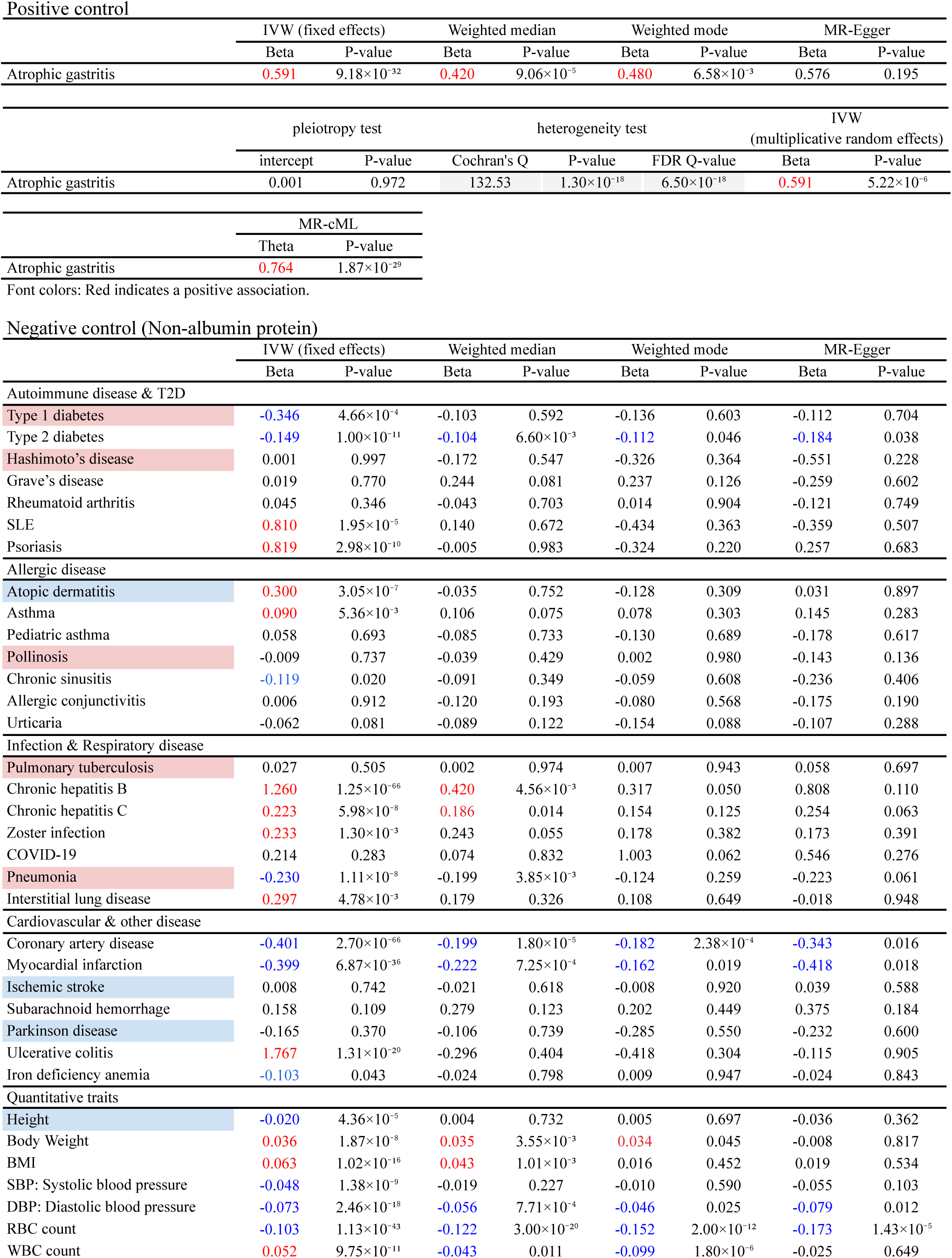
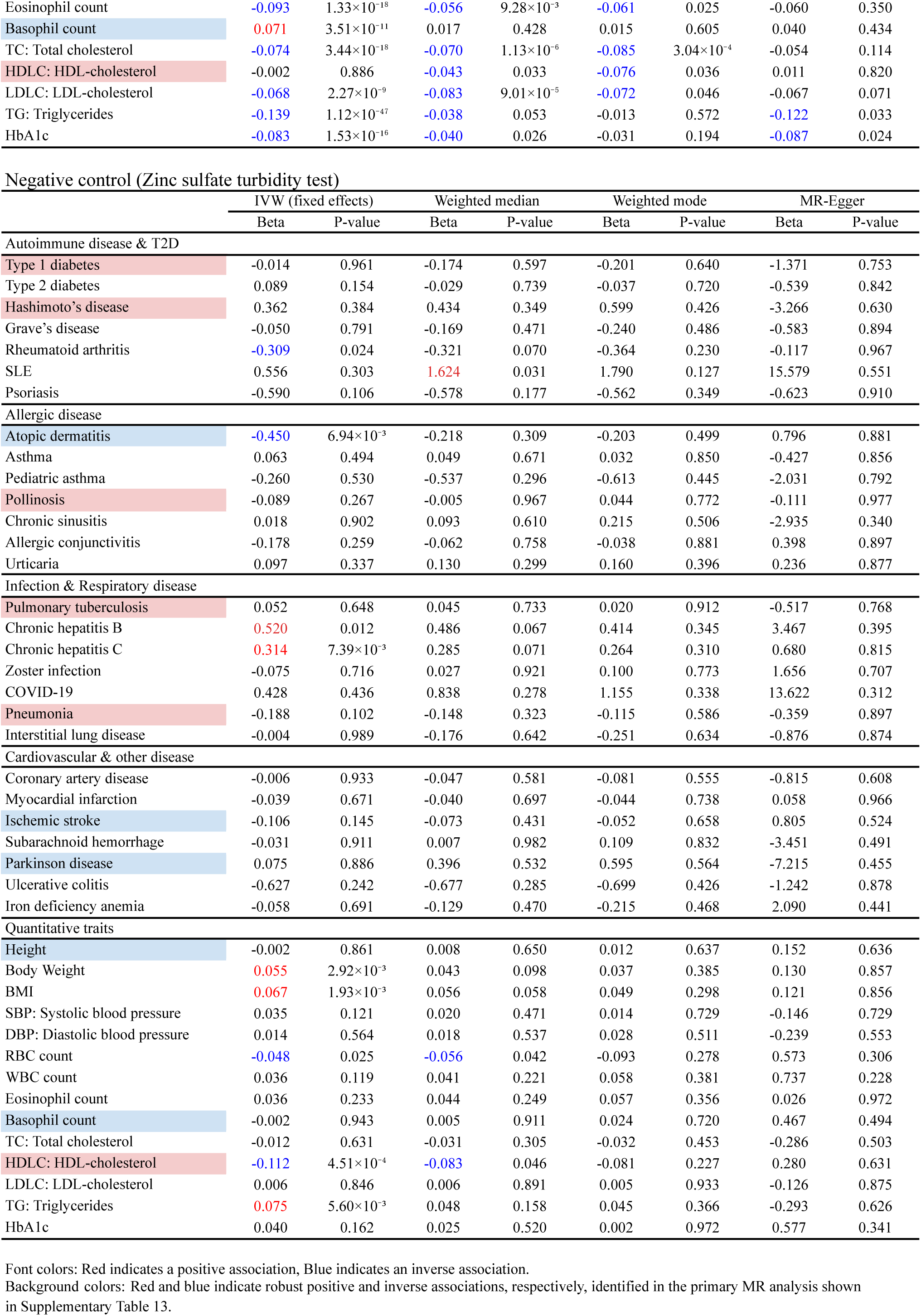
Mendelian randomization analysis (negative and positive control).

**Supplementary Table 15.**
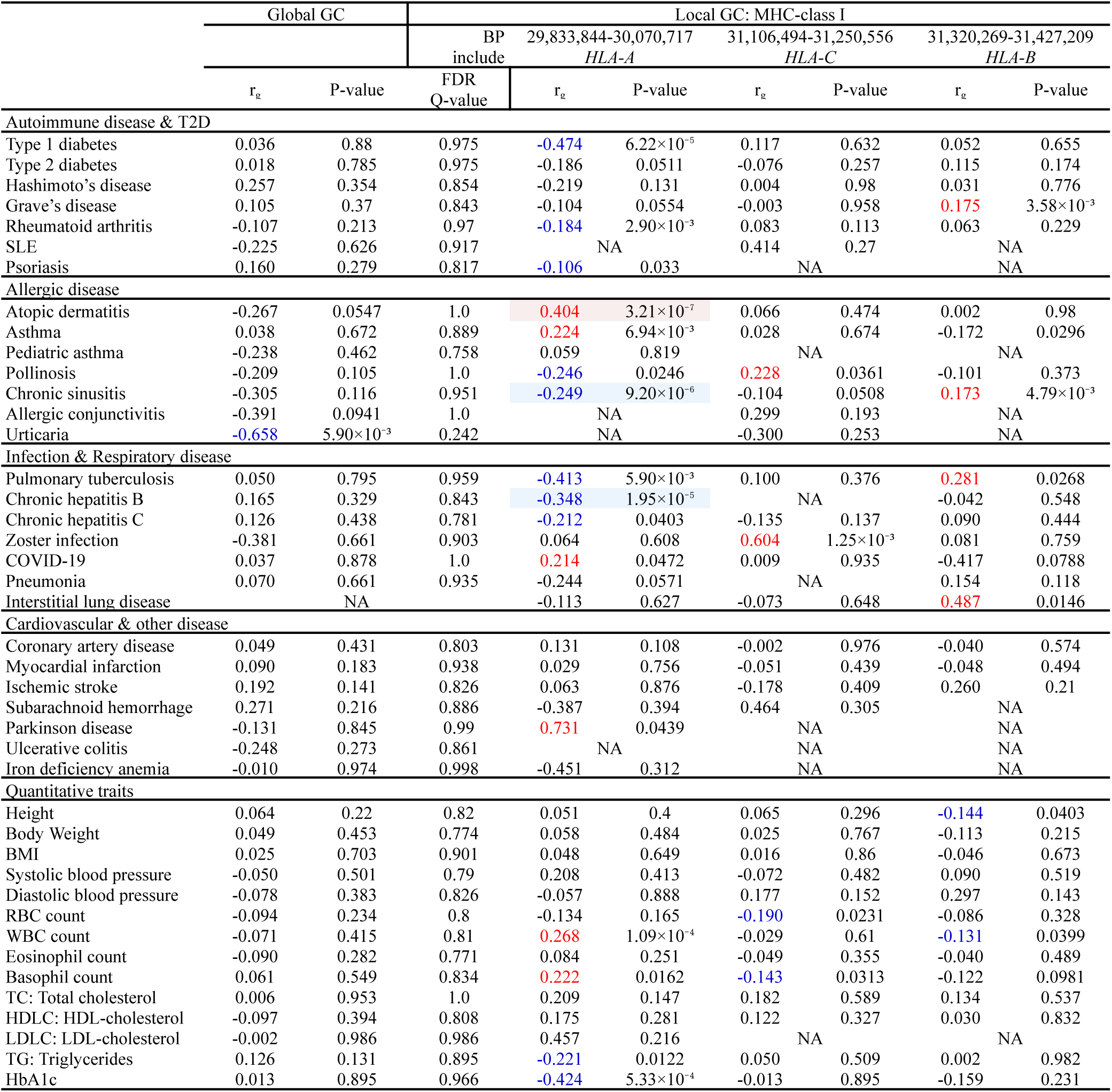
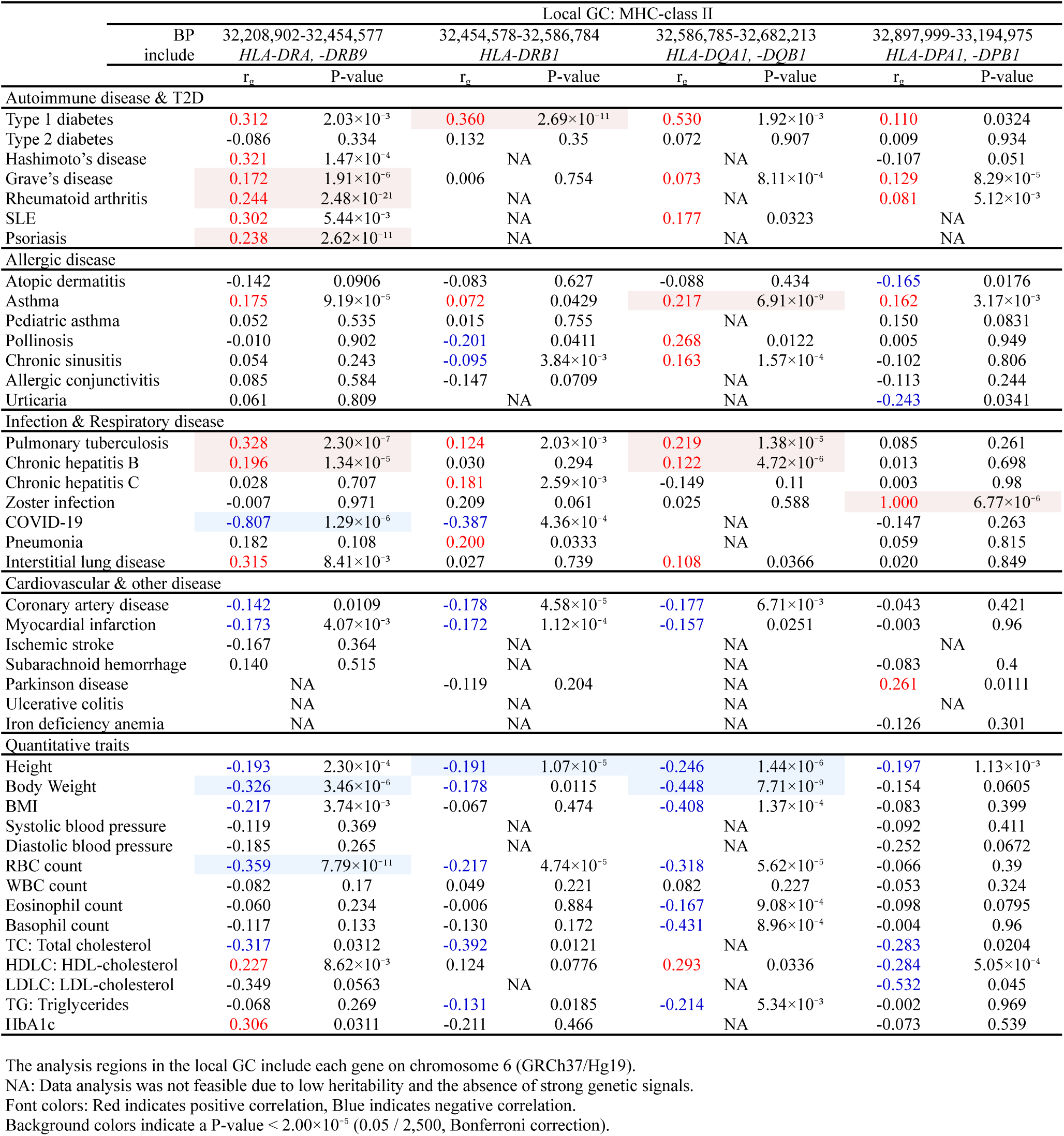
Genetic Correlation analysis / Local Genetic Correlation analysis (MHC region).

**Supplementary Table 16.**
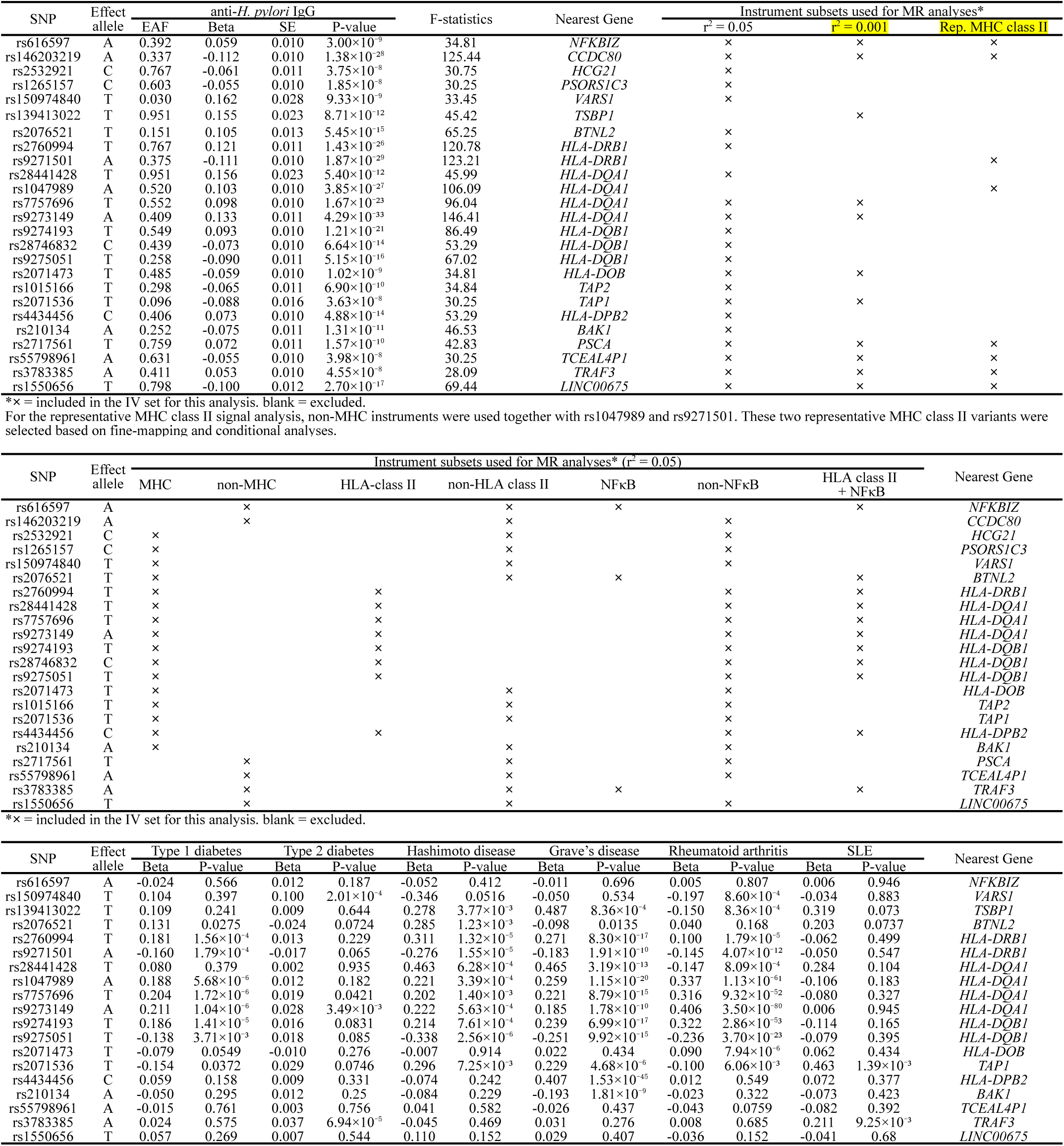
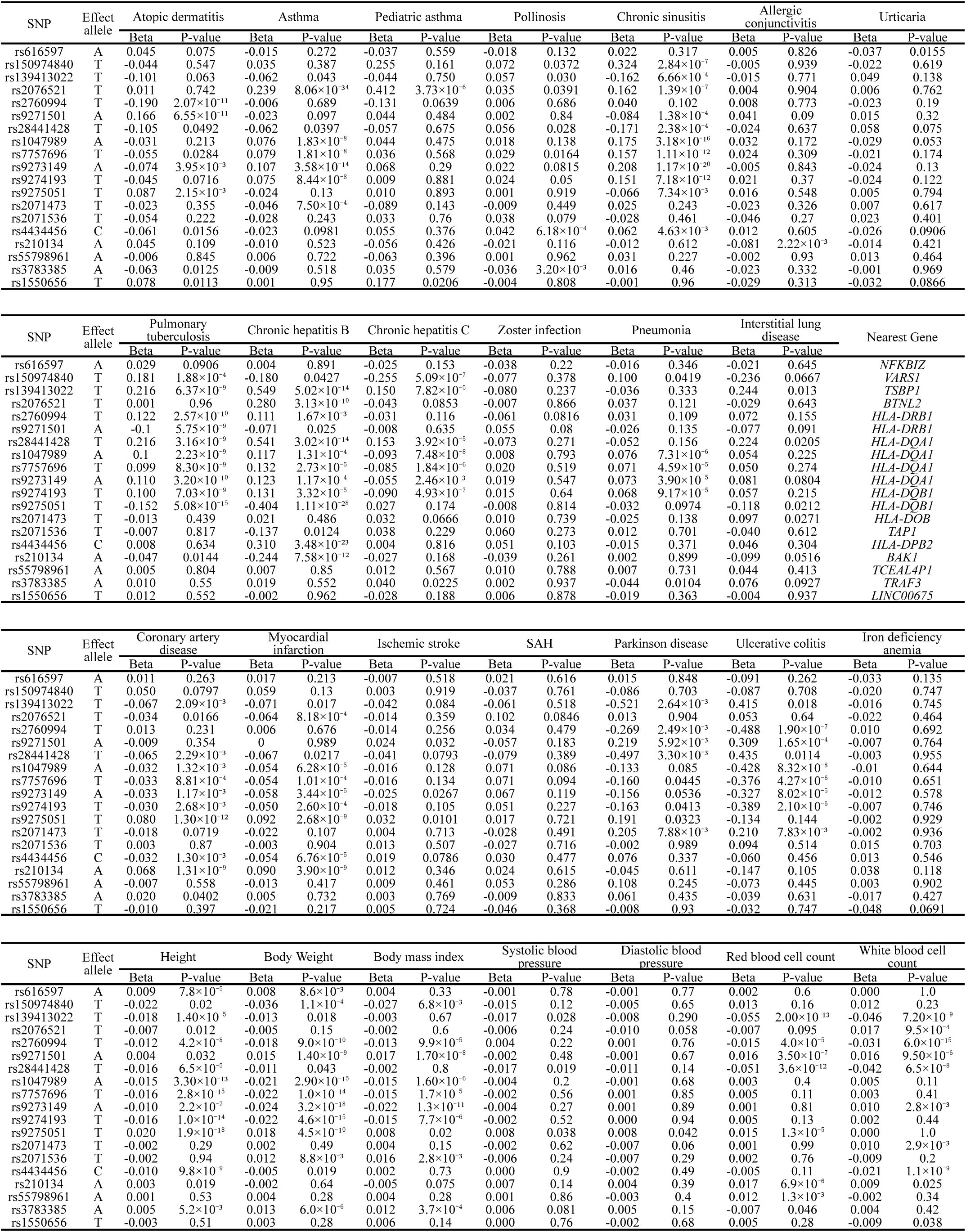
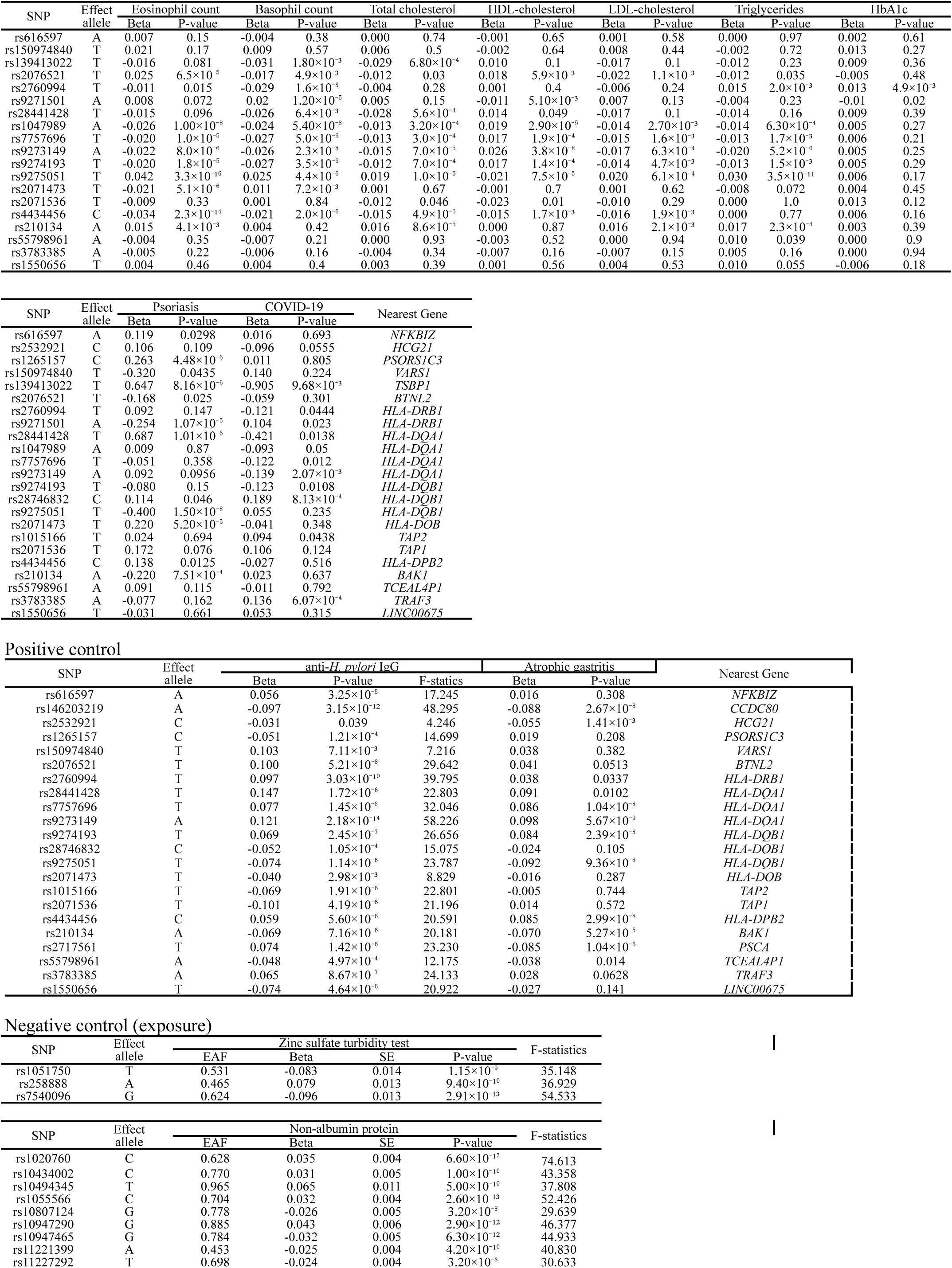
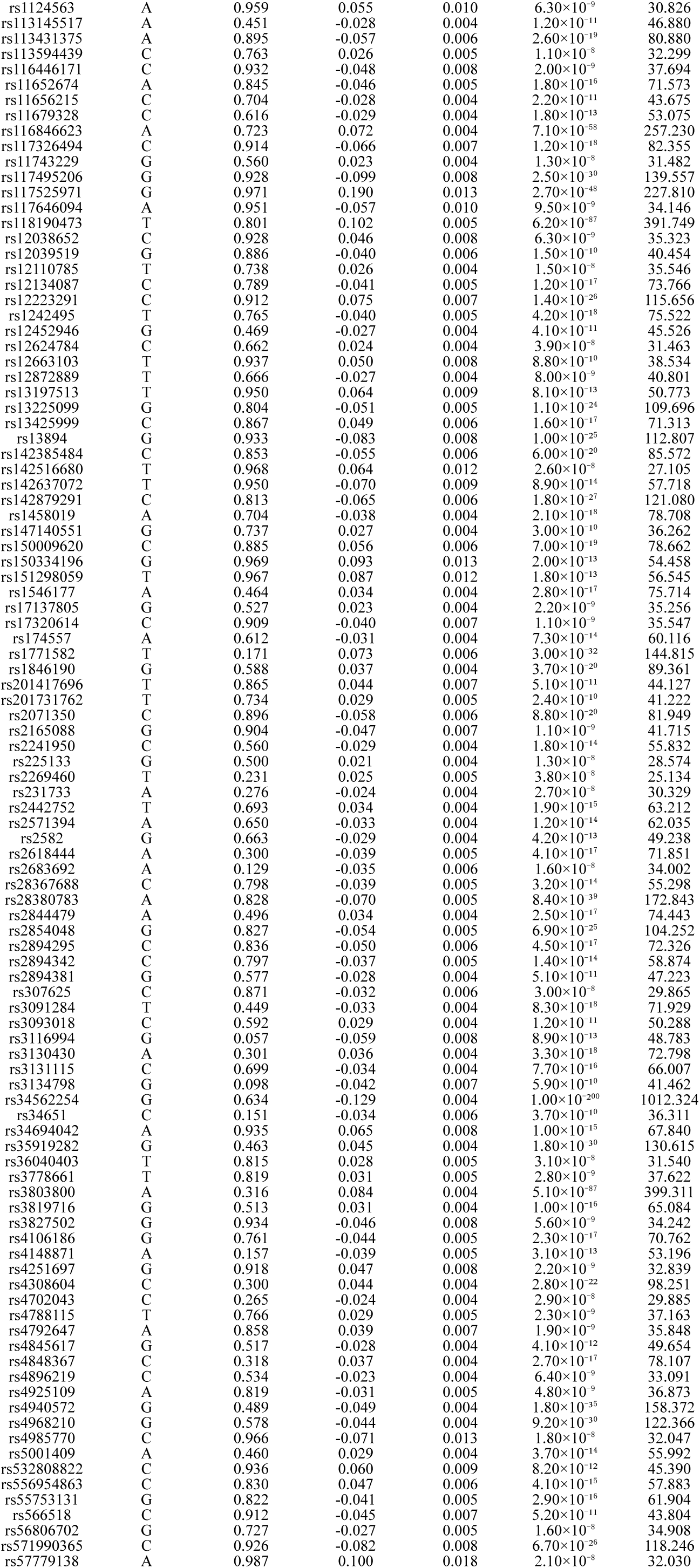
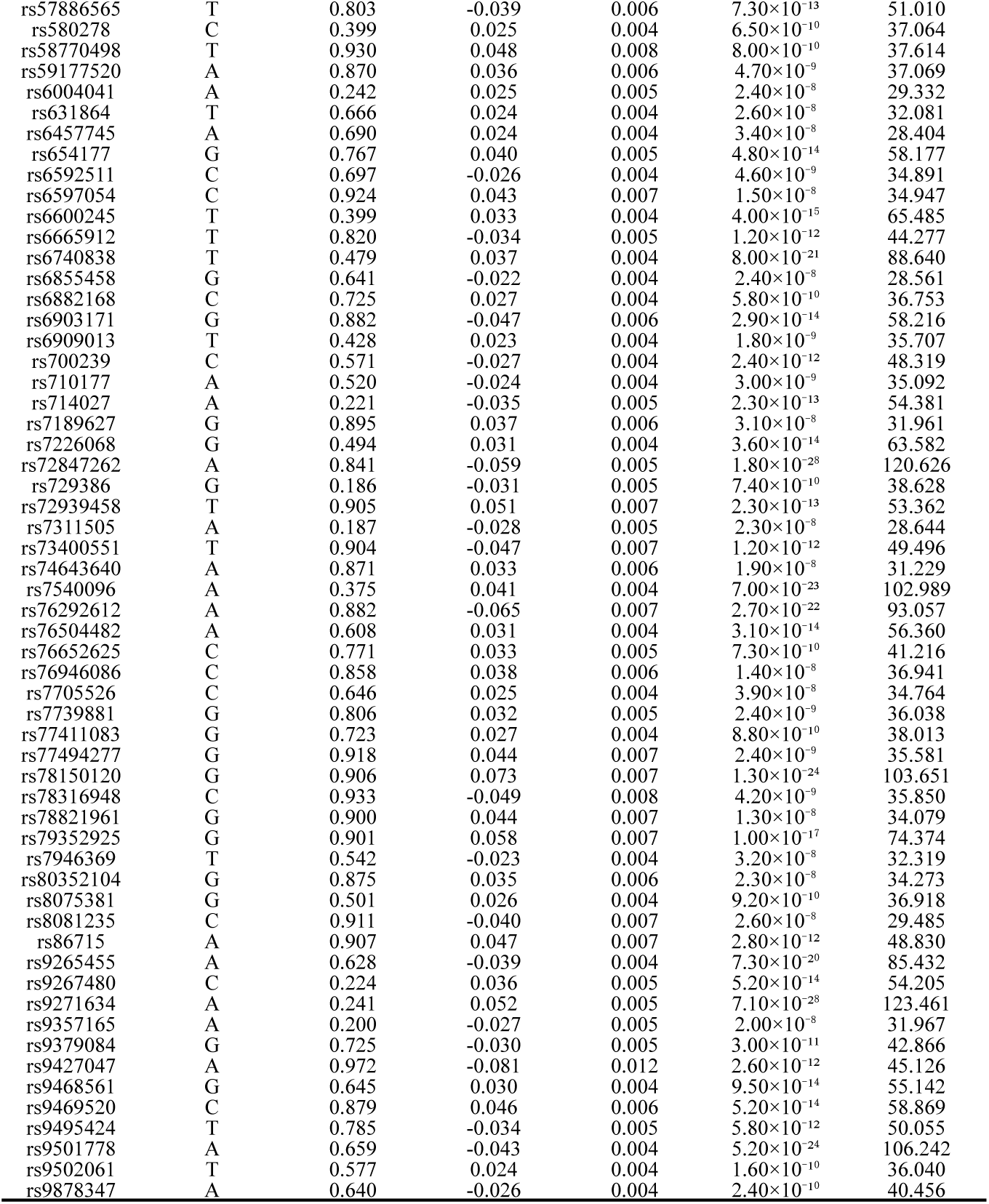
Variants used in the Mendelian randomization analysis and the comparison of their effect sizes. Of the 22 SNPs selected as instrumental variables through pruning from the GWAS summary statistics of Anti-*H. pylori* IgG, 16 overlapped with the summary statistics from BBJ and were used for most traits. For psoriasis and COVID-19, 20 SNPs were included.

**Supplementary Table 17.**
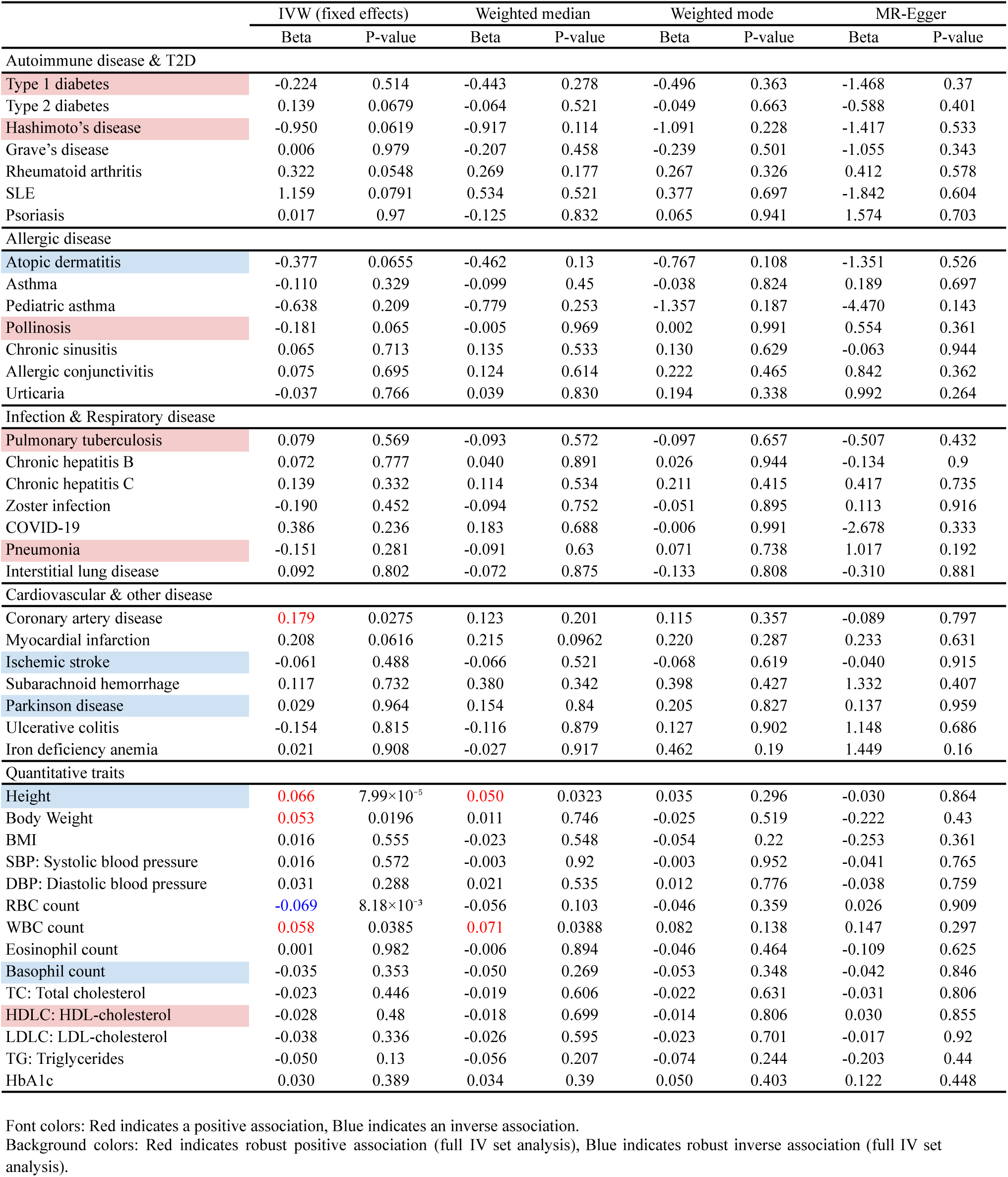
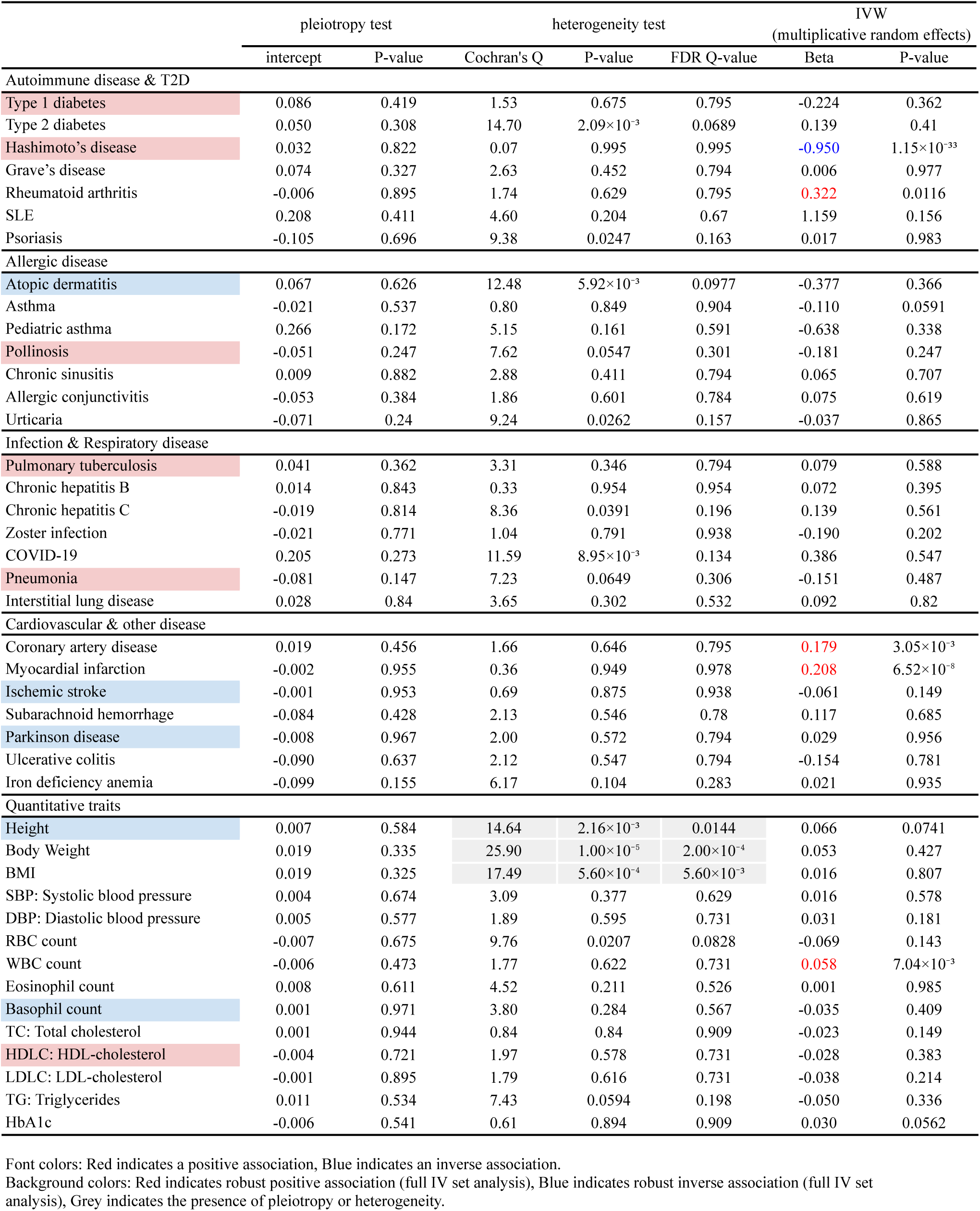
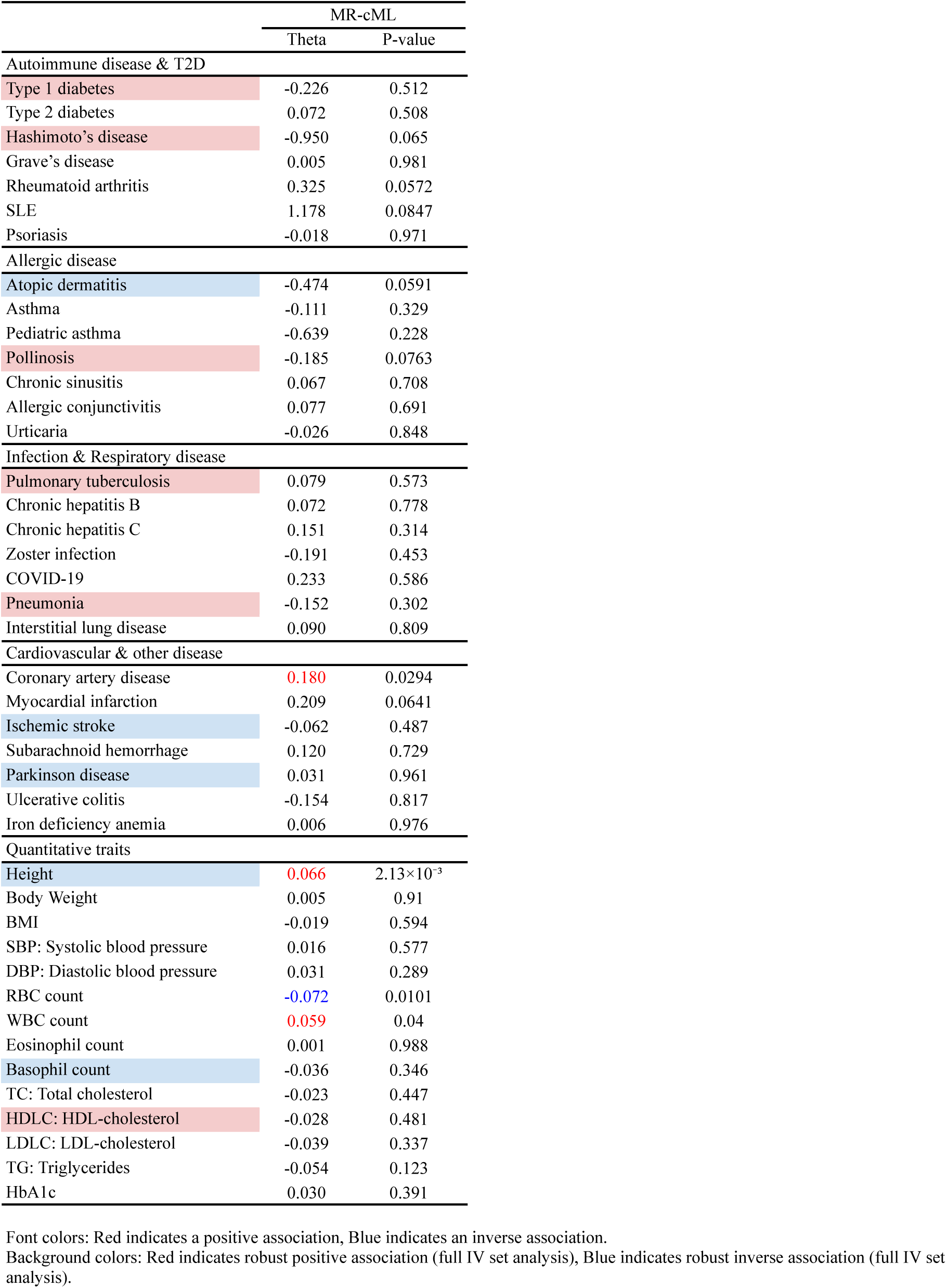
MR excluding extended MHC variants.

**Supplementary Table 18.** MR restricted to extended MHC variants.

|  | IVW (fixed effects) |  | Weighted median |  | Weighted mode |  | MR-Egger |  |
| --- | --- | --- | --- | --- | --- | --- | --- | --- |
|  | Beta | P-value | Beta | P-value | Beta | P-value | Beta | P-value |
| **Autoimmune disease & T2D** |  |  |  |  |  |  |  |  |
| Type 1 diabetes | 1.438 | 1.67×10<sup>-22</sup> | 1.527 | 1.05×10<sup>-12</sup> | 1.561 | 2.29×10<sup>-4</sup> | 1.425 | 0.0349 |
| Type 2 diabetes | 0.089 | 6.61×10<sup>-3</sup> | 0.146 | 3.24×10<sup>-3</sup> | 0.168 | 7.88×10<sup>-3</sup> | 0.316 | 0.197 |
| Hashimoto's disease | 1.637 | 7.80×10<sup>-14</sup> | 2.020 | 2.41×10<sup>-9</sup> | 2.173 | 4.95×10<sup>-5</sup> | 3.282 | 0.115 |
| Grave's disease | 1.830 | 7.83×10<sup>-76</sup> | 1.946 | 1.07×10<sup>-24</sup> | 2.198 | 8.69×10<sup>-7</sup> | 0.955 | 0.645 |
| Rheumatoid arthritis | 1.562 | 1.20×10<sup>-103</sup> | 0.540 | 1.00×10<sup>-3</sup> | 0.410 | 0.0504 | 2.655 | 0.212 |
| SLE | -0.062 | 0.826 | -0.103 | 0.799 | -0.291 | 0.547 | 0.686 | 0.679 |
| Psoriasis | 0.174 | 0.347 | 0.003 | 0.993 | 0.090 | 0.788 | 2.833 | 0.151 |
| **Allergic disease** |  |  |  |  |  |  |  |  |
| Atopic dermatitis | -0.617 | 1.51×10<sup>-12</sup> | -0.561 | 1.53×10<sup>-5</sup> | -0.580 | 2.89×10<sup>-3</sup> | -1.227 | 0.0711 |
| Asthma | 0.519 | 6.49×10<sup>-27</sup> | 0.289 | 0.017 | 0.493 | 0.0565 | 0.526 | 0.514 |
| Pediatric asthma | 0.438 | 0.0443 | 0.384 | 0.21 | 0.345 | 0.379 | -0.413 | 0.771 |
| Pollinosis | 0.208 | 7.75×10<sup>-7</sup> | 0.220 | 2.70×10<sup>-4</sup> | 0.235 | 0.0131 | 0.161 | 0.494 |
| Chronic sinusitis | 0.936 | 7.80×10<sup>-35</sup> | 0.810 | 1.15×10<sup>-5</sup> | 1.395 | 1.17×10<sup>-3</sup> | 1.537 | 0.156 |
| Allergic conjunctivitis | 0.134 | 0.105 | 0.057 | 0.603 | 0.023 | 0.871 | -0.508 | 0.142 |
| Urticaria | -0.120 | 0.0242 | -0.180 | 0.0135 | -0.191 | 0.0509 | 0.007 | 0.974 |
| **Infection & Respiratory disease** |  |  |  |  |  |  |  |  |
| Pulmonary tuberculosis | 0.850 | 2.25×10<sup>-46</sup> | 0.903 | 6.83×10<sup>-18</sup> | 0.955 | 8.14×10<sup>-6</sup> | 1.574 | 0.0109 |
| Chronic hepatitis B | 1.769 | 1.13×10<sup>-58</sup> | 1.138 | 1.19×10<sup>-9</sup> | 1.104 | 4.19×10<sup>-5</sup> | 0.224 | 0.898 |
| Chronic hepatitis C | -0.376 | 8.72×10<sup>-10</sup> | -0.408 | 5.72×10<sup>-5</sup> | -0.370 | 6.84×10<sup>-3</sup> | -0.341 | 0.613 |
| Zoster infection | -0.004 | 0.974 | 0.121 | 0.437 | 0.125 | 0.562 | -0.625 | 0.165 |
| COVID-19 | -0.797 | 1.75×10<sup>-7</sup> | -1.020 | 1.05×10<sup>-5</sup> | -1.023 | 2.78×10<sup>-3</sup> | -1.784 | 0.0329 |
| Pneumonia | 0.358 | 2.37×10<sup>-9</sup> | 0.379 | 8.22×10<sup>-5</sup> | 0.490 | 1.61×10<sup>-3</sup> | 0.520 | 0.222 |
| Interstitial lung disease | 0.497 | 1.55×10<sup>-3</sup> | 0.604 | 4.90×10<sup>-3</sup> | 0.597 | 0.0297 | 1.041 | 0.265 |
| **Cardiovascular & other disease** |  |  |  |  |  |  |  |  |
| Coronary artery disease | -0.277 | 1.37×10<sup>-15</sup> | -0.304 | 8.09×10<sup>-8</sup> | -0.311 | 1.02×10<sup>-3</sup> | -0.012 | 0.976 |
| Myocardial infarction | -0.428 | 2.08×10<sup>-19</sup> | -0.463 | 7.51×10<sup>-10</sup> | -0.494 | 6.09×10<sup>-4</sup> | -0.082 | 0.867 |
| Ischemic stroke | -0.144 | 1.53×10<sup>-4</sup> | -0.168 | 9.06×10<sup>-4</sup> | -0.169 | 0.0215 | -0.307 | 0.0688 |
| Subarachnoid hemorrhage | 0.334 | 0.0232 | 0.484 | 0.0111 | 0.488 | 0.0697 | 0.183 | 0.755 |
| Parkinson disease | -1.346 | 9.47×10<sup>-7</sup> | -1.538 | 8.78×10<sup>-5</sup> | -1.645 | 8.07×10<sup>-3</sup> | -2.189 | 0.155 |
| Ulcerative colitis | -1.740 | 7.83×10<sup>-10</sup> | -2.046 | 1.75×10<sup>-4</sup> | -3.239 | 3.28×10<sup>-4</sup> | -1.693 | 0.566 |
| Iron deficiency anemia | -0.066 | 0.386 | -0.083 | 0.387 | -0.076 | 0.559 | -0.054 | 0.859 |
| **Quantitative traits** |  |  |  |  |  |  |  |  |
| Height | -0.107 | 2.31×10<sup>-51</sup> | -0.094 | 4.11×10<sup>-12</sup> | -0.092 | 1.04×10<sup>-4</sup> | -0.139 | 0.0976 |
| Body Weight | -0.146 | 2.07×10<sup>-52</sup> | -0.154 | 1.80×10<sup>-16</sup> | -0.182 | 3.08×10<sup>-5</sup> | -0.258 | 0.0127 |
| BMI | -0.100 | 7.21×10<sup>-19</sup> | -0.116 | 2.90×10<sup>-9</sup> | -0.143 | 4.49×10<sup>-5</sup> | -0.218 | 0.0237 |
| SBP: Systolic blood pressure | -0.027 | 0.0207 | -0.025 | 0.162 | -0.023 | 0.338 | -0.066 | 0.268 |
| DBP: Diastolic blood pressure | -0.014 | 0.272 | 0.003 | 0.838 | 0.002 | 0.916 | -0.057 | 0.355 |
| RBC count | -0.056 | 4.04×10<sup>-7</sup> | -0.005 | 0.8 | 0.009 | 0.722 | -0.069 | 0.62 |
| WBC count | -0.048 | 6.20×10<sup>-5</sup> | 0.027 | 0.224 | 0.052 | 0.031 | 0.066 | 0.723 |
| Eosinophil count | -0.135 | 1.11×10<sup>-17</sup> | -0.148 | 1.38×10<sup>-8</sup> | -0.156 | 4.90×10<sup>-5</sup> | -0.094 | 0.716 |
| Basophil count | -0.205 | 1.66×10<sup>-37</sup> | -0.209 | 9.71×10<sup>-17</sup> | -0.228 | 9.93×10<sup>-6</sup> | -0.145 | 0.195 |
| TC: Total cholesterol | -0.112 | 9.91×10<sup>-19</sup> | -0.116 | 1.40×10<sup>-8</sup> | -0.128 | 5.14×10<sup>-4</sup> | -0.062 | 0.536 |
| HDLC: HDL-cholesterol | 0.109 | 7.36×10<sup>-11</sup> | 0.172 | 3.69×10<sup>-9</sup> | 0.180 | 2.68×10<sup>-4</sup> | 0.247 | 0.101 |
| LDLC: LDL-cholesterol | -0.124 | 3.19×10<sup>-13</sup> | -0.134 | 6.07×10<sup>-8</sup> | -0.142 | 9.45×10<sup>-4</sup> | -0.049 | 0.619 |
| TG: Triglycerides | -0.089 | 4.09×10<sup>-10</sup> | -0.126 | 7.60×10<sup>-8</sup> | -0.128 | 2.42×10<sup>-4</sup> | -0.102 | 0.528 |
| HbA1c | 0.031 | 0.039 | 0.052 | 0.0161 | 0.060 | 0.0443 | 0.142 | 0.0691 |

|  | pleiotropy test |  | heterogeneity test |  |  | IVW<br>(multiplicative random effects) |  |
| --- | --- | --- | --- | --- | --- | --- | --- |
|  | intercept | P-value | Cochran's Q | P-value | FDR Q-value | Beta | P-value |
| Autoimmune disease & T2D |  |  |  |  |  |  |  |
| Type 1 diabetes | 0.001 | 0.982 | 10.51 | 0.485 | 0.534 | 1.438 | 1.78×10<sup>-23</sup> |
| Type 2 diabetes | -0.023 | 0.328 | 36.39 | 1.46×10<sup>-4</sup> | 3.00×10<sup>-4</sup> | 0.089 | 0.135 |
| Hashimoto's disease | -0.168 | 0.392 | 54.63 | 9.04×10<sup>-8</sup> | 2.49×10<sup>-7</sup> | 1.637 | 7.98×10<sup>-4</sup> |
| Grave's disease | 0.089 | 0.662 | 278.13 | 3.50×10<sup>-53</sup> | 5.77×10<sup>-52</sup> | 1.830 | 2.48×10<sup>-4</sup> |
| Rheumatoid arthritis | -0.112 | 0.583 | 527.08 | 5.40×10<sup>-106</sup> | 1.80×10<sup>-104</sup> | 1.562 | 1.79×10<sup>-3</sup> |
| SLE | -0.076 | 0.641 | 22.15 | 0.0232 | 0.0333 | -0.062 | 0.877 |
| Psoriasis | -0.254 | 0.158 | 132.88 | 1.84×10<sup>-21</sup> | 1.52×10<sup>-20</sup> | 0.174 | 0.76 |
| Allergic disease |  |  |  |  |  |  |  |
| Atopic dermatitis | 0.062 | 0.324 | 35.76 | 1.85×10<sup>-4</sup> | 3.60×10<sup>-4</sup> | -0.617 | 8.74×10<sup>-5</sup> |
| Asthma | -0.001 | 0.993 | 172.65 | 3.36×10<sup>-31</sup> | 3.70×10<sup>-30</sup> | 0.519 | 6.70×10<sup>-3</sup> |
| Pediatric asthma | 0.087 | 0.539 | 28.07 | 3.16×10<sup>-3</sup> | 4.74×10<sup>-3</sup> | 0.438 | 0.208 |
| Pollinosis | 0.005 | 0.836 | 19.67 | 0.0501 | 0.0661 | 0.208 | 2.19×10<sup>-4</sup> |
| Chronic sinusitis | -0.061 | 0.548 | 120.51 | 1.43×10<sup>-20</sup> | 9.46×10<sup>-20</sup> | 0.936 | 1.99×10<sup>-4</sup> |
| Allergic conjunctivitis | 0.066 | 0.0637 | 11.74 | 0.383 | 0.67 | 0.134 | 0.117 |
| Urticaria | -0.013 | 0.551 | 11.21 | 0.426 | 0.67 | -0.120 | 0.0255 |
| Infection & Respiratory disease |  |  |  |  |  |  |  |
| Pulmonary tuberculosis | -0.074 | 0.168 | 58.95 | 1.45×10<sup>-8</sup> | 4.35×10<sup>-8</sup> | 0.850 | 6.56×10<sup>-10</sup> |
| Chronic hepatitis B | 0.157 | 0.37 | 175.54 | 8.55×10<sup>-32</sup> | 2.56×10<sup>-30</sup> | 1.769 | 5.28×10<sup>-5</sup> |
| Chronic hepatitis C | -0.004 | 0.956 | 75.80 | 9.52×10<sup>-12</sup> | 1.43×10<sup>-10</sup> | -0.376 | 0.0195 |
| Zoster infection | 0.063 | 0.154 | 11.33 | 0.416 | 0.67 | -0.004 | 0.974 |
| COVID-19 | 0.092 | 0.191 | 30.56 | 6.38×10<sup>-3</sup> | 0.0452 | -0.797 | 4.07×10<sup>-4</sup> |
| Pneumonia | -0.017 | 0.683 | 30.21 | 1.47×10<sup>-3</sup> | 2.42×10<sup>-3</sup> | 0.358 | 3.15×10<sup>-4</sup> |
| Interstitial lung disease | -0.056 | 0.538 | 22.04 | 0.0241 | 0.103 | 0.497 | 0.0253 |
| Cardiovascular & other disease |  |  |  |  |  |  |  |
| Coronary artery disease | -0.027 | 0.493 | 87.09 | 6.19×10<sup>-14</sup> | 3.41×10<sup>-13</sup> | -0.277 | 4.53×10<sup>-3</sup> |
| Myocardial infarction | -0.035 | 0.472 | 71.96 | 5.17×10<sup>-11</sup> | 2.13×10<sup>-10</sup> | -0.428 | 4.28×10<sup>-4</sup> |
| Ischemic stroke | 0.017 | 0.288 | 11.84 | 0.376 | 0.67 | -0.144 | 2.62×10<sup>-4</sup> |
| Subarachnoid hemorrhage | 0.015 | 0.789 | 7.34 | 0.771 | 0.907 | 0.334 | 5.43×10<sup>-3</sup> |
| Parkinson disease | 0.086 | 0.554 | 18.66 | 0.0675 | 0.0856 | -1.346 | 1.67×10<sup>-4</sup> |
| Ulcerative colitis | -0.005 | 0.987 | 68.11 | 2.79×10<sup>-10</sup> | 1.02×10<sup>-9</sup> | -1.740 | 0.0135 |
| Iron deficiency anemia | -0.001 | 0.967 | 3.63 | 0.979 | 0.987 | -0.066 | 0.132 |
| Quantitative traits |  |  |  |  |  |  |  |
| Height | 0.003 | 0.678 | 77.56 | 4.37×10<sup>-12</sup> | 1.25×10<sup>-11</sup> | -0.107 | 1.36×10<sup>-8</sup> |
| Body Weight | 0.011 | 0.204 | 62.48 | 3.20×10<sup>-9</sup> | 7.11×10<sup>-9</sup> | -0.146 | 1.63×10<sup>-10</sup> |
| BMI | 0.012 | 0.168 | 42.72 | 1.22×10<sup>-5</sup> | 2.03×10<sup>-5</sup> | -0.100 | 6.74×10<sup>-6</sup> |
| SBP: Systolic blood pressure | 0.004 | 0.492 | 16.01 | 0.141 | 0.156 | -0.027 | 0.0551 |
| DBP: Diastolic blood pressure | 0.004 | 0.462 | 16.16 | 0.135 | 0.156 | -0.014 | 0.365 |
| RBC count | 0.001 | 0.921 | 100.65 | 1.33×10<sup>-16</sup> | 8.86×10<sup>-16</sup> | -0.056 | 0.0939 |
| WBC count | -0.012 | 0.528 | 157.43 | 4.51×10<sup>-28</sup> | 4.51×10<sup>-27</sup> | -0.048 | 0.29 |
| Eosinophil count | -0.004 | 0.871 | 171.14 | 6.87×10<sup>-31</sup> | 1.37×10<sup>-29</sup> | -0.135 | 0.03 |
| Basophil count | -0.006 | 0.569 | 29.60 | 1.83×10<sup>-3</sup> | 2.61×10<sup>-3</sup> | -0.205 | 6.09×10<sup>-15</sup> |
| TC: Total cholesterol | -0.005 | 0.6 | 39.76 | 3.93×10<sup>-5</sup> | 6.05×10<sup>-5</sup> | -0.112 | 3.36×10<sup>-6</sup> |
| HDLC: HDL-cholesterol | -0.014 | 0.321 | 49.55 | 7.54×10<sup>-7</sup> | 1.51×10<sup>-6</sup> | 0.109 | 2.15×10<sup>-3</sup> |
| LDLC: LDL-cholesterol | -0.008 | 0.44 | 22.81 | 0.0188 | 0.025 | -0.124 | 4.21×10<sup>-7</sup> |
| TG: Triglycerides | 0.001 | 0.935 | 80.04 | 1.45×10<sup>-12</sup> | 4.84×10<sup>-12</sup> | -0.089 | 0.0205 |
| HbA1c | -0.011 | 0.13 | 18.33 | 0.0743 | 0.0928 | 0.031 | 0.11 |

|  | MR-cML |  |
| --- | --- | --- |
|  | Theta | P-value |
| Autoimmune disease & T2D |  |  |
| Type 1 diabetes | 1.456 | $1.87 \times 10^{-20}$ |
| Type 2 diabetes | 0.085 | 0.0161 |
| Hashimoto's disease | 2.059 | $3.92 \times 10^{-13}$ |
| Grave's disease | 2.362 | $1.29 \times 10^{-30}$ |
| Rheumatoid arthritis | 0.640 | $1.10 \times 10^{-3}$ |
| SLE | -0.011 | 0.971 |
| Psoriasis | -0.705 | 0.318 |
| Allergic disease |  |  |
| Atopic dermatitis | -0.482 | $1.90 \times 10^{-6}$ |
| Asthma | 0.649 | $4.66 \times 10^{-22}$ |
| Pediatric asthma | 0.215 | 0.362 |
| Pollinosis | 0.214 | $8.95 \times 10^{-7}$ |
| Chronic sinusitis | 1.360 | $7.67 \times 10^{-31}$ |
| Allergic conjunctivitis | 0.129 | 0.129 |
| Urticaria | -0.123 | 0.0231 |
| Infection & Respiratory disease |  |  |
| Pulmonary tuberculosis | 0.946 | $2.64 \times 10^{-31}$ |
| Chronic hepatitis B | 1.172 | $9.69 \times 10^{-10}$ |
| Chronic hepatitis C | -0.468 | $6.42 \times 10^{-10}$ |
| Zoster infection | -0.002 | 0.984 |
| COVID-19 | -0.834 | $1.09 \times 10^{-7}$ |
| Pneumonia | 0.382 | $7.46 \times 10^{-9}$ |
| Interstitial lung disease | 0.529 | $1.39 \times 10^{-3}$ |
| Cardiovascular & other disease |  |  |
| Coronary artery disease | -0.252 | $1.16 \times 10^{-5}$ |
| Myocardial infarction | -0.601 | $3.76 \times 10^{-20}$ |
| Ischemic stroke | -0.149 | $1.70 \times 10^{-4}$ |
| Subarachnoid hemorrhage | 0.337 | 0.0229 |
| Parkinson disease | -1.381 | $1.10 \times 10^{-6}$ |
| Ulcerative colitis | -2.636 | $1.04 \times 10^{-10}$ |
| Iron deficiency anemia | -0.066 | 0.387 |
| Quantitative traits |  |  |
| Height | -0.114 | $1.55 \times 10^{-27}$ |
| Body Weight | -0.163 | $2.86 \times 10^{-39}$ |
| BMI | -0.112 | $4.26 \times 10^{-16}$ |
| SBP: Systolic blood pressure | -0.028 | 0.0199 |
| DBP: Diastolic blood pressure | -0.014 | 0.263 |
| RBC count | 0.008 | 0.595 |
| WBC count | 0.051 | $3.65 \times 10^{-3}$ |
| Eosinophil count | -0.145 | $3.33 \times 10^{-12}$ |
| Basophil count | -0.213 | $3.87 \times 10^{-31}$ |
| TC: Total cholesterol | -0.122 | $1.81 \times 10^{-16}$ |
| HDLC: HDL-cholesterol | 0.138 | $5.04 \times 10^{-12}$ |
| LDLC: LDL-cholesterol | -0.128 | $1.08 \times 10^{-12}$ |
| TG: Triglycerides | -0.112 | $1.20 \times 10^{-9}$ |
| HbA1c | 0.032 | 0.0369 |

Font colors: Red indicates a positive association, Blue indicates an inverse association.
Background colors: Red indicates robust positive association (full IV set analysis), Blue indicates robust inverse association (full IV set analysis).

**Supplementary Table 19.** MR excluding HLA class II variants.

|  | IVW (fixed effects) |  | Weighted median |  | Weighted mode |  | MR-Egger |  |
| --- | --- | --- | --- | --- | --- | --- | --- | --- |
|  | Beta | P-value | Beta | P-value | Beta | P-value | Beta | P-value |
| **Autoimmune disease & T2D** |  |  |  |  |  |  |  |  |
| Type 1 diabetes | 0.518 | 0.023 | 0.642 | 0.0558 | 0.798 | 0.277 | 0.625 | 0.543 |
| Type 2 diabetes | 0.054 | 0.282 | -0.068 | 0.372 | -0.110 | 0.254 | -0.183 | 0.653 |
| Hashimoto's disease | -0.266 | 0.429 | -0.850 | 0.105 | -0.871 | 0.297 | -0.206 | 0.927 |
| Grave's disease | -0.076 | 0.616 | -0.292 | 0.205 | -0.307 | 0.251 | -1.255 | 0.431 |
| Rheumatoid arthritis | 0.051 | 0.644 | 0.330 | 0.0443 | 0.312 | 0.0892 | 0.085 | 0.932 |
| SLE | 0.447 | 0.306 | 0.520 | 0.4 | 0.540 | 0.517 | -0.793 | 0.761 |
| Psoriasis | -0.948 | 3.48×10<sup>-4</sup> | -1.517 | 4.44×10<sup>-4</sup> | -1.664 | 0.0359 | 0.118 | 0.96 |
| **Allergic disease** |  |  |  |  |  |  |  |  |
| Atopic dermatitis | -0.164 | 0.225 | 0.031 | 0.885 | 0.136 | 0.755 | -0.460 | 0.561 |
| Asthma | 0.453 | 1.25×10<sup>-9</sup> | 0.067 | 0.557 | 0.010 | 0.945 | 1.421 | 0.194 |
| Pediatric asthma | 0.711 | 0.035 | 0.713 | 0.18 | 0.991 | 0.405 | 1.349 | 0.572 |
| Pollinosis | 0.038 | 0.557 | 0.105 | 0.292 | 0.184 | 0.214 | 0.683 | 0.0751 |
| Chronic sinusitis | 0.459 | 9.42×10<sup>-5</sup> | 0.199 | 0.257 | 0.176 | 0.397 | 2.150 | 0.0213 |
| Allergic conjunctivitis | 0.255 | 0.0453 | 0.141 | 0.408 | 0.127 | 0.595 | 0.360 | 0.492 |
| Urticaria | -0.026 | 0.747 | 0.008 | 0.943 | -0.045 | 0.79 | 0.370 | 0.269 |
| **Infection & Respiratory disease** |  |  |  |  |  |  |  |  |
| Pulmonary tuberculosis | 0.252 | 6.14×10<sup>-3</sup> | 0.109 | 0.433 | 0.035 | 0.85 | 0.429 | 0.39 |
| Chronic hepatitis B | 0.836 | 7.05×10<sup>-7</sup> | 0.046 | 0.866 | -0.016 | 0.958 | 1.348 | 0.48 |
| Chronic hepatitis C | -0.189 | 0.0469 | -0.284 | 0.0788 | -0.425 | 0.204 | -0.957 | 0.214 |
| Zoster infection | -0.139 | 0.405 | -0.078 | 0.712 | -0.092 | 0.773 | -0.200 | 0.73 |
| COVID-19 | 0.048 | 0.808 | -0.266 | 0.348 | -0.323 | 0.426 | -1.249 | 0.27 |
| Pneumonia | 0.080 | 0.388 | 0.129 | 0.302 | 0.21 | 0.261 | 0.914 | 0.0327 |
| Interstitial lung disease | -0.110 | 0.649 | -0.165 | 0.636 | -0.160 | 0.767 | -0.596 | 0.638 |
| **Cardiovascular & other disease** |  |  |  |  |  |  |  |  |
| Coronary artery disease | -0.033 | 0.534 | 0.113 | 0.178 | 0.182 | 0.134 | -0.283 | 0.584 |
| Myocardial infarction | -0.090 | 0.223 | 0.196 | 0.0878 | 0.251 | 0.0486 | -0.338 | 0.63 |
| Ischemic stroke | -0.084 | 0.151 | -0.107 | 0.151 | -0.129 | 0.226 | -0.067 | 0.743 |
| Subarachnoid hemorrhage | 0.220 | 0.333 | 0.368 | 0.214 | 0.401 | 0.377 | 0.796 | 0.33 |
| Parkinson disease | -0.302 | 0.476 | 0.103 | 0.852 | 0.160 | 0.807 | 0.716 | 0.66 |
| Ulcerative colitis | -0.232 | 0.594 | 0.078 | 0.899 | 0.111 | 0.898 | 1.387 | 0.473 |
| Iron deficiency anemia | -0.112 | 0.339 | -0.169 | 0.305 | -0.168 | 0.555 | 0.287 | 0.5 |
| **Quantitative traits** |  |  |  |  |  |  |  |  |
| Height | 0.006 | 0.583 | 0.023 | 0.242 | 0.004 | 0.891 | -0.175 | 0.0471 |
| Body Weight | -0.015 | 0.315 | -0.031 | 0.176 | -0.032 | 0.2 | -0.282 | 0.024 |
| BMI | -0.024 | 0.177 | -0.044 | 0.094 | -0.040 | 0.283 | -0.222 | 0.0807 |
| SBP: Systolic blood pressure | -0.014 | 0.448 | -0.019 | 0.47 | -0.020 | 0.629 | -0.112 | 0.136 |
| DBP: Diastolic blood pressure | 0.006 | 0.769 | -0.001 | 0.967 | -0.013 | 0.797 | -0.125 | 0.127 |
| RBC count | -0.066 | 1.09×10<sup>-4</sup> | -0.049 | 0.0561 | -0.037 | 0.309 | 0.033 | 0.775 |
| WBC count | 0.031 | 0.099 | 0.072 | 0.0104 | 0.082 | 0.0638 | 0.211 | 0.114 |
| Eosinophil count | 0.070 | 3.80×10<sup>-3</sup> | 0.090 | 0.0381 | 0.076 | 0.459 | 0.087 | 0.692 |
| Basophil count | -0.064 | 9.46×10<sup>-3</sup> | -0.050 | 0.182 | -0.052 | 0.339 | -0.019 | 0.866 |
| TC: Total cholesterol | -0.044 | 0.0242 | -0.024 | 0.41 | -0.013 | 0.759 | -0.039 | 0.746 |
| HDLC: HDL-cholesterol | 0.034 | 0.183 | -0.007 | 0.84 | -0.010 | 0.832 | 0.154 | 0.233 |
| LDLC: LDL-cholesterol | -0.067 | 0.0102 | -0.031 | 0.436 | -0.008 | 0.907 | -0.070 | 0.622 |
| TG: Triglycerides | -0.057 | 9.05×10<sup>-3</sup> | -0.056 | 0.0744 | -0.074 | 0.196 | -0.174 | 0.225 |
| HbA1c | -0.009 | 0.686 | -0.010 | 0.743 | -0.035 | 0.522 | 0.040 | 0.618 |

|  | pleiotropy test |  | heterogeneity test |  |  | IVW<br>(multiplicative random effects) |  |
| --- | --- | --- | --- | --- | --- | --- | --- |
|  | intercept | P-value | Cochran's Q | P-value | FDR Q-value | Beta | P-value |
| **Autoimmune disease & T2D** |  |  |  |  |  |  |  |
| Type 1 diabetes | -0.008 | 0.912 | 11.48 | 0.176 | 0.224 | 0.518 | 0.0577 |
| Type 2 diabetes | 0.019 | 0.544 | 39.62 | $3.77 \times 10^{-6}$ | $1.38 \times 10^{-5}$ | 0.054 | 0.629 |
| Hashimoto's disease | -0.005 | 0.978 | 25.83 | $1.12 \times 10^{-3}$ | $2.85 \times 10^{-3}$ | -0.266 | 0.66 |
| Grave's disease | 0.092 | 0.438 | 66.62 | $2.31 \times 10^{-11}$ | $3.80 \times 10^{-10}$ | -0.076 | 0.862 |
| Rheumatoid arthritis | -0.003 | 0.971 | 46.69 | $1.75 \times 10^{-7}$ | $9.65 \times 10^{-7}$ | 0.051 | 0.848 |
| SLE | 0.097 | 0.62 | 21.33 | $6.33 \times 10^{-3}$ | 0.013 | 0.447 | 0.531 |
| Psoriasis | -0.078 | 0.642 | 60.35 | $7.97 \times 10^{-9}$ | $6.57 \times 10^{-8}$ | -0.948 | 0.127 |
| **Allergic disease** |  |  |  |  |  |  |  |
| Atopic dermatitis | 0.023 | 0.694 | 19.79 | 0.0112 | 0.0217 | -0.164 | 0.44 |
| Asthma | -0.076 | 0.34 | 125.69 | $2.21 \times 10^{-23}$ | $7.31 \times 10^{-22}$ | 0.453 | 0.125 |
| Pediatric asthma | -0.050 | 0.778 | 28.52 | $3.85 \times 10^{-4}$ | $1.27 \times 10^{-3}$ | 0.711 | 0.264 |
| Pollinosis | -0.051 | 0.0776 | 25.41 | $1.33 \times 10^{-3}$ | $3.13 \times 10^{-3}$ | 0.038 | 0.741 |
| Chronic sinusitis | -0.132 | 0.0452 | 44.01 | $5.67 \times 10^{-7}$ | $2.67 \times 10^{-6}$ | 0.459 | 0.0959 |
| Allergic conjunctivitis | -0.008 | 0.831 | 9.57 | 0.297 | 0.469 | 0.255 | 0.0671 |
| Urticaria | -0.031 | 0.219 | 11.17 | 0.192 | 0.36 | -0.026 | 0.785 |
| **Infection & Respiratory disease** |  |  |  |  |  |  |  |
| Pulmonary tuberculosis | -0.014 | 0.703 | 16.72 | 0.0332 | 0.0608 | 0.252 | 0.058 |
| Chronic hepatitis B | -0.040 | 0.775 | 73.11 | $1.18 \times 10^{-12}$ | $3.54 \times 10^{-11}$ | 0.836 | 0.101 |
| Chronic hepatitis C | 0.060 | 0.288 | 40.26 | $2.87 \times 10^{-6}$ | $4.31 \times 10^{-5}$ | -0.189 | 0.376 |
| Zoster infection | 0.005 | 0.912 | 4.30 | 0.829 | 0.888 | -0.139 | 0.256 |
| COVID-19 | 0.095 | 0.235 | 26.74 | $5.03 \times 10^{-3}$ | 0.0377 | 0.048 | 0.876 |
| Pneumonia | -0.065 | 0.0387 | 16.55 | 0.0351 | 0.061 | 0.080 | 0.548 |
| Interstitial lung disease | 0.038 | 0.687 | 16.01 | 0.0422 | 0.211 | -0.110 | 0.747 |
| **Cardiovascular & other disease** |  |  |  |  |  |  |  |
| Coronary artery disease | 0.020 | 0.612 | 55.02 | $4.38 \times 10^{-9}$ | $4.82 \times 10^{-8}$ | -0.033 | 0.813 |
| Myocardial infarction | 0.019 | 0.71 | 53.15 | $1.01 \times 10^{-8}$ | $6.66 \times 10^{-8}$ | -0.090 | 0.637 |
| Ischemic stroke | -0.001 | 0.93 | 1.43 | 0.994 | 0.994 | -0.084 | $6.80 \times 10^{-4}$ |
| Subarachnoid hemorrhage | -0.045 | 0.453 | 5.23 | 0.732 | 0.845 | 0.220 | 0.231 |
| Parkinson disease | -0.080 | 0.515 | 8.98 | 0.344 | 0.379 | -0.302 | 0.501 |
| Ulcerative colitis | -0.127 | 0.385 | 12.38 | 0.135 | 0.194 | -0.232 | 0.669 |
| Iron deficiency anemia | -0.031 | 0.334 | 8.50 | 0.387 | 0.527 | -0.112 | 0.354 |
| **Quantitative traits** |  |  |  |  |  |  |  |
| Height | 0.014 | 0.035 | 53.47 | $8.75 \times 10^{-9}$ | $8.75 \times 10^{-8}$ | 0.006 | 0.832 |
| Body Weight | 0.021 | 0.0248 | 59.14 | $6.88 \times 10^{-10}$ | $1.38 \times 10^{-8}$ | -0.015 | 0.712 |
| BMI | 0.016 | 0.0977 | 36.84 | $1.23 \times 10^{-5}$ | $6.16 \times 10^{-5}$ | -0.024 | 0.53 |
| SBP: Systolic blood pressure | 0.008 | 0.166 | 11.09 | 0.196 | 0.231 | -0.014 | 0.52 |
| DBP: Diastolic blood pressure | 0.010 | 0.1 | 13.52 | 0.0952 | 0.127 | 0.006 | 0.821 |
| RBC count | -0.008 | 0.381 | 29.70 | $2.39 \times 10^{-4}$ | $7.97 \times 10^{-4}$ | -0.066 | 0.0445 |
| WBC count | -0.014 | 0.15 | 33.63 | $4.73 \times 10^{-5}$ | $1.89 \times 10^{-4}$ | 0.031 | 0.421 |
| Eosinophil count | -0.001 | 0.937 | 46.56 | $1.86 \times 10^{-7}$ | $1.24 \times 10^{-6}$ | 0.070 | 0.23 |
| Basophil count | -0.004 | 0.676 | 12.31 | 0.138 | 0.172 | -0.064 | 0.0365 |
| TC: Total cholesterol | 0.000 | 0.96 | 21.27 | $6.46 \times 10^{-3}$ | 0.0129 | -0.044 | 0.167 |
| HDLC: HDL-cholesterol | -0.009 | 0.323 | 15.09 | 0.0574 | 0.0882 | 0.034 | 0.333 |
| LDLC: LDL-cholesterol | 0.000 | 0.982 | 16.94 | 0.0307 | 0.0512 | -0.067 | 0.0776 |
| TG: Triglycerides | 0.009 | 0.381 | 24.78 | $1.69 \times 10^{-3}$ | $4.23 \times 10^{-3}$ | -0.057 | 0.138 |
| HbA1c | -0.004 | 0.522 | 7.19 | 0.517 | 0.574 | -0.009 | 0.669 |

|  | MR-cML |  |
| --- | --- | --- |
|  | Theta | P-value |
| Autoimmune disease & T2D |  |  |
| Type 1 diabetes | 0.546 | 0.0225 |
| Type 2 diabetes | -0.063 | 0.317 |
| Hashimoto's disease | -0.790 | 0.0638 |
| Grave's disease | -0.310 | 0.103 |
| Rheumatoid arthritis | 0.402 | 2.24×10<sup>-3</sup> |
| SLE | 0.752 | 0.134 |
| Psoriasis | -1.785 | 1.85×10<sup>-6</sup> |
| Allergic disease |  |  |
| Atopic dermatitis | -0.176 | 0.212 |
| Asthma | 0.093 | 0.323 |
| Pediatric asthma | 0.225 | 0.606 |
| Pollinosis | 0.066 | 0.381 |
| Chronic sinusitis | 0.043 | 0.757 |
| Allergic conjunctivitis | 0.254 | 0.0561 |
| Urticaria | -0.024 | 0.779 |
| Infection & Respiratory disease |  |  |
| Pulmonary tuberculosis | 0.243 | 0.018 |
| Chronic hepatitis B | -0.010 | 0.961 |
| Chronic hepatitis C | -0.059 | 0.576 |
| Zoster infection | -0.142 | 0.401 |
| COVID-19 | -0.063 | 0.785 |
| Pneumonia | 0.098 | 0.327 |
| Interstitial lung disease | -0.124 | 0.623 |
| Cardiovascular & other disease |  |  |
| Coronary artery disease | 0.151 | 0.0525 |
| Myocardial infarction | 0.227 | 0.0151 |
| Ischemic stroke | -0.084 | 0.152 |
| Subarachnoid hemorrhage | 0.221 | 0.334 |
| Parkinson disease | -0.282 | 0.521 |
| Ulcerative colitis | -0.207 | 0.65 |
| Iron deficiency anemia | -0.124 | 0.317 |
| Quantitative traits |  |  |
| Height | 0.002 | 0.893 |
| Body Weight | -0.050 | 0.0179 |
| BMI | -0.048 | 0.0167 |
| SBP: Systolic blood pressure | -0.015 | 0.435 |
| DBP: Diastolic blood pressure | 0.007 | 0.737 |
| RBC count | -0.054 | 0.0209 |
| WBC count | 0.076 | 4.09×10<sup>-3</sup> |
| Eosinophil count | 0.100 | 7.79×10<sup>-3</sup> |
| Basophil count | -0.066 | 8.94×10<sup>-3</sup> |
| TC: Total cholesterol | -0.037 | 0.132 |
| HDLC: HDL-cholesterol | 0.035 | 0.198 |
| LDLC: LDL-cholesterol | -0.069 | 0.0127 |
| TG: Triglycerides | -0.057 | 0.0244 |
| HbA1c | -0.009 | 0.688 |

Font colors: Red indicates a positive association, Blue indicates an inverse association.
Background colors: Red indicates robust positive association (full IV set analysis), Blue indicates robust inverse association (full IV set analysis).

**Supplementary Table 20.** MR restricted to HLA class II variants.

|  | IVW (fixed effects) |  | Weighted median |  | Weighted mode |  | MR-Egger |  |
| --- | --- | --- | --- | --- | --- | --- | --- | --- |
|  | Beta | P-value | Beta | P-value | Beta | P-value | Beta | P-value |
| **Autoimmune disease & T2D** |  |  |  |  |  |  |  |  |
| Type 1 diabetes | 1.541 | 5.52×10<sup>-20</sup> | 1.562 | 1.05×10<sup>-11</sup> | 1.640 | 1.98×10<sup>-3</sup> | 1.256 | 0.238 |
| Type 2 diabetes | 0.120 | 1.35×10<sup>-3</sup> | 0.166 | 6.60×10<sup>-4</sup> | 0.175 | 0.0242 | 0.230 | 0.399 |
| Hashimoto's disease | 2.066 | 1.86×10<sup>-16</sup> | 2.148 | 2.61×10<sup>-10</sup> | 2.132 | 2.52×10<sup>-3</sup> | 4.103 | 0.112 |
| Grave's disease | 2.447 | 1.40×10<sup>-102</sup> | 2.244 | 2.04×10<sup>-24</sup> | 2.369 | 1.06×10<sup>-3</sup> | -1.308 | 0.422 |
| Rheumatoid arthritis | 2.098 | 7.20×10<sup>-142</sup> | 1.226 | 2.04×10<sup>-9</sup> | 0.567 | 0.0959 | 1.312 | 0.681 |
| SLE | -0.048 | 0.883 | -0.207 | 0.629 | -0.377 | 0.534 | 0.392 | 0.839 |
| Psoriasis | 0.935 | 2.93×10<sup>-5</sup> | 0.701 | 0.0349 | 0.394 | 0.273 | 1.269 | 0.725 |
| **Allergic disease** |  |  |  |  |  |  |  |  |
| Atopic dermatitis | -0.807 | 6.04×10<sup>-16</sup> | -0.565 | 8.54×10<sup>-5</sup> | -0.564 | 0.0104 | -0.918 | 0.304 |
| Asthma | 0.403 | 2.81×10<sup>-13</sup> | 0.444 | 2.92×10<sup>-4</sup> | 0.745 | 7.92×10<sup>-4</sup> | 0.576 | 0.585 |
| Pediatric asthma | 0.032 | 0.897 | 0.234 | 0.489 | 0.357 | 0.459 | -0.824 | 0.52 |
| Pollinosis | 0.207 | 1.61×10<sup>-5</sup> | 0.195 | 2.74×10<sup>-3</sup> | 0.191 | 0.0554 | -0.085 | 0.787 |
| Chronic sinusitis | 0.988 | 6.01×10<sup>-30</sup> | 0.909 | 5.17×10<sup>-6</sup> | 1.237 | 1.20×10<sup>-3</sup> | -0.059 | 0.972 |
| Allergic conjunctivitis | 0.053 | 0.572 | 0.044 | 0.72 | 0.012 | 0.945 | -0.290 | 0.538 |
| Urticaria | -0.151 | 0.0129 | -0.184 | 0.0181 | -0.198 | 0.0837 | 0.283 | 0.389 |
| **Infection & Respiratory disease** |  |  |  |  |  |  |  |  |
| Pulmonary tuberculosis | 0.994 | 2.43×10<sup>-48</sup> | 1.007 | 2.06×10<sup>-21</sup> | 0.964 | 1.07×10<sup>-4</sup> | 1.434 | 0.11 |
| Chronic hepatitis B | 1.871 | 2.11×10<sup>-50</sup> | 1.118 | 1.29×10<sup>-9</sup> | 1.070 | 4.52×10<sup>-4</sup> | -1.513 | 0.551 |
| Chronic hepatitis C | -0.355 | 4.07×10<sup>-7</sup> | -0.350 | 8.43×10<sup>-4</sup> | -0.369 | 0.0307 | 0.430 | 0.684 |
| Zoster infection | 0.026 | 0.832 | 0.144 | 0.37 | 0.154 | 0.522 | -1.037 | 0.131 |
| COVID-19 | -1.179 | 8.99×10<sup>-10</sup> | -1.096 | 1.96×10<sup>-5</sup> | -1.109 | 6.46×10<sup>-3</sup> | -1.679 | 0.151 |
| Pneumonia | 0.389 | 1.49×10<sup>-8</sup> | 0.438 | 7.20×10<sup>-6</sup> | 0.519 | 4.23×10<sup>-3</sup> | 0.358 | 0.636 |
| Interstitial lung disease | 0.733 | 4.47×10<sup>-5</sup> | 0.607 | 7.90×10<sup>-3</sup> | 0.606 | 0.0778 | 0.884 | 0.339 |
| **Cardiovascular & other disease** |  |  |  |  |  |  |  |  |
| Coronary artery disease | -0.302 | 3.00×10<sup>-14</sup> | -0.306 | 1.23×10<sup>-7</sup> | -0.313 | 4.32×10<sup>-3</sup> | 0.248 | 0.632 |
| Myocardial infarction | -0.462 | 2.06×10<sup>-17</sup> | -0.471 | 1.92×10<sup>-8</sup> | -0.483 | 2.49×10<sup>-3</sup> | 0.335 | 0.535 |
| Ischemic stroke | -0.157 | 3.08×10<sup>-4</sup> | -0.181 | 8.24×10<sup>-4</sup> | -0.175 | 0.0329 | -0.476 | 0.125 |
| Subarachnoid hemorrhage | 0.345 | 0.0406 | 0.460 | 0.0278 | 0.496 | 0.112 | -0.007 | 0.993 |
| Parkinson disease | -1.589 | 4.24×10<sup>-7</sup> | -1.689 | 5.12×10<sup>-5</sup> | -1.705 | 0.014 | -4.143 | 0.0546 |
| Ulcerative colitis | -2.191 | 1.38×10<sup>-11</sup> | -2.899 | 3.19×10<sup>-9</sup> | -3.323 | 4.97×10<sup>-4</sup> | -1.744 | 0.728 |
| Iron deficiency anemia | -0.020 | 0.821 | -0.068 | 0.521 | -0.078 | 0.601 | -0.187 | 0.663 |
| **Quantitative traits** |  |  |  |  |  |  |  |  |
| Height | -0.128 | 9.09×10<sup>-56</sup> | -0.100 | 2.00×10<sup>-10</sup> | -0.092 | 1.16×10<sup>-3</sup> | 0.033 | 0.631 |
| Body Weight | -0.171 | 1.67×10<sup>-54</sup> | -0.172 | 2.91×10<sup>-20</sup> | -0.184 | 3.11×10<sup>-4</sup> | -0.142 | 0.259 |
| BMI | -0.115 | 7.82×10<sup>-19</sup> | -0.135 | 9.43×10<sup>-14</sup> | -0.142 | 3.11×10<sup>-4</sup> | -0.204 | 0.149 |
| SBP: Systolic blood pressure | -0.025 | 0.0705 | -0.024 | 0.214 | -0.021 | 0.433 | -0.049 | 0.603 |
| DBP: Diastolic blood pressure | -0.013 | 0.341 | 0.004 | 0.826 | 0.006 | 0.768 | 0.016 | 0.833 |
| RBC count | -0.053 | 2.39×10<sup>-5</sup> | -0.008 | 0.732 | 0.007 | 0.75 | -0.185 | 0.456 |
| WBC count | -0.066 | 1.66×10<sup>-6</sup> | 0.027 | 0.267 | 0.043 | 0.0605 | -0.011 | 0.975 |
| Eosinophil count | -0.214 | 9.40×10<sup>-33</sup> | -0.171 | 1.08×10<sup>-9</sup> | -0.156 | 1.21×10<sup>-3</sup> | 0.274 | 0.127 |
| Basophil count | -0.241 | 1.30×10<sup>-39</sup> | -0.245 | 5.50×10<sup>-23</sup> | -0.280 | 3.42×10<sup>-4</sup> | -0.063 | 0.491 |
| TC: Total cholesterol | -0.128 | 1.23×10<sup>-18</sup> | -0.129 | 1.10×10<sup>-9</sup> | -0.129 | 4.79×10<sup>-3</sup> | 0.004 | 0.969 |
| HDLC: HDL-cholesterol | 0.118 | 9.36×10<sup>-10</sup> | 0.177 | 1.34×10<sup>-9</sup> | 0.188 | 1.32×10<sup>-3</sup> | 0.310 | 0.248 |
| LDLC: LDL-cholesterol | -0.135 | 4.98×10<sup>-12</sup> | -0.134 | 9.89×10<sup>-7</sup> | -0.134 | 0.0118 | 0.040 | 0.676 |
| TG: Triglycerides | -0.097 | 2.52×10<sup>-9</sup> | -0.136 | 2.80×10<sup>-8</sup> | -0.137 | 1.85×10<sup>-3</sup> | -0.007 | 0.979 |
| HbA1c | 0.053 | 2.05×10<sup>-3</sup> | 0.056 | 0.0145 | 0.052 | 0.146 | 0.089 | 0.382 |

|  | pleiotropy test |  | heterogeneity test |  |  | IVW<br>(multiplicative random effects) |  |
| --- | --- | --- | --- | --- | --- | --- | --- |
|  | intercept | P-value | Cochran's Q | P-value | FDR Q-value | Beta | P-value |
| **Autoimmune disease & T2D** |  |  |  |  |  |  |  |
| Type 1 diabetes | 0.031 | 0.767 | 7.29 | 0.295 | 0.347 | 1.541 | 1.01×10<sup>-16</sup> |
| Type 2 diabetes | -0.012 | 0.67 | 10.76 | 0.0962 | 0.138 | 0.120 | 0.0167 |
| Hashimoto's disease | -0.219 | 0.373 | 19.85 | 2.95×10<sup>-3</sup> | 5.73×10<sup>-3</sup> | 2.066 | 6.02×10<sup>-6</sup> |
| Grave's disease | 0.404 | 0.05 | 91.10 | 1.79×10<sup>-17</sup> | 2.96×10<sup>-16</sup> | 2.447 | 3.42×10<sup>-8</sup> |
| Rheumatoid arthritis | 0.085 | 0.8 | 309.89 | 6.20×10<sup>-64</sup> | 2.05×10<sup>-62</sup> | 2.098 | 4.18×10<sup>-4</sup> |
| SLE | -0.047 | 0.816 | 7.48 | 0.279 | 0.341 | -0.048 | 0.895 |
| Psoriasis | -0.036 | 0.924 | 52.53 | 1.46×10<sup>-9</sup> | 9.65×10<sup>-9</sup> | 0.935 | 0.158 |
| **Allergic disease** |  |  |  |  |  |  |  |
| Atopic dermatitis | 0.012 | 0.893 | 14.97 | 0.0205 | 0.0338 | -0.807 | 3.05×10<sup>-7</sup> |
| Asthma | -0.019 | 0.865 | 73.86 | 6.57×10<sup>-14</sup> | 5.42×10<sup>-13</sup> | 0.403 | 0.0374 |
| Pediatric asthma | 0.092 | 0.495 | 5.86 | 0.439 | 0.483 | 0.032 | 0.896 |
| Pollinosis | 0.032 | 0.363 | 10.75 | 0.0964 | 0.138 | 0.207 | 1.27×10<sup>-3</sup> |
| Chronic sinusitis | 0.113 | 0.535 | 86.61 | 1.53×10<sup>-16</sup> | 1.69×10<sup>-15</sup> | 0.988 | 2.77×10<sup>-3</sup> |
| Allergic conjunctivitis | 0.037 | 0.46 | 2.49 | 0.869 | 0.981 | 0.053 | 0.381 |
| Urticaria | -0.047 | 0.2 | 8.17 | 0.226 | 0.613 | -0.151 | 0.0331 |
| **Infection & Respiratory disease** |  |  |  |  |  |  |  |
| Pulmonary tuberculosis | -0.047 | 0.569 | 29.35 | 5.23×10<sup>-5</sup> | 1.57×10<sup>-4</sup> | 0.994 | 3.95×10<sup>-11</sup> |
| Chronic hepatitis B | 0.364 | 0.203 | 116.18 | 1.03×10<sup>-22</sup> | 3.10×10<sup>-21</sup> | 1.871 | 6.92×10<sup>-4</sup> |
| Chronic hepatitis C | -0.085 | 0.456 | 52.83 | 1.27×10<sup>-9</sup> | 1.91×10<sup>-8</sup> | -0.355 | 0.0878 |
| Zoster infection | 0.115 | 0.117 | 7.90 | 0.245 | 0.613 | 0.026 | 0.853 |
| COVID-19 | 0.053 | 0.627 | 6.51 | 0.369 | 0.656 | -1.179 | 4.03×10<sup>-9</sup> |
| Pneumonia | 0.003 | 0.967 | 24.89 | 3.58×10<sup>-4</sup> | 9.84×10<sup>-4</sup> | 0.389 | 5.44×10<sup>-3</sup> |
| Interstitial lung disease | -0.016 | 0.861 | 2.88 | 0.823 | 0.981 | 0.733 | 3.90×10<sup>-9</sup> |
| **Cardiovascular & other disease** |  |  |  |  |  |  |  |
| Coronary artery disease | -0.059 | 0.301 | 44.28 | 6.50×10<sup>-8</sup> | 3.06×10<sup>-7</sup> | -0.302 | 5.16×10<sup>-3</sup> |
| Myocardial infarction | -0.086 | 0.166 | 30.26 | 3.50×10<sup>-5</sup> | 1.16×10<sup>-4</sup> | -0.462 | 1.57×10<sup>-4</sup> |
| Ischemic stroke | 0.034 | 0.262 | 10.84 | 0.0934 | 0.4 | -0.157 | 7.26×10<sup>-3</sup> |
| Subarachnoid hemorrhage | 0.038 | 0.665 | 4.37 | 0.626 | 0.918 | 0.345 | 0.0165 |
| Parkinson disease | 0.275 | 0.176 | 9.63 | 0.141 | 0.194 | -1.589 | 6.55×10<sup>-5</sup> |
| Ulcerative colitis | -0.048 | 0.927 | 49.72 | 5.35×10<sup>-9</sup> | 2.94×10<sup>-8</sup> | -2.191 | 0.0189 |
| Iron deficiency anemia | 0.018 | 0.689 | 1.10 | 0.981 | 0.981 | -0.020 | 0.597 |
| **Quantitative traits** |  |  |  |  |  |  |  |
| Height | -0.017 | 0.052 | 33.79 | 7.40×10<sup>-6</sup> | 1.64×10<sup>-5</sup> | -0.128 | 3.36×10<sup>-11</sup> |
| Body Weight | -0.003 | 0.8 | 24.08 | 5.04×10<sup>-4</sup> | 9.16×10<sup>-4</sup> | -0.171 | 8.51×10<sup>-15</sup> |
| BMI | 0.010 | 0.479 | 22.06 | 1.18×10<sup>-3</sup> | 1.97×10<sup>-3</sup> | -0.115 | 3.80×10<sup>-6</sup> |
| SBP: Systolic blood pressure | 0.003 | 0.789 | 9.84 | 0.132 | 0.176 | -0.025 | 0.158 |
| DBP: Diastolic blood pressure | -0.003 | 0.689 | 5.87 | 0.438 | 0.461 | -0.013 | 0.336 |
| RBC count | 0.014 | 0.582 | 80.55 | 2.75×10<sup>-15</sup> | 2.75×10<sup>-14</sup> | -0.053 | 0.249 |
| WBC count | -0.006 | 0.863 | 120.21 | 1.47×10<sup>-23</sup> | 2.94×10<sup>-22</sup> | -0.066 | 0.285 |
| Eosinophil count | -0.053 | 0.0206 | 51.96 | 1.90×10<sup>-9</sup> | 5.42×10<sup>-9</sup> | -0.214 | 5.12×10<sup>-5</sup> |
| Basophil count | -0.019 | 0.0848 | 5.58 | 0.472 | 0.472 | -0.241 | 1.78×10<sup>-42</sup> |
| TC: Total cholesterol | -0.014 | 0.239 | 15.26 | 0.0183 | 0.0282 | -0.128 | 3.29×10<sup>-8</sup> |
| HDLC: HDL-cholesterol | -0.021 | 0.443 | 40.12 | 4.31×10<sup>-7</sup> | 1.08×10<sup>-6</sup> | 0.118 | 0.018 |
| LDLC: LDL-cholesterol | -0.019 | 0.105 | 7.39 | 0.287 | 0.337 | -0.135 | 4.84×10<sup>-10</sup> |
| TG: Triglycerides | -0.010 | 0.741 | 61.71 | 2.02×10<sup>-11</sup> | 8.09×10<sup>-11</sup> | -0.097 | 0.0631 |
| HbA1c | -0.004 | 0.705 | 7.09 | 0.313 | 0.348 | 0.053 | 4.55×10<sup>-3</sup> |

|  | MR-cML |  |
| --- | --- | --- |
|  | Theta | P-value |
| Autoimmune disease & T2D |  |  |
| Type 1 diabetes | 1.560 | $8.02 \times 10^{-18}$ |
| Type 2 diabetes | 0.127 | $1.52 \times 10^{-3}$ |
| Hashimoto's disease | 2.280 | $5.84 \times 10^{-14}$ |
| Grave's disease | 2.347 | $3.46 \times 10^{-28}$ |
| Rheumatoid arthritis | 3.139 | $4.96 \times 10^{-62}$ |
| SLE | -0.051 | 0.877 |
| Psoriasis | 0.188 | 0.432 |
| Allergic disease |  |  |
| Atopic dermatitis | -0.735 | $3.33 \times 10^{-7}$ |
| Asthma | 0.738 | $5.11 \times 10^{-20}$ |
| Pediatric asthma | 0.039 | 0.878 |
| Pollinosis | 0.210 | $2.42 \times 10^{-5}$ |
| Chronic sinusitis | 1.405 | $8.64 \times 10^{-28}$ |
| Allergic conjunctivitis | 0.053 | 0.573 |
| Urticaria | -0.157 | 0.0126 |
| Infection & Respiratory disease |  |  |
| Pulmonary tuberculosis | 1.065 | $1.26 \times 10^{-31}$ |
| Chronic hepatitis B | 1.123 | $6.03 \times 10^{-11}$ |
| Chronic hepatitis C | -0.487 | $3.39 \times 10^{-9}$ |
| Zoster infection | 0.029 | 0.816 |
| COVID-19 | -1.154 | $1.82 \times 10^{-9}$ |
| Pneumonia | 0.427 | $1.19 \times 10^{-7}$ |
| Interstitial lung disease | 0.736 | $5.53 \times 10^{-5}$ |
| Cardiovascular & other disease |  |  |
| Coronary artery disease | -0.338 | $1.45 \times 10^{-9}$ |
| Myocardial infarction | -0.581 | $1.46 \times 10^{-16}$ |
| Ischemic stroke | -0.165 | $3.73 \times 10^{-4}$ |
| Subarachnoid hemorrhage | 0.348 | 0.0405 |
| Parkinson disease | -1.626 | $7.21 \times 10^{-7}$ |
| Ulcerative colitis | -3.203 | $3.30 \times 10^{-15}$ |
| Iron deficiency anemia | -0.020 | 0.82 |
| Quantitative traits |  |  |
| Height | -0.143 | $1.39 \times 10^{-23}$ |
| Body Weight | -0.177 | $8.16 \times 10^{-39}$ |
| BMI | -0.122 | $1.20 \times 10^{-15}$ |
| SBP: Systolic blood pressure | -0.025 | 0.0685 |
| DBP: Diastolic blood pressure | -0.014 | 0.344 |
| RBC count | 0.019 | 0.278 |
| WBC count | 0.040 | 0.0512 |
| Eosinophil count | -0.159 | $2.99 \times 10^{-13}$ |
| Basophil count | -0.244 | $4.83 \times 10^{-31}$ |
| TC: Total cholesterol | -0.135 | $2.51 \times 10^{-14}$ |
| HDLC: HDL-cholesterol | 0.164 | $2.28 \times 10^{-9}$ |
| LDLC: LDL-cholesterol | -0.137 | $3.27 \times 10^{-11}$ |
| TG: Triglycerides | -0.128 | $7.68 \times 10^{-8}$ |
| HbA1c | 0.054 | $2.13 \times 10^{-3}$ |

Font colors: Red indicates a positive association, Blue indicates an inverse association.
Background colors: Red indicates robust positive association (full IV set analysis), Blue indicates robust inverse association (full IV set analysis).

**Supplementary Table 21.** MR restricted to NF-κB–related variants.

|  | IVW (fixed effects) |  | Weighted median |  | Weighted mode |  | MR-Egger |  |
| --- | --- | --- | --- | --- | --- | --- | --- | --- |
|  | Beta | P-value | Beta | P-value | Beta | P-value | Beta | P-value |
| Autoimmune disease & T2D |  |  |  |  |  |  |  |  |
| Type 1 diabetes | 0.525 | 0.177 | 0.539 | 0.244 | 0.739 | 0.354 | 2.007 | 0.375 |
| Type 2 diabetes | 0.577 | 0.139 | 0.627 | 0.204 | 0.961 | 0.296 | 2.508 | 0.324 |
| Hashimoto's disease | 0.822 | 0.154 | -0.178 | 0.821 | -0.870 | 0.634 | 6.667 | 0.185 |
| Grave's disease | -0.358 | 0.169 | -0.398 | 0.231 | -0.613 | 0.316 | -2.276 | 0.239 |
| Rheumatoid arthritis | 0.243 | 0.201 | 0.213 | 0.332 | 0.119 | 0.682 | 0.658 | 0.499 |
| SLE | 1.893 | 0.0111 | 1.703 | 0.0587 | 1.638 | 0.279 | 1.334 | 0.827 |
| Psoriasis | -0.537 | 0.282 | -1.491 | 0.0253 | -1.559 | 0.135 | -3.289 | 0.615 |
| Allergic disease |  |  |  |  |  |  |  |  |
| Atopic dermatitis | -0.007 | 0.974 | 0.169 | 0.58 | 0.255 | 0.552 | 0.699 | 0.814 |
| Asthma | 0.944 | 1.82×10<sup>-13</sup> | -0.194 | 0.32 | -0.222 | 0.324 | 5.016 | 0.0963 |
| Pediatric asthma | 1.798 | 1.87×10<sup>-3</sup> | 0.954 | 0.31 | -0.008 | 0.996 | 8.078 | 0.2 |
| Pollinosis | -0.092 | 0.407 | -0.079 | 0.604 | 0.161 | 0.452 | 1.289 | 0.184 |
| Chronic sinusitis | 0.907 | 6.37×10<sup>-6</sup> | 0.544 | 0.0693 | 0.343 | 0.491 | 2.879 | 0.151 |
| Allergic conjunctivitis | -0.059 | 0.785 | 0.042 | 0.868 | 0.056 | 0.87 | 0.335 | 0.733 |
| Urticaria | -0.155 | 0.268 | 0.012 | 0.946 | 0.041 | 0.845 | 0.406 | 0.723 |
| Infection & Respiratory disease |  |  |  |  |  |  |  |  |
| Pulmonary tuberculosis | 0.194 | 0.217 | 0.139 | 0.46 | 0.077 | 0.757 | -0.317 | 0.663 |
| Chronic hepatitis B | 1.347 | 3.19×10<sup>-6</sup> | 0.344 | 0.485 | 0.196 | 0.699 | 5.344 | 0.118 |
| Chronic hepatitis C | -0.135 | 0.407 | -0.409 | 0.0386 | -0.416 | 0.194 | -1.188 | 0.537 |
| Zoster infection | -0.210 | 0.462 | -0.098 | 0.774 | -0.034 | 0.939 | 0.099 | 0.937 |
| COVID-19 | 0.445 | 0.227 | -0.039 | 0.932 | -0.252 | 0.612 | -3.040 | 0.425 |
| Pneumonia | -0.110 | 0.489 | -0.055 | 0.788 | 0.135 | 0.63 | 1.434 | 0.232 |
| Interstitial lung disease | 0.107 | 0.797 | -0.287 | 0.564 | -0.305 | 0.655 | -1.407 | 0.611 |
| Cardiovascular & other disease |  |  |  |  |  |  |  |  |
| Coronary artery disease | -0.004 | 0.967 | 0.032 | 0.784 | -0.301 | 0.416 | -1.017 | 0.193 |
| Myocardial infarction | -0.184 | 0.143 | -0.048 | 0.791 | 0.199 | 0.675 | -1.475 | 0.193 |
| Ischemic stroke | -0.084 | 0.405 | -0.125 | 0.284 | -0.130 | 0.447 | -0.272 | 0.577 |
| Subarachnoid hemorrhage | 0.525 | 0.177 | 0.539 | 0.244 | 0.739 | 0.354 | 2.007 | 0.375 |
| Parkinson disease | 0.412 | 0.57 | 0.218 | 0.784 | 0.173 | 0.875 | -0.630 | 0.843 |
| Ulcerative colitis | -0.382 | 0.607 | -0.391 | 0.646 | 0.199 | 0.846 | 2.219 | 0.546 |
| Iron deficiency anemia | -0.340 | 0.0905 | -0.294 | 0.184 | -0.254 | 0.457 | 0.010 | 0.991 |
| Quantitative traits |  |  |  |  |  |  |  |  |
| Height | 0.040 | 0.0357 | 0.034 | 0.191 | -0.063 | 0.178 | -0.277 | 0.27 |
| Body Weight | 0.074 | 3.80×10<sup>-3</sup> | 0.053 | 0.117 | -0.021 | 0.648 | -0.322 | 0.169 |
| BMI | 0.061 | 0.0415 | 0.028 | 0.461 | 0.011 | 0.79 | -0.214 | 0.405 |
| SBP: Systolic blood pressure | -0.004 | 0.897 | -0.032 | 0.412 | -0.042 | 0.378 | -0.180 | 0.431 |
| DBP: Diastolic blood pressure | -0.025 | 0.447 | -0.038 | 0.358 | -0.062 | 0.361 | -0.265 | 0.256 |
| RBC count | -0.053 | 0.0693 | -0.058 | 0.133 | -0.072 | 0.269 | -0.073 | 0.795 |
| WBC count | 0.090 | 4.72×10<sup>-3</sup> | 0.085 | 0.0425 | 0.124 | 0.235 | 0.289 | 0.261 |
| Eosinophil count | 0.125 | 2.52×10<sup>-3</sup> | 0.141 | 0.017 | 0.178 | 0.145 | 0.522 | 0.224 |
| Basophil count | -0.124 | 3.43×10<sup>-3</sup> | -0.124 | 0.0158 | -0.145 | 0.143 | -0.233 | 0.355 |
| TC: Total cholesterol | -0.069 | 0.0393 | -0.078 | 0.0574 | -0.096 | 0.23 | -0.201 | 0.333 |
| HDLC: HDL-cholesterol | 0.043 | 0.337 | 0.030 | 0.617 | 0.002 | 0.991 | 0.460 | 0.205 |
| LDLC: LDL-cholesterol | -0.126 | 5.36×10<sup>-3</sup> | -0.151 | 9.82×10<sup>-3</sup> | -0.183 | 0.152 | -0.355 | 0.374 |
| TG: Triglycerides | -0.030 | 0.421 | -0.035 | 0.454 | -0.068 | 0.444 | -0.307 | 0.255 |
| HbA1c | -0.015 | 0.709 | -0.017 | 0.694 | -0.033 | 0.615 | -0.103 | 0.588 |

|  | pleiotropy test |  | heterogeneity test |  |  | IVW<br>(multiplicative random effects) |  |
| --- | --- | --- | --- | --- | --- | --- | --- |
|  | intercept | P-value | Cochran's Q | P-value | FDR Q-value | Beta | P-value |
| **Autoimmune disease & T2D** |  |  |  |  |  |  |  |
| Type 1 diabetes | -0.106 | 0.454 | 1.44 | 0.486 | 0.6 | 0.525 | 0.112 |
| Type 2 diabetes | -0.138 | 0.386 | 3.31 | 0.191 | 0.287 | 0.577 | 0.25 |
| Hashimoto's disease | -0.419 | 0.201 | 9.61 | 8.19×10<sup>-3</sup> | 0.0241 | 0.822 | 0.516 |
| Grave's disease | 0.138 | 0.268 | 5.55 | 0.0624 | 0.131 | -0.358 | 0.409 |
| Rheumatoid arthritis | -0.030 | 0.628 | 0.49 | 0.781 | 0.8 | 0.243 | 0.01 |
| SLE | 0.040 | 0.923 | 3.53 | 0.171 | 0.266 | 1.893 | 0.056 |
| Psoriasis | 0.199 | 0.653 | 10.55 | 5.12×10<sup>-3</sup> | 0.0215 | -0.537 | 0.639 |
| **Allergic disease** |  |  |  |  |  |  |  |
| Atopic dermatitis | -0.051 | 0.805 | 9.52 | 8.59×10<sup>-3</sup> | 0.0241 | -0.007 | 0.988 |
| Asthma | -0.291 | 0.113 | 94.38 | 3.21×10<sup>-21</sup> | 1.35×10<sup>-19</sup> | 0.944 | 0.284 |
| Pediatric asthma | -0.448 | 0.243 | 12.38 | 2.05×10<sup>-3</sup> | 0.0144 | 1.798 | 0.211 |
| Pollinosis | -0.099 | 0.165 | 14.53 | 7.00×10<sup>-4</sup> | 5.88×10<sup>-3</sup> | -0.092 | 0.759 |
| Chronic sinusitis | -0.141 | 0.207 | 8.91 | 0.0116 | 0.0287 | 0.907 | 0.0324 |
| Allergic conjunctivitis | -0.028 | 0.68 | 0.93 | 0.628 | 0.733 | -0.059 | 0.689 |
| Urticaria | -0.040 | 0.623 | 4.72 | 0.0942 | 0.157 | -0.155 | 0.471 |
| **Infection & Respiratory disease** |  |  |  |  |  |  |  |
| Pulmonary tuberculosis | 0.037 | 0.505 | 1.70 | 0.427 | 0.579 | 0.194 | 0.181 |
| Chronic hepatitis B | -0.285 | 0.15 | 18.26 | 1.08×10<sup>-4</sup> | 1.13×10<sup>-3</sup> | 1.347 | 0.123 |
| Chronic hepatitis C | 0.076 | 0.561 | 9.52 | 8.58×10<sup>-3</sup> | 0.0241 | -0.135 | 0.704 |
| Zoster infection | -0.022 | 0.799 | 1.00 | 0.606 | 0.727 | -0.210 | 0.298 |
| COVID-19 | 0.248 | 0.37 | 11.52 | 3.16×10<sup>-3</sup> | 0.019 | 0.445 | 0.614 |
| Pneumonia | -0.111 | 0.208 | 9.37 | 9.24×10<sup>-3</sup> | 0.0243 | -0.110 | 0.749 |
| Interstitial lung disease | 0.109 | 0.576 | 3.19 | 0.203 | 0.294 | 0.107 | 0.839 |
| **Cardiovascular & other disease** |  |  |  |  |  |  |  |
| Coronary artery disease | 0.073 | 0.185 | 11.20 | 3.70×10<sup>-3</sup> | 0.0194 | -0.004 | 0.986 |
| Myocardial infarction | 0.093 | 0.21 | 10.73 | 4.68×10<sup>-3</sup> | 0.0215 | -0.184 | 0.528 |
| Ischemic stroke | 0.014 | 0.671 | 0.65 | 0.722 | 0.778 | -0.084 | 0.145 |
| Subarachnoid hemorrhage | -0.106 | 0.454 | 1.44 | 0.486 | 0.6 | 0.525 | 0.112 |
| Parkinson disease | 0.075 | 0.739 | 0.34 | 0.844 | 0.844 | 0.412 | 0.168 |
| Ulcerative colitis | -0.187 | 0.482 | 1.44 | 0.486 | 0.6 | -0.382 | 0.545 |
| Iron deficiency anemia | -0.025 | 0.691 | 0.53 | 0.766 | 0.8 | -0.340 | 1.04×10<sup>-3</sup> |
| **Quantitative traits** |  |  |  |  |  |  |  |
| Height | 0.023 | 0.23 | 29.66 | 3.63×10<sup>-7</sup> | 7.62×10<sup>-6</sup> | 0.040 | 0.586 |
| Body Weight | 0.028 | 0.133 | 22.63 | 1.22×10<sup>-5</sup> | 1.71×10<sup>-4</sup> | 0.074 | 0.39 |
| BMI | 0.020 | 0.32 | 10.13 | 6.31×10<sup>-3</sup> | 0.0221 | 0.061 | 0.365 |
| SBP: Systolic blood pressure | 0.013 | 0.424 | 4.68 | 0.0961 | 0.157 | -0.004 | 0.932 |
| DBP: Diastolic blood pressure | 0.017 | 0.269 | 5.85 | 0.0536 | 0.118 | -0.025 | 0.657 |
| RBC count | 0.001 | 0.941 | 4.68 | 0.0964 | 0.157 | -0.053 | 0.235 |
| WBC count | -0.014 | 0.346 | 4.84 | 0.0887 | 0.157 | 0.090 | 0.0695 |
| Eosinophil count | -0.029 | 0.276 | 10.18 | 6.16×10<sup>-3</sup> | 0.0221 | 0.125 | 0.181 |
| Basophil count | 0.008 | 0.575 | 0.88 | 0.646 | 0.733 | -0.124 | 9.72×10<sup>-6</sup> |
| TC: Total cholesterol | 0.009 | 0.445 | 2.41 | 0.3 | 0.42 | -0.069 | 0.0603 |
| HDLC: HDL-cholesterol | -0.030 | 0.216 | 8.25 | 0.0161 | 0.0376 | 0.043 | 0.637 |
| LDLC: LDL-cholesterol | 0.016 | 0.495 | 4.66 | 0.0974 | 0.157 | -0.126 | 0.0681 |
| TG: Triglycerides | 0.020 | 0.269 | 5.39 | 0.0677 | 0.135 | -0.030 | 0.624 |
| HbA1c | 0.006 | 0.621 | 0.66 | 0.719 | 0.778 | -0.015 | 0.515 |

|  | MR-cML |  |
| --- | --- | --- |
|  | Theta | P-value |
| Autoimmune disease & T2D |  |  |
| Type 1 diabetes | 0.529 | 0.181 |
| Type 2 diabetes | 0.587 | 0.146 |
| Hashimoto's disease | 0.465 | 0.62 |
| Grave's disease | -0.361 | 0.193 |
| Rheumatoid arthritis | 0.244 | 0.205 |
| SLE | 1.940 | 0.0128 |
| Psoriasis | -0.916 | 0.196 |
| Allergic disease |  |  |
| Atopic dermatitis | 0.043 | 0.872 |
| Asthma | -0.219 | 0.218 |
| Pediatric asthma | 1.214 | 0.307 |
| Pollinosis | -0.363 | 0.116 |
| Chronic sinusitis | 0.898 | $1.34 \times 10^{-3}$ |
| Allergic conjunctivitis | -0.059 | 0.787 |
| Urticaria | -0.156 | 0.289 |
| Infection & Respiratory disease |  |  |
| Pulmonary tuberculosis | 0.196 | 0.22 |
| Chronic hepatitis B | 0.408 | 0.469 |
| Chronic hepatitis C | -0.211 | 0.304 |
| Zoster infection | -0.211 | 0.464 |
| COVID-19 | 0.238 | 0.65 |
| Pneumonia | -0.172 | 0.405 |
| Interstitial lung disease | 0.104 | 0.807 |
| Cardiovascular & other disease |  |  |
| Coronary artery disease | 0.101 | 0.557 |
| Myocardial infarction | -0.074 | 0.746 |
| Ischemic stroke | -0.084 | 0.405 |
| Subarachnoid hemorrhage | 0.529 | 0.181 |
| Parkinson disease | 0.412 | 0.571 |
| Ulcerative colitis | -0.391 | 0.603 |
| Iron deficiency anemia | -0.341 | 0.0941 |
| Quantitative traits |  |  |
| Height | 0.137 | $1.21 \times 10^{-5}$ |
| Body Weight | 0.186 | $2.13 \times 10^{-5}$ |
| BMI | 0.057 | 0.125 |
| SBP: Systolic blood pressure | -0.005 | 0.873 |
| DBP: Diastolic blood pressure | -0.027 | 0.426 |
| RBC count | -0.056 | 0.0682 |
| WBC count | 0.092 | $7.41 \times 10^{-3}$ |
| Eosinophil count | 0.144 | $3.68 \times 10^{-3}$ |
| Basophil count | -0.124 | $4.67 \times 10^{-3}$ |
| TC: Total cholesterol | -0.071 | 0.0411 |
| HDLC: HDL-cholesterol | 0.033 | 0.554 |
| LDLC: LDL-cholesterol | -0.132 | $6.36 \times 10^{-3}$ |
| TG: Triglycerides | -0.030 | 0.456 |
| HbA1c | -0.015 | 0.71 |

Font colors: Red indicates a positive association, Blue indicates an inverse association.
Background colors: Red indicates robust positive association (full IV set analysis), Blue indicates robust inverse association (full IV set analysis).

**Supplementary Table 22.** MR excluding NF-κB–related variants.

|  | IVW (fixed effects) |  | Weighted median |  | Weighted mode |  | MR-Egger |  |
| --- | --- | --- | --- | --- | --- | --- | --- | --- |
|  | Beta | P-value | Beta | P-value | Beta | P-value | Beta | P-value |
| **Autoimmune disease & T2D** |  |  |  |  |  |  |  |  |
| Type 1 diabetes | 1.262 | 2.30×10<sup>-18</sup> | 1.514 | 1.95×10<sup>-12</sup> | 1.614 | 7.17×10<sup>-5</sup> | 1.536 | 0.0799 |
| Type 2 diabetes | 0.093 | 3.72×10<sup>-3</sup> | 0.128 | 0.0103 | 0.169 | 0.0141 | 0.327 | 0.105 |
| Hashimoto's disease | 1.289 | 1.89×10<sup>-9</sup> | 1.793 | 1.44×10<sup>-7</sup> | 2.050 | 1.57×10<sup>-4</sup> | 3.158 | 0.109 |
| Grave's disease | 1.810 | 4.01×10<sup>-77</sup> | 1.788 | 3.91×10<sup>-20</sup> | 2.183 | 2.18×10<sup>-6</sup> | 1.544 | 0.394 |
| Rheumatoid arthritis | 1.524 | 9.80×10<sup>-103</sup> | 0.730 | 1.27×10<sup>-5</sup> | 0.448 | 0.0165 | 2.647 | 0.158 |
| SLE | -0.117 | 0.673 | -0.059 | 0.883 | -0.165 | 0.749 | 0.026 | 0.985 |
| Psoriasis | 0.243 | 0.182 | 0.419 | 0.165 | 0.209 | 0.486 | 3.090 | 0.0805 |
| **Allergic disease** |  |  |  |  |  |  |  |  |
| Atopic dermatitis | -0.660 | 1.34×10<sup>-14</sup> | -0.565 | 8.72×10<sup>-6</sup> | -0.605 | 1.65×10<sup>-3</sup> | -1.366 | 0.0174 |
| Asthma | 0.350 | 1.44×10<sup>-13</sup> | 0.227 | 0.0322 | 0.014 | 0.939 | 0.438 | 0.382 |
| Pediatric asthma | 0.063 | 0.766 | 0.295 | 0.332 | 0.355 | 0.387 | -0.956 | 0.31 |
| Pollinosis | 0.181 | 1.19×10<sup>-5</sup> | 0.165 | 5.67×10<sup>-3</sup> | 0.155 | 0.0575 | 0.192 | 0.369 |
| Chronic sinusitis | 0.786 | 5.09×10<sup>-26</sup> | 0.596 | 2.11×10<sup>-4</sup> | 0.385 | 0.275 | 1.619 | 0.101 |
| Allergic conjunctivitis | 0.150 | 0.0634 | 0.097 | 0.36 | 0.078 | 0.599 | -0.345 | 0.263 |
| Urticaria | -0.100 | 0.0543 | -0.180 | 0.0155 | -0.194 | 0.0422 | 0.028 | 0.903 |
| **Infection & Respiratory disease** |  |  |  |  |  |  |  |  |
| Pulmonary tuberculosis | 0.805 | 2.54×10<sup>-43</sup> | 0.876 | 4.37×10<sup>-17</sup> | 0.945 | 2.68×10<sup>-6</sup> | 1.655 | 5.04×10<sup>-3</sup> |
| Chronic hepatitis B | 1.524 | 8.43×10<sup>-46</sup> | 0.923 | 7.24×10<sup>-7</sup> | 0.920 | 1.20×10<sup>-4</sup> | 0.697 | 0.674 |
| Chronic hepatitis C | -0.318 | 1.18×10<sup>-7</sup> | -0.322 | 1.35×10<sup>-3</sup> | -0.349 | 0.0197 | -0.325 | 0.6 |
| Zoster infection | -0.008 | 0.94 | 0.114 | 0.435 | 0.119 | 0.586 | -0.467 | 0.248 |
| COVID-19 | -0.752 | 4.42×10<sup>-7</sup> | -1.013 | 2.81×10<sup>-6</sup> | -1.041 | 9.10×10<sup>-4</sup> | -1.860 | 0.0162 |
| Pneumonia | 0.333 | 1.53×10<sup>-8</sup> | 0.357 | 2.35×10<sup>-4</sup> | 0.483 | 2.37×10<sup>-3</sup> | 0.585 | 0.129 |
| Interstitial lung disease | 0.479 | 1.85×10<sup>-3</sup> | 0.603 | 4.36×10<sup>-3</sup> | 0.596 | 0.0222 | 1.271 | 0.124 |
| **Cardiovascular & other disease** |  |  |  |  |  |  |  |  |
| Coronary artery disease | -0.235 | 5.48×10<sup>-12</sup> | -0.256 | 1.93×10<sup>-5</sup> | -0.259 | 5.71×10<sup>-3</sup> | -0.115 | 0.76 |
| Myocardial infarction | -0.350 | 5.94×10<sup>-14</sup> | -0.433 | 4.88×10<sup>-7</sup> | -0.457 | 2.03×10<sup>-3</sup> | -0.222 | 0.648 |
| Ischemic stroke | -0.137 | 2.20×10<sup>-4</sup> | -0.169 | 7.35×10<sup>-4</sup> | -0.175 | 0.0181 | -0.263 | 0.0818 |
| Subarachnoid hemorrhage | 0.269 | 0.0618 | 0.451 | 0.0174 | 0.476 | 0.0966 | 0.446 | 0.412 |
| Parkinson disease | -1.345 | 5.85×10<sup>-7</sup> | -1.554 | 9.13×10<sup>-5</sup> | -1.669 | 0.0108 | -1.927 | 0.157 |
| Ulcerative colitis | -1.646 | 2.95×10<sup>-9</sup> | -1.786 | 9.21×10<sup>-4</sup> | -3.237 | 7.30×10<sup>-4</sup> | -2.451 | 0.355 |
| Iron deficiency anemia | -0.013 | 0.864 | -0.069 | 0.47 | -0.079 | 0.549 | 0.004 | 0.989 |
| **Quantitative traits** |  |  |  |  |  |  |  |  |
| Height | -0.097 | 4.54×10<sup>-44</sup> | -0.090 | 1.06×10<sup>-11</sup> | -0.096 | 3.87×10<sup>-4</sup> | -0.144 | 0.0909 |
| Body Weight | -0.142 | 4.01×10<sup>-51</sup> | -0.150 | 1.55×10<sup>-15</sup> | -0.182 | 2.70×10<sup>-5</sup> | -0.256 | 6.88×10<sup>-3</sup> |
| BMI | -0.102 | 3.26×10<sup>-20</sup> | -0.110 | 7.21×10<sup>-9</sup> | -0.138 | 9.64×10<sup>-5</sup> | -0.206 | 0.0126 |
| SBP: Systolic blood pressure | -0.023 | 0.0473 | -0.022 | 0.185 | -0.014 | 0.533 | -0.056 | 0.287 |
| DBP: Diastolic blood pressure | -0.004 | 0.728 | 0.005 | 0.771 | 0.004 | 0.85 | -0.054 | 0.276 |
| RBC count | -0.059 | 6.52×10<sup>-8</sup> | -0.008 | 0.698 | 0.007 | 0.76 | -0.030 | 0.811 |
| WBC count | -0.049 | 4.01×10<sup>-5</sup> | 0.027 | 0.192 | 0.055 | 8.94×10<sup>-3</sup> | 0.014 | 0.931 |
| Eosinophil count | -0.147 | 1.91×10<sup>-21</sup> | -0.134 | 1.83×10<sup>-7</sup> | -0.147 | 5.06×10<sup>-5</sup> | -0.182 | 0.378 |
| Basophil count | -0.186 | 2.06×10<sup>-32</sup> | -0.202 | 8.43×10<sup>-15</sup> | -0.232 | 4.33×10<sup>-6</sup> | -0.222 | 0.0873 |
| TC: Total cholesterol | -0.102 | 2.08×10<sup>-16</sup> | -0.116 | 6.94×10<sup>-9</sup> | -0.119 | 1.38×10<sup>-3</sup> | -0.091 | 0.34 |
| HDLC: HDL-cholesterol | 0.094 | 1.00×10<sup>-8</sup> | 0.094 | 3.42×10<sup>-3</sup> | 0.189 | 4.06×10<sup>-3</sup> | 0.217 | 0.108 |
| LDLC: LDL-cholesterol | -0.109 | 7.31×10<sup>-11</sup> | -0.122 | 2.33×10<sup>-6</sup> | -0.125 | 2.21×10<sup>-3</sup> | -0.075 | 0.41 |
| TG: Triglycerides | -0.090 | 1.01×10<sup>-10</sup> | -0.121 | 1.54×10<sup>-7</sup> | -0.127 | 1.75×10<sup>-4</sup> | -0.076 | 0.589 |
| HbA1c | 0.037 | 0.0116 | 0.056 | 6.98×10<sup>-3</sup> | 0.055 | 0.07 | 0.139 | 0.034 |

|  | pleiotropy test |  | heterogeneity test |  |  | IVW<br>(multiplicative random effects) |  |
| --- | --- | --- | --- | --- | --- | --- | --- |
|  | intercept | P-value | Cochran's Q | P-value | FDR Q-value | Beta | P-value |
| **Autoimmune disease & T2D** |  |  |  |  |  |  |  |
| Type 1 diabetes | -0.027 | 0.727 | 25.81 | 0.0114 | 0.0177 | 1.262 | 2.52×10<sup>-9</sup> |
| Type 2 diabetes | -0.023 | 0.215 | 32.59 | 1.12×10<sup>-3</sup> | 1.96×10<sup>-3</sup> | 0.093 | 0.0784 |
| Hashimoto's disease | -0.186 | 0.306 | 66.33 | 1.55×10<sup>-9</sup> | 3.62×10<sup>-9</sup> | 1.289 | 0.0106 |
| Grave's disease | 0.026 | 0.877 | 267.40 | 3.18×10<sup>-50</sup> | 6.68×10<sup>-49</sup> | 1.810 | 8.23×10<sup>-5</sup> |
| Rheumatoid arthritis | -0.112 | 0.518 | 534.55 | 9.70×10<sup>-107</sup> | 4.07×10<sup>-105</sup> | 1.524 | 1.26×10<sup>-3</sup> |
| SLE | -0.014 | 0.914 | 19.72 | 0.0725 | 0.0982 | -0.117 | 0.742 |
| Psoriasis | -0.265 | 0.0899 | 129.65 | 2.49×10<sup>-20</sup> | 1.49×10<sup>-19</sup> | 0.243 | 0.65 |
| **Allergic disease** |  |  |  |  |  |  |  |
| Atopic dermatitis | 0.070 | 0.161 | 32.83 | 1.03×10<sup>-3</sup> | 1.88×10<sup>-3</sup> | -0.660 | 3.22×10<sup>-6</sup> |
| Asthma | -0.009 | 0.853 | 86.56 | 2.28×10<sup>-13</sup> | 7.98×10<sup>-13</sup> | 0.350 | 5.92×10<sup>-3</sup> |
| Pediatric asthma | 0.101 | 0.263 | 16.71 | 0.161 | 0.199 | 0.063 | 0.801 |
| Pollinosis | -0.001 | 0.956 | 20.71 | 0.0547 | 0.0766 | 0.181 | 8.58×10<sup>-4</sup> |
| Chronic sinusitis | -0.083 | 0.359 | 134.53 | 7.59×10<sup>-23</sup> | 5.31×10<sup>-22</sup> | 0.786 | 1.63×10<sup>-3</sup> |
| Allergic conjunctivitis | 0.049 | 0.106 | 11.94 | 0.451 | 0.486 | 0.150 | 0.0628 |
| Urticaria | -0.013 | 0.562 | 15.97 | 0.193 | 0.225 | -0.100 | 0.0953 |
| **Infection & Respiratory disease** |  |  |  |  |  |  |  |
| Pulmonary tuberculosis | -0.085 | 0.0888 | 73.35 | 7.52×10<sup>-11</sup> | 2.11×10<sup>-10</sup> | 0.805 | 2.38×10<sup>-8</sup> |
| Chronic hepatitis B | 0.082 | 0.605 | 195.01 | 3.48×10<sup>-35</sup> | 4.87×10<sup>-34</sup> | 1.524 | 4.25×10<sup>-4</sup> |
| Chronic hepatitis C | 0.001 | 0.992 | 84.43 | 5.85×10<sup>-13</sup> | 1.89×10<sup>-12</sup> | -0.318 | 0.0458 |
| Zoster infection | 0.046 | 0.238 | 11.39 | 0.496 | 0.521 | -0.008 | 0.939 |
| COVID-19 | 0.100 | 0.11 | 32.36 | 5.75×10<sup>-3</sup> | 9.29×10<sup>-3</sup> | -0.752 | 5.86×10<sup>-4</sup> |
| Pneumonia | -0.025 | 0.478 | 32.38 | 1.21×10<sup>-3</sup> | 2.03×10<sup>-3</sup> | 0.333 | 5.72×10<sup>-4</sup> |
| Interstitial lung disease | -0.079 | 0.303 | 22.82 | 0.0293 | 0.0424 | 0.479 | 0.024 |
| **Cardiovascular & other disease** |  |  |  |  |  |  |  |
| Coronary artery disease | -0.012 | 0.74 | 98.71 | 9.96×10<sup>-16</sup> | 4.18×10<sup>-15</sup> | -0.235 | 0.0163 |
| Myocardial infarction | -0.013 | 0.785 | 87.72 | 1.36×10<sup>-13</sup> | 5.19×10<sup>-13</sup> | -0.350 | 5.48×10<sup>-3</sup> |
| Ischemic stroke | 0.012 | 0.362 | 12.35 | 0.418 | 0.462 | -0.137 | 2.71×10<sup>-4</sup> |
| Subarachnoid hemorrhage | -0.018 | 0.732 | 7.98 | 0.787 | 0.806 | 0.269 | 0.022 |
| Parkinson disease | 0.058 | 0.643 | 19.08 | 0.0867 | 0.114 | -1.345 | 7.42×10<sup>-5</sup> |
| Ulcerative colitis | 0.080 | 0.748 | 71.16 | 1.94×10<sup>-10</sup> | 5.09×10<sup>-10</sup> | -1.646 | 0.0148 |
| Iron deficiency anemia | -0.002 | 0.95 | 7.14 | 0.849 | 0.849 | -0.013 | 0.824 |
| **Quantitative traits** |  |  |  |  |  |  |  |
| Height | 0.005 | 0.546 | 107.12 | 2.21×10<sup>-17</sup> | 1.16×10<sup>-16</sup> | -0.097 | 3.16×10<sup>-6</sup> |
| Body Weight | 0.011 | 0.151 | 68.71 | 5.57×10<sup>-10</sup> | 1.38×10<sup>-9</sup> | -0.142 | 3.27×10<sup>-10</sup> |
| BMI | 0.010 | 0.146 | 40.15 | 6.78×10<sup>-5</sup> | 1.29×10<sup>-4</sup> | -0.102 | 4.78×10<sup>-7</sup> |
| SBP: Systolic blood pressure | 0.003 | 0.508 | 16.14 | 0.185 | 0.222 | -0.023 | 0.0872 |
| DBP: Diastolic blood pressure | 0.005 | 0.296 | 13.83 | 0.312 | 0.354 | -0.004 | 0.746 |
| RBC count | -0.003 | 0.81 | 105.90 | 3.86×10<sup>-17</sup> | 1.80×10<sup>-16</sup> | -0.059 | 0.0689 |
| WBC count | -0.006 | 0.687 | 149.82 | 6.16×10<sup>-26</sup> | 6.47×10<sup>-25</sup> | -0.049 | 0.245 |
| Eosinophil count | 0.004 | 0.856 | 139.23 | 8.59×10<sup>-24</sup> | 7.22×10<sup>-23</sup> | -0.147 | 5.24×10<sup>-3</sup> |
| Basophil count | 0.004 | 0.759 | 48.13 | 2.96×10<sup>-6</sup> | 6.22×10<sup>-6</sup> | -0.186 | 3.25×10<sup>-9</sup> |
| TC: Total cholesterol | -0.001 | 0.898 | 45.07 | 1.00×10<sup>-5</sup> | 2.00×10<sup>-5</sup> | -0.102 | 2.23×10<sup>-5</sup> |
| HDLC: HDL-cholesterol | -0.012 | 0.327 | 52.43 | 5.20×10<sup>-7</sup> | 1.15×10<sup>-6</sup> | 0.094 | 6.12×10<sup>-3</sup> |
| LDLC: LDL-cholesterol | -0.003 | 0.701 | 23.77 | 0.0218 | 0.0327 | -0.109 | 3.69×10<sup>-6</sup> |
| TG: Triglycerides | -0.001 | 0.916 | 81.00 | 2.66×10<sup>-12</sup> | 7.98×10<sup>-12</sup> | -0.090 | 0.0128 |
| HbA1c | -0.010 | 0.092 | 16.77 | 0.158 | 0.199 | 0.037 | 0.0327 |

|  | MR-cML |  |
| --- | --- | --- |
|  | Theta | P-value |
| Autoimmune disease & T2D |  |  |
| Type 1 diabetes | 1.387 | $6.66 \times 10^{-16}$ |
| Type 2 diabetes | 0.089 | $9.17 \times 10^{-3}$ |
| Hashimoto's disease | 1.770 | $2.74 \times 10^{-8}$ |
| Grave's disease | 2.271 | $1.02 \times 10^{-33}$ |
| Rheumatoid arthritis | 0.582 | $1.50 \times 10^{-6}$ |
| SLE | -0.073 | 0.801 |
| Psoriasis | -0.063 | 0.778 |
| Allergic disease |  |  |
| Atopic dermatitis | -0.546 | $1.34 \times 10^{-7}$ |
| Asthma | 0.017 | 0.811 |
| Pediatric asthma | 0.075 | 0.732 |
| Pollinosis | 0.186 | $1.20 \times 10^{-5}$ |
| Chronic sinusitis | 0.296 | $9.02 \times 10^{-3}$ |
| Allergic conjunctivitis | 0.147 | 0.0772 |
| Urticaria | -0.104 | 0.0515 |
| Infection & Respiratory disease |  |  |
| Pulmonary tuberculosis | 0.890 | $7.15 \times 10^{-28}$ |
| Chronic hepatitis B | 0.852 | $1.11 \times 10^{-9}$ |
| Chronic hepatitis C | -0.336 | $7.73 \times 10^{-4}$ |
| Zoster infection | -0.007 | 0.949 |
| COVID-19 | -0.789 | $2.44 \times 10^{-7}$ |
| Pneumonia | 0.353 | $2.89 \times 10^{-8}$ |
| Interstitial lung disease | 0.508 | $1.64 \times 10^{-3}$ |
| Cardiovascular & other disease |  |  |
| Coronary artery disease | -0.162 | $3.56 \times 10^{-4}$ |
| Myocardial infarction | -0.340 | $7.49 \times 10^{-3}$ |
| Ischemic stroke | -0.142 | $2.36 \times 10^{-4}$ |
| Subarachnoid hemorrhage | 0.272 | 0.061 |
| Parkinson disease | -1.378 | $6.84 \times 10^{-7}$ |
| Ulcerative colitis | -2.364 | $2.07 \times 10^{-7}$ |
| Iron deficiency anemia | -0.014 | 0.855 |
| Quantitative traits |  |  |
| Height | -0.115 | $1.06 \times 10^{-26}$ |
| Body Weight | -0.162 | $2.29 \times 10^{-34}$ |
| BMI | -0.113 | $3.35 \times 10^{-17}$ |
| SBP: Systolic blood pressure | -0.024 | 0.0454 |
| DBP: Diastolic blood pressure | -0.004 | 0.718 |
| RBC count | 0.001 | 0.952 |
| WBC count | 0.042 | 0.0113 |
| Eosinophil count | -0.127 | $1.31 \times 10^{-11}$ |
| Basophil count | -0.197 | $1.83 \times 10^{-27}$ |
| TC: Total cholesterol | -0.110 | $2.73 \times 10^{-15}$ |
| HDLC: HDL-cholesterol | 0.119 | $1.21 \times 10^{-10}$ |
| LDLC: LDL-cholesterol | -0.112 | $1.30 \times 10^{-10}$ |
| TG: Triglycerides | -0.113 | $3.10 \times 10^{-10}$ |
| HbA1c | 0.038 | 0.0109 |

Font colors: Red indicates a positive association, Blue indicates an inverse association.
Background colors: Red indicates robust positive association (full IV set analysis), Blue indicates robust inverse association (full IV set analysis).

**Supplementary Table 23.**
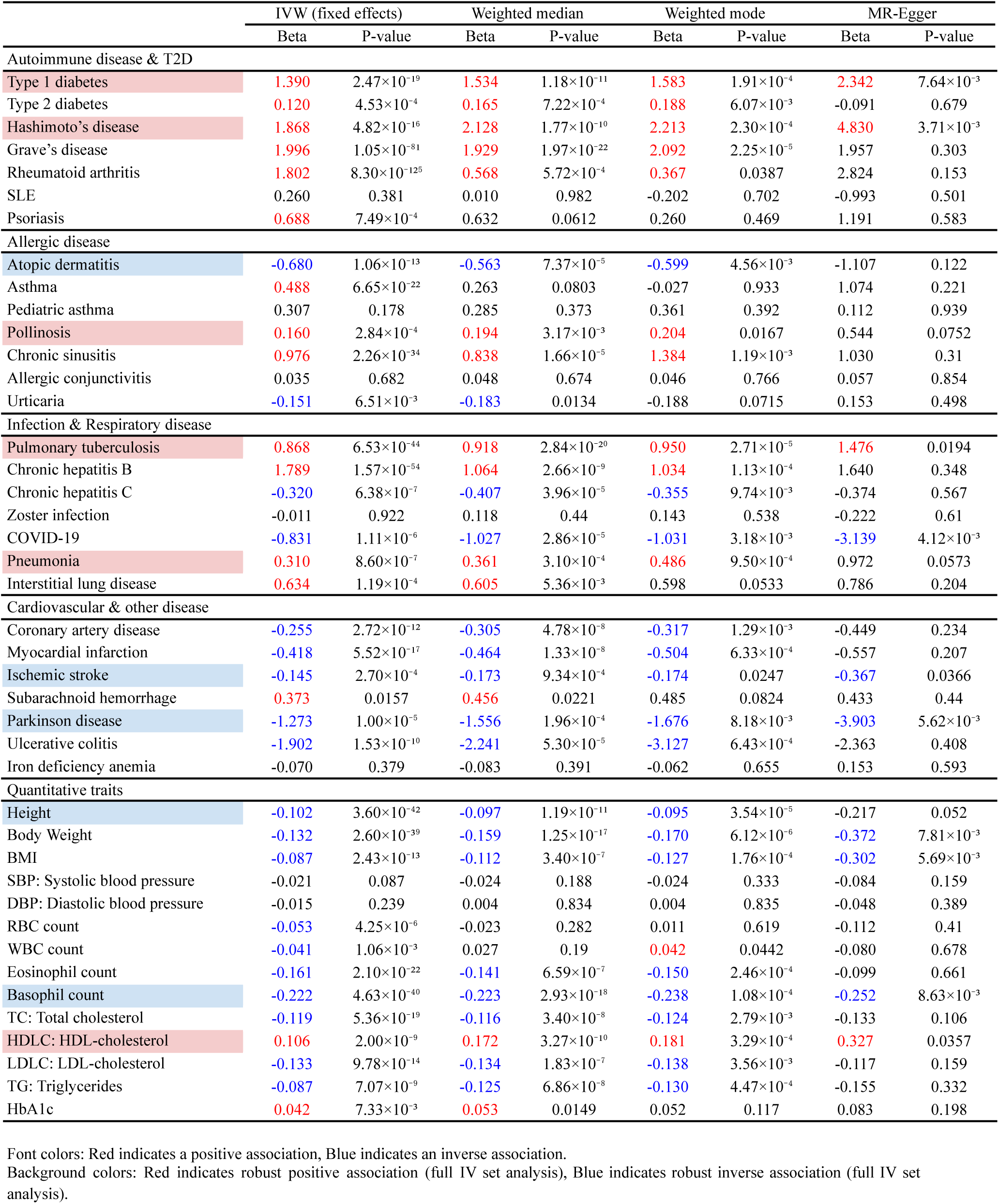
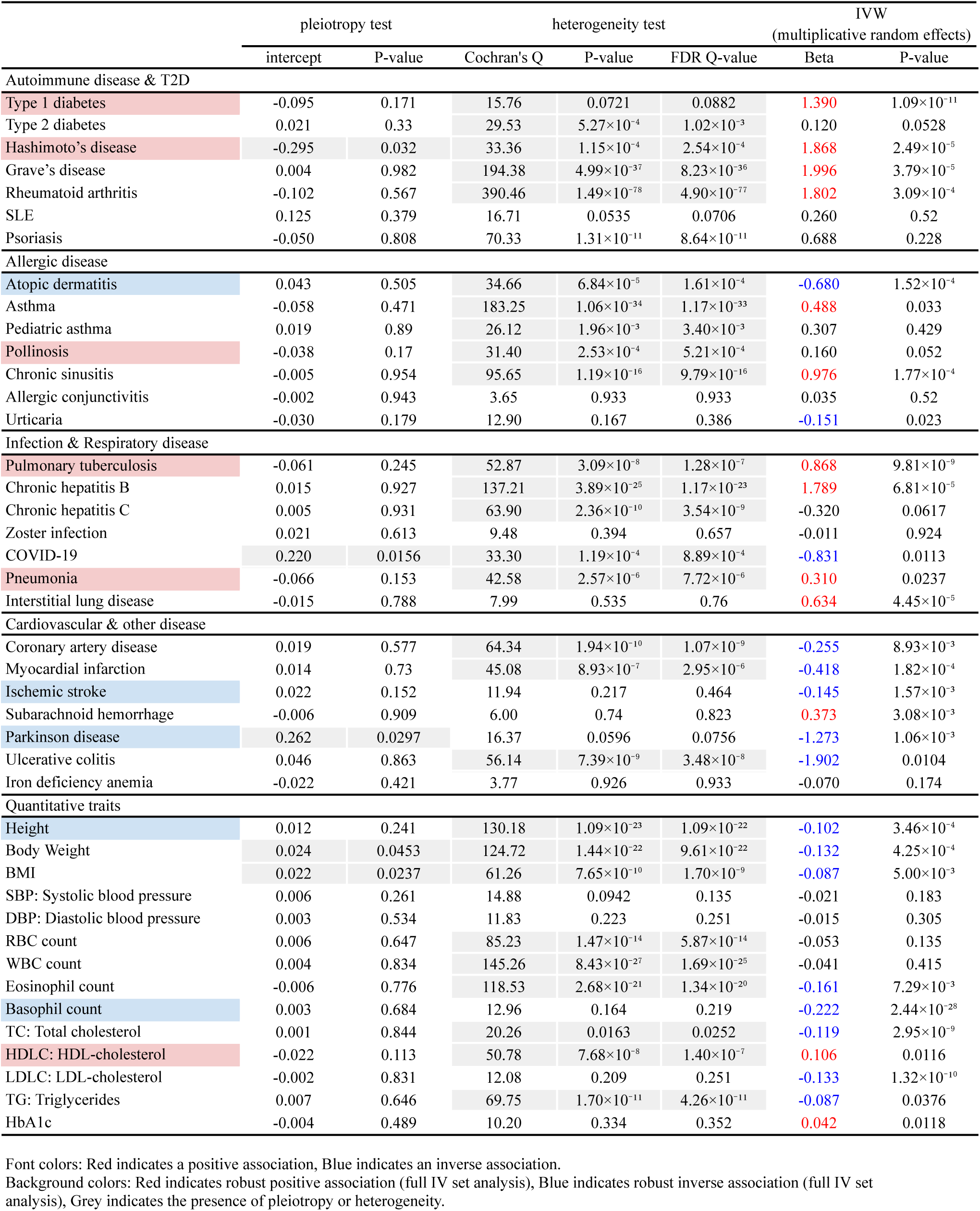
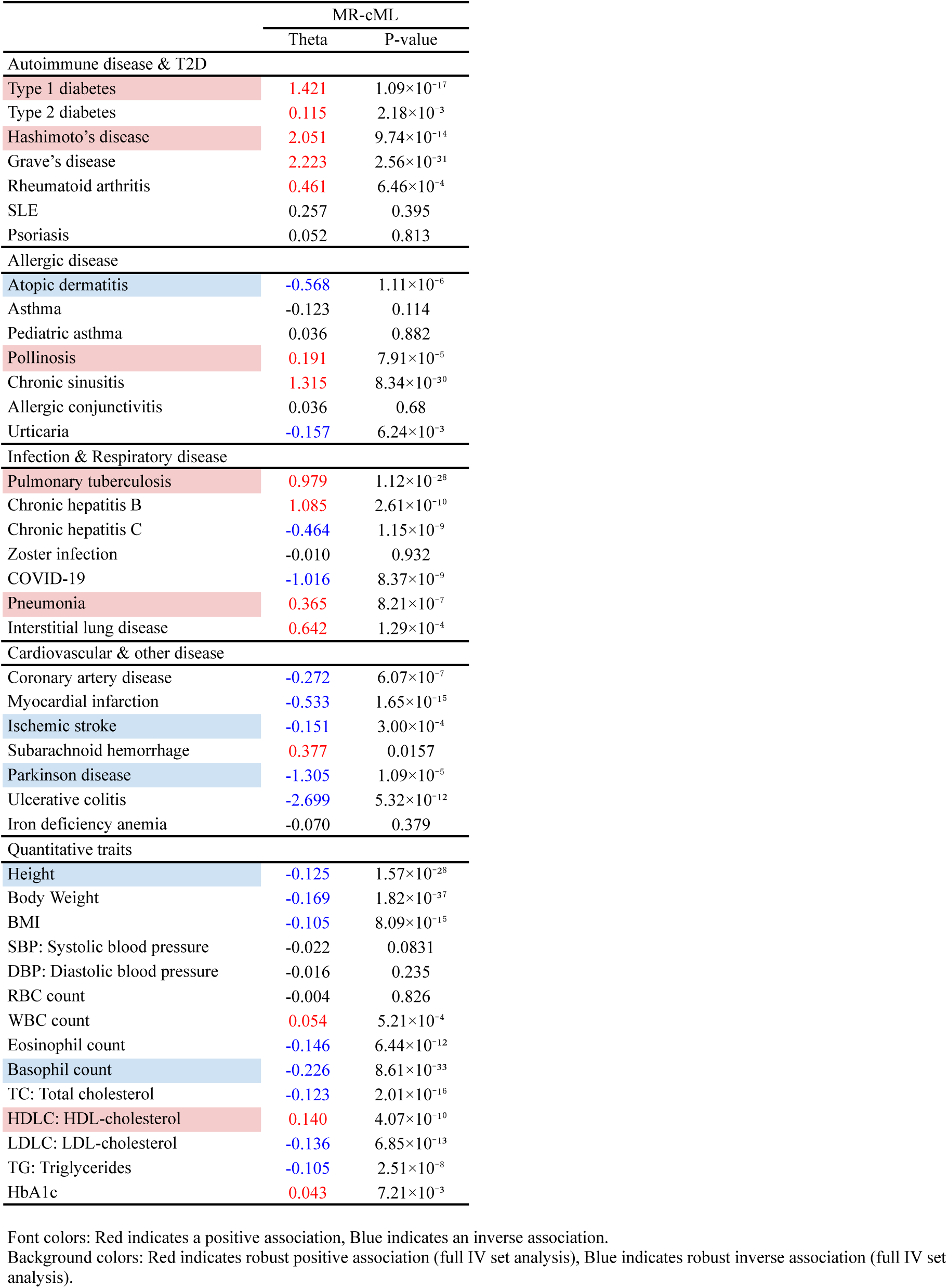
MR restricted to HLA class II and NF-κB–related variants.

|  | IVW (fixed effects) |  | Weighted median |  | Weighted mode |  | MR-Egger |  |
| --- | --- | --- | --- | --- | --- | --- | --- | --- |
|  | Beta | P-value | Beta | P-value | Beta | P-value | Beta | P-value |
| **Autoimmune disease & T2D** |  |  |  |  |  |  |  |  |
| Type 1 diabetes | 1.390 | 2.47×10<sup>-19</sup> | 1.534 | 1.18×10<sup>-11</sup> | 1.583 | 1.91×10<sup>-4</sup> | 2.342 | 7.64×10<sup>-3</sup> |
| Type 2 diabetes | 0.120 | 4.53×10<sup>-4</sup> | 0.165 | 7.22×10<sup>-4</sup> | 0.188 | 6.07×10<sup>-3</sup> | -0.091 | 0.679 |
| Hashimoto's disease | 1.868 | 4.82×10<sup>-16</sup> | 2.128 | 1.77×10<sup>-10</sup> | 2.213 | 2.30×10<sup>-4</sup> | 4.830 | 3.71×10<sup>-3</sup> |
| Grave's disease | 1.996 | 1.05×10<sup>-81</sup> | 1.929 | 1.97×10<sup>-22</sup> | 2.092 | 2.25×10<sup>-5</sup> | 1.957 | 0.303 |
| Rheumatoid arthritis | 1.802 | 8.30×10<sup>-125</sup> | 0.568 | 5.72×10<sup>-4</sup> | 0.367 | 0.0387 | 2.824 | 0.153 |
| SLE | 0.260 | 0.381 | 0.010 | 0.982 | -0.202 | 0.702 | -0.993 | 0.501 |
| Psoriasis | 0.688 | 7.49×10<sup>-4</sup> | 0.632 | 0.0612 | 0.260 | 0.469 | 1.191 | 0.583 |
| **Allergic disease** |  |  |  |  |  |  |  |  |
| Atopic dermatitis | -0.680 | 1.06×10<sup>-13</sup> | -0.563 | 7.37×10<sup>-5</sup> | -0.599 | 4.56×10<sup>-3</sup> | -1.107 | 0.122 |
| Asthma | 0.488 | 6.65×10<sup>-22</sup> | 0.263 | 0.0803 | -0.027 | 0.933 | 1.074 | 0.221 |
| Pediatric asthma | 0.307 | 0.178 | 0.285 | 0.373 | 0.361 | 0.392 | 0.112 | 0.939 |
| Pollinosis | 0.160 | 2.84×10<sup>-4</sup> | 0.194 | 3.17×10<sup>-3</sup> | 0.204 | 0.0167 | 0.544 | 0.0752 |
| Chronic sinusitis | 0.976 | 2.26×10<sup>-34</sup> | 0.838 | 1.66×10<sup>-5</sup> | 1.384 | 1.19×10<sup>-3</sup> | 1.030 | 0.31 |
| Allergic conjunctivitis | 0.035 | 0.682 | 0.048 | 0.674 | 0.046 | 0.766 | 0.057 | 0.854 |
| Urticaria | -0.151 | 6.51×10<sup>-3</sup> | -0.183 | 0.0134 | -0.188 | 0.0715 | 0.153 | 0.498 |
| **Infection & Respiratory disease** |  |  |  |  |  |  |  |  |
| Pulmonary tuberculosis | 0.868 | 6.53×10<sup>-44</sup> | 0.918 | 2.84×10<sup>-20</sup> | 0.950 | 2.71×10<sup>-5</sup> | 1.476 | 0.0194 |
| Chronic hepatitis B | 1.789 | 1.57×10<sup>-54</sup> | 1.064 | 2.66×10<sup>-9</sup> | 1.034 | 1.13×10<sup>-4</sup> | 1.640 | 0.348 |
| Chronic hepatitis C | -0.320 | 6.38×10<sup>-7</sup> | -0.407 | 3.96×10<sup>-5</sup> | -0.355 | 9.74×10<sup>-3</sup> | -0.374 | 0.567 |
| Zoster infection | -0.011 | 0.922 | 0.118 | 0.44 | 0.143 | 0.538 | -0.222 | 0.61 |
| COVID-19 | -0.831 | 1.11×10<sup>-6</sup> | -1.027 | 2.86×10<sup>-5</sup> | -1.031 | 3.18×10<sup>-3</sup> | -3.139 | 4.12×10<sup>-3</sup> |
| Pneumonia | 0.310 | 8.60×10<sup>-7</sup> | 0.361 | 3.10×10<sup>-4</sup> | 0.486 | 9.50×10<sup>-4</sup> | 0.972 | 0.0573 |
| Interstitial lung disease | 0.634 | 1.19×10<sup>-4</sup> | 0.605 | 5.36×10<sup>-3</sup> | 0.598 | 0.0533 | 0.786 | 0.204 |
| **Cardiovascular & other disease** |  |  |  |  |  |  |  |  |
| Coronary artery disease | -0.255 | 2.72×10<sup>-12</sup> | -0.305 | 4.78×10<sup>-8</sup> | -0.317 | 1.29×10<sup>-3</sup> | -0.449 | 0.234 |
| Myocardial infarction | -0.418 | 5.52×10<sup>-17</sup> | -0.464 | 1.33×10<sup>-8</sup> | -0.504 | 6.33×10<sup>-4</sup> | -0.557 | 0.207 |
| Ischemic stroke | -0.145 | 2.70×10<sup>-4</sup> | -0.173 | 9.34×10<sup>-4</sup> | -0.174 | 0.0247 | -0.367 | 0.0366 |
| Subarachnoid hemorrhage | 0.373 | 0.0157 | 0.456 | 0.0221 | 0.485 | 0.0824 | 0.433 | 0.44 |
| Parkinson disease | -1.273 | 1.00×10<sup>-5</sup> | -1.556 | 1.96×10<sup>-4</sup> | -1.676 | 8.18×10<sup>-3</sup> | -3.903 | 5.62×10<sup>-3</sup> |
| Ulcerative colitis | -1.902 | 1.53×10<sup>-10</sup> | -2.241 | 5.30×10<sup>-5</sup> | -3.127 | 6.43×10<sup>-4</sup> | -2.363 | 0.408 |
| Iron deficiency anemia | -0.070 | 0.379 | -0.083 | 0.391 | -0.062 | 0.655 | 0.153 | 0.593 |
| **Quantitative traits** |  |  |  |  |  |  |  |  |
| Height | -0.102 | 3.60×10<sup>-42</sup> | -0.097 | 1.19×10<sup>-11</sup> | -0.095 | 3.54×10<sup>-5</sup> | -0.217 | 0.052 |
| Body Weight | -0.132 | 2.60×10<sup>-39</sup> | -0.159 | 1.25×10<sup>-17</sup> | -0.170 | 6.12×10<sup>-6</sup> | -0.372 | 7.81×10<sup>-3</sup> |
| BMI | -0.087 | 2.43×10<sup>-13</sup> | -0.112 | 3.40×10<sup>-7</sup> | -0.127 | 1.76×10<sup>-4</sup> | -0.302 | 5.69×10<sup>-3</sup> |
| SBP: Systolic blood pressure | -0.021 | 0.087 | -0.024 | 0.188 | -0.024 | 0.333 | -0.084 | 0.159 |
| DBP: Diastolic blood pressure | -0.015 | 0.239 | 0.004 | 0.834 | 0.004 | 0.835 | -0.048 | 0.389 |
| RBC count | -0.053 | 4.25×10<sup>-6</sup> | -0.023 | 0.282 | 0.011 | 0.619 | -0.112 | 0.41 |
| WBC count | -0.041 | 1.06×10<sup>-3</sup> | 0.027 | 0.19 | 0.042 | 0.0442 | -0.080 | 0.678 |
| Eosinophil count | -0.161 | 2.10×10<sup>-22</sup> | -0.141 | 6.59×10<sup>-7</sup> | -0.150 | 2.46×10<sup>-4</sup> | -0.099 | 0.661 |
| Basophil count | -0.222 | 4.63×10<sup>-40</sup> | -0.223 | 2.93×10<sup>-18</sup> | -0.238 | 1.08×10<sup>-4</sup> | -0.252 | 8.63×10<sup>-3</sup> |
| TC: Total cholesterol | -0.119 | 5.36×10<sup>-19</sup> | -0.116 | 3.40×10<sup>-8</sup> | -0.124 | 2.79×10<sup>-3</sup> | -0.133 | 0.106 |
| HDLC: HDL-cholesterol | 0.106 | 2.00×10<sup>-9</sup> | 0.172 | 3.27×10<sup>-10</sup> | 0.181 | 3.29×10<sup>-4</sup> | 0.327 | 0.0357 |
| LDLC: LDL-cholesterol | -0.133 | 9.78×10<sup>-14</sup> | -0.134 | 1.83×10<sup>-7</sup> | -0.138 | 3.56×10<sup>-3</sup> | -0.117 | 0.159 |
| TG: Triglycerides | -0.087 | 7.07×10<sup>-9</sup> | -0.125 | 6.86×10<sup>-8</sup> | -0.130 | 4.47×10<sup>-4</sup> | -0.155 | 0.332 |
| HbA1c | 0.042 | 7.33×10<sup>-3</sup> | 0.053 | 0.0149 | 0.052 | 0.117 | 0.083 | 0.198 |

|  | pleiotropy test |  | heterogeneity test |  |  | IVW<br>(multiplicative random effects) |  |
| --- | --- | --- | --- | --- | --- | --- | --- |
|  | intercept | P-value | Cochran's Q | P-value | FDR Q-value | Beta | P-value |
| **Autoimmune disease & T2D** |  |  |  |  |  |  |  |
| Type 1 diabetes | -0.095 | 0.171 | 15.76 | 0.0721 | 0.0882 | 1.390 | 1.09×10<sup>-11</sup> |
| Type 2 diabetes | 0.021 | 0.33 | 29.53 | 5.27×10<sup>-4</sup> | 1.02×10<sup>-3</sup> | 0.120 | 0.0528 |
| Hashimoto's disease | -0.295 | 0.032 | 33.36 | 1.15×10<sup>-4</sup> | 2.54×10<sup>-4</sup> | 1.868 | 2.49×10<sup>-5</sup> |
| Grave's disease | 0.004 | 0.982 | 194.38 | 4.99×10<sup>-37</sup> | 8.23×10<sup>-36</sup> | 1.996 | 3.79×10<sup>-5</sup> |
| Rheumatoid arthritis | -0.102 | 0.567 | 390.46 | 1.49×10<sup>-78</sup> | 4.90×10<sup>-77</sup> | 1.802 | 3.09×10<sup>-4</sup> |
| SLE | 0.125 | 0.379 | 16.71 | 0.0535 | 0.0706 | 0.260 | 0.52 |
| Psoriasis | -0.050 | 0.808 | 70.33 | 1.31×10<sup>-11</sup> | 8.64×10<sup>-11</sup> | 0.688 | 0.228 |
| **Allergic disease** |  |  |  |  |  |  |  |
| Atopic dermatitis | 0.043 | 0.505 | 34.66 | 6.84×10<sup>-5</sup> | 1.61×10<sup>-4</sup> | -0.680 | 1.52×10<sup>-4</sup> |
| Asthma | -0.058 | 0.471 | 183.25 | 1.06×10<sup>-34</sup> | 1.17×10<sup>-33</sup> | 0.488 | 0.033 |
| Pediatric asthma | 0.019 | 0.89 | 26.12 | 1.96×10<sup>-3</sup> | 3.40×10<sup>-3</sup> | 0.307 | 0.429 |
| Pollinosis | -0.038 | 0.17 | 31.40 | 2.53×10<sup>-4</sup> | 5.21×10<sup>-4</sup> | 0.160 | 0.052 |
| Chronic sinusitis | -0.005 | 0.954 | 95.65 | 1.19×10<sup>-16</sup> | 9.79×10<sup>-16</sup> | 0.976 | 1.77×10<sup>-4</sup> |
| Allergic conjunctivitis | -0.002 | 0.943 | 3.65 | 0.933 | 0.933 | 0.035 | 0.52 |
| Urticaria | -0.030 | 0.179 | 12.90 | 0.167 | 0.386 | -0.151 | 0.023 |
| **Infection & Respiratory disease** |  |  |  |  |  |  |  |
| Pulmonary tuberculosis | -0.061 | 0.245 | 52.87 | 3.09×10<sup>-8</sup> | 1.28×10<sup>-7</sup> | 0.868 | 9.81×10<sup>-9</sup> |
| Chronic hepatitis B | 0.015 | 0.927 | 137.21 | 3.89×10<sup>-25</sup> | 1.17×10<sup>-23</sup> | 1.789 | 6.81×10<sup>-5</sup> |
| Chronic hepatitis C | 0.005 | 0.931 | 63.90 | 2.36×10<sup>-10</sup> | 3.54×10<sup>-9</sup> | -0.320 | 0.0617 |
| Zoster infection | 0.021 | 0.613 | 9.48 | 0.394 | 0.657 | -0.011 | 0.924 |
| COVID-19 | 0.220 | 0.0156 | 33.30 | 1.19×10<sup>-4</sup> | 8.89×10<sup>-4</sup> | -0.831 | 0.0113 |
| Pneumonia | -0.066 | 0.153 | 42.58 | 2.57×10<sup>-6</sup> | 7.72×10<sup>-6</sup> | 0.310 | 0.0237 |
| Interstitial lung disease | -0.015 | 0.788 | 7.99 | 0.535 | 0.76 | 0.634 | 4.45×10<sup>-5</sup> |
| **Cardiovascular & other disease** |  |  |  |  |  |  |  |
| Coronary artery disease | 0.019 | 0.577 | 64.34 | 1.94×10<sup>-10</sup> | 1.07×10<sup>-9</sup> | -0.255 | 8.93×10<sup>-3</sup> |
| Myocardial infarction | 0.014 | 0.73 | 45.08 | 8.93×10<sup>-7</sup> | 2.95×10<sup>-6</sup> | -0.418 | 1.82×10<sup>-4</sup> |
| Ischemic stroke | 0.022 | 0.152 | 11.94 | 0.217 | 0.464 | -0.145 | 1.57×10<sup>-3</sup> |
| Subarachnoid hemorrhage | -0.006 | 0.909 | 6.00 | 0.74 | 0.823 | 0.373 | 3.08×10<sup>-3</sup> |
| Parkinson disease | 0.262 | 0.0297 | 16.37 | 0.0596 | 0.0756 | -1.273 | 1.06×10<sup>-3</sup> |
| Ulcerative colitis | 0.046 | 0.863 | 56.14 | 7.39×10<sup>-9</sup> | 3.48×10<sup>-8</sup> | -1.902 | 0.0104 |
| Iron deficiency anemia | -0.022 | 0.421 | 3.77 | 0.926 | 0.933 | -0.070 | 0.174 |
| **Quantitative traits** |  |  |  |  |  |  |  |
| Height | 0.012 | 0.241 | 130.18 | 1.09×10<sup>-23</sup> | 1.09×10<sup>-22</sup> | -0.102 | 3.46×10<sup>-4</sup> |
| Body Weight | 0.024 | 0.0453 | 124.72 | 1.44×10<sup>-22</sup> | 9.61×10<sup>-22</sup> | -0.132 | 4.25×10<sup>-4</sup> |
| BMI | 0.022 | 0.0237 | 61.26 | 7.65×10<sup>-10</sup> | 1.70×10<sup>-9</sup> | -0.087 | 5.00×10<sup>-3</sup> |
| SBP: Systolic blood pressure | 0.006 | 0.261 | 14.88 | 0.0942 | 0.135 | -0.021 | 0.183 |
| DBP: Diastolic blood pressure | 0.003 | 0.534 | 11.83 | 0.223 | 0.251 | -0.015 | 0.305 |
| RBC count | 0.006 | 0.647 | 85.23 | 1.47×10<sup>-14</sup> | 5.87×10<sup>-14</sup> | -0.053 | 0.135 |
| WBC count | 0.004 | 0.834 | 145.26 | 8.43×10<sup>-27</sup> | 1.69×10<sup>-25</sup> | -0.041 | 0.415 |
| Eosinophil count | -0.006 | 0.776 | 118.53 | 2.68×10<sup>-21</sup> | 1.34×10<sup>-20</sup> | -0.161 | 7.29×10<sup>-3</sup> |
| Basophil count | 0.003 | 0.684 | 12.96 | 0.164 | 0.219 | -0.222 | 2.44×10<sup>-28</sup> |
| TC: Total cholesterol | 0.001 | 0.844 | 20.26 | 0.0163 | 0.0252 | -0.119 | 2.95×10<sup>-9</sup> |
| HDLC: HDL-cholesterol | -0.022 | 0.113 | 50.78 | 7.68×10<sup>-8</sup> | 1.40×10<sup>-7</sup> | 0.106 | 0.0116 |
| LDLC: LDL-cholesterol | -0.002 | 0.831 | 12.08 | 0.209 | 0.251 | -0.133 | 1.32×10<sup>-10</sup> |
| TG: Triglycerides | 0.007 | 0.646 | 69.75 | 1.70×10<sup>-11</sup> | 4.26×10<sup>-11</sup> | -0.087 | 0.0376 |
| HbA1c | -0.004 | 0.489 | 10.20 | 0.334 | 0.352 | 0.042 | 0.0118 |

|  | MR-cML |  |
| --- | --- | --- |
|  | Theta | P-value |
| Autoimmune disease & T2D |  |  |
| Type 1 diabetes | 1.421 | $1.09 \times 10^{-17}$ |
| Type 2 diabetes | 0.115 | $2.18 \times 10^{-3}$ |
| Hashimoto's disease | 2.051 | $9.74 \times 10^{-14}$ |
| Grave's disease | 2.223 | $2.56 \times 10^{-31}$ |
| Rheumatoid arthritis | 0.461 | $6.46 \times 10^{-4}$ |
| SLE | 0.257 | 0.395 |
| Psoriasis | 0.052 | 0.813 |
| Allergic disease |  |  |
| Atopic dermatitis | -0.568 | $1.11 \times 10^{-6}$ |
| Asthma | -0.123 | 0.114 |
| Pediatric asthma | 0.036 | 0.882 |
| Pollinosis | 0.191 | $7.91 \times 10^{-5}$ |
| Chronic sinusitis | 1.315 | $8.34 \times 10^{-30}$ |
| Allergic conjunctivitis | 0.036 | 0.68 |
| Urticaria | -0.157 | $6.24 \times 10^{-3}$ |
| Infection & Respiratory disease |  |  |
| Pulmonary tuberculosis | 0.979 | $1.12 \times 10^{-28}$ |
| Chronic hepatitis B | 1.085 | $2.61 \times 10^{-10}$ |
| Chronic hepatitis C | -0.464 | $1.15 \times 10^{-9}$ |
| Zoster infection | -0.010 | 0.932 |
| COVID-19 | -1.016 | $8.37 \times 10^{-9}$ |
| Pneumonia | 0.365 | $8.21 \times 10^{-7}$ |
| Interstitial lung disease | 0.642 | $1.29 \times 10^{-4}$ |
| Cardiovascular & other disease |  |  |
| Coronary artery disease | -0.272 | $6.07 \times 10^{-7}$ |
| Myocardial infarction | -0.533 | $1.65 \times 10^{-15}$ |
| Ischemic stroke | -0.151 | $3.00 \times 10^{-4}$ |
| Subarachnoid hemorrhage | 0.377 | 0.0157 |
| Parkinson disease | -1.305 | $1.09 \times 10^{-5}$ |
| Ulcerative colitis | -2.699 | $5.32 \times 10^{-12}$ |
| Iron deficiency anemia | -0.070 | 0.379 |
| Quantitative traits |  |  |
| Height | -0.125 | $1.57 \times 10^{-28}$ |
| Body Weight | -0.169 | $1.82 \times 10^{-37}$ |
| BMI | -0.105 | $8.09 \times 10^{-15}$ |
| SBP: Systolic blood pressure | -0.022 | 0.0831 |
| DBP: Diastolic blood pressure | -0.016 | 0.235 |
| RBC count | -0.004 | 0.826 |
| WBC count | 0.054 | $5.21 \times 10^{-4}$ |
| Eosinophil count | -0.146 | $6.44 \times 10^{-12}$ |
| Basophil count | -0.226 | $8.61 \times 10^{-33}$ |
| TC: Total cholesterol | -0.123 | $2.01 \times 10^{-16}$ |
| HDLC: HDL-cholesterol | 0.140 | $4.07 \times 10^{-10}$ |
| LDLC: LDL-cholesterol | -0.136 | $6.85 \times 10^{-13}$ |
| TG: Triglycerides | -0.105 | $2.51 \times 10^{-8}$ |
| HbA1c | 0.043 | $7.21 \times 10^{-3}$ |

Font colors: Red indicates a positive association, Blue indicates an inverse association.
Background colors: Red indicates robust positive association (full IV set analysis), Blue indicates robust inverse association (full IV set analysis).

**Supplementary Table 24.** MR stringent LD clumping (r² < 0.001 within a 10 Mb window).

|  | IVW (fixed effects) |  | Weighted median |  | Weighted mode |  | MR-Egger |  |
| --- | --- | --- | --- | --- | --- | --- | --- | --- |
|  | Beta | P-value | Beta | P-value | Beta | P-value | Beta | P-value |
| **Autoimmune disease & T2D** |  |  |  |  |  |  |  |  |
| Type 1 diabetes | 1.041 | $5.08 \times 10^{-8}$ | 1.133 | $3.00 \times 10^{-4}$ | 1.641 | $1.66 \times 10^{-3}$ | 2.144 | 0.078 |
| Type 2 diabetes | 0.143 | $7.65 \times 10^{-4}$ | 0.199 | $3.53 \times 10^{-4}$ | 0.182 | 0.020 | 0.010 | 0.967 |
| Hashimoto's disease | 0.814 | $4.15 \times 10^{-3}$ | 1.057 | 0.020 | 1.808 | $6.94 \times 10^{-3}$ | 3.672 | 0.060 |
| Grave's disease | 1.082 | $3.83 \times 10^{-17}$ | 0.954 | $1.59 \times 10^{-4}$ | 1.279 | $3.85 \times 10^{-3}$ | 2.687 | 0.107 |
| Rheumatoid arthritis | 1.717 | $7.15 \times 10^{-75}$ | 0.421 | 0.040 | 0.307 | 0.219 | 3.173 | 0.115 |
| SLE | 0.163 | 0.657 | 0.064 | 0.899 | 0.012 | 0.983 | -0.837 | 0.686 |
| Psoriasis | 0.354 | 0.158 | 0.407 | 0.264 | 0.223 | 0.568 | 2.085 | 0.225 |
| **Allergic disease** |  |  |  |  |  |  |  |  |
| Atopic dermatitis | -0.453 | $6.38 \times 10^{-5}$ | -0.561 | $1.66 \times 10^{-4}$ | -0.613 | $4.80 \times 10^{-3}$ | -0.926 | 0.128 |
| Asthma | 0.377 | $1.79 \times 10^{-9}$ | 0.312 | 0.069 | 0.796 | $5.90 \times 10^{-4}$ | 0.983 | 0.101 |
| Pediatric asthma | 0.002 | 0.995 | 0.392 | 0.296 | 0.422 | 0.328 | -0.065 | 0.948 |
| Pollinosis | 0.074 | 0.176 | 0.146 | 0.063 | 0.181 | 0.045 | 0.647 | 0.021 |
| Chronic sinusitis | 0.776 | $3.61 \times 10^{-15}$ | 0.362 | 0.146 | 1.522 | $8.61 \times 10^{-4}$ | 1.439 | 0.225 |
| Allergic conjunctivitis | 0.075 | 0.483 | 0.032 | 0.813 | 0.018 | 0.918 | 0.122 | 0.705 |
| Urticaria | -0.094 | 0.173 | -0.181 | 0.056 | -0.193 | 0.135 | 0.170 | 0.573 |
| **Infection & Respiratory disease** |  |  |  |  |  |  |  |  |
| Pulmonary tuberculosis | 0.653 | $2.85 \times 10^{-17}$ | 0.679 | $4.83 \times 10^{-7}$ | 0.845 | $8.06 \times 10^{-4}$ | 1.379 | 0.026 |
| Chronic hepatitis B | 1.011 | $1.05 \times 10^{-12}$ | 0.826 | $8.15 \times 10^{-5}$ | 0.845 | 0.012 | 2.375 | 0.047 |
| Chronic hepatitis C | -0.185 | 0.020 | -0.391 | $1.34 \times 10^{-3}$ | -0.440 | 0.015 | -0.230 | 0.747 |
| Zoster infection | -0.058 | 0.680 | 0.079 | 0.661 | 0.113 | 0.620 | 0.169 | 0.690 |
| COVID-19 | -0.584 | $3.76 \times 10^{-3}$ | -0.915 | $4.37 \times 10^{-4}$ | -0.950 | $9.96 \times 10^{-3}$ | -2.925 | $6.06 \times 10^{-3}$ |
| Pneumonia | 0.250 | $1.36 \times 10^{-3}$ | 0.320 | $9.90 \times 10^{-3}$ | 0.575 | $3.60 \times 10^{-3}$ | 0.980 | 0.057 |
| Interstitial lung disease | 0.520 | 0.011 | 0.543 | 0.039 | 0.491 | 0.166 | 1.164 | 0.105 |
| **Cardiovascular & other disease** |  |  |  |  |  |  |  |  |
| Coronary artery disease | -0.143 | $1.55 \times 10^{-3}$ | -0.197 | $4.22 \times 10^{-3}$ | -0.276 | $6.22 \times 10^{-3}$ | -0.658 | $7.14 \times 10^{-3}$ |
| Myocardial infarction | -0.238 | $1.21 \times 10^{-4}$ | -0.313 | $6.97 \times 10^{-4}$ | -0.469 | $2.01 \times 10^{-3}$ | -0.843 | 0.021 |
| Ischemic stroke | -0.150 | $2.32 \times 10^{-3}$ | -0.171 | $6.87 \times 10^{-3}$ | -0.176 | 0.048 | -0.282 | 0.094 |
| Subarachnoid hemorrhage | 0.328 | 0.085 | 0.480 | 0.036 | 0.516 | 0.089 | 0.750 | 0.222 |
| Parkinson disease | -1.049 | $3.25 \times 10^{-3}$ | -1.158 | 0.019 | -1.149 | 0.074 | -2.767 | 0.046 |
| Ulcerative colitis | -1.405 | $1.24 \times 10^{-4}$ | -1.762 | $2.12 \times 10^{-3}$ | -2.309 | 0.013 | -1.813 | 0.470 |
| Iron deficiency anemia | -0.064 | 0.515 | -0.098 | 0.437 | -0.097 | 0.549 | 0.182 | 0.545 |
| **Quantitative traits** |  |  |  |  |  |  |  |  |
| Height | -0.049 | $1.57 \times 10^{-7}$ | -0.058 | $6.39 \times 10^{-4}$ | -0.075 | $9.43 \times 10^{-3}$ | -0.217 | 0.040 |
| Body Weight | -0.103 | $1.35 \times 10^{-16}$ | -0.132 | $3.29 \times 10^{-6}$ | -0.153 | $4.34 \times 10^{-4}$ | -0.356 | 0.013 |
| BMI | -0.091 | $5.33 \times 10^{-10}$ | -0.122 | $3.17 \times 10^{-7}$ | -0.132 | $5.77 \times 10^{-4}$ | -0.278 | 0.022 |
| SBP: Systolic blood pressure | -0.016 | 0.299 | -0.022 | 0.257 | -0.023 | 0.311 | -0.096 | 0.078 |
| DBP: Diastolic blood pressure | 0.011 | 0.497 | 0.006 | 0.764 | 0.006 | 0.804 | -0.049 | 0.334 |
| RBC count | -0.044 | $2.16 \times 10^{-3}$ | 0.006 | 0.791 | 0.011 | 0.590 | -0.048 | 0.750 |
| WBC count | 0.025 | 0.112 | 0.072 | $6.80 \times 10^{-4}$ | 0.074 | 0.012 | -0.040 | 0.766 |
| Eosinophil count | -0.101 | $7.10 \times 10^{-7}$ | -0.120 | $6.93 \times 10^{-5}$ | -0.147 | $3.43 \times 10^{-3}$ | -0.270 | 0.031 |
| Basophil count | -0.151 | $3.66 \times 10^{-13}$ | -0.165 | $4.84 \times 10^{-7}$ | -0.192 | $1.97 \times 10^{-3}$ | -0.307 | 0.022 |
| TC: Total cholesterol | -0.084 | $3.70 \times 10^{-7}$ | -0.101 | $5.61 \times 10^{-5}$ | -0.112 | $4.01 \times 10^{-3}$ | -0.199 | 0.037 |
| HDLc: HDL-cholesterol | 0.111 | $3.04 \times 10^{-7}$ | 0.142 | $3.07 \times 10^{-5}$ | 0.174 | $3.08 \times 10^{-3}$ | 0.290 | 0.026 |
| LDLC: LDL-cholesterol | -0.090 | $4.46 \times 10^{-5}$ | -0.120 | $6.32 \times 10^{-5}$ | -0.125 | $6.52 \times 10^{-3}$ | -0.170 | 0.066 |
| TG: Triglycerides | -0.100 | $6.80 \times 10^{-8}$ | -0.125 | $1.01 \times 10^{-6}$ | -0.128 | $1.94 \times 10^{-3}$ | -0.213 | 0.018 |
| HbA1c | 0.034 | 0.084 | 0.048 | 0.057 | 0.049 | 0.102 | 0.079 | 0.207 |

|  | pleiotropy test |  | heterogeneity test |  |  | IVW<br>(multiplicative random effects) |  |
| --- | --- | --- | --- | --- | --- | --- | --- |
|  | intercept | P-value | Cochran's Q | P-value | FDR Q-value | Beta | P-value |
| **Autoimmune disease & T2D** |  |  |  |  |  |  |  |
| Type 1 diabetes | -0.107 | 0.288 | 24.72 | $8.49 \times 10^{-4}$ | $1.62 \times 10^{-3}$ | 1.041 | $3.74 \times 10^{-3}$ |
| Type 2 diabetes | 0.013 | 0.562 | 22.77 | $1.87 \times 10^{-3}$ | $3.14 \times 10^{-3}$ | 0.143 | 0.062 |
| Hashimoto's disease | -0.278 | 0.103 | 36.52 | $5.79 \times 10^{-6}$ | $1.87 \times 10^{-5}$ | 0.814 | 0.209 |
| Grave's disease | -0.156 | 0.273 | 108.85 | $1.59 \times 10^{-20}$ | $3.34 \times 10^{-19}$ | 1.082 | 0.033 |
| Rheumatoid arthritis | -0.142 | 0.401 | 277.61 | $3.62 \times 10^{-56}$ | $1.52 \times 10^{-54}$ | 1.717 | $3.65 \times 10^{-3}$ |
| SLE | 0.098 | 0.608 | 21.88 | $2.66 \times 10^{-3}$ | $4.30 \times 10^{-3}$ | 0.163 | 0.801 |
| Psoriasis | -0.166 | 0.276 | 34.07 | $1.67 \times 10^{-5}$ | $5.01 \times 10^{-5}$ | 0.354 | 0.523 |
| **Allergic disease** |  |  |  |  |  |  |  |
| Atopic dermatitis | 0.046 | 0.374 | 17.94 | 0.012 | 0.018 | -0.453 | 0.013 |
| Asthma | -0.059 | 0.250 | 60.08 | $1.46 \times 10^{-10}$ | $8.76 \times 10^{-10}$ | 0.377 | 0.040 |
| Pediatric asthma | 0.007 | 0.943 | 8.37 | 0.301 | 0.372 | 0.002 | 0.995 |
| Pollinosis | -0.056 | 0.027 | 25.77 | $5.53 \times 10^{-4}$ | $1.16 \times 10^{-3}$ | 0.074 | 0.480 |
| Chronic sinusitis | -0.065 | 0.531 | 90.72 | $8.79 \times 10^{-17}$ | $7.38 \times 10^{-16}$ | 0.776 | 0.029 |
| Allergic conjunctivitis | -0.005 | 0.876 | 3.90 | 0.791 | 0.811 | 0.075 | 0.347 |
| Urticaria | -0.026 | 0.362 | 14.52 | 0.043 | 0.062 | -0.094 | 0.345 |
| **Infection & Respiratory disease** |  |  |  |  |  |  |  |
| Pulmonary tuberculosis | -0.071 | 0.148 | 38.72 | $2.21 \times 10^{-6}$ | $7.74 \times 10^{-6}$ | 0.653 | $3.26 \times 10^{-4}$ |
| Chronic hepatitis B | -0.132 | 0.179 | 45.07 | $1.33 \times 10^{-7}$ | $5.08 \times 10^{-7}$ | 1.011 | $4.99 \times 10^{-3}$ |
| Chronic hepatitis C | 0.004 | 0.946 | 52.89 | $3.90 \times 10^{-9}$ | $2.05 \times 10^{-8}$ | -0.185 | 0.399 |
| Zoster infection | -0.022 | 0.571 | 4.82 | 0.682 | 0.716 | -0.058 | 0.619 |
| COVID-19 | 0.212 | 0.012 | 27.63 | $2.56 \times 10^{-4}$ | $6.07 \times 10^{-4}$ | -0.584 | 0.145 |
| Pneumonia | -0.071 | 0.110 | 32.74 | $2.96 \times 10^{-5}$ | $8.29 \times 10^{-5}$ | 0.250 | 0.139 |
| Interstitial lung disease | -0.063 | 0.303 | 7.83 | 0.348 | 0.418 | 0.520 | 0.016 |
| **Cardiovascular & other disease** |  |  |  |  |  |  |  |
| Coronary artery disease | 0.050 | 0.016 | 27.59 | $2.60 \times 10^{-4}$ | $6.07 \times 10^{-4}$ | -0.143 | 0.111 |
| Myocardial infarction | 0.059 | 0.054 | 27.11 | $3.18 \times 10^{-4}$ | $7.03 \times 10^{-4}$ | -0.238 | 0.051 |
| Ischemic stroke | 0.013 | 0.358 | 2.48 | 0.928 | 0.928 | -0.150 | $3.17 \times 10^{-7}$ |
| Subarachnoid hemorrhage | -0.041 | 0.445 | 5.06 | 0.652 | 0.716 | 0.328 | 0.043 |
| Parkinson disease | 0.167 | 0.148 | 10.14 | 0.181 | 0.237 | -1.049 | 0.015 |
| Ulcerative colitis | 0.040 | 0.859 | 30.14 | $8.95 \times 10^{-5}$ | $2.35 \times 10^{-4}$ | -1.405 | 0.064 |
| Iron deficiency anemia | -0.024 | 0.391 | 6.52 | 0.480 | 0.560 | -0.064 | 0.500 |
| **Quantitative traits** |  |  |  |  |  |  |  |
| Height | 0.016 | 0.074 | 103.19 | $2.36 \times 10^{-19}$ | $2.48 \times 10^{-18}$ | -0.049 | 0.172 |
| Body Weight | 0.025 | 0.038 | 104.02 | $1.59 \times 10^{-19}$ | $2.23 \times 10^{-18}$ | -0.103 | 0.032 |
| BMI | 0.018 | 0.069 | 49.59 | $1.74 \times 10^{-8}$ | $7.31 \times 10^{-8}$ | -0.091 | 0.020 |
| SBP: Systolic blood pressure | 0.008 | 0.108 | 9.92 | 0.193 | 0.246 | -0.016 | 0.383 |
| DBP: Diastolic blood pressure | 0.006 | 0.219 | 4.90 | 0.672 | 0.716 | 0.011 | 0.417 |
| RBC count | 0.000 | 0.979 | 72.24 | $5.20 \times 10^{-13}$ | $3.64 \times 10^{-12}$ | -0.044 | 0.340 |
| WBC count | 0.006 | 0.609 | 51.14 | $8.62 \times 10^{-9}$ | $4.02 \times 10^{-8}$ | 0.025 | 0.556 |
| Eosinophil count | 0.017 | 0.111 | 25.62 | $5.88 \times 10^{-4}$ | $1.18 \times 10^{-3}$ | -0.101 | $9.55 \times 10^{-3}$ |
| Basophil count | 0.015 | 0.146 | 24.55 | $9.10 \times 10^{-4}$ | $1.66 \times 10^{-3}$ | -0.151 | $1.04 \times 10^{-4}$ |
| TC: Total cholesterol | 0.011 | 0.152 | 21.49 | $3.11 \times 10^{-3}$ | $4.85 \times 10^{-3}$ | -0.084 | $3.71 \times 10^{-3}$ |
| HDLC: HDL-cholesterol | -0.017 | 0.101 | 24.04 | $1.12 \times 10^{-3}$ | $1.96 \times 10^{-3}$ | 0.111 | $5.72 \times 10^{-3}$ |
| LDLC: LDL-cholesterol | 0.008 | 0.304 | 10.38 | 0.168 | 0.227 | -0.090 | $8.04 \times 10^{-4}$ |
| TG: Triglycerides | 0.011 | 0.119 | 14.31 | 0.046 | 0.064 | -0.100 | $1.61 \times 10^{-4}$ |
| HbA1c | -0.004 | 0.418 | 5.93 | 0.548 | 0.622 | 0.034 | 0.060 |

|  | MR-cML |  |
| --- | --- | --- |
|  | Theta | P-value |
| Autoimmune disease & T2D |  |  |
| Type 1 diabetes | 1.197 | $8.98 \times 10^{-7}$ |
| Type 2 diabetes | 0.136 | $4.00 \times 10^{-3}$ |
| Hashimoto's disease | 0.776 | 0.210 |
| Grave's disease | 0.291 | 0.595 |
| Rheumatoid arthritis | 0.436 | $9.63 \times 10^{-3}$ |
| SLE | 0.303 | 0.454 |
| Psoriasis | 0.067 | 0.806 |
| Allergic disease |  |  |
| Atopic dermatitis | -0.483 | $7.56 \times 10^{-5}$ |
| Asthma | -0.127 | 0.176 |
| Pediatric asthma | 0.019 | 0.949 |
| Pollinosis | 0.099 | 0.105 |
| Chronic sinusitis | -0.060 | 0.765 |
| Allergic conjunctivitis | 0.076 | 0.481 |
| Urticaria | -0.100 | 0.163 |
| Infection & Respiratory disease |  |  |
| Pulmonary tuberculosis | 0.770 | $5.81 \times 10^{-12}$ |
| Chronic hepatitis B | 0.780 | $6.45 \times 10^{-7}$ |
| Chronic hepatitis C | -0.413 | $5.11 \times 10^{-4}$ |
| Zoster infection | -0.058 | 0.680 |
| COVID-19 | -0.845 | $1.20 \times 10^{-4}$ |
| Pneumonia | 0.067 | 0.793 |
| Interstitial lung disease | 0.526 | 0.011 |
| Cardiovascular & other disease |  |  |
| Coronary artery disease | -0.155 | $1.79 \times 10^{-3}$ |
| Myocardial infarction | -0.248 | $1.74 \times 10^{-3}$ |
| Ischemic stroke | -0.151 | $2.48 \times 10^{-3}$ |
| Subarachnoid hemorrhage | 0.332 | 0.084 |
| Parkinson disease | -1.062 | $3.70 \times 10^{-3}$ |
| Ulcerative colitis | -1.660 | $5.79 \times 10^{-3}$ |
| Iron deficiency anemia | -0.068 | 0.500 |
| Quantitative traits |  |  |
| Height | 0.056 | $8.90 \times 10^{-3}$ |
| Body Weight | -0.157 | $2.62 \times 10^{-14}$ |
| BMI | -0.118 | $1.02 \times 10^{-11}$ |
| SBP: Systolic blood pressure | -0.016 | 0.291 |
| DBP: Diastolic blood pressure | 0.011 | 0.498 |
| RBC count | -0.006 | 0.697 |
| WBC count | 0.062 | $2.40 \times 10^{-4}$ |
| Eosinophil count | -0.105 | $8.74 \times 10^{-6}$ |
| Basophil count | -0.158 | $3.89 \times 10^{-12}$ |
| TC: Total cholesterol | -0.090 | $5.87 \times 10^{-7}$ |
| HDLC: HDL-cholesterol | 0.118 | $4.63 \times 10^{-7}$ |
| LDLC: LDL-cholesterol | -0.092 | $5.04 \times 10^{-5}$ |
| TG: Triglycerides | -0.103 | $1.28 \times 10^{-7}$ |
| HbA1c | 0.034 | 0.081 |

Font colors: Red indicates a positive association, Blue indicates an inverse association.

**Supplementary Table 25.** MR analysis using representative MHC class II signal variants.

|  | IVW (fixed effects) |  | Weighted median |  | Weighted mode |  | MR-Egger |  |
| --- | --- | --- | --- | --- | --- | --- | --- | --- |
|  | Beta | P-value | Beta | P-value | Beta | P-value | Beta | P-value |
| **Autoimmune disease & T2D** |  |  |  |  |  |  |  |  |
| Type 1 diabetes | 0.892 | 3.66×10<sup>-5</sup> | 1.027 | 1.08×10<sup>-3</sup> | 1.564 | 7.55×10<sup>-3</sup> | 2.376 | 0.165 |
| Type 2 diabetes | 0.139 | 3.71×10<sup>-3</sup> | 0.133 | 0.025 | 0.124 | 0.113 | -0.146 | 0.617 |
| Hashimoto's disease | 1.016 | 1.59×10<sup>-3</sup> | 1.075 | 0.013 | 2.347 | 9.21×10<sup>-3</sup> | 4.437 | 0.093 |
| Grave's disease | 1.244 | 9.35×10<sup>-18</sup> | 0.861 | 6.30×10<sup>-3</sup> | 0.066 | 0.923 | 2.977 | 0.133 |
| Rheumatoid arthritis | 1.479 | 2.69×10<sup>-44</sup> | 0.742 | 1.01×10<sup>-3</sup> | 0.35 | 0.346 | 3.416 | 0.127 |
| SLE | 0.304 | 0.465 | 0.412 | 0.446 | 0.329 | 0.631 | -2.294 | 0.232 |
| Psoriasis | 0.724 | 0.011 | 0.268 | 0.565 | 0.343 | 0.646 | 2.836 | 0.217 |
| **Allergic disease** |  |  |  |  |  |  |  |  |
| Atopic dermatitis | -0.706 | 3.98×10<sup>-8</sup> | -0.64 | 2.66×10<sup>-3</sup> | -0.616 | 0.147 | -1.811 | 0.153 |
| Asthma | 0.236 | 8.66×10<sup>-4</sup> | 0.133 | 0.21 | 0.115 | 0.431 | 0.927 | 0.121 |
| Pediatric asthma | -0.255 | 0.424 | -0.173 | 0.694 | -0.017 | 0.976 | -1.164 | 0.436 |
| Pollinosis | -0.03 | 0.631 | -0.012 | 0.881 | 0.037 | 0.681 | 0.518 | 0.098 |
| Chronic sinusitis | 0.754 | 1.52×10<sup>-11</sup> | 0.542 | 1.69×10<sup>-3</sup> | 0.507 | 0.112 | 1.857 | 0.14 |
| Allergic conjunctivitis | 0.003 | 0.98 | 0.104 | 0.547 | 0.26 | 0.419 | 0.109 | 0.846 |
| Urticaria | -0.139 | 0.074 | -0.183 | 0.078 | -0.196 | 0.181 | 0.1 | 0.82 |
| **Infection & Respiratory disease** |  |  |  |  |  |  |  |  |
| Pulmonary tuberculosis | 0.598 | 7.99×10<sup>-12</sup> | 0.697 | 7.12×10<sup>-7</sup> | 0.897 | 1.28×10<sup>-3</sup> | 1.298 | 0.121 |
| Chronic hepatitis B | 0.557 | 4.92×10<sup>-4</sup> | 0.528 | 0.019 | 0.602 | 0.108 | 1.302 | 0.113 |
| Chronic hepatitis C | -0.181 | 0.045 | -0.029 | 0.839 | 0.093 | 0.611 | -0.55 | 0.56 |
| Zoster infection | -0.21 | 0.187 | -0.106 | 0.604 | 0.017 | 0.954 | -0.184 | 0.748 |
| COVID-19 | -0.307 | 0.169 | -0.757 | 5.67×10<sup>-3</sup> | -0.769 | 0.046 | -2.921 | 0.026 |
| Pneumonia | 0.226 | 0.01 | 0.222 | 0.078 | 0.216 | 0.255 | 1.324 | 0.041 |
| Interstitial lung disease | 0.406 | 0.078 | 0.54 | 0.067 | 0.504 | 0.167 | 0.833 | 0.373 |
| **Cardiovascular & other disease** |  |  |  |  |  |  |  |  |
| Coronary artery disease | 0.01 | 0.848 | 0.089 | 0.234 | 0.096 | 0.37 | -0.364 | 0.295 |
| Myocardial infarction | -0.068 | 0.33 | 0.029 | 0.776 | 0.063 | 0.655 | -0.505 | 0.33 |
| Ischemic stroke | -0.137 | 0.014 | -0.16 | 0.018 | -0.177 | 0.096 | -0.272 | 0.22 |
| Subarachnoid hemorrhage | 0.407 | 0.06 | 0.5 | 0.055 | 0.539 | 0.134 | 1.4 | 0.125 |
| Parkinson disease | -0.985 | 0.015 | -1.329 | 0.016 | -1.565 | 0.071 | -2.846 | 0.118 |
| Ulcerative colitis | -2.129 | 2.94×10<sup>-7</sup> | -2.235 | 3.74×10<sup>-4</sup> | -2.974 | 0.012 | -5.059 | 0.123 |
| Iron deficiency anemia | -0.001 | 0.996 | -0.044 | 0.754 | -0.028 | 0.876 | 0.547 | 0.225 |
| **Quantitative traits** |  |  |  |  |  |  |  |  |
| Height | -0.025 | 0.016 | -0.03 | 0.072 | -0.021 | 0.401 | -0.217 | 0.148 |
| Body Weight | -0.083 | 4.55×10<sup>-9</sup> | -0.102 | 2.04×10<sup>-5</sup> | -0.125 | 2.80×10<sup>-3</sup> | -0.43 | 0.031 |
| BMI | -0.085 | 2.90×10<sup>-7</sup> | -0.126 | 8.66×10<sup>-8</sup> | -0.131 | 1.95×10<sup>-3</sup> | -0.375 | 0.018 |
| SBP: Systolic blood pressure | 0.001 | 0.942 | 0 | 0.99 | 0.009 | 0.762 | -0.042 | 0.544 |
| DBP: Diastolic blood pressure | 0.013 | 0.481 | 0.01 | 0.664 | 0.004 | 0.88 | -0.044 | 0.508 |
| RBC count | -0.066 | 4.98×10<sup>-5</sup> | -0.049 | 0.136 | -0.001 | 0.984 | -0.042 | 0.783 |
| WBC count | -0.007 | 0.688 | 0.049 | 0.077 | 0.061 | 0.117 | -0.11 | 0.506 |
| Eosinophil count | -0.095 | 3.96×10<sup>-5</sup> | -0.071 | 0.034 | -0.064 | 0.199 | -0.295 | 0.147 |
| Basophil count | -0.139 | 3.56×10<sup>-9</sup> | -0.151 | 9.11×10<sup>-6</sup> | -0.187 | 5.66×10<sup>-3</sup> | -0.326 | 0.07 |
| TC: Total cholesterol | -0.059 | 1.44×10<sup>-3</sup> | -0.046 | 0.074 | -0.039 | 0.349 | -0.122 | 0.161 |
| HDLC: HDL-cholesterol | 0.073 | 2.97×10<sup>-3</sup> | 0.083 | 0.019 | 0.112 | 0.072 | 0.271 | 0.086 |
| LDLC: LDL-cholesterol | -0.075 | 2.68×10<sup>-3</sup> | -0.068 | 0.04 | -0.064 | 0.166 | -0.127 | 0.21 |
| TG: Triglycerides | -0.046 | 0.03 | -0.036 | 0.285 | -0.099 | 0.251 | -0.08 | 0.615 |
| HbA1c | 0.055 | 0.013 | 0.053 | 0.045 | 0.057 | 0.138 | 0.14 | 0.131 |

|  | pleiotropy test |  | heterogeneity test |  |  | IVW<br>(multiplicative random effects) |  |
| --- | --- | --- | --- | --- | --- | --- | --- |
|  | intercept | P-value | Cochran's Q | P-value | FDR Q-value | Beta | P-value |
| **Autoimmune disease & T2D** |  |  |  |  |  |  |  |
| Type 1 diabetes | -0.131 | 0.33 | 19.553 | $1.52 \times 10^{-3}$ | $3.54 \times 10^{-3}$ | 0.892 | 0.037 |
| Type 2 diabetes | 0.025 | 0.331 | 14.796 | 0.011 | 0.02 | 0.139 | 0.092 |
| Hashimoto's disease | -0.301 | 0.15 | 25.096 | $1.34 \times 10^{-4}$ | $4.04 \times 10^{-4}$ | 1.016 | 0.159 |
| Grave's disease | -0.153 | 0.315 | 56.424 | $6.65 \times 10^{-11}$ | $6.98 \times 10^{-10}$ | 1.244 | 0.011 |
| Rheumatoid arthritis | -0.171 | 0.317 | 133.198 | $4.98 \times 10^{-27}$ | $2.09 \times 10^{-25}$ | 1.479 | $6.83 \times 10^{-3}$ |
| SLE | 0.229 | 0.171 | 9.282 | 0.098 | 0.147 | 0.304 | 0.592 |
| Psoriasis | -0.183 | 0.316 | 22.364 | $4.46 \times 10^{-4}$ | $1.17 \times 10^{-3}$ | 0.724 | 0.231 |
| **Allergic disease** |  |  |  |  |  |  |  |
| Atopic dermatitis | 0.097 | 0.323 | 29.907 | $1.54 \times 10^{-5}$ | $8.08 \times 10^{-5}$ | -0.706 | 0.025 |
| Asthma | -0.061 | 0.2 | 25.076 | $1.35 \times 10^{-4}$ | $4.04 \times 10^{-4}$ | 0.236 | 0.137 |
| Pediatric asthma | 0.08 | 0.518 | 7.09 | 0.214 | 0.3 | -0.255 | 0.502 |
| Pollinosis | -0.048 | 0.076 | 13.031 | 0.023 | 0.037 | -0.03 | 0.766 |
| Chronic sinusitis | -0.097 | 0.317 | 38.727 | $2.69 \times 10^{-7}$ | $1.89 \times 10^{-6}$ | 0.754 | 0.015 |
| Allergic conjunctivitis | -0.009 | 0.843 | 6.764 | 0.239 | 0.324 | 0.003 | 0.983 |
| Urticaria | -0.021 | 0.576 | 10.893 | 0.054 | 0.083 | -0.139 | 0.227 |
| **Infection & Respiratory disease** |  |  |  |  |  |  |  |
| Pulmonary tuberculosis | -0.062 | 0.329 | 26.546 | $6.99 \times 10^{-5}$ | $2.67 \times 10^{-4}$ | 0.598 | $3.00 \times 10^{-3}$ |
| Chronic hepatitis B | -0.066 | 0.292 | 7.903 | 0.162 | 0.234 | 0.557 | $5.57 \times 10^{-3}$ |
| Chronic hepatitis C | 0.033 | 0.678 | 34.463 | $1.93 \times 10^{-6}$ | $1.16 \times 10^{-5}$ | -0.181 | 0.446 |
| Zoster infection | -0.002 | 0.961 | 3.009 | 0.699 | 0.752 | -0.21 | 0.089 |
| COVID-19 | 0.219 | 0.032 | 20.11 | $1.19 \times 10^{-3}$ | $2.94 \times 10^{-3}$ | -0.307 | 0.493 |
| Pneumonia | -0.097 | 0.061 | 24.172 | $2.01 \times 10^{-4}$ | $5.63 \times 10^{-4}$ | 0.226 | 0.244 |
| Interstitial lung disease | -0.038 | 0.62 | 4.953 | 0.422 | 0.492 | 0.406 | 0.077 |
| **Cardiovascular & other disease** |  |  |  |  |  |  |  |
| Coronary artery disease | 0.033 | 0.265 | 17.661 | $3.40 \times 10^{-3}$ | $6.50 \times 10^{-3}$ | 0.01 | 0.919 |
| Myocardial infarction | 0.039 | 0.372 | 18.921 | $1.99 \times 10^{-3}$ | $4.37 \times 10^{-3}$ | -0.068 | 0.617 |
| Ischemic stroke | 0.012 | 0.494 | 2.046 | 0.843 | 0.853 | -0.137 | $1.19 \times 10^{-4}$ |
| Subarachnoid hemorrhage | -0.087 | 0.224 | 3.423 | 0.635 | 0.702 | 0.407 | 0.023 |
| Parkinson disease | 0.164 | 0.246 | 6.584 | 0.253 | 0.333 | -0.985 | 0.033 |
| Ulcerative colitis | 0.258 | 0.302 | 18.816 | $2.08 \times 10^{-3}$ | $4.37 \times 10^{-3}$ | -2.129 | $8.22 \times 10^{-3}$ |
| Iron deficiency anemia | -0.048 | 0.208 | 6.483 | 0.262 | 0.334 | -0.001 | 0.997 |
| **Quantitative traits** |  |  |  |  |  |  |  |
| Height | 0.017 | 0.172 | 79.858 | $8.98 \times 10^{-16}$ | $1.26 \times 10^{-14}$ | -0.025 | 0.547 |
| Body Weight | 0.031 | 0.051 | 89.628 | $8.04 \times 10^{-18}$ | $1.69 \times 10^{-16}$ | -0.083 | 0.166 |
| BMI | 0.026 | 0.035 | 41.65 | $6.93 \times 10^{-8}$ | $5.82 \times 10^{-7}$ | -0.085 | 0.076 |
| SBP: Systolic blood pressure | 0.004 | 0.514 | 5.225 | 0.389 | 0.467 | 0.001 | 0.943 |
| DBP: Diastolic blood pressure | 0.005 | 0.382 | 2.827 | 0.727 | 0.763 | 0.013 | 0.348 |
| RBC count | -0.002 | 0.865 | 26.995 | $5.72 \times 10^{-5}$ | $2.40 \times 10^{-4}$ | -0.066 | 0.081 |
| WBC count | 0.009 | 0.514 | 28.979 | $2.34 \times 10^{-5}$ | $1.09 \times 10^{-4}$ | -0.007 | 0.867 |
| Eosinophil count | 0.018 | 0.272 | 25.109 | $1.33 \times 10^{-4}$ | $4.04 \times 10^{-4}$ | -0.095 | 0.067 |
| Basophil count | 0.016 | 0.215 | 17.533 | $3.59 \times 10^{-3}$ | $6.56 \times 10^{-3}$ | -0.139 | $1.62 \times 10^{-3}$ |
| TC: Total cholesterol | 0.005 | 0.41 | 6.212 | 0.286 | 0.353 | -0.059 | $4.25 \times 10^{-3}$ |
| HDLC: HDL-cholesterol | -0.017 | 0.159 | 14.719 | 0.012 | 0.02 | 0.073 | 0.083 |
| LDLC: LDL-cholesterol | 0.005 | 0.561 | 4.503 | 0.48 | 0.544 | -0.075 | $1.56 \times 10^{-3}$ |
| TG: Triglycerides | 0.003 | 0.818 | 17.751 | $3.27 \times 10^{-3}$ | $6.50 \times 10^{-3}$ | -0.046 | 0.249 |
| HbA1c | -0.008 | 0.292 | 1.969 | 0.853 | 0.853 | 0.055 | $7.58 \times 10^{-5}$ |

|  | MR-cML |  |
| --- | --- | --- |
|  | Theta | P-value |
| Autoimmune disease & T2D |  |  |
| Type 1 diabetes | 0.976 | 9.56×10<sup>-4</sup> |
| Type 2 diabetes | 0.126 | 0.022 |
| Hashimoto's disease | 0.024 | 0.984 |
| Grave's disease | 0.014 | 0.954 |
| Rheumatoid arthritis | 0.476 | 0.075 |
| SLE | 0.293 | 0.493 |
| Psoriasis | 0.316 | 0.514 |
| Allergic disease |  |  |
| Atopic dermatitis | -0.425 | 0.033 |
| Asthma | 0.033 | 0.708 |
| Pediatric asthma | -0.25 | 0.445 |
| Pollinosis | -0.022 | 0.743 |
| Chronic sinusitis | 0.362 | 0.017 |
| Allergic conjunctivitis | 0.007 | 0.954 |
| Urticaria | -0.153 | 0.07 |
| Infection & Respiratory disease |  |  |
| Pulmonary tuberculosis | 0.691 | 1.17×10<sup>-5</sup> |
| Chronic hepatitis B | 0.558 | 9.57×10<sup>-4</sup> |
| Chronic hepatitis C | 0.112 | 0.305 |
| Zoster infection | -0.211 | 0.189 |
| COVID-19 | -0.571 | 0.022 |
| Pneumonia | 0.095 | 0.501 |
| Interstitial lung disease | 0.411 | 0.078 |
| Cardiovascular & other disease |  |  |
| Coronary artery disease | 0.12 | 0.093 |
| Myocardial infarction | 0.103 | 0.262 |
| Ischemic stroke | -0.138 | 0.014 |
| Subarachnoid hemorrhage | 0.411 | 0.059 |
| Parkinson disease | -0.996 | 0.016 |
| Ulcerative colitis | -2.032 | 3.19×10<sup>-4</sup> |
| Iron deficiency anemia | -0.003 | 0.981 |
| Quantitative traits |  |  |
| Height | 0.001 | 0.954 |
| Body Weight | -0.147 | 2.55×10<sup>-11</sup> |
| BMI | -0.117 | 3.46×10<sup>-9</sup> |
| SBP: Systolic blood pressure | 0.001 | 0.948 |
| DBP: Diastolic blood pressure | 0.013 | 0.482 |
| RBC count | -0.099 | 2.29×10<sup>-4</sup> |
| WBC count | 0.056 | 9.94×10<sup>-3</sup> |
| Eosinophil count | -0.036 | 0.234 |
| Basophil count | -0.146 | 1.21×10<sup>-8</sup> |
| TC: Total cholesterol | -0.059 | 2.38×10<sup>-3</sup> |
| HDLC: HDL-cholesterol | 0.07 | 0.016 |
| LDLC: LDL-cholesterol | -0.076 | 3.07×10<sup>-3</sup> |
| TG: Triglycerides | -0.053 | 0.036 |
| HbA1c | 0.055 | 0.014 |

Font colors: Red indicates a positive association, Blue indicates an inverse association.

**Supplementary Table 26.**
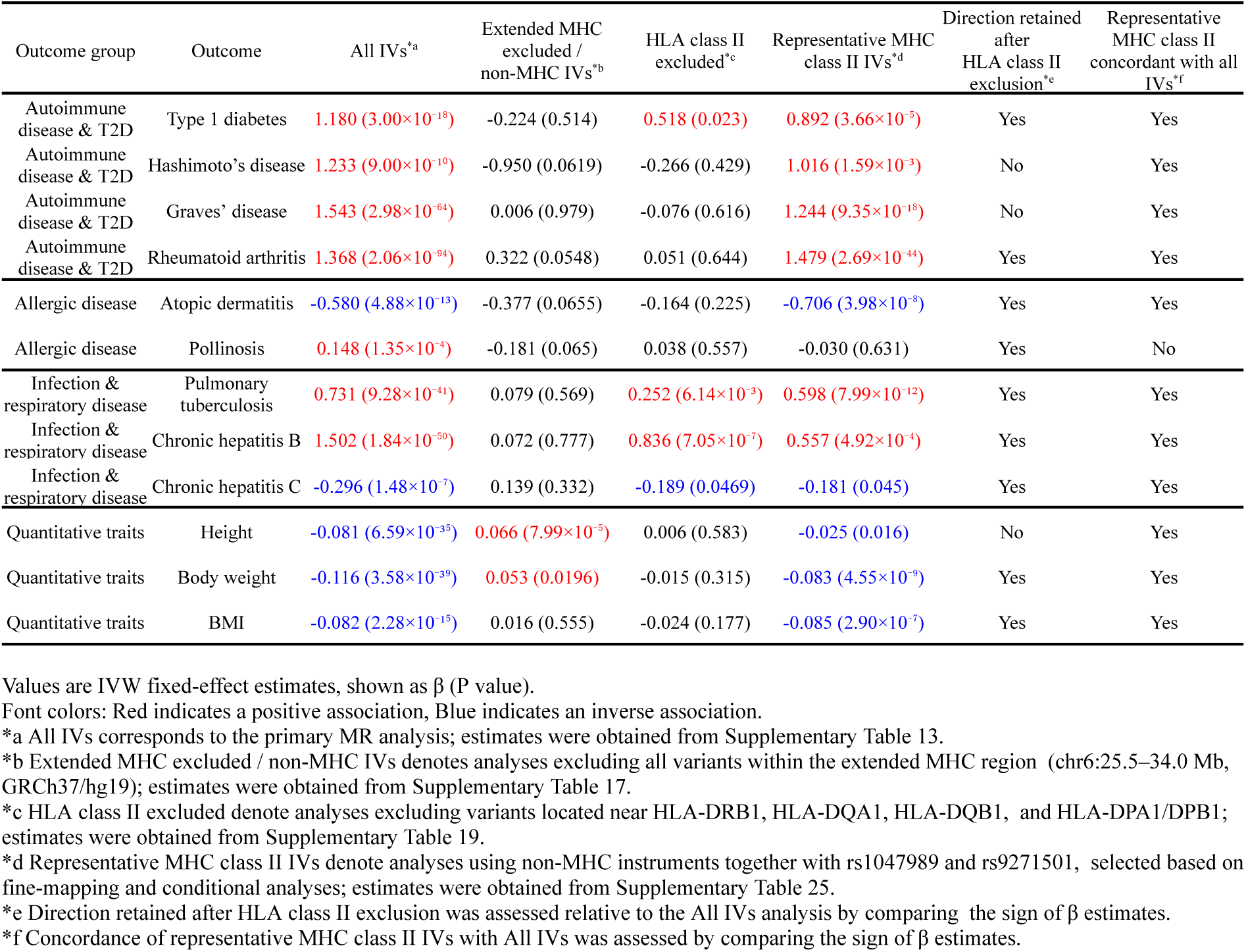
MHC-dependence sensitivity analysis of MR findings. Values are IVW fixed-effect estimates, shown as β (P value). Directional retention was assessed relative to the all-IV analysis by comparing the sign of β estimates.

**Supplementary Table 27.**
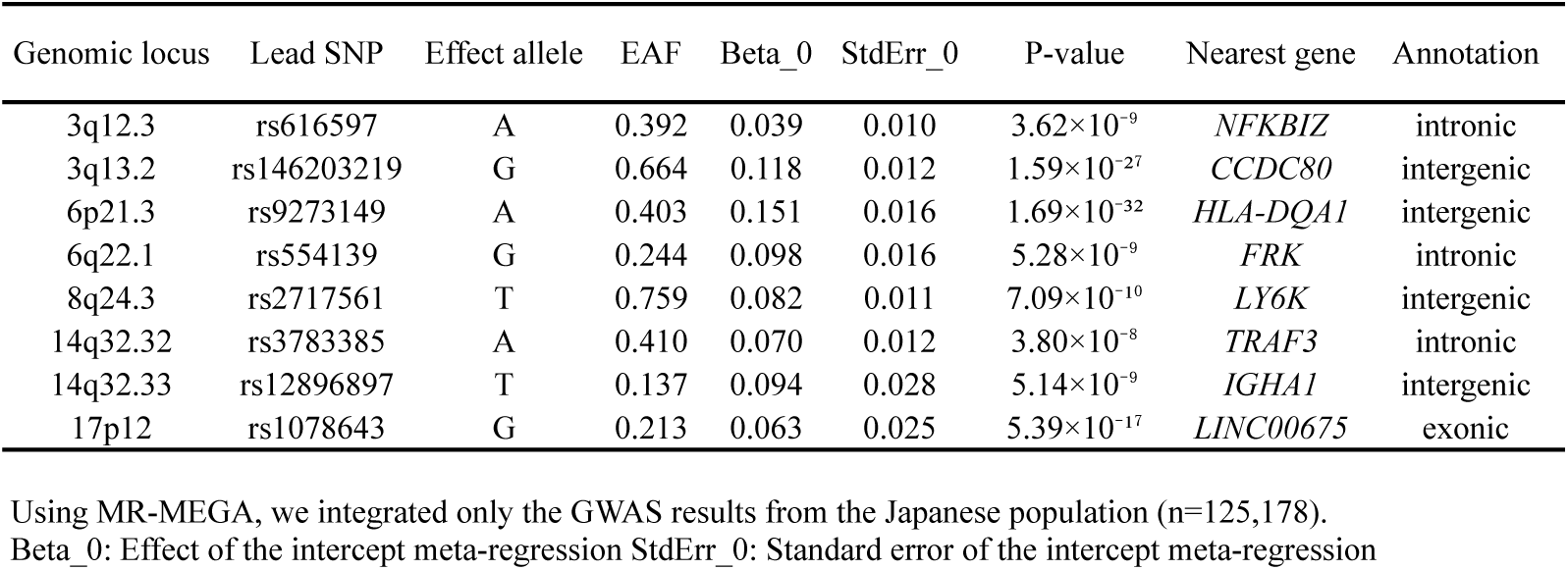
Meta-GWAS in Japan (using MR-MEGA). A list of regions reaching genome-wide significance.

| Genomic locus | Lead SNP | Effect allele | EAF | Beta_0 | StdErr_0 | P-value | Nearest gene | Annotation |
| --- | --- | --- | --- | --- | --- | --- | --- | --- |
| 3q12.3 | rs616597 | A | 0.392 | 0.039 | 0.010 | $3.62 \times 10^{-9}$ | *NFKBIZ* | intronic |
| 3q13.2 | rs146203219 | G | 0.664 | 0.118 | 0.012 | $1.59 \times 10^{-27}$ | *CCDC80* | intergenic |
| 6p21.3 | rs9273149 | A | 0.403 | 0.151 | 0.016 | $1.69 \times 10^{-32}$ | *HLA-DQA1* | intergenic |
| 6q22.1 | rs554139 | G | 0.244 | 0.098 | 0.016 | $5.28 \times 10^{-9}$ | *FRK* | intronic |
| 8q24.3 | rs2717561 | T | 0.759 | 0.082 | 0.011 | $7.09 \times 10^{-10}$ | *LY6K* | intergenic |
| 14q32.32 | rs3783385 | A | 0.410 | 0.070 | 0.012 | $3.80 \times 10^{-8}$ | *TRAF3* | intronic |
| 14q32.33 | rs12896897 | T | 0.137 | 0.094 | 0.028 | $5.14 \times 10^{-9}$ | *IGHA1* | intergenic |
| 17p12 | rs1078643 | G | 0.213 | 0.063 | 0.025 | $5.39 \times 10^{-17}$ | *LINC00675* | exonic |

Using MR-MEGA, we integrated only the GWAS results from the Japanese population (n=125,178). Beta\_0: Effect of the intercept meta-regression StdErr\_0: Standard error of the intercept meta-regression

## Notes

### Competing Interest Statement

The authors have declared no competing interest.

### Author Declarations

Informed consent for participation was obtained for all study subjects and approval was given by the Institutional Review Boards of Tohoku University (TMM Project), National Cancer Center (JPHC), Aichi Cancer Center (J-MICC, HERPACC), and Yamagata University (Yamagata Study).

### Summary of Updates

This version incorporates major revisions prepared in June 2026. We reframed anti-H. pylori IgG antibody titers as a composite host-response phenotype because antibody levels reflect both susceptibility to persistent infection and subsequent immune activation. To better characterize what this phenotype captures and assess the stability of its genetic associations, we expanded analyses of infection history, eradication-related antibody dynamics and older participants. HLA variants may influence both antibody responses and immune-related diseases. We therefore expanded the MR sensitivity and control analyses to examine whether correlations between genetic instruments or broader immune pathways could account for the observed associations. These analyses informed a revised interpretation centered on directional consistency and shared immunogenetic architecture, with clearer discussion of the biological meaning and methodological limits of the findings. The main text and supplementary materials have been reorganized around this framework.

