## Supplementary Information (including Supplementary Figures 1-12) for "Genome-wide association and Mendelian randomization analyses of host response to *Helicobacter pylori* infection implicate HLA polymorphisms and immune-mediated diseases"

**Table of contents**


Supplementary Note 2
- Cohorts 2
- TMM Cohorts 2
  - Cohort dataset 2
  - Genotype Data 3
    - *Genotype Calling* 3
    - *Genotype Imputation* 3
  - Principal Component Analysis 3
  - Traits 4
  - Post-imputation Quality Control 4
  - Genome-wide Association Studies 4
- Other Cohorts 5
- Ethics 5
- Validation of anti–*H. pylori* IgG antibody titers 6
- GWAS sensitivity analyses 7
- Supplementary information on Mendelian randomization 8
  - Study design 8
  - Sensitivity analyses using HLA-based instruments 9
  - Sensitivity analyses for specificity and directionality 10
- Supplementary discussion 11
  - The association of *LINC00675* 11
  - The association of *NT5DC1* and *FRK* 11
  - *H. pylori* and allergic and growth-related traits 11
  - *H. pylori* and psoriasis 12
  - *H. pylori* and tuberculosis 12
  - *H. pylori* and COVID-19 12

Supplementary Figures 14
- Supplementary Figure 1 14
- Supplementary Figure 2 15
- Supplementary Figure 3 16
- Supplementary Figure 4 17
- Supplementary Figure 5 18
- Supplementary Figure 6 20
- Supplementary Figure 7 21
- Supplementary Figure 8 23
- Supplementary Figure 9 25
- Supplementary Figure 10 26
- Supplementary Figure 11 28
- Supplementary Figure 12 29

Supplementary References 30

Consortia 34

#### Supplementary Note

##### Cohorts

This study utilized data from 125,178 participants across the following six cohorts in Japan:

- Tohoku Medical Megabank Organization (TMM) Community-Based Cohort (n=75,074)[1]
  - Specific health checkups in Iwate and Miyagi prefectures (n=56,967)
  - Community Support Centers of Tohoku University Tohoku Medical Megabank organization (ToMMo) in Miyagi prefecture (n=11,442)
  - Satellites of Iwate Tohoku Medical Megabank organization at Iwate Medical University (IMM) in Iwate prefecture (n=6,665)
- TMM Birth and Three-Generation Cohort (n=30,837)[2]
- Yamagata Study (n=5,266)[3]
- Japan Public Health Center-based Prospective Study (JPHC) (n=5,344)[4]
- Japan Multi-Institutional Collaborative Cohort Study (J-MICC Study) (n=3,468)[5]
- Hospital-based Epidemiologic Research Program at Aichi Cancer Center (HERPACC) (n=5,189)[6]

The cross-ancestry meta-analysis incorporated the results from the above cohorts and the findings of previous GWAS performed on populations of European descent[7]. Suk Yee Lam and others' GWAS encompassed data from Rotterdam Study (RS)-I, RS-II, Study of Health in Pomerania (SHIP), SHIP-TREND, Framingham Heart Study (FHS), Multi-Ethnic Study of Atherosclerosis (MESA), and the Generation R (GenR) study cohorts, comprising a total of 15,685 participants.

##### TMM Cohorts

##### Cohort dataset:

The TMM Project was launched with the aims of reconstruction from the Great East Japan Earthquake and establishment of personalized healthcare and medicine.[8] In the project, two prospective cohort studies were performed: the TMM Community-Based Cohort (TMM CommCohort) Study and the TMM Birth and Three-Generation (TMM BirThree) Cohort Study, which together recruited a total of more than 150,000 participants. In the TMM CommCohort Study, a total of more than 80,000 residents aged 20 years or older living in Iwate and Miyagi Prefectures, located on the Pacific side of Tohoku (northeastern) region of Honshu (the main island of Japan), were enrolled from May 2013 to March 2016.[1] On the other hand, in the TMM BirThree Cohort Study, a total of more than 70,000 participants, including fetuses and their parents, siblings, grandparents, and extended family members in Miyagi Prefectures were enrolled from July 2013 to March 2017.[2]

We used six subsets of the TMM CommCohort and TMM BirThree Cohort with available genotype data. Notably, the CommCohort consists of the specific health checkups and the Community Support Centers, and Satellites. We used 56,967 individuals from the specific health checkups in Iwate and Miyagi prefectures: 49,507 individuals genotyped on the Japonica Array version 2 (Dataset 1) and 9,336 individuals genotyped on the Japonica Array NEO (Dataset 2). Additionally, we included 11,842 individuals from the Community Support Centers of ToMMo in Miyagi prefecture (Dataset 3) and 6,806 individuals from the Satellites of Iwate ToMMo at Iwate Medical University in Iwate Prefecture (Dataset 4), both genotyped on the Japonica Array version 2. The TMM BirThree Cohort was divided into two datasets: 16,765 individuals genotyped on the Japonica Array version 2 (Dataset 5) and 14,454 individuals genotyped on the Japonica Array NEO (Dataset 6). We analyzed each of these subsets separately (**Supplementary Table 1 and 2**).

**Genotype Data:**

Out of all the participants in the two cohorts of the TMM Project, about 140,000 individuals have so far been genotyped on the Japonica Array version 2[9] and on the Japonica Array NEO[10]. The individuals were divided into 29 batches, each of which consisted of 50 plates (96 wells per plate), i.e 4,800 individuals, and a series of steps, from genotype calling to imputation, were performed for each batch.

***Genotype Calling:***

Following the Best Practices Genotyping Analysis Workflow provided by Thermo Fisher Scientific Inc., raw data (CEL files) were processed using the Affymetrix Power Tools (APT) v1.17.0, and the samples that failed to meet any of the following criteria were excluded: dish quality control (QC) values $\geq 0.82$, sample QC call rate $\geq 0.97$, plate pass rate (proportion of samples passing the former two criteria to all sample on a plate) $\geq 0.95$, and average call rate in passing samples per plate $\geq 0.985$. We performed "association tests" by assuming in turn one plate (96 individuals) as cases and the remaining 49 plates as controls, and confirmed that average P-values were distributed around 0.5, i.e., no significant plate effects were observed. Markers were classified, based on cluster resolution, into i) "PolyHighResolution", ii) "MonoHighResolution", iii) "NoMinorHom", iv) "Hemizygous", v) "CallRateBelowThreshold", vi) "Other", and vii) "OTV (OffTargetVariant). The markers classified into i)-iv) were defined as "recommended" and retained for further downstream steps. After statistical genetic QC procedures for excessive heterozygosity, sex discrepancy, cryptic relatedness, and population stratification were performed, individuals with a call rate < 0.95, heterozygosity outliers (3SD from mean), and variant sites that failed to meet any of the following criteria were excluded: P-value for Hardy-Weinberg equilibrium $\geq 1\times10^{-5}$, minor allele frequency (MAF) $\geq 0.01$, call rate (Mendelian inconsistent genotype calls were treated as missing) $\geq 0.99$. We also excluded one of each pair (or trio) of variants with the same positions. For the variants with a minor allele count of less than 6, the genotypes were replaced with those called separately in each plate, since this was shown to increase calling accuracy for extremely rare variants.[11]

***Genotype Imputation:***

The genotype data were pre-phased using SHAPEIT2[12], along with the —duohmm option[13], which incorporates information on relatedness between individuals to increase phasing accuracy. The phased genotypes were subsequently imputed using IMPUTE4[14]. In this study, a cross-imputed reference panel combining the 3.5KJPNv2 (N = 3052)[15] and the 1000 Genomes Project phase 3 panel (N = 2504)[16] was used. As the 3.5KJPNv2 panel is derived from certain Japanese populations, some variants may not be observed in other ancestral populations. Therefore, in order to obtain genotype data for a sufficient number of variants and significantly contribute to large-scale, multi-ancestral metaGWAS as a representative of the East Asian population, it will be necessary to impute the genotype calls using a cross-imputed panel generated from multiple reference panels.

**Principal Component Analysis:**

We performed principal component (PC) analysis to obtain PC scores, which were used as covariates to adjust for population stratification, as described later in the article. First, we excluded variant sites that failed to meet any of the following criteria: P-value for Hardy-Weinberg equilibrium $\geq 0.05$, MAF $\geq 0.05$, call rate $\geq 0.99$. Next, we performed linkage disequilibrium (LD)-based pruning with a threshold of $r^2 \geq 0.3$, a window size of 1,500 kb, and a step size of 150 sites, resulting in a total of about 22,000 sites retained. The PC analysis was performed in the reduced dataset using PLINK 2.0[17], and 10 calculated PC scores were projected to the original data set to be used to adjust for population stratification.

**Traits:**

Genome-wide association studies (GWAS) were performed for host response to *H. pylori*. Anti-*H. pylori* IgG antibody titers were measured using latex agglutination (Denka Seiken Co., Ltd., Tokyo, Japan), and an indicator for current or previous infection was defined as IgG antibody titers greater than 3 U/mL. In TMM Cohorts, individuals with the 25% highest IgG distribution comprised the case group, and individuals with the 75% lowest IgG distribution comprised the control group. Phenotypic values were obtained by regressing the raw values on age, sex, and 10 PC scores mentioned above, and by transforming the residuals by rank-based inverse normalization.[18]

**Post-imputation Quality Control:**

From the 29 batches encompassing 140,000 individuals, genotype data corresponding to any of the six datasets were extracted and subsequently combined. Individuals meeting any of the following criteria were excluded from the datasets: those who had withdrawn their consent as of October 28, 2021, those with inconsistent sex declarations between self-reporting and genotyping, those with a history of gastric cancer, and those with PC scores 1 or 2 that were $\geq 4$ standard deviations from the mean. Individuals who appeared in both datasets 1 and 3 or in both datasets 1 and 4 were excluded from dataset 1. Similarly, individuals present in both datasets 5 and 1 or in both datasets 5 and 3 were excluded from dataset 5. Following the filtering process, the remaining participants for datasets 1 through 6 were as follows: 47,831 in dataset 1, 9,136 in dataset 2, 11,442 in dataset 3, 6,665 in dataset 4, 16,552 in dataset 5, and 14,285 in dataset 6.

**Genome-wide Association Studies:**

We performed separate GWAS for each of the six datasets. A linear mixed model was employed for the GWAS, utilizing the Genome-wide Complex Trait Analysis (GCTA) software (version 1.94.0 beta)[19], as this cohort contained a substantial number of closely related individuals. The regression included age, sex, and top 10 PCs as covariates.

#### Other Cohorts

Yamagata Study[3] is a community-based prospective cohort study, performed in the Yamagata prefecture of Japan. It focuses on various health aspects among adult Japanese individuals aged 40 years and older. One aspect of the study investigates the relationship between lifestyle factors and various health outcomes, including mortality and social support within the community. The study has collected comprehensive data on the diet, health behaviors, and medical history of its participants, aiming to contribute to preventive healthcare strategies and enhance the understanding of factors influencing health and longevity in the region.

Japan Public Health Center-based Prospective Study (JPHC Study)[4] is a large-scale cohort study initiated in 1990, primarily focusing on investigating lifestyle habits and their relationships with the incidence of chronic diseases such as cancer, cardiovascular disease, and other lifestyle-related diseases. The study covers approximately 140,000 residents aged 40 to 69 years across 11 public health center areas in Japan. It aims to establish evidence to benefit health maintenance and improve disease prevention strategies through comprehensive data collection, including dietary habits, physical activity, smoking, and alcohol consumption. The JPHC Study has contributed significantly to understanding the impact of lifestyle factors on health and has published numerous research findings.

Japan Multi-Institutional Collaborative Cohort Study (J-MICC Study)[5] was established in 2005 to investigate gene-environment interactions and their impact on lifestyle-related diseases, particularly cancer. It is a large-scale, prospective cohort study, enrolling individuals aged 35 to 69 years from various regions of Japan. The study's main goals include identifying biomarkers for early cancer diagnosis and understanding how genetic traits interact with lifestyle factors to influence disease risk.

Hospital-based Epidemiologic Research Program at Aichi Cancer Center (HERPACC)[6] was initiated in 1988 to explore the risk and protective factors for cancer through hospital-based epidemiology. HERPACC focuses on large-scale case-referent studies across main cancer sites such as stomach, colorectal, lung, breast, and uterine cancers, evaluating the influence of lifestyle factors like diet, smoking, and exercise on cancer incidence in the Japanese population. This research aims to develop practical cancer prevention strategies and educate outpatients on reducing cancer risk.

Different platforms for genome-wide genotyping were employed by cohorts using standard procedures of the manufacturer. In genotyping, the Illumina platform was utilized by Yamagata Study (Human660W-Quad, HumanOmniExpressExome arrays, or Japonica Array Neo), JPHC (HumanOmniExpressExome BeadChip arrays), J-MICC (HumanOmniExpressExome arrays), and HERPACC (Human610-Quad, HumanCoreExome, or Infinium Asian Screening arrays). Additionally, a part of the Yamagata study employed the Japonica Array Neo.

To aid meta-analysis, JPHC, J-MICC, and HERPACC datasets were imputed to the 1000 Genomes Project phase 3 panel. In the Yamagata study, a cross-imputed reference panel combining the 3.5KJPNv2 and the 1000 Genomes Project phase 3 panel was used. Genome wide association analyses were performed in individual cohorts with adjustment for sex, age and study specific covariates.

#### Ethics

Informed consent for participation was obtained for all study subjects and approval was given by the Institutional Review Boards of Tohoku University (TMM Project), National Cancer Center (JPHC), Aichi Cancer Center (J-MICC, HERPACC), and Yamagata University (Yamagata Study).

**Validation of anti–*H. pylori* IgG antibody titers**

To characterize anti–*H. pylori* IgG antibody titers as a composite host phenotype reflecting susceptibility to persistent infection and subsequent immune activation, we analyzed self-reported infection data from the ToMMo CommCohort (Datasets 1, 2, and 3; see **Supplementary Table 1**). Of the 68,549 participants with available data on anti-*H. pylori* IgG antibody titers, 68,543 also had data on Pepsinogen (PG) 1/2 ratios. Among them, 12,481 participants reported a history of *H. pylori* infection, while 56,062 participants did not provide infection status. PG1 and PG2 levels were measured using LIA, and the PG1/2 ratio was calculated as PG1 divided by PG2. We compared IgG antibody titers and PG1/2 ratios between the self-reported infection group and the no-report group using the Mann-Whitney *U* test. Additionally, the correlation between IgG antibody titers and PG1/2 ratios was assessed across all participants using Spearman's rank correlation coefficient. To further assess potential bias arising from antibody decline following eradication therapy, participants who self-reported a history of *H. pylori* infection were stratified by eradication status, and the proportions of individuals with high antibody titers (above the top 25 % of each dataset) were compared between groups.

Compared to the non-reporting group (n = 56,062), the self-reported infection group had significantly higher IgG titers (Mann-Whitney U test: W = 222,591,616, P-value < $2.2\times10^{-16}$; **Supplementary Fig. 1a**) and significantly lower PG1/2 ratios (W = 425,587,934, P-value < $2.2\times10^{-16}$; **Supplementary Fig. 1b**). Furthermore, a significant negative correlation was observed between IgG titers and PG1/2 ratios (Spearman's $\rho$ = -0.691, P-value < $2.2\times10^{-16}$; **Supplementary Fig. 1c**) across all participants. The PG1/2 ratio is known to decrease due to *H. pylori* infection and associated gastric mucosal changes, such as intestinal metaplasia and atrophic gastritis[20][21]. The consistent associations between IgG titers, PG1/2 ratios, and self-reported infection status support the validity of anti-*H. pylori* IgG antibody titers as a composite host phenotype reflecting susceptibility to persistent infection and its associated immune and gastric mucosal changes as previously pointed out[22]. Among the 12,481 participants who reported a history of *H. pylori* infection, 5,381 (43.1%) exhibited high antibody titers (above the top 25% within each dataset). Of these, 4,283 individuals had undergone eradication therapy prior to antibody measurement, and 1,027 of them still maintained high titers after eradication. Assuming a comparable baseline distribution of antibody titers, approximately 819 individuals (19.1% of the eradicated group; 6.6% of all self-reported infected participants) were estimated to have experienced a substantial post-eradication decline in antibody levels. These results indicate that potential bias due to eradication-related antibody decline is likely limited in this cohort. In addition, among individuals for whom the interval between eradication therapy and antibody measurement was available, we conducted a detailed analysis of 2,349 participants whose antibody measurements had been obtained within 8.5 years after eradication. In this subset, anti–*H. pylori* IgG antibody titers did not decline rapidly after eradication, but instead decreased gradually over periods ranging from several months to several years. These findings further support the interpretation that anti–*H. pylori* IgG levels reflect an ongoing or accumulated biological process rather than a transient indicator of infection status alone **(Supplementary Fig. 1d)**.

**GWAS sensitivity analyses**

To further evaluate the robustness of our GWAS results with respect to potential bias related to antibody dynamics following *H. pylori* eradication therapy, and environmental heterogeneity in pathogen encounter, we performed a series of sub-analyses. First, we re-ran the analysis in the ToMMo CommCohort after excluding individuals with self-reported H. pylori eradication to assess whether eradication-related changes in antibody levels affected the genetic associations (n = 61,156; 13,919 cases with high anti-*H. pylori* IgG levels and 47,237 controls). Second, we investigated the effect of different antibody cutoff thresholds by performing three additional GWAS: (i) a unified cutoff of 3 U/mL across all cohorts (N = 125,178; 58,106 cases and 67,072 controls); (ii) an extreme group comparison of the top 25% vs ≤ 3 U/mL within the TMM dataset (n = 87,557; 27,010 cases and 60,547 controls); and (iii) another extreme comparison of ≥ 10 U/mL vs ≤ 3 U/mL (n = 93,765; 33,218 cases and 60547 controls). These sub-analyses were designed to test the stability of key association signals excluding participants potentially affected by eradication-related antibody decline, across alternative phenotype definitions, and within a subgroup with a higher likelihood of prior pathogen encounter.

In the eradication-exclusion analysis using the TMM CommCohort (top 25% vs others; n = 61,156; 13,919 cases and 47,237 controls), the major association signals—including those in the HLA class II region—were reproduced at the genome-wide significance level (**Supplementary Fig. 4a and Supplementary Table 3**). The strongest association was in the intronic region of *HLA-DQB1* at 6p21.3 (lead SNP: rs28746807, OR = 1.157, 95% CI: 1.130 – 1.185, P-value = $1.62\times10^{-24}$). The second strongest was in the intergenic region between *CCDC80* and *CD200R1L* at 3q13.2 (lead SNP: rs146203219, OR = 1.138, 95% CI: 1.110 – 1.166, P-value = $1.62\times10^{-18}$), and the third strongest was in the exonic region of *LINC00675* at 17p12 (lead SNP: rs1078643, OR = 1.150, 95% CI: 1.112 – 1.189, P-value = $5.79\times10^{-16}$).

In the cutoff-variation analyses, which were performed to test the robustness of the main combined GWAS meta-analysis (N = 125,178; 37,410 cases and 87,768 controls), all major regions identified in the primary analysis—*CCDC80*, *HLA-DQA1*, *LINC00675*, *PSCA*, and *TRAF3*—remained genome-wide significant when using the unified 3 U/mL threshold (**Supplementary Fig. 4b)**. For example, rs9271146 in the intergenic region between *HLA-DQA1* and *HLA-DQB1* at 6p21.3 retained strong significance (OR = 1.155, 95% CI: 1.126 – 1.184, P-value = $1.12\times10^{-29}$), and rs146203219 in the intergenic region between *CCDC80* and *CD200R1L* at 3q13.2 remained robust (OR = 1.121, 95% CI: 1.100 – 1.142, P-value = $2.03\times10^{-33}$). Although the extreme group comparisons (top 25% vs ≤ 3 U/mL; ≥ 10 U/mL vs ≤ 3 U/mL) had smaller sample sizes and therefore slightly reduced significance, consistent associations persisted in the *HLA-DQA1*, *CCDC80*, and *LINC00675* (**Supplementary Fig. 4c, 4d and Supplementary Table 3)**.

To further reduce the potential influence of environmental heterogeneity in pathogen encounter, we additionally performed a GWAS restricted to individuals aged 60 years or older (top 25% vs others; n = 50,428; 15,659 cases and 34,769 controls), among whom prior *H. pylori* encounter is expected to be more common in older Japanese birth cohorts[23]. In this analysis, the major association signals remained concentrated in the MHC class II region, consistent with the results of the primary analysis **(Supplementary Fig. 4e and Supplementary Table 3)**.

**Supplementary information on Mendelian randomization**

**Study design:**

In observational studies, when adjusting for confounders is challenging, the Instrumental Variable Method (IV method) can be applied to estimate causal effects without bias[24]. The IV method estimates the causal effect of exposure-outcome (β) indirectly, using the causal effects of IV-exposure (β1) and IV-outcome (β2), thus deducing β (=β2/β1) indirectly. This approach requires the fulfillment of three key assumptions:

- Related to exposure (Assumption 1)
- Influences the outcome only through exposure (Assumption 2)
- Not associated with any unmeasured confounders affecting both exposure and outcome (Assumption 3)

In Mendelian randomization (MR), gene variants (G) are used as instrumental variables (IV)[25]. Gene variants related to exposure are typically selected from existing GWAS results performed in the same ancestral population, satisfying Assumption 1. Due to Mendel's laws of inheritance, it is presumed that the distribution of background factors between groups with and without specific gene variants will be generally equal, thereby likely fulfilling Assumption 3. If a gene variant does not affect the outcome except through exposure, Assumption 2 is also met, and thus all major assumptions of the IV method are considered satisfied. Deviation from Assumption 2 is referred to as Horizontal Pleiotropy, which is a significant challenge in MR, but various sensitivity analysis techniques have been developed to address this issue.

In our association analyses, the primary signals for anti–*H. pylori* IgG antibody titers were consistently observed in the HLA class II region (near HLA-DQA1/DQB1). An independent European study likewise identified the same region as the major locus for *H. pylori*–related serological traits[26]. While such convergence does not, by itself, validate these variants as specific instruments, it indicates that a substantial proportion of the genetic architecture underlying host response to *H. pylori* maps to this region, thereby motivating careful evaluation of HLA-based IVs. Although pleiotropy within the HLA region is a recognized limitation in MR analyses of infectious exposures[27], our sensitivity analyses indicate that the present findings are unlikely to be driven solely by generalized HLA pleiotropy. Rather, the overall pattern is most consistent with a biologically meaningful component related to genetic liability to high anti–*H. pylori* IgG levels, rather than nonspecific immune pleiotropy. Mechanistically, the HLA class II region plausibly modulates immune responses to *H. pylori*. For example, *H. pylori* infection suppresses CIITA (class II transactivator) expression in macrophages, reducing HLA-DR surface expression[28]. HLA polymorphisms also alter class II gene expression efficiency and peptide-binding spectra[29], and specific HLA-DR alleles can trigger autoimmune responses through structural mimicry between microbial and self-antigens[30].

Therefore, although the IV assumptions may not be grossly violated, the concentration of signals within a highly pleiotropic immunogenetic region, together with the composite nature of the exposure, warrants cautious interpretation of MR estimates as reflecting directionally consistent associations rather than definitive causal effects.

In this study, the TwoSample MR (version 0.5.7) package[31] was used for MR analysis to evaluate the potential directionally consistent associations between genetic liability to high anti-*H. pylori* IgG levels and related diseases. Summary statistics for the outcome traits[32][33] are listed in **Supplementary Table 12**.

**Sensitivity analyses using HLA-based instruments:**

*H. pylori* infection has been implicated in several extra-gastroduodenal conditions, supporting the evaluation of host-response genetic signals beyond gastric disease[34]. To further evaluate the regional and functional structure of the association signals, we performed regional and function-based robustness analyses. First, to evaluate whether the associations were driven by the MHC/HLA region, we repeated the MR analyses using instruments that were (i) restricted to the extended MHC region (chr6:25.5–34.0 Mb, GRCh37/hg19), (ii) restricted to the HLA class II region (near *HLA-DRB1*, *DQA1*, *DQB1*, and *DPA1/DPB1*), or, in parallel, using instruments in which (iii) all MHC variants or (iv) all HLA class II variants were excluded. Excluding all variants within the extended MHC region from the IV set markedly attenuated and destabilized the directionally consistent associations observed in the primary analysis. In contrast, when only variants within the HLA class II region were removed, the direction of effect was largely preserved for major outcomes, including type 1 diabetes. Conversely, restricting the instruments to HLA class II variants preserved statistical support for several associations **(Supplementary Table 17, 18, 19, and 20)**.

Because *H. pylori* strongly activates NF-κB signaling, we next performed analogous analyses in which instruments were either restricted to variants near NF-κB–related genes (*NFKBIZ*, *TRAF3*, *BTNL2*) or these variants were excluded from the full IV set. Although this limited subset alone did not yield statistically significant effects, excluding these variants from the full IV set reduced the significance of several associations, consistent with a contribution of variation near NF-κB–related genes to the observed association signals **(Supplementary Table 21 and 22)**. We further conducted functionally informed subset analyses using variants selected on the basis of biological relevance to *H. pylori* immunity—specifically, variants within the HLA class II region and those near NF-κB–related genes. Using this combined instrument set, estimated effects remained directionally consistent, and most associations retained statistical significance **(Supplementary Table 23)**.

In addition, we repeated the key MR analyses after applying more stringent LD clumping criteria ($r^2 < 0.001$ within a 10 Mb window) to further reduce the potential influence of local correlation among instruments. The core immune-related association pattern was retained under this stricter IV set. This result supports the robustness of the central pattern to more stringent control of residual local correlation among instruments, particularly within the MHC region **(Supplementary Table 24)**.

Finally, because complete exclusion of the MHC region may also remove a biologically meaningful component of the host-response phenotype, we performed a representative MHC class II signal sensitivity analysis using variants selected based on our fine-mapping and conditional analyses. This approach is consistent with recent MR work on anti-EBNA1 IgG and multiple sclerosis, where MHC-centered antibody-response instruments were treated as informative while LD and pleiotropy were explicitly addressed[35]. In this analysis, the MHC class II contribution was represented by two variants supported by our fine-mapping and conditional analyses: rs1047989, a missense variant in HLA-DQA1, and rs9271501, the residual lead variant that emerged after conditioning on rs1047989. These variants showed low-to-modest pairwise LD ($D' = 0.23$, $r^2 = 0.05$) and correspond to two partly independent MHC class II signal components related to HLA-DQA1 and HLA-DRB1/DQB1 allelic polymorphisms. We then repeated the MR analyses using non-MHC instruments together with these two representative variants. The core immune-related association pattern was retained and was broadly concordant with that observed in the primary and stringent LD-clumping analyses (**Supplementary Table 25**).

We further summarized these complementary IV-set analyses in a comparative MHC-dependence sensitivity table, which contrasts IVW estimates across the full all-IV set, the extended MHC–excluded/non-MHC IV set, the HLA class II–excluded IV set, and the representative MHC class II IV set described above **(Supplementary Table 26)**. Overall, these regional exclusion, region-restricted, stringent LD-clumping, and representative-signal analyses support the biological relevance of HLA class II antigen-presentation signals within the anti–*H. pylori* IgG host-response phenotype. At the same time, the preservation of effect directions after excluding HLA class II variants for several key outcomes indicates that the broader HLA-enriched MR pattern is not uniformly attributable to HLA class II alone. The overall pattern is therefore most consistent with a shared host-response architecture involving, but not limited to, HLA class II, rather than an artifact generated simply by redundant inclusion of multiple locally correlated MHC variants.

**Sensitivity analyses for specificity and directionality:**

To verify the specificity of the MR results, we performed negative and positive control analyses. As a mechanism-specific negative control, we tested the hypothesis that the observed associations might reflect generalized immune activation or a tendency toward higher globulin levels, rather than antigen-specific immune responses to *H. pylori*. Anti–*H. pylori* IgG antibody titers reflect antibody responses to a specific antigen; however, if the major source of inter-individual variation were instead a nonspecific predisposition toward higher overall immunoglobulin levels, similar associations would be expected for biomarkers that capture antigen-independent globulin levels. We therefore performed additional MR analyses using non-albumin protein (NAP) and zinc sulfate turbidity test (ZTT) as negative control exposures. GWAS summary statistics for NAP were obtained from the BioBank Japan PheWeb resource. For ZTT, we used published lead variants from ZTT-associated loci reported in the BioBank Japan quantitative-trait GWAS[36]. Details of the sample sizes and IV variants are provided in **Supplementary Table 12 and 16**. ZTT is a classical assay that reflects increases in serum γ-globulin, particularly IgG, whereas NAP reflects a broader non-albumin globulin fraction. Because neither ZTT nor NAP measures antibody responses to a specific antigen, they can be interpreted as proxies for generalized immune activity rather than antigen-specific humoral responses. If the associations observed in the primary MR analyses were driven mainly by generalized immune activation or hyperglobulinemia, similar directionally consistent associations would be expected when ZTT or NAP was used as the exposure. However, these negative-control MR analyses did not show a robust and directionally concordant pattern across multiple MR methods corresponding to the primary anti–*H. pylori* IgG results, particularly for the autoimmune outcomes of interest **(Supplementary Table 14)**. These findings argue against the interpretation that the primary associations are explained solely by a nonspecific tendency toward increased immunoglobulin levels. Taken together, these mechanism-specific negative control analyses suggest that the associations between anti–*H. pylori* IgG antibody titers and autoimmune disease outcomes are difficult to explain solely by generalized globulin elevation or nonspecific immune activation, and are more consistent with mechanisms related to antigen-specific host responses to *H. pylori*.

For positive controls, we performed MR analyses using atrophic gastritis measured by the pepsinogen (PG) method as the outcome. Atrophic gastritis was defined biochemically as PG I < 70 ng/mL and a PG I/II ratio < 3.0[37]. Because the GWAS data for anti–*H. pylori* IgG antibody titers and atrophic gastritis were derived from the same participants, we used the replication GWAS dataset as the exposure (genetic liability to anti–*H. pylori* IgG levels) and the discovery GWAS dataset as the outcome (atrophic gastritis). Although the MR-Egger analysis did not show a statistically significant association, possibly due to reduced statistical power resulting from sample splitting, the primary IVW and other sensitivity analyses retained directionally consistent associations (**Supplementary Table 14**), supporting the biological relevance of the selected genetic instruments.

Furthermore, we performed MR analyses with exposure–outcome roles swapped; however, no evidence of directionally consistent associations was observed when diseases or traits were treated as exposures and genetic liability to high anti–*H. pylori* IgG levels as the outcome. However, these results depend on the power of the GWAS results (summary statistics) used, and it cannot be ruled out that future updates to summary statistics and increased detection power might reveal new directionally consistent associations.

#### Supplementary discussion

##### Supplementary discussion on the association of *LINC00675*:

In the discovery and combined GWAS meta-analyses, we additionally identified a significant association of anti-*H. pylori* IgG antibody titers with a SNP on the *LINC00675* gene on chromosome 17p12, while the association was suggestive in the replication GWAS. *LINC00675*, a long non-coding RNA (lncRNA), has been previously implicated in the progression of several cancers, including gastric and pancreatic cancers[38][39], underscoring its potential relevance in the context of *H. pylori* infection. Some studies have reported that *LINC00675* suppresses cell proliferation and metastasis in colorectal cancer by acting on miR-942 and Wnt/beta-catenin signaling[40]. The lack of replication at the genome-wide significant level may stem from sample heterogeneity, variations in genotyping platforms, and differences in imputation reference panels utilized across the discovery and replication GWAS (**Supplementary Table 1**). Additionally, a smaller number of variants in linkage disequilibrium (LD) near the *LINC00675* gene might have contributed to this inconsistency (**Supplementary Fig. 11**).

##### Supplementary discussion on the association of *NT5DC1* and *FRK*:

In addition to our Japanese cohort, we conducted a cross-ancestry GWAS meta-analysis by incorporating previous GWAS results from European populations. This cross-ancestry analysis confirmed the associations identified in the Japanese GWAS without significant loss of significance post-integration. Furthermore, a novel significant association was observed in the intronic region of *NT5DC1* at 6q22.1. However, this variant demonstrated effect-size heterogeneity between Japanese and European populations. A meta-analysis restricted to the Japanese population utilizing MR-MEGA software revealed a more robust association at the intronic region of *FRK* (Fyn Related Src Family Tyrosine Kinase), which is located in proximity to *NT5DC1* (**Supplementary Fig. 12 and Supplementary Table 27**). This suggests that the novel association might have emerged due to the application of MR-MEGA software rather than through enhanced association from the cross-ancestry meta-analysis. *NT5DC1* has been implicated in neurological disorders, although its specific role in *H. pylori* infection remains uncertain. In contrast, *FRK* has been characterized as a tumor suppressor, playing significant roles in regulating cell proliferation and growth inhibition[41].

##### Supplementary discussion on *H. pylori* and allergic and growth-related traits

The directionally consistent associations with traits such as pollinosis and height may appear less intuitive at first glance. The positive associations with pollinosis may reflect population-specific factors in Japan, where adult-onset Japanese cedar pollinosis is highly prevalent[42]. Early-life *H. pylori* infection can induce immune tolerance through regulatory T-cell education, whereas chronic or adult-acquired infection may promote systemic inflammation[43], increase gastric and intestinal mucosal permeability[44], and alter immune tolerance[28], potentially enhancing allergic sensitization later in life. In adult-onset pollinosis, particularly among immunosenescent individuals, persistent *H. pylori* infection may plausibly act as an adjuvant sustaining low-grade inflammation, consistent with mechanisms described for *H. pylori*–related immune dysregulation[45]. In contrast, the inverse association with height is biologically plausible. Chronic *H. pylori* infection can damage gastric X/A-like (ghrelin-producing) cells, leading to reduced ghrelin secretion, which regulates both appetite and growth hormone (GH) release[46]. Reduced ghrelin levels, together with impaired IGF-1 secretion, may suppress linear growth and appetite[47]. Moreover, chronic infection can impair iron and vitamin $B_{12}$ absorption[48], further inhibiting growth-plate function. These mechanisms provide a coherent biological explanation for the inverse associations observed with height and weight. However, certain host genetic variants predisposing to persistent *H. pylori* colonization may also correlate with socioeconomic or nutritional factors[49]; therefore, residual confounding and survival bias cannot be entirely excluded. While these mechanisms are consistent with the direction of our MR results, the precise causal pathways remain speculative and require validation in independent cohorts.

**Supplementary discussion on *H. pylori* and psoriasis:**

Past epidemiological analyses have shown a positive correlation between *H. pylori* infection and psoriasis[50], with reports indicating that eradication of *H. pylori* could enhance the treatment efficacy for psoriasis[51]. However, studies in Taiwan and Iran did not demonstrate significant associations[52][53], leading to inconsistent results. Recent research has shown a positive correlation between *H. pylori* infection and psoriasis only in patients with moderate to severe symptoms[54], suggesting a complex interplay of mechanisms behind this relationship. In our MR analysis, no significant association was found between genetic liability to high anti-*H. pylori* IgG levels and psoriasis. Psoriasis, like Behçet's disease[55][56] and ankylosing spondylitis[57][58][59], is noted for having a stronger involvement of HLA-class I genes regions in disease susceptibility compared to HLA-class II genes regions[60][61][62]. The summary statistics for psoriasis in our study showed relatively weak associations in the HLA-class II region (especially near the *HLA-DRB1* gene), which might have affected the results of the MR analysis.

**Supplementary discussion on *H. pylori* and tuberculosis:**

Epidemiological studies have yielded inconsistent results regarding the relationship between *H. pylori* infection and tuberculosis. Reports vary by region, with some indicating that *H. pylori* infection reduces the risk of tuberculosis (USA)[63] and some finding no significant association (Japan, Peru)[64][65]. From the perspective of reduced risk, mechanisms like the hygiene hypothesis and bystander effects suggest that *H. pylori* infection might enhance immunity against other bacteria and viruses, including tuberculosis[66][67]. However, it has also been noted that persistent *H. pylori* infection may induce immune tolerance[64][68][69][70], potentially increasing susceptibility to other infectious diseases. Given the likely association of these infections with poor hygiene and low socioeconomic status, interpreting such studies necessitates careful consideration of potential confounding factors. In our MR analysis, genetic liability to high anti-*H. pylori* IgG levels showed positive associations with tuberculosis. While it is challenging to completely eliminate the influence of biases, local $r_g$ analysis within the MHC-class II region showed local positive correlations, and HLA association analysis identified shared risk HLA-class II alleles (HLA-DR9[71]), suggest that part of this association may reflect a shared immunogenetic architecture.

**Supplementary discussion on *H. pylori* and COVID-19:**

In our MR analysis, genetic liability to high anti-*H. pylori* IgG levels showed nominal inverse associations with COVID-19 susceptibility; however, evidence of horizontal pleiotropy in the MR-Egger test raises uncertainty regarding the robustness of this relationship. Epidemiological studies have also not reached a consensus on the link between *H. pylori* infection and COVID-19. Some data suggest a negative correlation, as lower COVID-19 incidence rates are observed in developing countries[72] with higher *H. pylori* infection rates; however, such ecological comparisons are highly susceptible to confounding by socioeconomic, demographic, and healthcare-related factors. Conversely, there are reports of exacerbated gastrointestinal symptoms in COVID-19 patients with *H. pylori* infection[73]. Moreover, individuals carrying the HLA gene polymorphism DR9 might have a higher risk of severe COVID-19 outcomes[74]. It is essential to recognize that susceptibility to infection and the risk of severe disease following infection may be influenced by distinct genetic factors. For instance, a study from Turkey suggested that HLA alleles associated with the risk of *H. pylori* infection do not necessarily coincide with those causing severe conditions like gastritis or gastric cancer following infection[75]. Given the evidence of pleiotropy and the complex immunogenetic architecture underlying both host responses to *H. pylori* and viral infections, these findings should not be interpreted as evidence of a direct protective effect of *H. pylori*–related host immune response on COVID-19 susceptibility. Further validation in independent datasets is warranted.

### Supplementary Figures

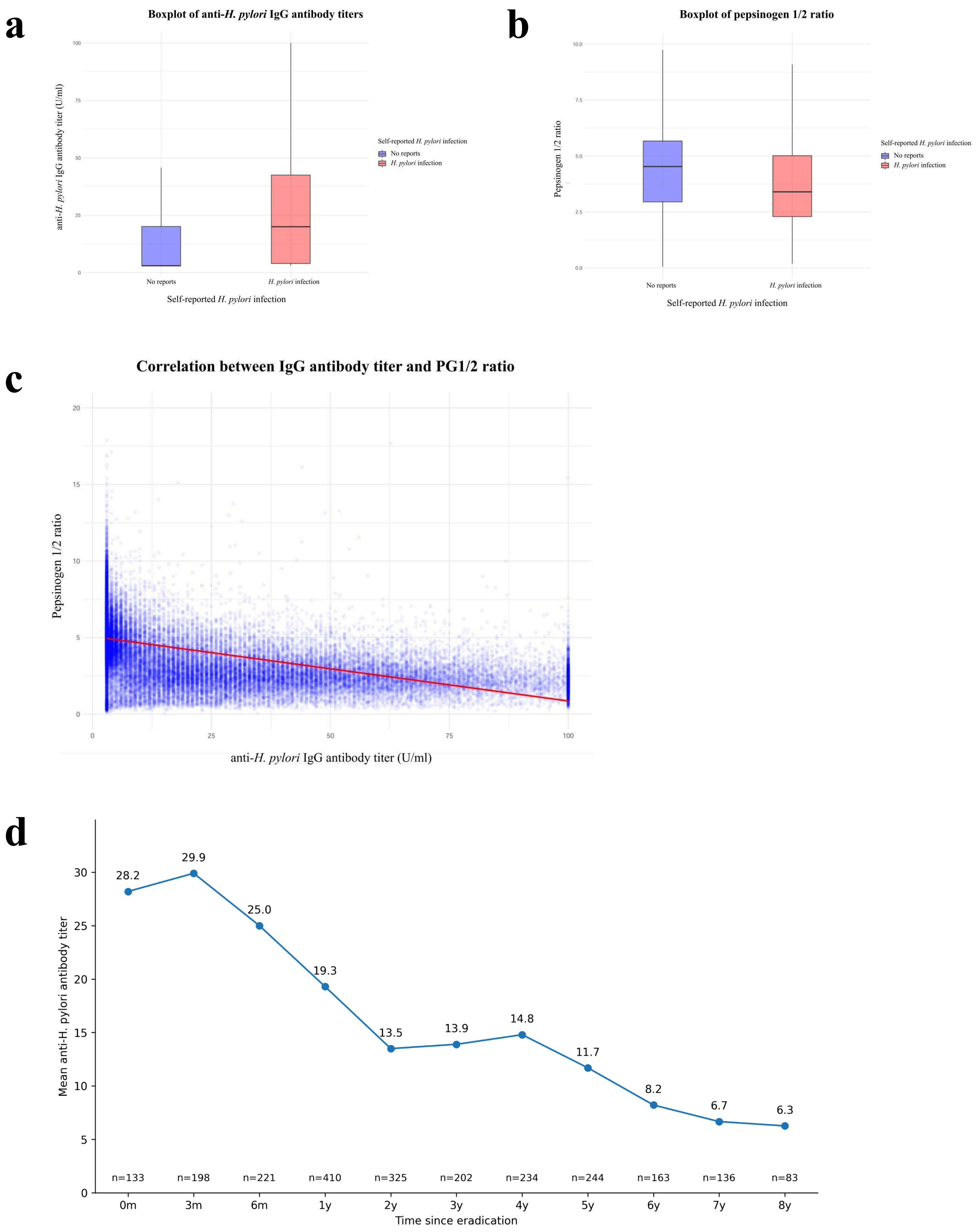

**Supplementary Fig. 1**

**Comparison of IgG antibody titers and pepsinogen (PG) 1/2 ratios by self-reported *H. pylori* infection status, and correlation between these biomarkers.**

The no-report group consists of participants from the TMM CommCohort (Dataset 1, 2, and 3; see **Supplementary Table 1**) who were included in the study but did not provide self-reported infection status. **a,** Distributions of IgG antibody titers in participants who self-reported *H. pylori* infection (n = 12,481) and those who did not report infection (n = 56,062). The median IgG antibody titer was higher in the self-reported *H. pylori*-positive group compared to the no-report group. In the no-report group, the first quartile (Q1), median (Q2), and third quartile (Q3) were 3.0, 3.0, and 20.1 U/ml, respectively, whereas in the self-reported *H. pylori*-positive group, they were 4.0, 20.0, and 42.5 U/ml, respectively. A Mann–Whitney *U* test indicated a statistically significant difference between the two groups ($W$ = 222,591,616, P-value < $2.2\times10^{-16}$). **b,** Distributions of the pepsinogen (PG) 1/2 ratio in the same two groups. The median PG1/2 ratio was lower in the self-reported *H. pylori*-positive group. In the no-report group, Q1, Q2, and Q3 were 3.0, 4.5, and 5.7, respectively, whereas in the self-reported *H. pylori*-positive group, they were 2.3, 3.4, and 5.0, respectively. A Mann–Whitney *U* test indicated a statistically significant difference ($W$ = 425,587,934, P-value < $2.2\times10^{-16}$). **c,** Correlation analysis among all participants (n = 68,543) demonstrated a significant negative correlation between IgG antibody titers and the PG1/2 ratio (Spearman's $\rho$ = -0.691, P-value < $2.2\times10^{-16}$). **d,** Mean anti-*H. pylori* IgG antibody titers according to the interval between eradication therapy and antibody measurement among participants with self-reported *H. pylori* infection and available information on the timing of eradication therapy. Antibody titers showed a gradual decline over several years after eradication, rather than a rapid decrease immediately after treatment. The number of participants in each time category is shown below the x-axis.

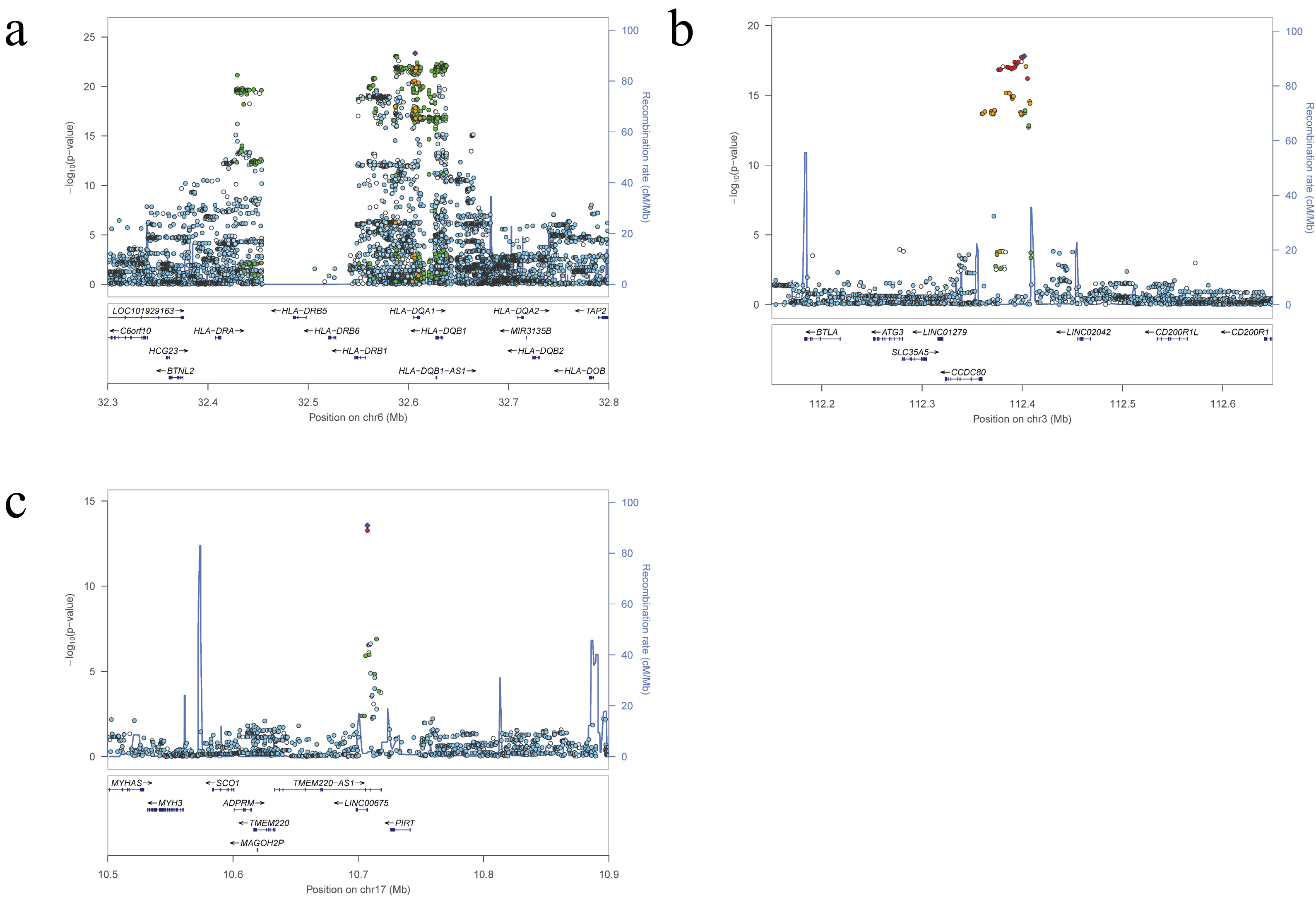


**Supplementary Fig. 2**

**Association signals around the significant region in 6p21.3, 3q13.2, and 17p12 loci of the discovery GWAS.**

The panel is association signals around significant locus. The X-axis represents chromosomal positions (GRC37/hg19) and the Y-axis represents -log10 P-values. The lead variant is shown in purple. Colors represent the degree of LD ($r^2$) between each variant and the lead variant. LD was calculated based on the 1000 Genomes Mar 2012 ASN data. **a,** Association signals around the significant region in 6p21.3 loci. Regional association plot showing SNP significance and genes around lead SNP rs28383359. **b,** Association signals around the significant region in 3q13.2 loci. Regional association plot showing SNP significance and genes around lead SNP rs146203219. **c,** Association signals around the significant region in 17p12 loci. Regional association plot showing SNP significance and genes around lead SNP rs1078643.

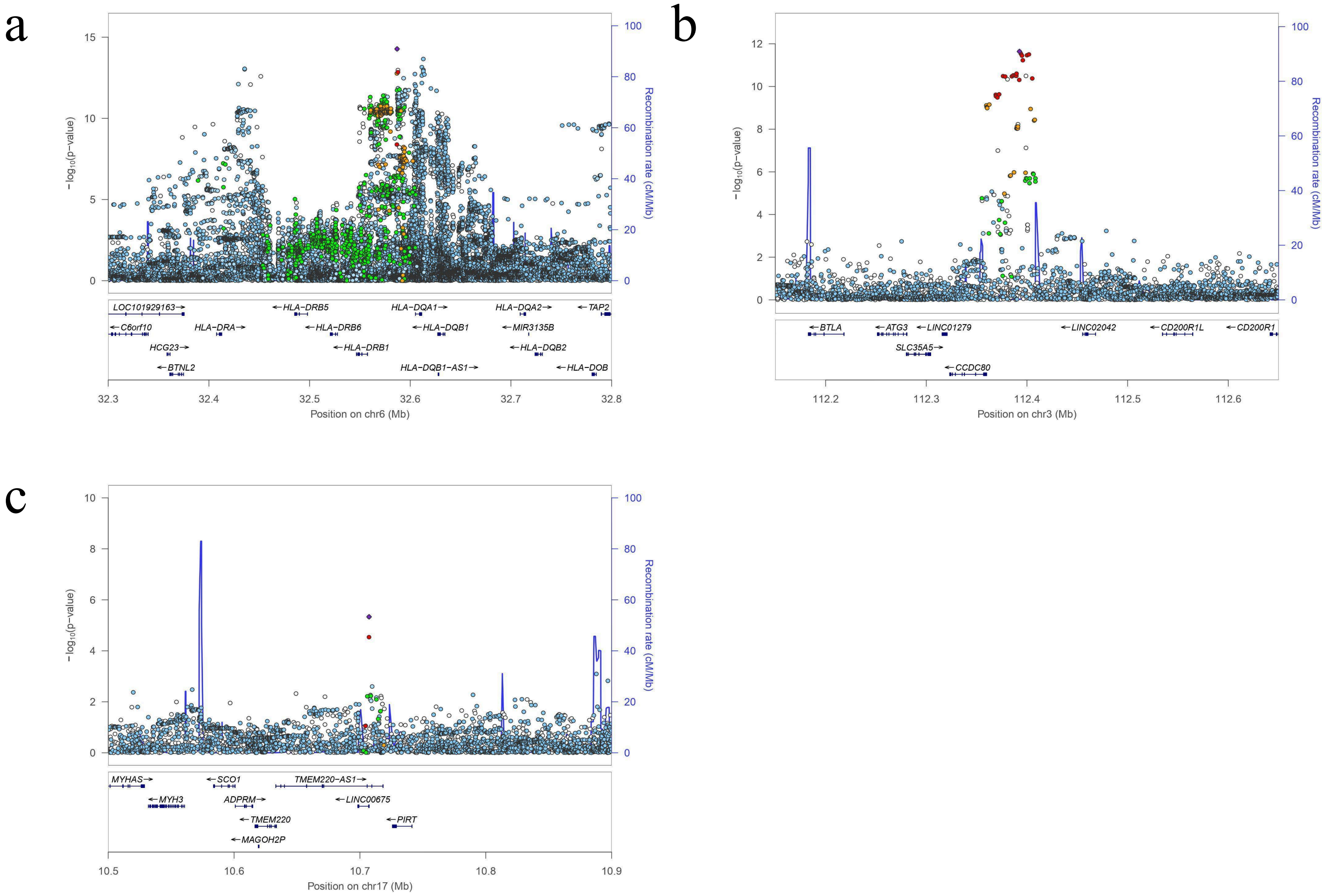


**Supplementary Fig. 3**

**Association signals around the significant region in 6p21.3, 3q13.2, and 17p12 loci of the replication GWAS.**

The panel is association signals around significant locus. The X-axis represents chromosomal positions (GRC37/hg19) and the Y-axis represents -log10 P-values. The lead variant is shown in purple. Colors represent the degree of LD ($r^2$) between each variant and the lead variant. LD was calculated based on the 1000 Genomes Mar 2012 ASN data. **a,** Association signals around the significant region in 6p21.3 loci. Regional association plot showing SNP significance and genes around lead SNP rs9271367. **b,** Association signals around the significant region in 3q13.2 loci. Regional association plot showing SNP significance and genes around lead SNP rs12634199. **c,** Association signals around the significant region in 17p12 loci. Regional association plot showing SNP significance and genes around lead SNP rs1550656.

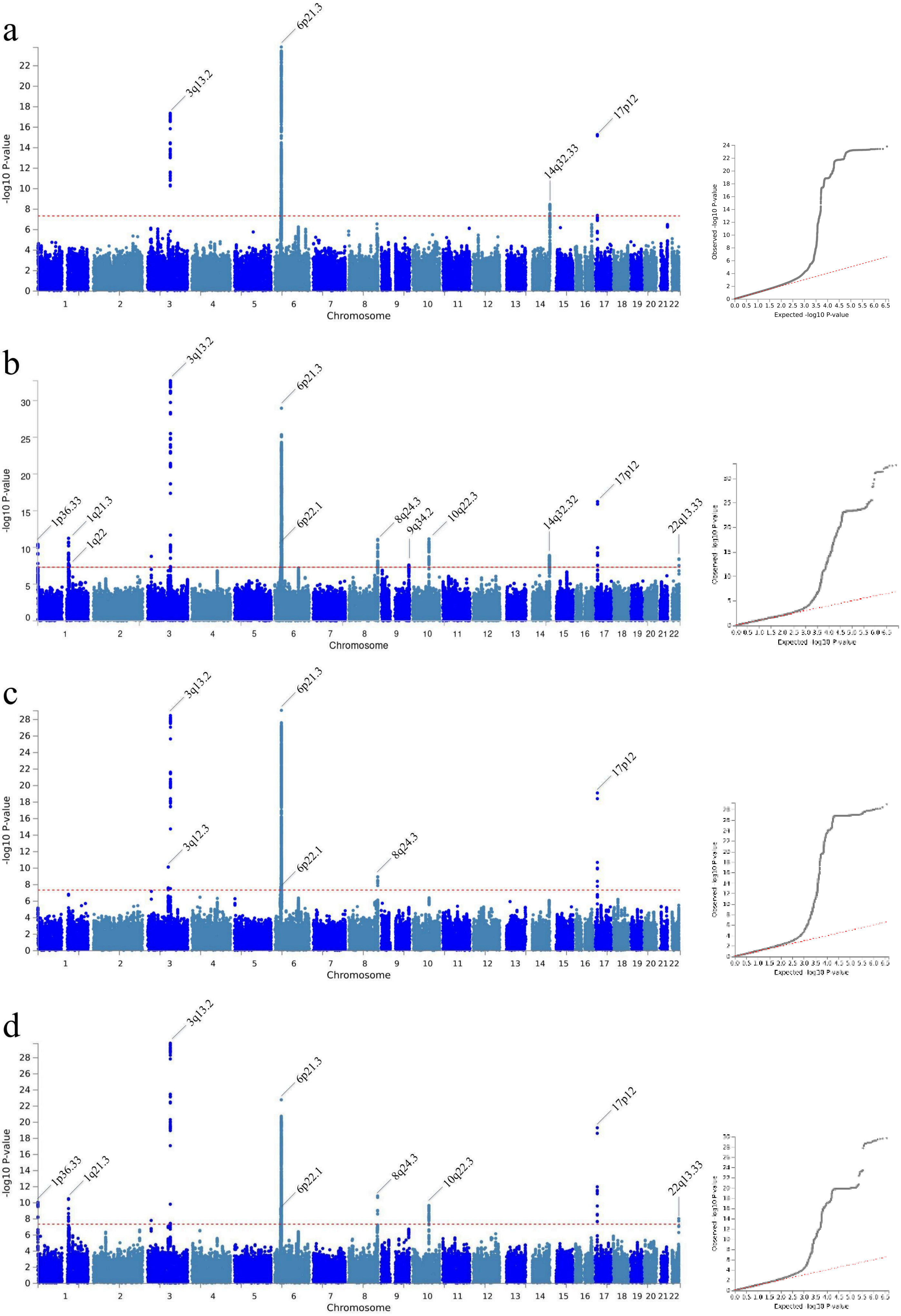
a
3q13.2
6p21.3
14q32.33
17p12
-log10 P-value
Chromosome
Observed -log10 P-value
Expected -log10 P-value
b
1p36.33
1q21.3
1q22
3q13.2
6p21.3
6p22.1
8q24.3
9q34.2
10q22.3
14q32.32
17p12
22q13.33
c
3q13.2
3q12.3
6p21.3
6p22.1
8q24.3
17p12
d
1p36.33
1q21.3
3q13.2
6p21.3
6p22.1
8q24.3
10q22.3
17p12
22q13.33

e

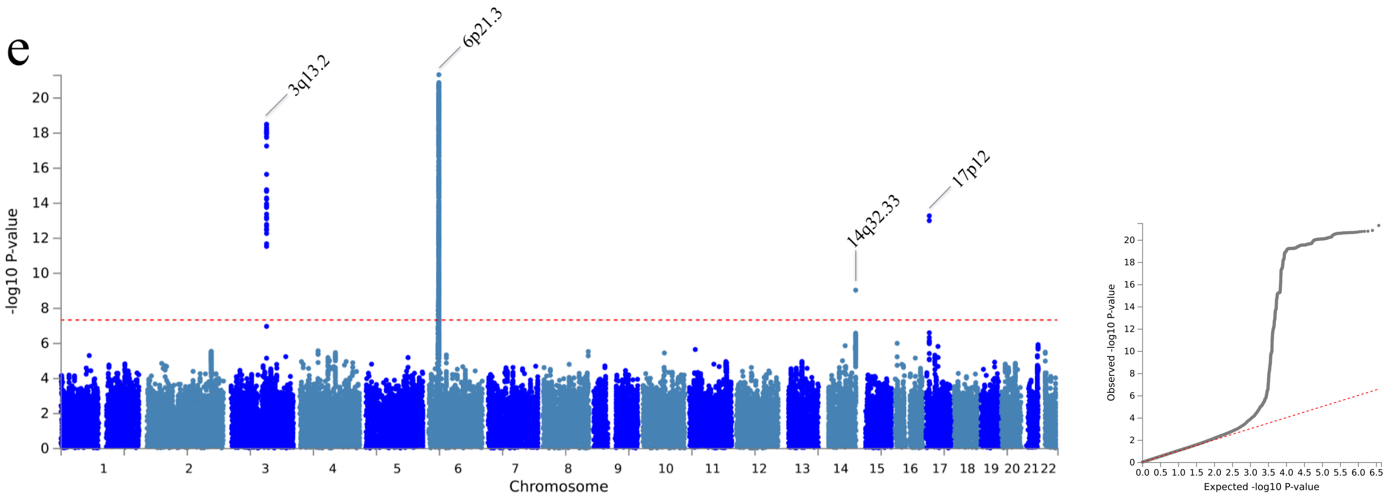


**Supplementary Fig. 4**
**Manhattan and Q-Q plots of the GWAS sensitivity analyses**

Manhattan plots and Quantile-Quantile (Q-Q) plots were generated using the FUMA platform (v1.6.1). **a,** Plots of the *H. pylori* eradication-exclusion GWAS. P-values were derived from the meta-analysis of 13,919 cases and 47,237 controls of Japanese ancestry. The x-axis represents the chromosomal positions and the y-axis represents the -log10 P-values. The red dashed line indicates the genome-wide significance threshold (P-value $< 5\times10^{-8}$). Variants are plotted against GRCh37 (hg19). In the Q-Q plot, the negative logarithm of the observed (y-axis) and the expected (x-axis) P-value were plotted for each SNP (dot), and the red dashed line ($y = x$) indicates the null hypothesis of no true association. The regression genomic inflation factor (λGC score) is 1.041. **b,** Plot of the cutoff-variation GWAS (3 U/ml threshold). P-values were derived from the meta-analysis of 58,106 cases and 67,072 controls of Japanese ancestry. The λGC score is 1.047. **c,** Plots of the cutoff-variation GWAS (top 25 % vs ≤ 3 U/ml). P-values were derived from the meta-analysis of 27,010 cases and 60,547 controls of Japanese ancestry. The λGC score is 1.059. **d,** Plots of the cutoff-variation GWAS (≥ 10 U/ml vs ≤ 3 U/ml). P-values were derived from the meta-analysis of 33,218 cases and 60,547 controls of Japanese ancestry. The λGC score is 1.058. **e,** Plots of the GWAS restricted to individuals aged 60 years or older. P-values were derived from the meta-analysis of 15,659 cases and 34,769 controls of Japanese ancestry. The λGC score is 1.051.

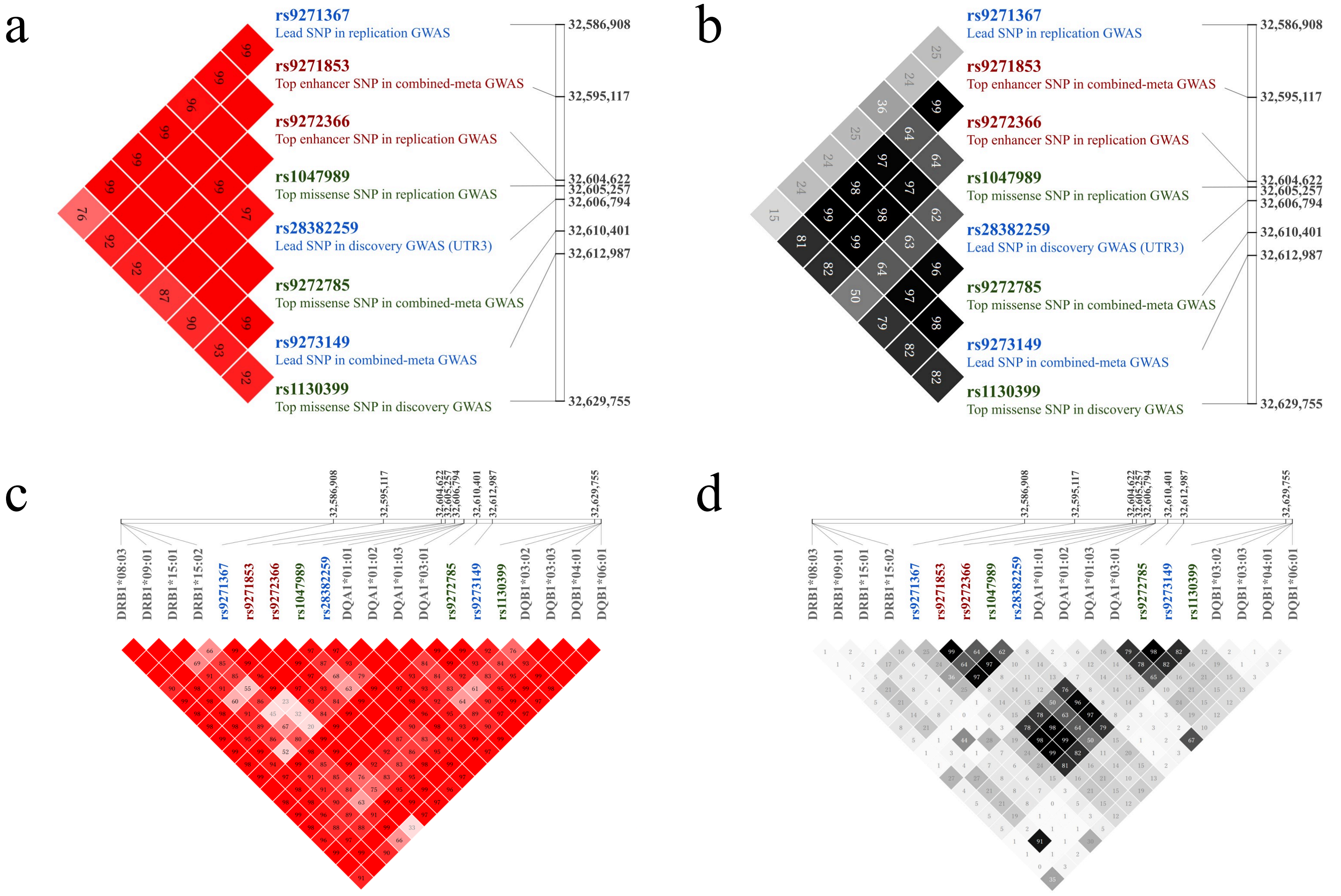


**Supplementary Fig. 5**

**LD heatmap of the relationship between GWAS Top variants and HLA-DRB1, -DQA1, and -DQB1 alleles.**

Linkage disequilibrium (LD) scale transitions from white to red to represent the coefficient of disequilibrium (D') values, and from white to black to represent the squared correlation coefficient ($r^2$) values. Base pair (BP) positions on chromosome 6 are referenced to the GRCh37 (hg19) genome assembly. **a,** D' between GWAS Top variants. **b,** $r^2$ between GWAS Top variants. **c,** D' between GWAS Top variants and HLA-DRB1, -DQA1, and -DQB1 alleles. **d,** $r^2$ between GWAS Top variants and HLA-DRB1, -DQA1, and -DQB1 alleles.

a

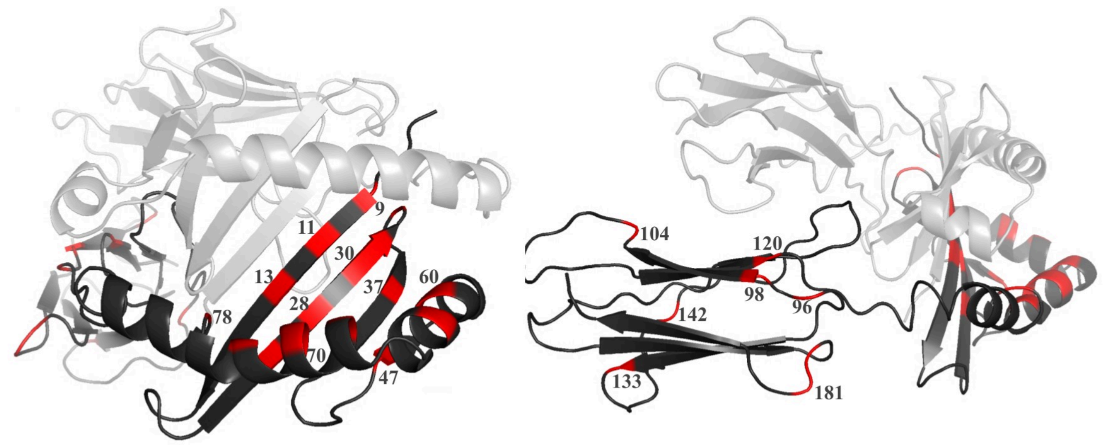


| P | 9 | 6 | 4 | 4 | 4,7 | 6 | | | 9 | 7 | 9 | 7 | 7 | 4,7 | 4 | 4 | 4 |
|---|---|---|---|---|---|---|---|---|---|---|---|---|---|---|---|---|---|
| AA | 9 | 11 | 13 | 26 | 28 | 30 | 31 | 33 | 37 | 47 | 57 | 60 | 67 | 70 | 71 | 74 | 78 |
| 04:05 | E | V | H | F | D | Y | F | H | Y | Y | S | Y | L | Q | R | A | Y |
| 08:03 | E | S | G | F | D | Y | F | N | Y | Y | S | Y | I | D | R | L | Y |
| 09:01 | K | D | F | Y | H | G | I | N | N | Y | V | S | F | R | R | E | V |
| 13:02 | E | S | S | F | D | Y | F | N | N | F | D | Y | I | D | E | A | Y |
| 15:01 | W | P | R | F | D | Y | F | N | S | F | D | Y | I | Q | A | A | Y |
| 15:02 | W | P | R | F | D | Y | F | N | S | F | D | Y | I | Q | A | A | Y |

| | | | | | | | | |
|---|---|---|---|---|---|---|---|---|
| AA | 96 | 98 | 104 | 120 | 133 | 142 | 180 | 181 |
| 04:05 | Y | E | A | N | R | V | L | T |
| 08:03 | H | K | S | S | R | V | V | T |
| 09:01 | H | E | A | S | R | V | V | M |
| 13:02 | H | K | S | S | R | V | V | T |
| 15:01 | Q | K | S | S | L | M | V | T |
| 15:02 | Q | K | S | S | L | M | V | T |

b

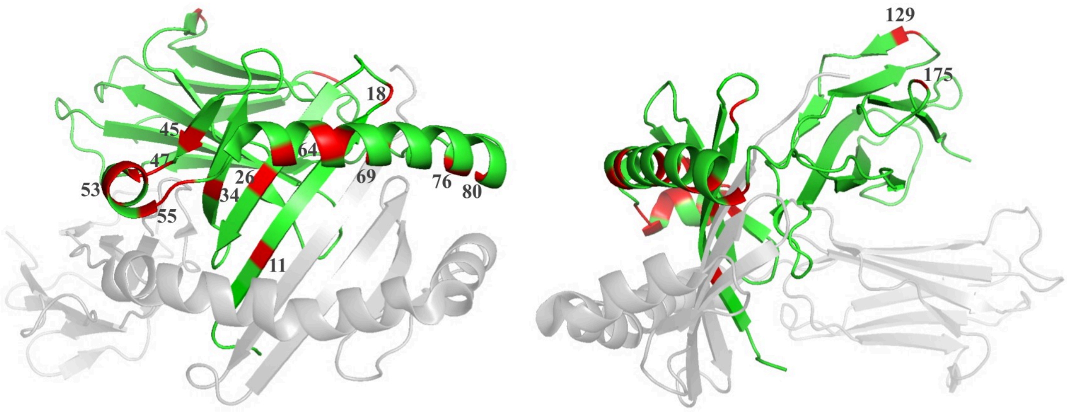


| P | 1 | | | 1 | | | | | | | 1 | 1 | | | | 6 | 9 | |
|---|---|---|---|---|---|---|---|---|---|---|---|---|---|---|---|---|---|---|
| AA | 11 | 18 | 26 | 34 | 45 | 47 | 48 | 50 | 52 | 53 | 55 | 56 | 61 | 64 | 66 | 69 | 76 | 80 |
| 01:01 | C | F | T | E | A | R | W | E | S | K | G | G | G | R | M | A | M | Y |
| 01:02 | C | F | T | Q | A | R | W | E | S | K | G | G | G | R | M | A | M | Y |
| 01:03 | C | F | T | Q | A | R | W | E | S | K | G | G | G | R | M | A | M | Y |
| 03:01 | Y | S | S | E | V | Q | L | L | R | R | R | R | F | T | I | L | V | S |
| 04:01 | Y | S | T | Q | V | C | L | V | R | Q | R | - | F | T | I | T | L | S |
| 05:01 | Y | S | T | Q | V | C | L | V | R | Q | R | - | F | T | I | L | L | S |

| | | |
|---|---|---|
| AA | 129 | 175 |
| 01:01 | Q | Q |
| 01:02 | Q | Q |
| 01:03 | Q | Q |
| 03:01 | H | E |
| 04:01 | H | E |
| 05:01 | H | K |

c

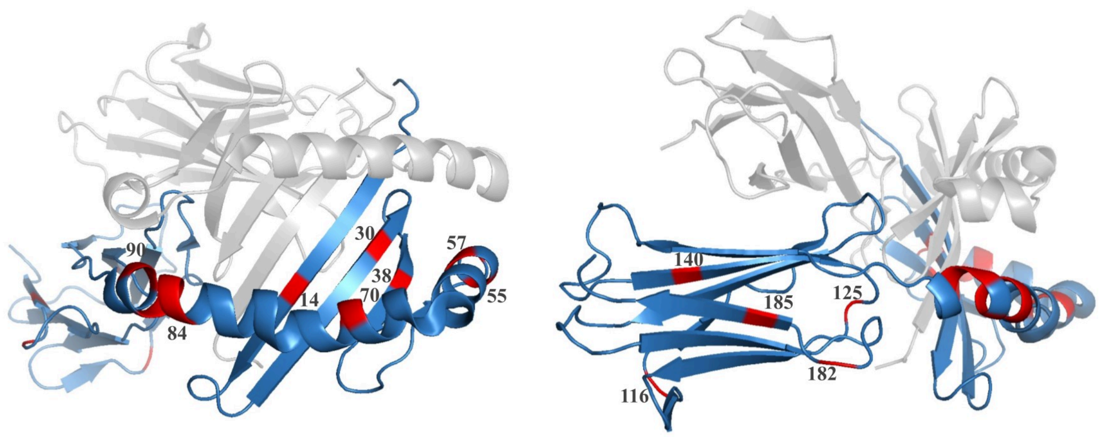


| P | | 6 | 9 | | | 9 | 7 | | | | 1 | 1 | | |
|---|---|---|---|---|---|---|---|---|---|---|---|---|---|---|
| AA | 14 | 30 | 38 | 53 | 55 | 57 | 70 | 71 | 84 | 85 | 86 | 87 | 89 | 90 |
| 03:01 | M | Y | A | L | P | D | R | T | Q | L | E | L | T | T |
| 03:02 | M | Y | A | L | P | A | R | T | Q | L | E | L | T | T |
| 03:03 | M | Y | A | L | P | D | R | T | Q | L | E | L | T | T |
| 04:01 | M | Y | A | L | R | D | E | D | Q | L | E | L | T | T |
| 06:01 | M | Y | V | Q | R | D | R | T | E | V | A | F | G | I |
| 06:02 | M | Y | A | Q | R | D | G | T | E | V | A | F | G | I |

| | | | | | |
|---|---|---|---|---|---|
| AA | 116 | 125 | 140 | 182 | 185 |
| 03:01 | V | A | T | N | T |
| 03:02 | V | A | T | N | I |
| 03:03 | V | A | T | N | I |
| 04:01 | V | A | T | N | I |
| 06:01 | V | G | A | S | T |
| 06:02 | V | G | A | S | T |

**Supplementary Fig. 6**
**The mapping of the associated amino acids onto the HLA-DRB1, -DQA1, and -DQB1 protein structures.**

**a,** The mapping of the associated amino acids with anti-*H. pylori* IgG antibody titers onto the HLA-DRB1 protein. P refers to the peptide pocket number, and AA refers to the amino acid position. Red font indicates amino acids associated with high antibody titers, while blue font indicates those associated with low antibody titers. **b,** The mapping of the associated amino acids onto the HLA-DQA1 protein. **c,** The mapping of the associated amino acids onto the HLA-DQB1 protein.

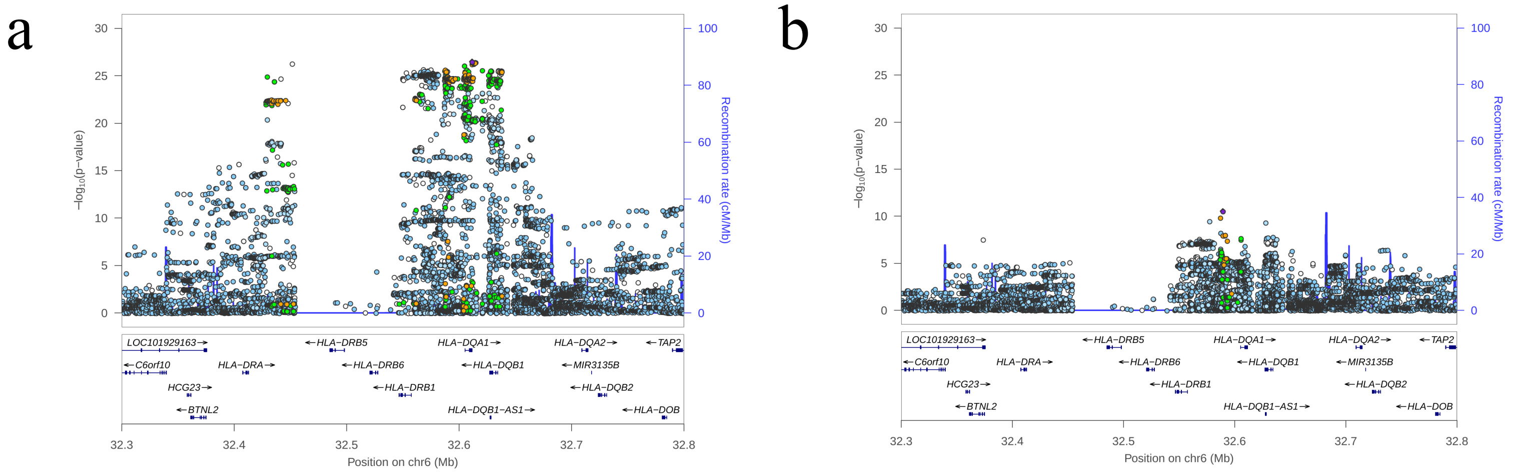


**Supplementary Fig. 7**

**Association signals around the significant region in 6p21.3 of the primary and conditional GWAS.**

The panel is association signals around significant locus. The X-axis represents chromosomal positions (GRC37/hg19) and the Y-axis represents -log10 P-values. The lead variant is shown in purple. Colors represent the degree of LD ($r^2$) between each variant and the lead variant. LD was calculated based on the 1000 Genomes Mar 2012 ASN data. **a,** Primary GWAS meta-analysis: P-values were derived from the meta-analysis of 27,034 cases and 78,897 controls of TMM cohorts. Regional association plot showing SNP significance and genes around lead SNP rs9273002 (P-value = $3.32\times10^{-27}$). **b,** Conditional GWAS (rs1047989 as covariate): Association signals in the same region from a conditional GWAS. Regional association plot showing SNP significance and genes around lead SNP rs9271501 (P-value = $2.89\times10^{-11}$).

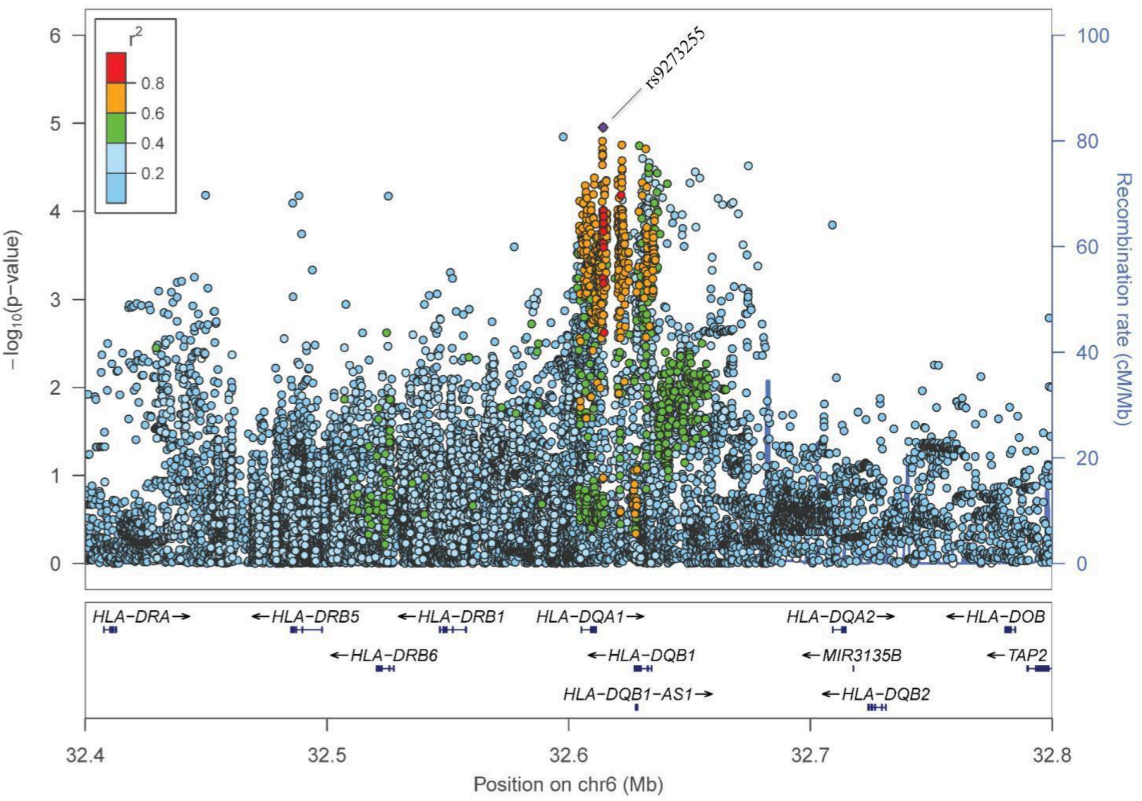


**Supplementary Fig. 8**
**Association signals around the significant region in 6p21.3 loci (previous European study).**

Regional association plot showing SNP significance and genes around lead SNP rs9273255. The panel is association signals around significant locus. The X-axis represents chromosomal positions (GRCh37/hg19) and the Y-axis represents -log10 P-values. The lead variant is shown in purple. Colors represent the degree of LD ($r^2$) between each variant and the lead variant. LD was calculated based on the 1000 Genomes Mar 2012 EUR data.

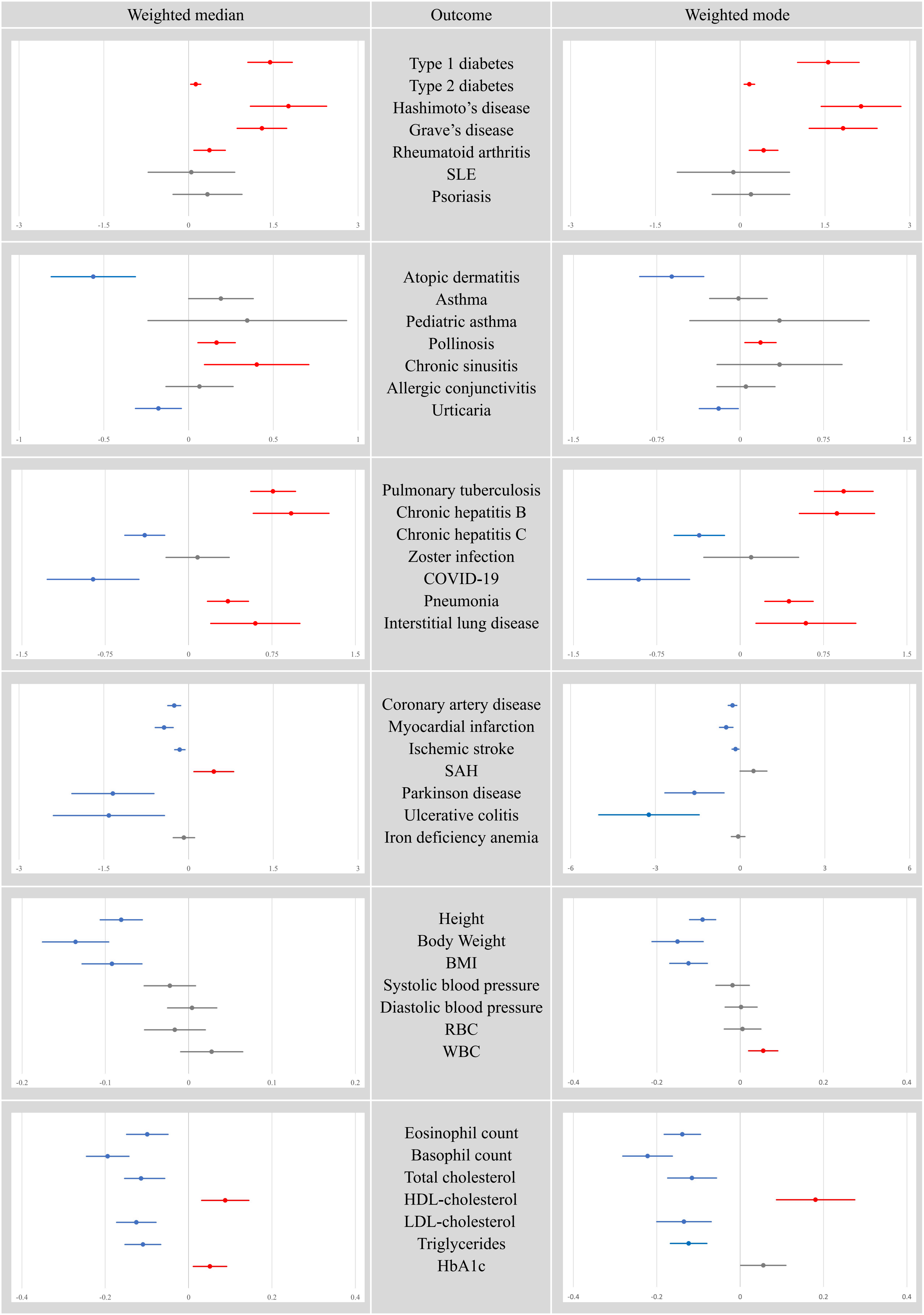
Weighted median
Outcome
Weighted mode
Type 1 diabetes
Type 2 diabetes
Hashimoto's disease
Grave's disease
Rheumatoid arthritis
SLE
Psoriasis
-3 -1.5 0 1.5 3
Atopic dermatitis
Asthma
Pediatric asthma
Pollinosis
Chronic sinusitis
Allergic conjunctivitis
Urticaria
-1 -0.5 0 0.5 1
-1.5 -0.75 0 0.75 1.5
Pulmonary tuberculosis
Chronic hepatitis B
Chronic hepatitis C
Zoster infection
COVID-19
Pneumonia
Interstitial lung disease
-1.5 -0.75 0 0.75 1.5
Coronary artery disease
Myocardial infarction
Ischemic stroke
SAH
Parkinson disease
Ulcerative colitis
Iron deficiency anemia
-3 -1.5 0 1.5 3
-6 -3 0 3 6
Height
Body Weight
BMI
Systolic blood pressure
Diastolic blood pressure
RBC
WBC
-0.2 -0.1 0 0.1 0.2
-0.4 -0.2 0 0.2 0.4
Eosinophil count
Basophil count
Total cholesterol
HDL-cholesterol
LDL-cholesterol
Triglycerides
HbA1c
-0.4 -0.2 0 0.2 0.4

**Supplementary Fig. 9**
**Forest plot of Mendelian randomization analysis (Weighted median, Weighted mode).**

Forest plot of Mendelian randomization (Weighted median and Weighted mode) for the association between genetic liability to high anti-*H. pylori* IgG levels and other diseases. The X-axis shows the Beta coefficients for the presence of each disease (95% CI). The red line indicates a positive association, the blue line indicates an inverse association, and the gray line indicates no significant association.

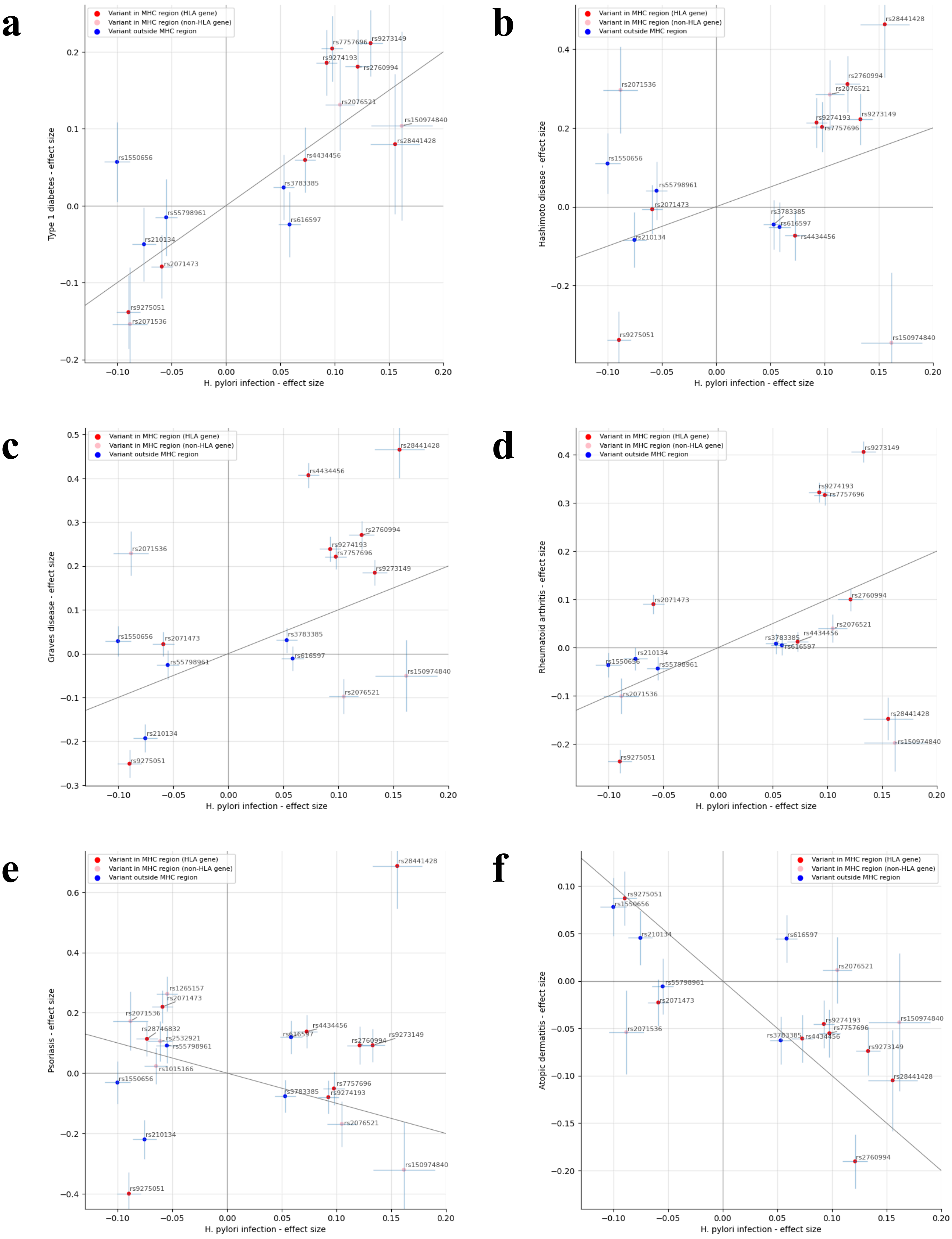

a
Variant in MHC region (HLA gene)
Variant in MHC region (non-HLA gene)
Variant outside MHC region
Type 1 diabetes - effect size
H. pylori infection - effect size
rs7757696
rs9273149
rs9274193
rs2760994
rs2076521
rs150974840
rs28441428
rs1550656
rs4434456
rs3783385
rs616597
rs55798961
rs210134
rs2071473
rs9275051
rs2071536
b
Hashimoto disease - effect size
c
Graves disease - effect size
d
Rheumatoid arthritis - effect size
e
Psoriasis - effect size
rs1265157
rs28746832
rs2532921
rs1015166
f
Atopic dermatitis - effect size

**Supplementary Fig. 10**
**Per-allele effect-size comparison between anti-*H. pylori* IgG levels and other diseases.**

Per-allele effect size comparison of variants used in the Mendelian randomization analysis between anti-*H. pylori* IgG levels and other diseases. Data are presented as effect size (BETA) ± standard error (s.e.). The X-axis represents the effect size for IgG levels, while the Y-axis represents the effect size for other diseases. Variants near the HLA gene in the MHC region are shown in red, those in the MHC region but not the HLA gene are shown in pink, and variants outside the MHC region are shown in blue. **a,** Comparison between IgG levels and Type 1 diabetes. **b,** Comparison between IgG levels and Hashimoto's disease. **c,** Comparison between IgG levels and Grave's disease. **d,** Comparison between IgG levels and Rheumatoid arthritis. **e,** Comparison between IgG levels and Psoriasis. **f,** Comparison between IgG levels and Atopic dermatitis.

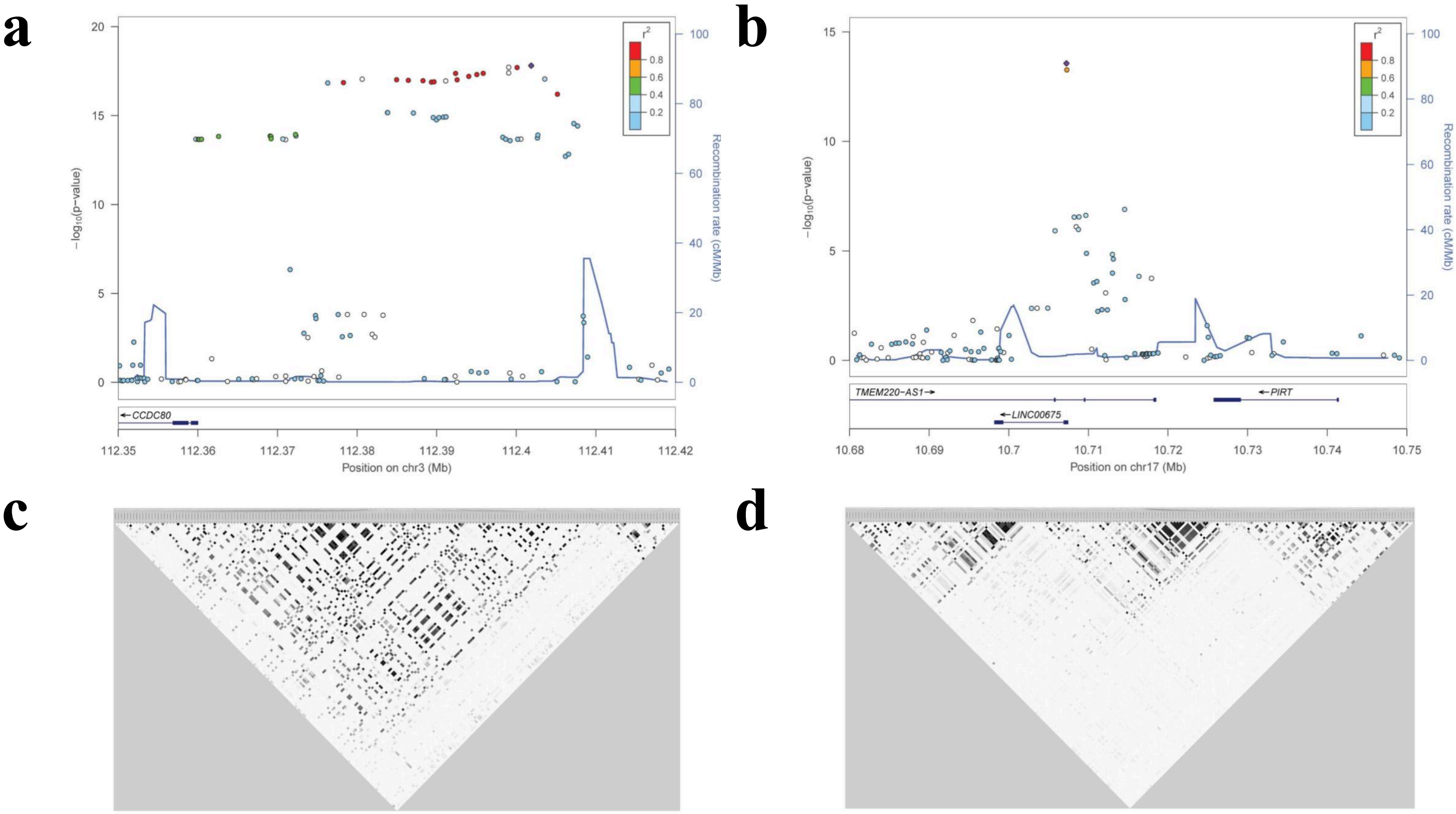


**Supplementary Fig. 11**

**Association signals around the significant regions in 3q13.2 and 17p12 loci.**

**a,** Association signals around the significant regions in 3q13.2 loci. Regional association plot showing SNP significance and genes around lead SNP rs146203219. The panel is association signals around significant locus in Discovery GWAS. The X-axis represents chromosomal positions (GRC37/hg19) and the Y-axis represents -log10 P-values. The lead variant is shown in purple. Colors represent the degree of LD ($r^2$) between each variant and the lead variant. LD was calculated based on the 1000 Genomes Mar 2012 ASN data. **b,** Association signals around the significant regions in 17p12 loci. Regional association plot showing SNP significance and genes around lead SNP rs1078643. **c,** LD heatmap corresponding to **a**. The dataset includes EAS populations from 1KGP (Phase 3), with a minor allele frequency (MAF) filter of 0.01. LD scale transitions from white to black to represent squared correlation coefficient ($r^2$) values. **d,** LD heatmap corresponding to **b**.

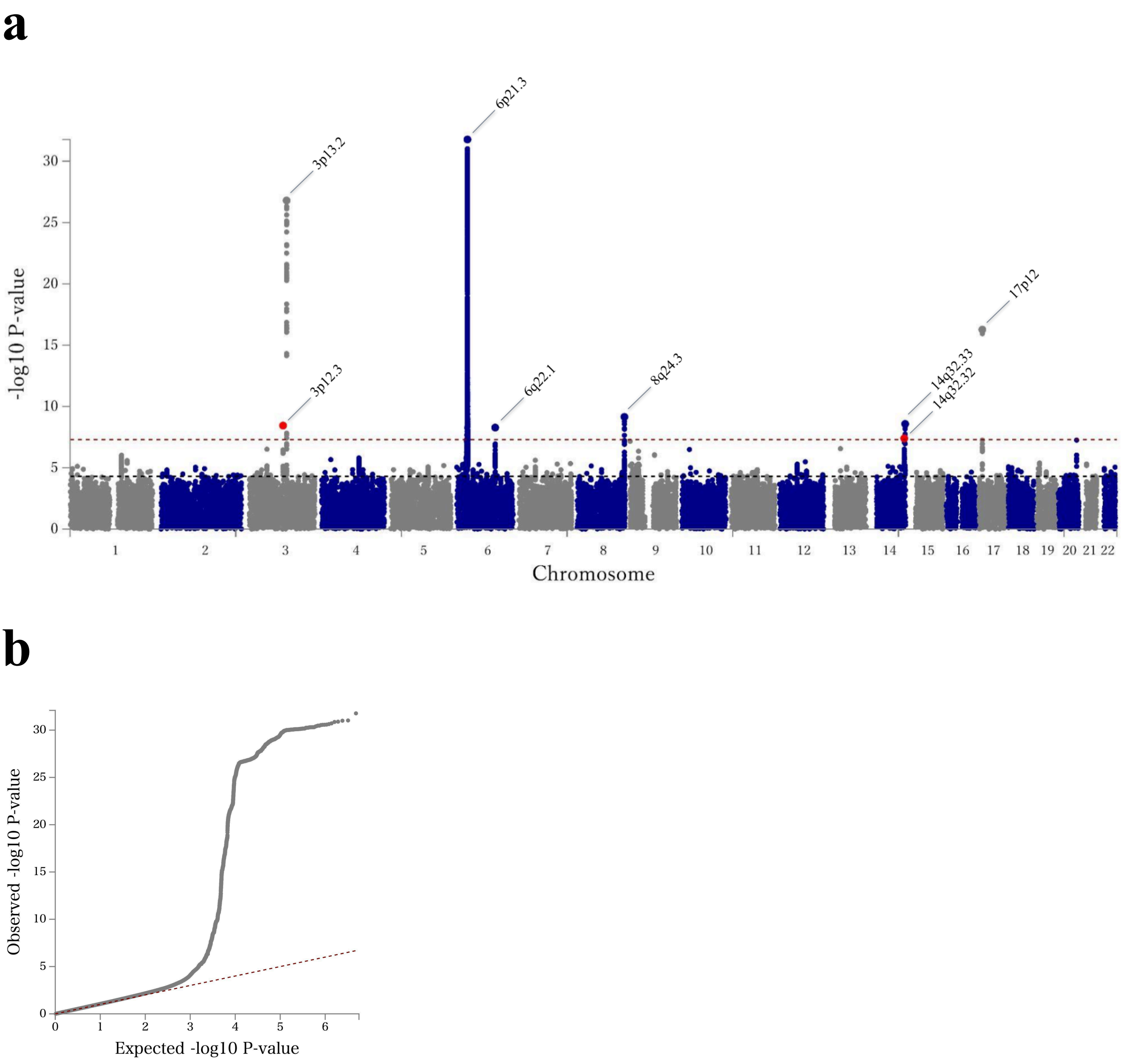


**Supplementary Fig. 12**

**Manhattan plot of the Meta-GWAS in Japan (using MR-MEGA).**

**a.** Manhattan plot of the Meta-GWAS in Japan. P-values were derived from the meta-analysis of 37,410 cases and 87,768 controls of Japanese ancestry. The X-axis represents the chromosomal positions and the Y-axis represents the – log10 P-values. The red dashed line indicates the genome-wide significance threshold (P-value $< 5\times10^{-8}$). The black dashed line indicates the suggestive threshold (P-value $< 5\times10^{-5}$). Variants are plotted against GRCh37 (hg19). **b,** Q-Q plots of the GWAS. The negative logarithm of the observed (y-axis) and the expected (x-axis) P-value was plotted for each SNP (dot), and the red dashed line ($y = x$) indicates the null hypothesis of no true association. The regression genomic inflation factor ($\lambda$ score) is 1.046.

**Consortia**

**the Tohoku Medical Megabank Project**

Hikaru Abe[1], Michiaki Abe[1], Momoka Abe[1], Naomi Abe[1], Noriko Abe[1], Tomomi Abe[1], Yuto Abe[1], Shizuko Ahiko[1], Kayo Aiki[1], Hiromi Aizawa[1], Yukari Akiyama[1], Hayato Anzawa[1], Eri Aoki[1], Yuichi Aoki[1], Hiroko Arai[1], Misaki Arakawa[1], Yukie Asano[1], Liam Baird[1], Ayano Chiba[1], Haruna Chiba[1], Ippei Chiba[1], Kenji Chiba[1], Tetsuo Chiba[1], Hisako Endo[1], Reika Fue[1], Futaba Fujishiro[1], Yayoi Fujita[1], Waka Fukunaga[1], Mami Funata[1], Takamitsu Funayama[1], Sho Furuhashi[1], Nobuo Fuse[1], Junko Fushimi[1], Kumiko Fushiya[1], Tomomi Gamo[1], Chinatsu Gocho[1], Katsuhiro Gonoi[1], Maki Goto[1], Takahiko Goto[1], Yukie Goto[1], Kaori Gouko[1], Michiko Haga[1], Yoko Haga[1], Hiroko Hamada[1], Yumiko Hamaie[1], Yohei Hamanaka[1], Mika Hanazawa[1], Yukari Hara[1], Atsushi Hasegawa[1], Asuka Hatakeyama[1], Sumika Hatakeyama[1], Nozomi Hatanaka[1], Rieko Hatanaka[1], Takanori Hidaka[1], Kenji Hino[1], Hiroe Hirama[1], Ikuo Hirano[1], Sachiko Hirano[1], Takumi Hirata[1], Masahiro Hiratsuka[1], Yuki Hiratsuka[1], Ikuko Hirayama[1], Eiji Hishinuma[1], Takako Hoshi[1], Atsushi Hozawa[1], Keisuke Ido[1], Nobuko Igari[1], Chikako Iida[1], Katsuko Imai[1], Makiko Inoue[1], Reiko Inoue[1], Rumi Irie[1], Motoko Ishida[1], Noriko Ishida[1], Eri Ishigaka[1], Chihiro Ishii[1], Kaori Ishii[1], Osamu Ishii[1], Tadashi Ishii[1], Tatsuro Ishikawa[1], Mami Ishikuro[1], Kazutoshi Ishimori[1], Miho Itabashi[1], Kumiko Ito[1], Maiko Ito[1], Masumi Ito[1], Mayumi Ito[1], Megumi Ito[1], Natsuko Ito[1], Rie Ito[1], Saori Ito[1], Fumihiko Iwabuchi[1], Maki Iwabuchi[1], Yoko Izumi[1], Yoshiko Izumi[1], Masataka Kambe[1], Takanari Kanno[1], Mayu Kano[1], Naoko Kasahara[1], Hinako Kashiwa[1], Kiyomi Katahira[1], Mayumi Kato[1], Yukie Kato[1], Fumiki Katsuoka[1], Takeshi Kawabata[1], Rika Kawada[1], Aoi Kawagoe[1], Hiroshi Kawame[1], Junko Kawashima[1], Yukako Kawashima[1], Junko Kikuchi[1], Tsuyoshi Kikukawa[1], Masahiro Kikuya[1], Masae Kimura[1], Michiko Kimura[1], Kengo Kinoshita[1], Ikuko Kishi[1], Tomoko Kishimoto[1], Tamie Kitaura[1], Mika Kobayashi[1], Tadao Kobayashi[1], Tomoko Kobayashi[1], Eiichi N. Kodama[1], Shun Kodate[1], Mana Kogure[1], Toshisada Kohagizawa[1], Naomi Kohketsu[1], Noa Koida[1], Chie Koide[1], Mika Koide[1], Toshihiko Koike[1], Kaname Kojima[1], Junko Komatsu[1], Ayumi Kondo[1], Riyo Konno[1], Yukie Konno[1], Sachie Koreeda[1], Seizo Koshiba[1], Takuya Koyama[1], Hisaaki Kudo[1], Kazuki Kumada[1], Ryoko Kumadaki[1], Rika Kumagai[1], Toshie Kumagai[1], Yuko Kumagai[1], Yasuto Kunii[1], Miho Kuriki[1], Shinichi Kuriyama[1], Emiko Kurokawa[1], Seiko Kurota[1], Hisako Kusano[1], Bin Li[1], Donghan Li[1], Xue Li[1], Kanako Maeshibu[1], Keiko Maeta[1], Satoshi Makino[1], Hiroko Matsubara[1], Naomi Matsukawa[1], Masako Matsumoto[1], Takako Matsuoka[1], Yuka Matsushita[1], Motomichi Matsuzaki[1], Hirohito Metoki[1], Sayaka Minakawa[1], Yuki Minami[1], Suzuki Mirei[1], Kyoko Mitate[1], Satomi Mito[1], Ayako Miura[1], Noriko Miura[1], Ryo Miyagi[1], Akiko Miyazawa[1], Satoshi Mizuno[1], Akiko Mochida[1], Mika Momii[1], Hiroko Mori[1], Naoko Mori[1], Hozumi Motohashi[1], Ikuko N. Motoike[1], Shunji Mugikura[1], Keiko Murakami[1], Takahisa Murakami[1], Masato Nagai[1,2], Satoshi Nagaie[1], Fuji Nagami[1], Toko Naganuma[1], Tatsuo Nagasaka[1], Sachiko Nagase[1], Kumiko Nakagawa[1], Taku Nakai[1], Noriko Nakajo[1], Kyoko Nakamichi[1], Chie Nakamura[1], Naoki Nakamura[1], Tomohiro Nakamura[1], Yuko Nakasato[1], Kumi Nakaya[1], Naoki Nakaya[1], Kei Nanatani[1], Akira Narita[1], Yuka Narita[1], Yasuhisa Nemoto[1], Hafumi Nishi[1], Kohji Nishida[1], Ichiko Nishijima[1], Momo Nishiyama[1], Takahiro Nobukuni[1], Kotaro Nochioka[1], Aoi Noda[1], Kenichi Noguchi[1], Kiriko Nozoe[1], Rie Nunokawa[1], Taku Obara[1], Tomoko Obara[1], Kaori Ogasawara[1], Satoru Ogawa[1], Soichi Ogishima[1], Natsuki Oguma[1], Nahoko Ohi[1], Namiko Ohisa[1], Kinuko Ohneda[1], Hayami Ohori[1], Miri Oikawa[1], Yumi Oikawa[1], Yumiko Ojima[1], Yumi Okada[1], Yasunobu Okamura[1], Hiroshi Okuda[1], Mitsuko Okuda[1], Ayako Okumoto[1], Akane Ono[1], Chiaki Ono[1], Genki Onodera[1], Kaname Onodera[1], Masako Onodera[1], Midori Onuma[1], Tomomi Onuma[1], Keiichiro Oohashi[1], Masumi Oomachi[1], Kazuya Ootomo[1], Yukie Oouchi[1], Masatsugu Orui[1], Mayumi Osada[1], Tamae Osanai[1], Reiko Ota[1], Noriko Otake[1], Sumie Otomo[1], Akihito Otsuki[1], Yoko Otsuki[1], Yuki Oyama[1], Keiko Oyamada[1], Yoko Ozawa[1], Satomi Obara[1], Daisuke Saigusa[1], Asami Saito[1], Hisako Saito[1], Kazue Saito[1], Manami Saito[1], Megumi Saito[1], Ritsumi Saito[1], Sakae Saito[1], Tomo Saito[1], Yoko Saito[1], Yuki Saito[1], Yoshinobu Saitoh[1], Hiroko Sakai[1], Masaki Sakaida[1], Hiroshi Sakamono[1], Hiromi Sakamoto[1], Kana Sakamoto[1], Mia Sakamoto[1], Kasumi Sakurai[1], Miyuki Sakurai[1], Rieko Sakurai[1], Mika Sakurai-Yageta[1], Kana Sasaki[1], Miho Sasaki[1], Tadashi Sasaki[1], Yukari Sasaki[1], Yukie Sasaki[1], Chika Sato[1], Hirokazu Sato[1], Michiyo Sato[1], Miho Sato[1], Mitsuharu Sato[1], Miu Sato[1], Naoko Sato[1], Reiko Sato[1], Satoshi Sato[1], Shiho Sato[1], Taku Sato[1], Yoshiko Sato[1], Youko Sato[1], Yui Sato[1], Michihiro Satoh[1], Ayako Sekiya[1], Mariko Seo[1], Yoshiko Shima[1], Muneaki Shimada[1], Atsushi Shimizu[1,2], Ritsuko Shimizu[1], Genki Shinoda[1], Nobuyuki Shirakawa[1], Matsuyuki Shirota[1], Hiroe Shoji[1], Ikuko Shoji[1], Mariko Shoji[1],

Midori Shoji[1], Wakako Shoji[1], Satomi Someya[1], Shinya Sonobe[1], Itsumi Sou[1], Rie Suenaga[1], Yasuko Suenaga[1], Mayumi Suga[1], Rika Sugai[1], Junichi Sugawara[1], Megumi Sugawara[1], Michiko Sugawara[1], Nanako Sugawara[1], Saori Sugawara[1], Yuki Sugawara[1], Sachiyo Sugimoto[1], Airi Suzuki[1], Ayano Suzuki[1], Keiko P. Suzuki[1], Michirou Suzuki[1], Mikiko Suzuki[1], Norio Suzuki[1], Rie Suzuki[1], Ryoko Suzuki[1], Takafumi Suzuki[1], Tatsuya Suzuki[1], Yoichi Suzuki[1], Shu Tadaka[1], Keiko Taguchi[1], Nozomi Taiji[1], Makiko Taira[1], Kaori Takagi[1], Emi Takahashi[1], Harumi Takahashi[1], Junko Takahashi[1], Megumi Takahashi[1], Noriko Takahashi[1], Rieko Takahashi[1], Yukiko Takahashi[1], Mayuko Takasawa[1], Masato Takase[1], Jun Takayama[1], Miho Takeuchi[1], Sayaka Takita[1], Toru Tamahara[1], Gen Tamiya[1], Naomi Tamura[1], Akari Tanaka[1], Saiko Tanaka[1], Chihiro Tanno[1], Naoko Tanno[1], Keiko Tateno[1], Minoru Tateno[1], Chika Terui[1], Mihoko Toki[1], Sayuri Tokioka[1], Etsuko Tomita[1], Hiroaki Tomita[1], Mai Tomizuka[1], Naho Tsuchiya[1], Miyuki Tsuda[1], Tomomi Tsumuraya[1], Junko Tsunasawa[1], Issei Tsunoda[1], Juri Uchiya[1], Akiko Ueda[1], Yuriko Ueki[1], Fumihiko Ueno[1], Keiko Umeda[1], Akira Uruno[1], Ikuko Wada[1], Tomoko Wada[1], Mika Wagatsuma[1], Hitoshi Watanabe[1], Kanako Watanabe[1], Kazue Watanabe[1], Nobuo Yaegashi[1], Mika Yagyu[1], Etsuko Yamada[1], Yumi Yamaguchi-Kabata[1], Hiroko Yamamoto[1], Masayuki Yamamoto[1], Yukari Yamauchi[1], Mika Yamazaki[1], Jun Yasuda[1], Hang Yin[1], Hiroshi Yokota[1], Manami Yokoyama[1], Marie Yokoyama[1], Tomoko Yokoyama[1], Yuko Yoshida[1], Mizue Yoshino[1], Zhiqian Yu[1], Lin Zhang[1], Makoto Sasaki[2], Yasushi Ishigaki[2], Koichi Asahi[2], Ryoichi Tanaka[2], Kozo Tanno[2], Kotaro Otsuka[2], Naoyuki Nishiya[2], Mitsuko Iwabuchi[2], Fumitaka Tanaka[2], Shinichi Omama[2], Hiroshi Akasaka[2], Kouhei Hashizume[2], Noriko Takebe[2], Kazuhiro Yoshikawa[2], Yuka Kotozaki[2], Yorihiko Koeda[2], Takahiro Mikami[2], Takahito Nasu[2], Junko Akai[2], Nobuyuki Takanashi[2], Kasumi Hannokizawa[2], Hideki Ohmomo[2], Shohei Komaki[2], Mamoru Satoh[2], Fumiaki Takanashi[2], Yoichi Sutoh[2], Fumio Yamashita[2], Yutaka Hasegawa[2], Motoki Nakao[2], Yayoi Yamasaki[2], Shiori Minabe[2], Tsuyoshi Hachiya[2], Nobuhiro Suzumori[2], Yukiko Toya[2], Akiko Yoshida[2], Satoshi Nishizuka[2], and Ryujin Endo[2].

**the Biobank Japan Project**

Koichi Matsuda[3,4], Takayuki Morisaki[4,5], Yukinori Okada[6], Yoichiro Kamatani[7], Kaori Muto[8], Akiko Nagai[8], Yoji Sagiya[4], Natsuhiko Kumasaka[9], Yoichi Furukawa[10], Yuji Yamanashi[5], Yoshinori Murakami[5], Yusuke Nakamura[5], Wataru Obara[11], Ken Yamaji[12], Kazuhisa Takahashi[13], Satoshi Asai[14,15], Yasuo Takahashi[15], Shinichi Higashiue[16], Shuzo Kobayashi[16], Hiroki Yamaguchi[17], Yasunobu Nagata[17], Satoshi Wakita[17], Chikako Nito[18], Yu-ki Iwasaki[19], Shigeo Murayama[20], Kozo Yoshimori[21], Yoshio Miki[22], Daisuke Obata[23], Masahiko Higashiyama[24], Akihide Masumoto[25], Yoshinobu Koga[25], and Yukihiro Koretsune[26].

1. Tohoku Medical Megabank Organization, Tohoku University, Sendai, Japan
2. Iwate Tohoku Medical Megabank Organization, Iwate Medical University, Iwate, Japan
3. Laboratory of Genome Technology, Human Genome Center, Institute of Medical Science, The University of Tokyo, Tokyo, Japan.
4. Laboratory of Clinical Genome Sequencing, Graduate School of Frontier Sciences, The University of Tokyo, Tokyo, Japan.
5. The Institute of Medical Science, The University of Tokyo, Tokyo, Japan.
6. Department of Genome Informatics, Graduate School of Medicine, The University of Tokyo, Tokyo, Japan.
7. Laboratory of Complex Trait Genomics, Graduate School of Frontier Sciences, The University of Tokyo, Tokyo, Japan.
8. Department of Public Policy, Institute of Medical Science, The University of Tokyo, Tokyo, Japan.
9. Division of Digital Genomics, Institute of Medical Science, The University of Tokyo, Tokyo, Japan.
10. Division of Clinical Genome Research, Institute of Medical Science, The University of Tokyo, Tokyo, Japan.
11. Department of Urology, Iwate Medical University, Iwate, Japan.
12. Department of Internal Medicine and Rheumatology, Juntendo University Graduate School of Medicine, Tokyo, Japan.
13. Department of Respiratory Medicine, Juntendo University Graduate School of Medicine, Tokyo, Japan.

14. Division of Pharmacology, Department of Biomedical Science, Nihon University School of Medicine, Tokyo, Japan.
15. Division of Genomic Epidemiology and Clinical Trials, Clinical Trials Research Center, Nihon University. School of Medicine, Tokyo, Japan.
16. Tokushukai Group, Tokyo, Japan.
17. Department of Hematology, Nippon Medical School, Tokyo, Japan.
18. Laboratory for Clinical Research, Collaborative Research Center, Nippon Medical School, Tokyo, Japan.
19. Department of Cardiovascular Medicine, Nippon Medical School, Tokyo, Japan.
20. Tokyo Metropolitan Geriatric Hospital and Institute of Gerontology, Tokyo, Japan.
21. Fukujuji Hospital, Japan Anti-Tuberculosis Association, Tokyo, Japan.
22. The Cancer Institute Hospital of the Japanese Foundation for Cancer Research, Tokyo, Japan.
23. Center for Clinical Research and Advanced Medicine, Shiga University of Medical Science, Shiga, Japan.
24. Department of General Thoracic Surgery, Osaka International Cancer Institute, Osaka, Japan.
25. Iizuka Hospital, Fukuoka, Japan.
26. National Hospital Organization Osaka National Hospital, Osaka, Japan.
